# A scoping review of trauma informed approaches in acute, crisis, emergency and residential mental health care

**DOI:** 10.1101/2023.01.16.22283243

**Authors:** Katherine R. K. Saunders, Elizabeth McGuinness, Phoebe Barnett, Una Foye, Jessica Sears, Sophie Carlisle, Felicity Allman, Vasiliki Tzouvara, Merle Schlief, Norha Vera San Juan, Ruth Stuart, Jessica Griffiths, Rebecca Appleton, Paul McCrone, Rachel Rowan Olive, Patrick Nyikavaranda, Tamar Jeynes, Lizzie Mitchell, Alan Simpson, Sonia Johnson, Kylee Trevillion

**Affiliations:** NIHR Mental Health Policy Research Unit, Institute of Psychiatry, Psychology and Neuroscience, King’s College London, London, UK; NIHR Mental Health Policy Research Unit, Division of Psychiatry, University College London, UK; Centre for Outcomes Research and Effectiveness, Research Department of Clinical, Educational, & Health Psychology, University College London, London, UK; National Collaborating Centre for Mental Health, Royal College of Psychiatrists, London, UK; South London and Maudsley NHS Foundation Trust, London, UK; Section of Women’s Mental Health, King’s College London, London, UK; Care for Long Term Conditions Research Division, King’s College London, London, UK; Institute for Lifecourse Development, University of Greenwich, London, UK; School of Health Sciences, University of Greenwich, London, UK; NIHR Mental Health Policy Research Unit Lived Experience Working Group, Division of Psychiatry, University College London, UK

## Abstract

Experiences of trauma in childhood and adulthood are highly prevalent among service users accessing acute, crisis, emergency, and residential mental health services. These settings, and restraint and seclusion practices used, can be extremely traumatic, leading to a growing awareness for the need for trauma informed care (TIC). The aim of TIC is to acknowledge the prevalence and impact of trauma and create a safe environment to prevent re-traumatisation. This scoping review maps the TIC approaches delivered in these settings and reports related service user and staff experiences and attitudes, staff wellbeing, and service use outcomes.

We searched seven databases (EMBASE; PsycINFO; MEDLINE; Web of Science; Social Policy and Practice; Maternity and Infant Care Database; Cochrane Library Trials Register) between 24/02/2022-10/03/2022, used backwards and forwards citation tracking, and consulted academic and lived experience experts, identifying 4244 potentially relevant studies. Thirty-one studies were included.

Most studies (n=23) were conducted in the USA and were based in acute mental health services (n=16). We identified few trials, limiting inferences that can be drawn from the findings. The Six Core Strategies (n=7) and the Sanctuary Model (n=6) were the most commonly reported approaches. Rates of restraint and seclusion reportedly decreased. Some service users reported feeling trusted and cared for, while staff reported feeling empathy for service users and having a greater understanding of trauma. Staff reported needing training to deliver TIC effectively.

TIC principles should be at the core of all mental health service delivery. Implementing TIC approaches may integrate best practice into mental health care, although significant time and financial resources are required to implement organisational change at scale. Most evidence is preliminary in nature, and confined to acute and residential services, with little evidence on community crisis or emergency services. Clinical and research developments should prioritise lived experience expertise in addressing these gaps.

## Introduction

The concept of providing ‘trauma informed care’ (TIC) in healthcare settings has developed in response to increasing recognition that potentially traumatic experiences throughout the life course are associated with subsequent psychological distress and a range of mental health problems (1–4). ‘Trauma’ is a broad term which has no universally agreed definition. The Substance Abuse and Mental Health Services Administration (SAMHSA) defined trauma as ‘an event, series of events, or set of circumstances that is experienced by an individual as physically or emotionally harmful or life threatening and that has lasting adverse effects on the individual’s functioning and mental, physical, social, emotional, or spiritual well-being’ (5). Traumatic experiences can include physical, sexual and/or emotional abuse, neglect, exposure to violence or conflict, physical or mental illness (personal experience or that of a family member), and systemic or social traumas (6, 7).

Research has shown that individuals engaged with mental health services report high levels of childhood and adulthood trauma (2, 8–10). There is a high prevalence of trauma among service users in acute services, including among women (2, 11), those with psychosis (12, 13), and “personality disorder” diagnoses (14) (which is a particularly controversial diagnosis (15) as a result of the stigma associated with this diagnostic label and disparities in quality of care experienced (16–19)). Electronic health record evidence shows that service users with a history of abuse during childhood have more comorbidities and are more likely to have inpatient admissions versus service users without a similar history (20). Similarly, among people with long-term mental health conditions, rates of childhood trauma and adversity are high, with both experiences theorised as aetiological factors for mental health conditions (21–23). Staff in acute services are also affected by trauma experienced at work, which are highlighted as a source of stress and create a cycle of ‘reciprocal traumatisation’ (24, 25).

Inpatient, crisis, emergency and residential mental health care settings (typology of care categories derived from an exploration of the range, accessibility, and quality of acute psychiatric services (26)) are used by service users experiencing severe mental health episodes. Such settings include acute wards, community crisis teams, psychiatry liaison teams within emergency departments, and mental health crisis houses. Compulsory detention under mental health legislation, e.g., the Mental Health Act in the UK, which affects many service users in these settings, has the potential to traumatise or retraumatise (27). Routine staff procedures for managing the behaviour of distressed service users in inpatient settings, including seclusion and restraint, have also been shown to be destabilising and retraumatising among people who have experienced trauma (6, 28) as well as constituting a traumatic experience in their own right (24, 29). The power imbalances in these settings can create abusive dynamics, as well as mirror previous abusive relationships and situations (6), engendering mistrust and creating a harmful, rather than healing and supportive, environment.

In principle, TIC centres an understanding of the prevalence and impact of trauma, recognises trauma, responds comprehensively to trauma and takes steps to avoid re-traumatisation (5). The aim of TIC in healthcare is to create a therapeutic environment where communication, empathy and collaboration are prioritised. The TIC literature in healthcare is varied and lacks an agreed definition. However, Sweeney and Taggart (2018), who both write from dual perspectives as researchers and trauma survivors, developed an adapted definition of TIC (5, 30, 31) which we have used as a working definition throughout this scoping review. They define TIC as ‘a programme or organisational/system approach that: (1) understands and acknowledges the links between trauma and mental health, (2) adopts a broad definition of trauma which recognises social trauma and the intersectionality of multiple traumas, (3) undertakes sensitive enquiry into trauma experiences, (4) refers individuals to evidence-based trauma-specific support, (5) addresses vicarious trauma and re-traumatisation, (6) prioritises trustworthiness and transparency in communications, (7) seeks to establish collaborative relationships with service users, (8) adopts a strengths-based approach to care, (9) prioritises emotional and physical safety of service users, (10) works in partnership with trauma survivors to design, deliver and evaluate services.’ Although TIC provides a service structure which supports service users who have experienced trauma, there may be benefits for all service users in contact with mental health services. TIC principles are also helpful more broadly for avoiding traumatisation of service users.

TIC within inpatient, crisis, emergency, and residential mental health care settings is fairly newly established, and there is a lack of research mapping its use. The aim of this scoping review is to identify, map and explore the TIC models and approaches used in these settings, and to review impacts on and experiences of service users and staff. We also highlight gaps and variability in literature and service provision. TIC is a broad term, and it has been applied in numerous and varied ways in mental health care. In this review, we describe each application of TIC as a ‘trauma informed approach’.

This scoping review will answer the following primary research question:

1. What trauma informed approaches are used in acute, crisis, emergency, and residential mental health care?

Within each trauma informed approach identified, we will answer the following secondary research questions:

2. What is known about service user and carer expectations and experiences of TIC in acute, crisis, emergency, and residential mental health care?
3. How does TIC in acute, crisis, emergency, and residential mental health care impact on service user outcomes?
4. What is known about staff attitudes, expectations, and experiences of delivering TIC in acute, crisis, emergency, and residential mental health care?
5. How does TIC impact on staff practices and staff wellbeing acute, crisis, emergency, and residential mental health care?
6. How does TIC in acute, crisis, emergency and residential mental health care impact on service use and service costs, and what evidence exists about their cost-effectiveness?

## Methods

### Study design

This scoping review was conducted in accordance with the Preferred Reporting Items for Systematic Reviews and Meta-Analyses Extension for Scoping Reviews (PRISMA-ScR (32)), using a framework for conducting scoping reviews (33). The PRISMA-ScR checklist can be seen in Appendix 1. The protocol was registered with the Open Science Framework ahead of conducting the searches (https://osf.io/2b5w7). The review was steered by a team including academic experts, clinical researchers, and experts by experience and/or profession, with lived experience researchers contributing to the development of the research questions, the data extraction form, the interpretation, and the manuscript draft.

### Eligibility criteria

#### Population

Service users, or people who support or care for service users, of any age (both adults and children), gender or sexuality, or staff members (of any gender and sexuality) were included.

#### Setting

We included studies that focused (or provided disaggregated data) on care delivered within acute, crisis, emergency settings, or residential mental health settings; acute and crisis settings include inpatient, community-based crisis, hospital emergency department, acute day units and crisis houses. Forensic mental health and substance use acute, crisis and inpatient settings were also included. We excluded studies from general population prison settings, where there is debate as to whether TIC can be delivered in carceral settings (34), and residential settings where the primary purpose of the setting was not to provide mental health or psychiatric care (e.g., foster care or residential schools).

#### Intervention

Trauma informed care interventions. Programmes aiming to reduce restrictive practices in psychiatric settings were not included without explicit reference to TIC within the programme.

#### Outcomes

We included studies reporting any positive and adverse individual-, interpersonal-, service- and/or system-level outcomes, including outcomes from the implementation, use or testing of TIC. Individual-level outcomes are related to service user or staff experiences, attitudes, and expectations; interpersonal outcomes occur because of interactions between staff and service users; service-level outcomes include TIC procedures that occur on an individual service level; and system-level outcomes refer to broader organisational outcomes related to TIC implementation. We included studies exploring service user, staff and carer expectations and experiences of TIC approaches.

#### Types of studies

We included qualitative, quantitative, or mixed-method research study designs. To map TIC provision, service descriptions, evaluations, audits, and case studies of individual service provision were also included. We excluded reviews, conference abstracts with no associated paper, protocols, editorials, policy briefings, books/book chapters, personal blogs/commentaries, and BSc and MSc theses. We included non-English studies that our team could translate (English, German, Spanish). Both peer-reviewed and grey literature sources were eligible.

### Search strategy

A three-step search strategy was used. Firstly, we searched seven databases between 24/02/2022 and 10/03/2022: EMBASE; PsycINFO; MEDLINE; Web of Science; Social Policy and Practice; Maternity and Infant Care Database (formerly MIDIRS); Cochrane Library Trials Register. An example full search strategy can be seen in Appendix 2. Searches were also run in one electronic grey literature database (Social Care Online); two pre-print servers (medRxiv and PsyArXiv), and two PhD thesis websites (EThOS and DART). The search strategy used terms adapted from related reviews (35–44). We added specific health economic search terms. No date or language limits were applied to searches. Secondly, forward citation searching was conducted using Web of Science for all studies meeting inclusion criteria. Reference lists of all included studies were checked for relevant studies. Finally, international experts, networks on TIC in mental health care, and lived experience networks were contacted to identify additional studies.

### Study selection

All studies identified through database searches were independently title and abstract screened by KS, KT and NVSJ, with 20% double screened. All full texts of potentially relevant studies were double screened independently by KS and KT, with disagreements resolved through discussion. Screening was conducted using Covidence (45). Studies identified through forwards and backwards citation searching and expert recommendation were screened by KS, EMG, and NVSJ.

### Charting and organising the data

A data extraction form based on the research questions and potential outcomes was developed using Microsoft excel and revised collaboratively with the working group. Information on the study design, research and analysis methods, population characteristics, mental health care setting, and TIC approach were extracted alongside data relating to our primary and secondary outcomes. The form was piloted on three included papers and relevant revisions made. Data extraction was completed by KS, EMG, VT, SC, and FA, with over 50% double extracted to check for accuracy by KS and EMG.

### Data synthesis process

Data relevant for each research question was synthesised narratively by KS, EMG, JS, UF, VT, FA, and MS. Question 1 was grouped by approach and reported by setting. Where data was available, evidence for questions 2-6 was synthesised within each TIC approach. We answered the a priori questions set out in our protocol, however we also answered two further questions, based on their relevance for informing policy and implementation strategy: “What are the reported difficulties in implementing trauma informed approaches and interventions?” and “What is required to implement TIC effectively?”

Both quantitative and qualitative data were narratively synthesised together for each question. Areas of heterogeneity were considered throughout this process and highlighted. The categorisation and synthesis of the trauma informed approaches were discussed and validated by KS, EMG, JS, and KT.

## Results

The database search returned 4146 studies from which 2759 potentially relevant full-text studies were identified. Additional search methods identified 96 studies. Overall, 31 studies met inclusion criteria and were included in this review. The PRISMA diagram can be seen in Figure 1. Characteristics of all included studies are shown in Table 1.

**Figure 1:**
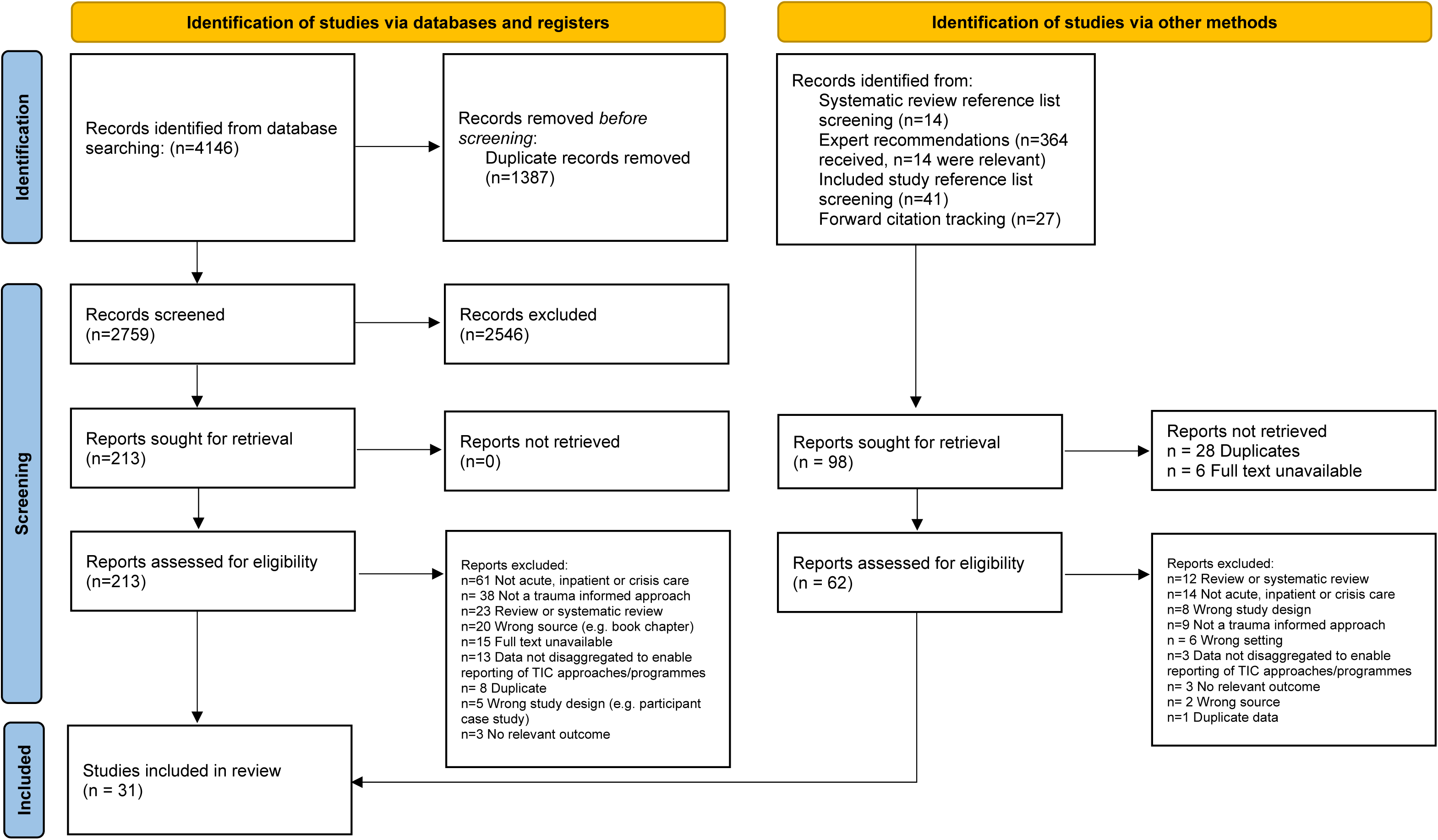
PRISMA diagram demonstrating the search strategy From: Page MJ, McKenzie JE, Bossuyt PM, Boutron I, Hoffmann TC, Mulrow CD, et al. The PRISMA 2020 statement: an updated guideline for reporting systematic reviews. BMJ 2021;372:n71. doi: 10.1136/bmj.n71. For more information, visit: http://www.prisma-statement.org/

**Table 1.**
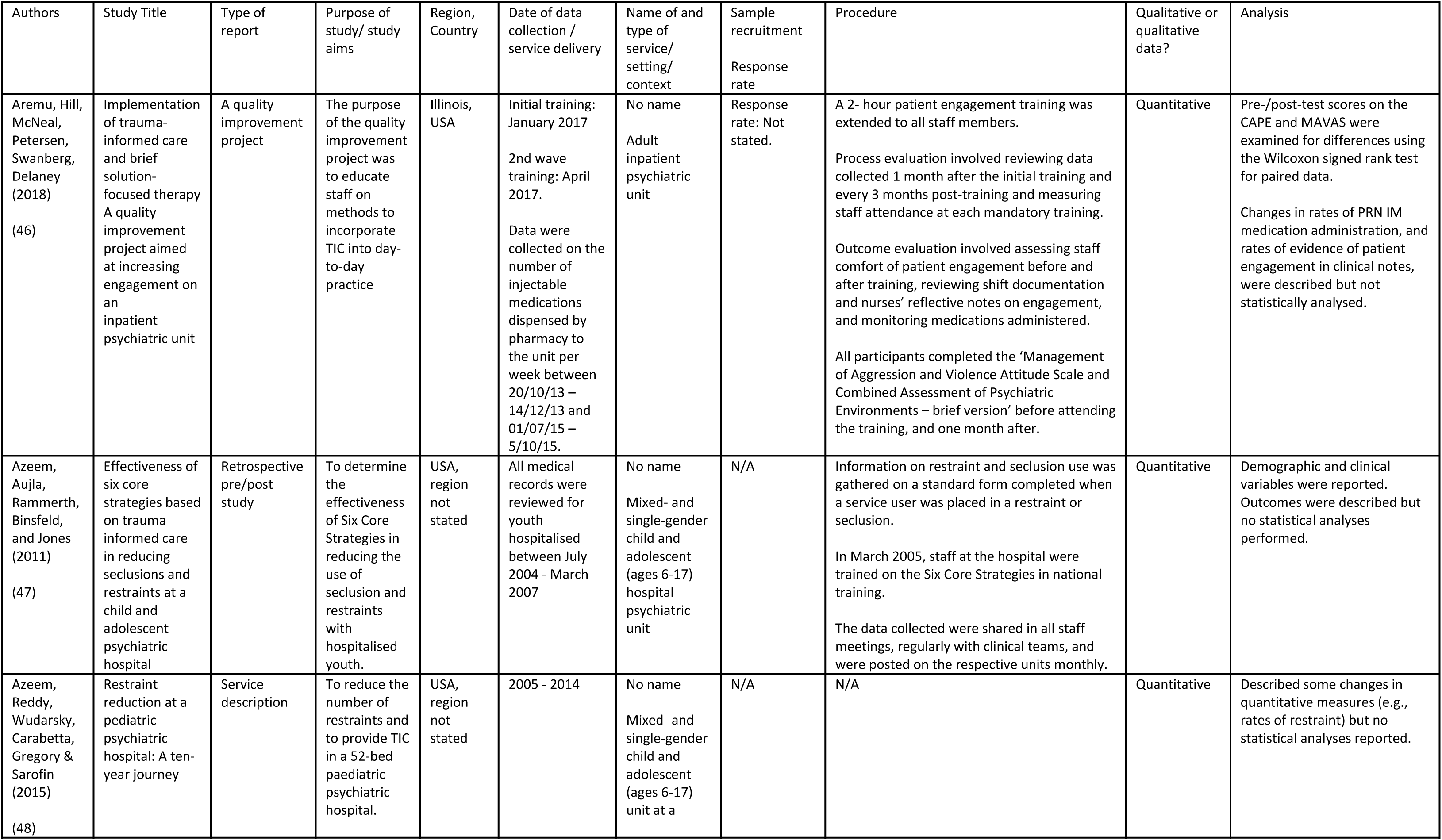
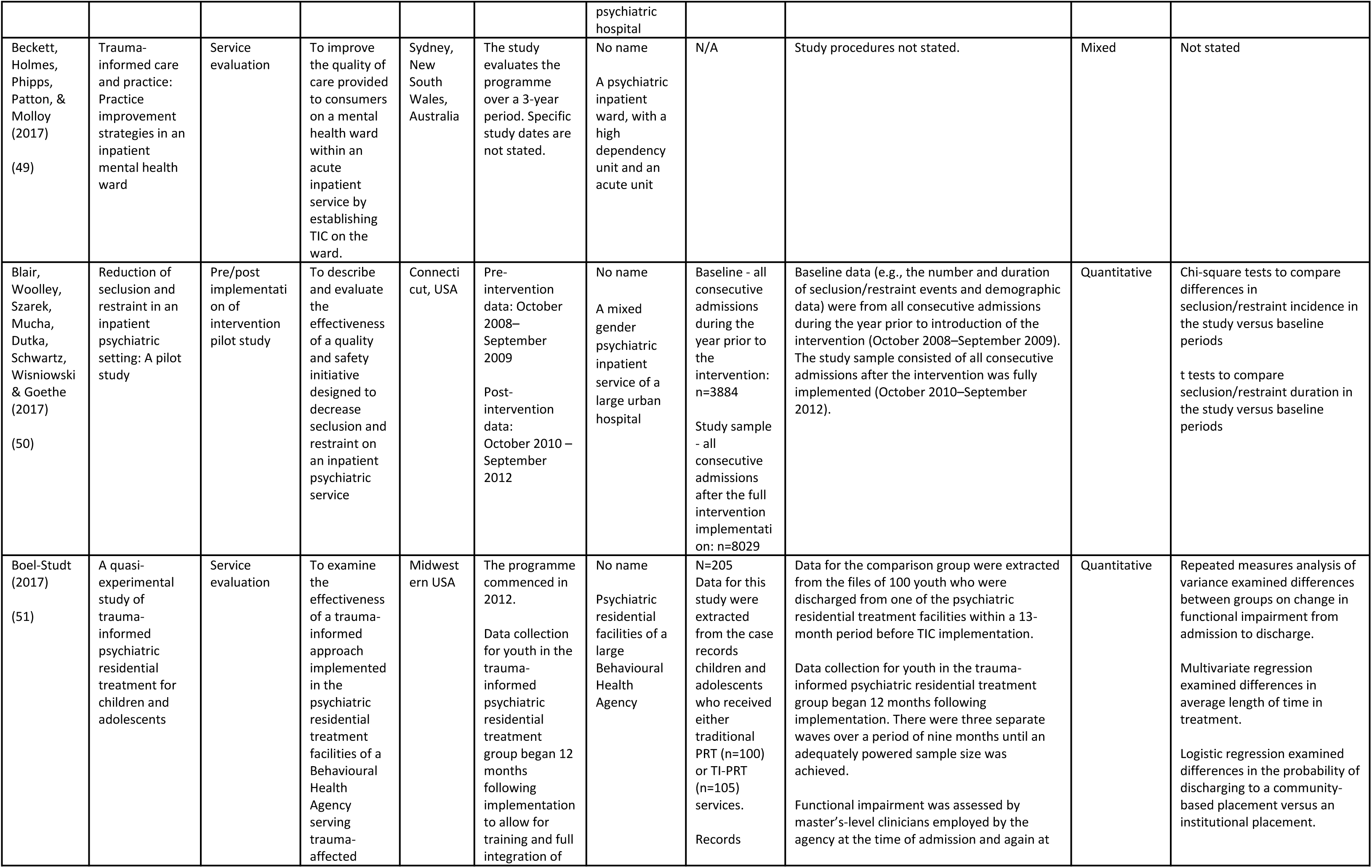
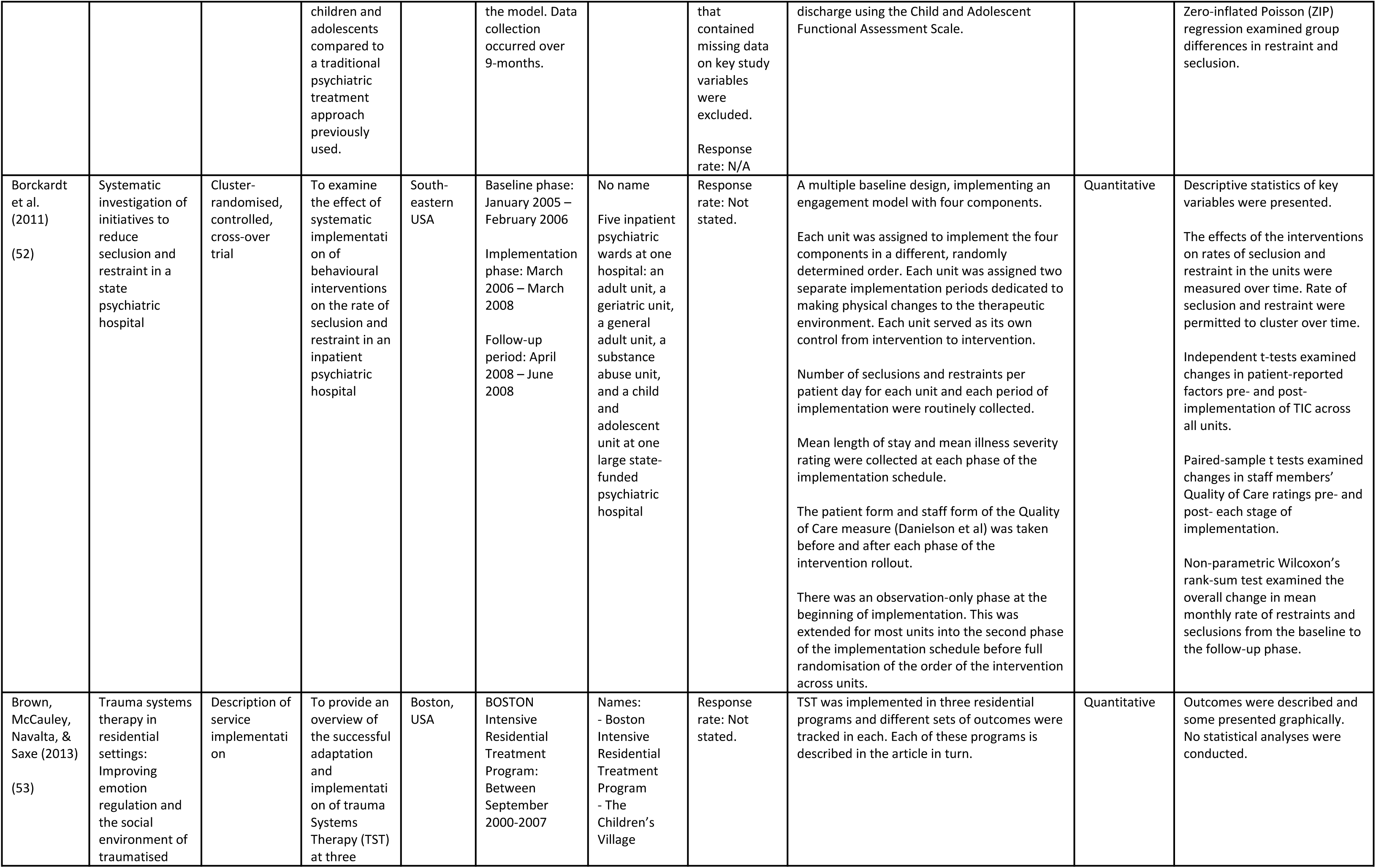
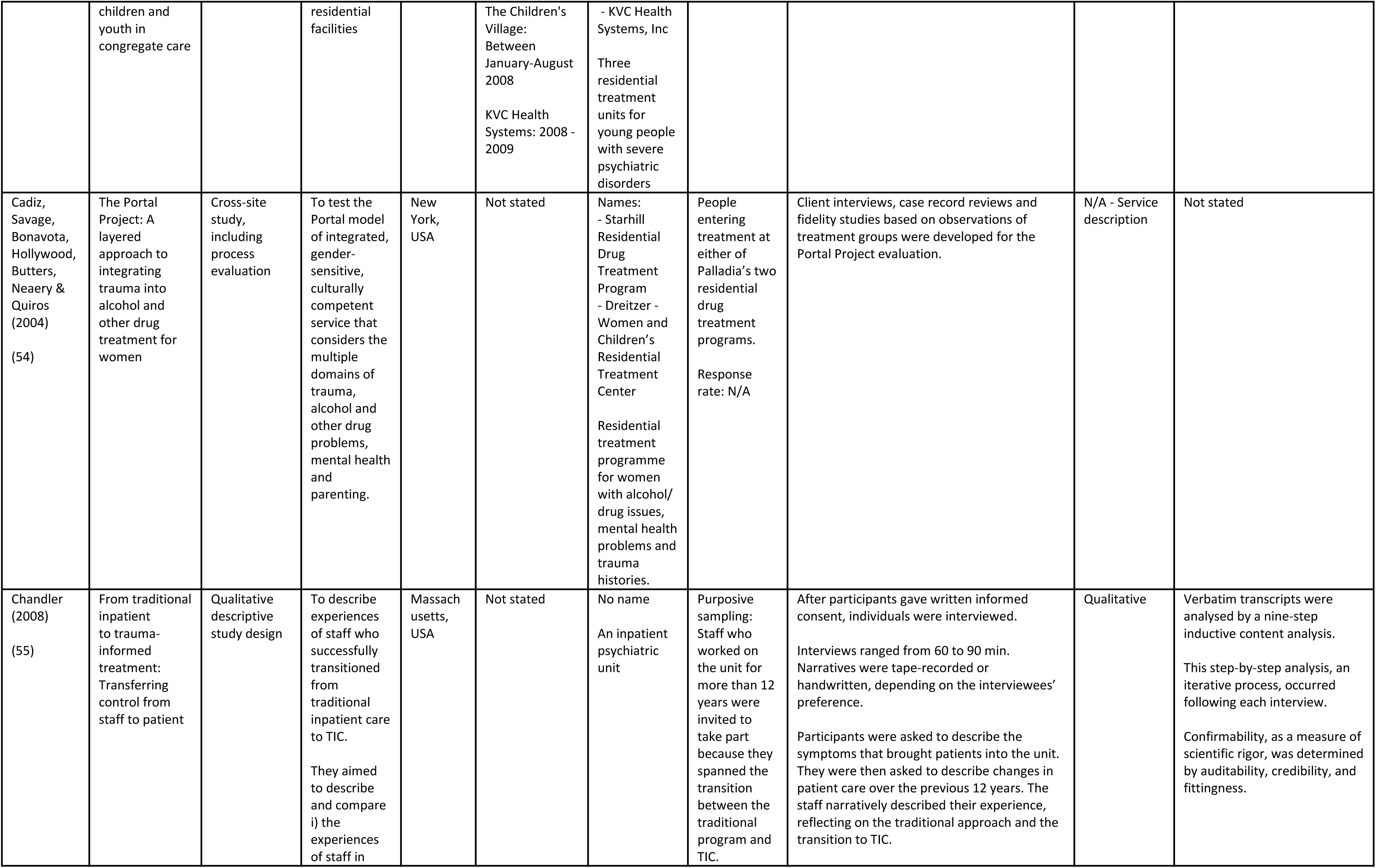
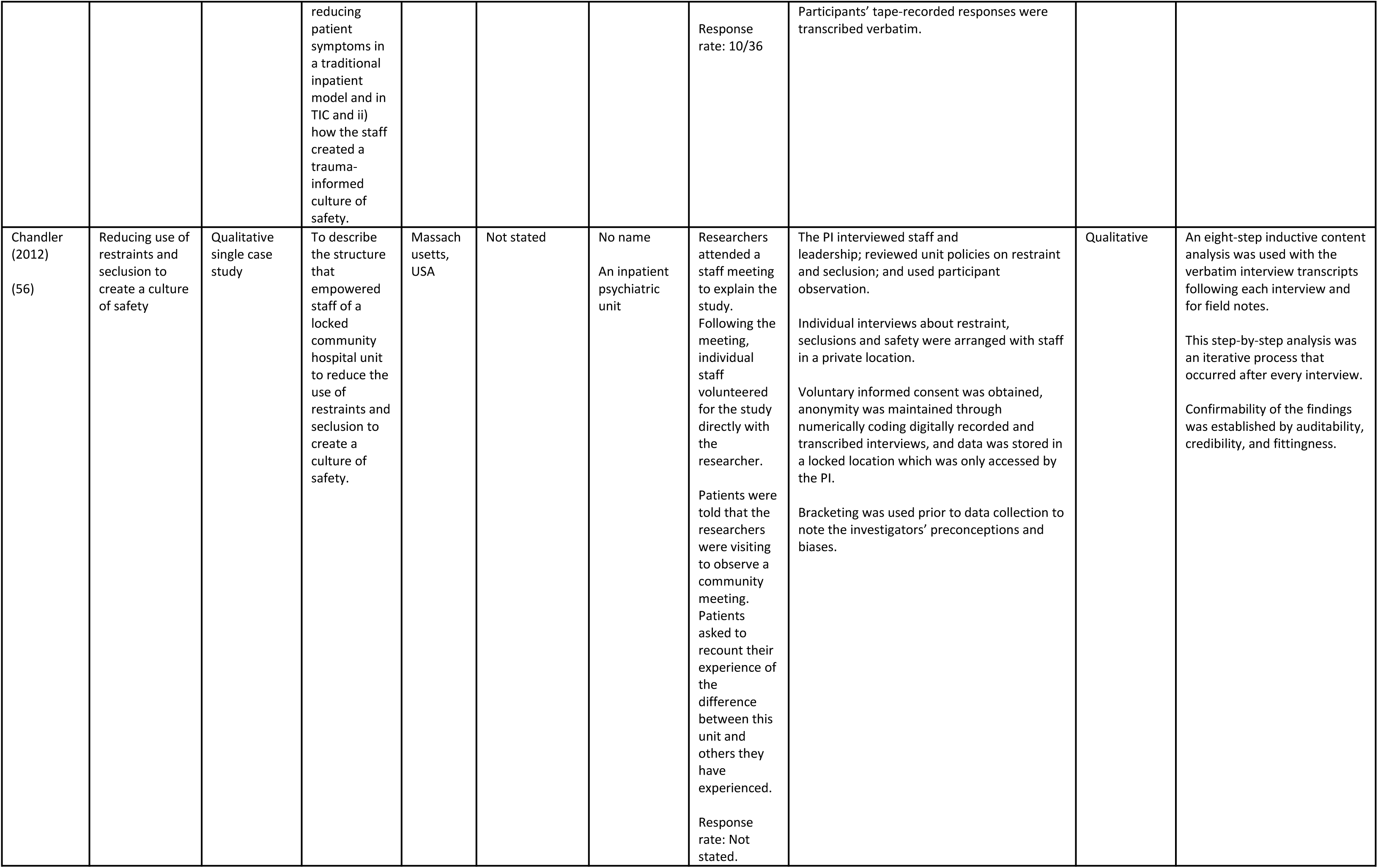
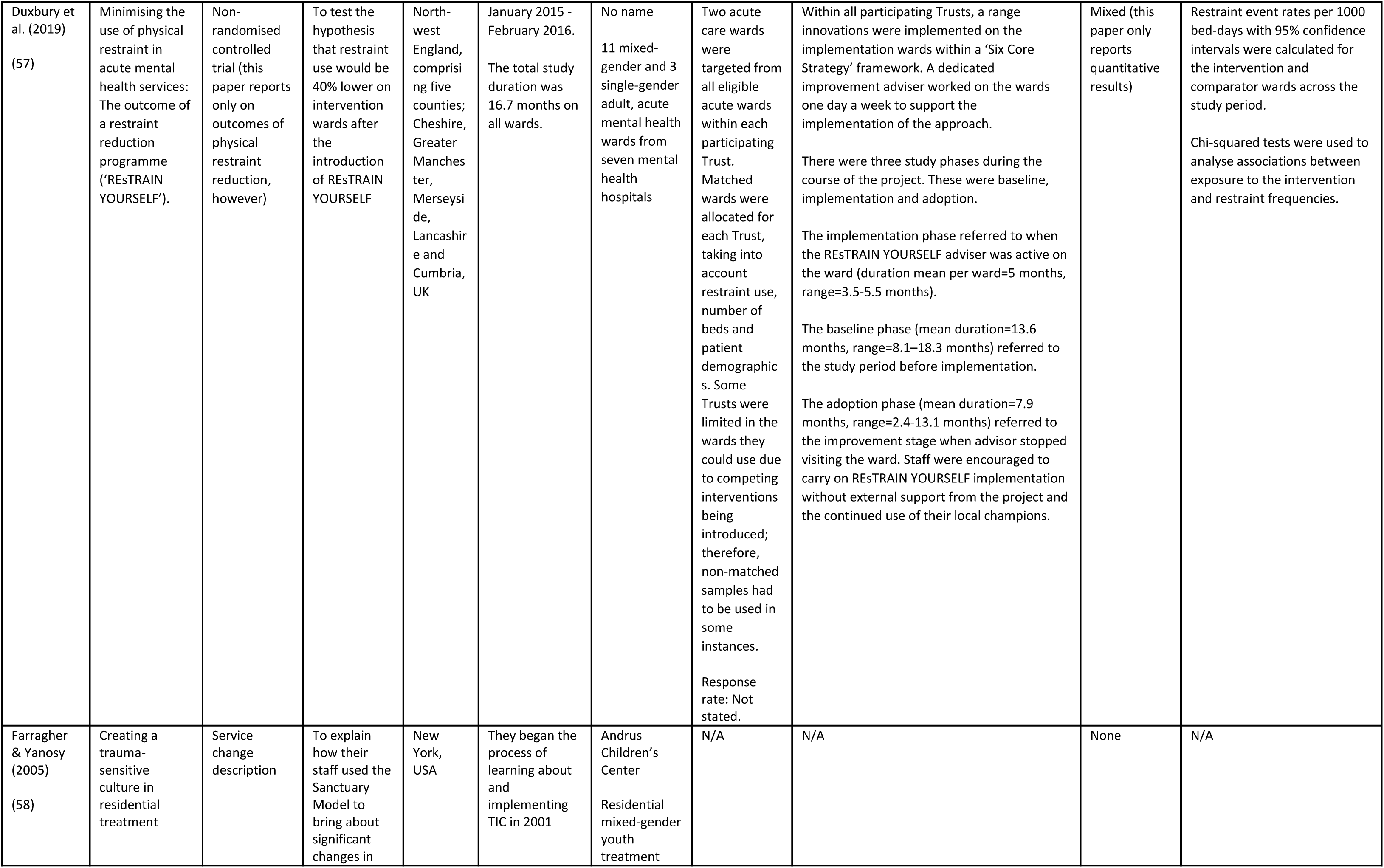
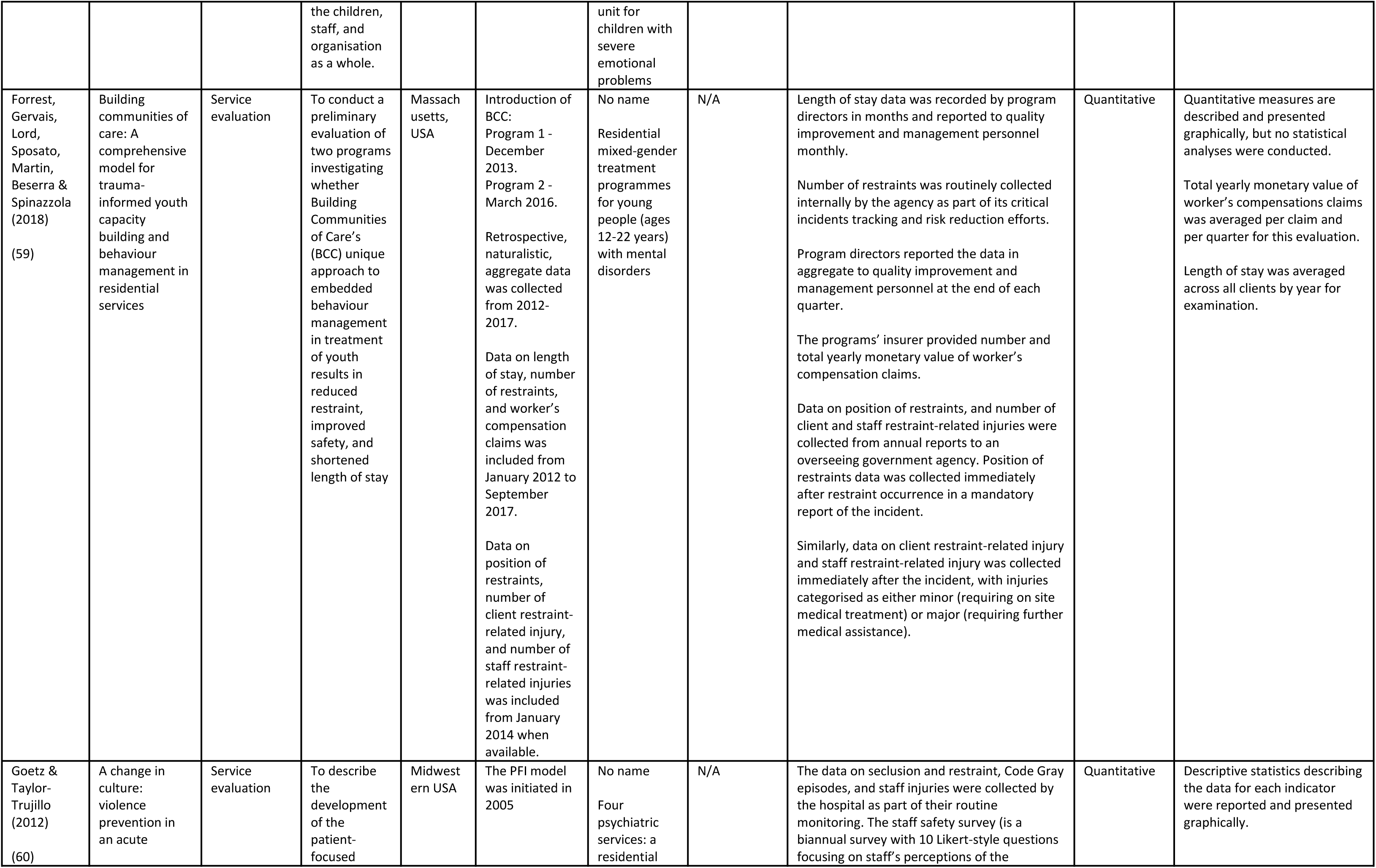
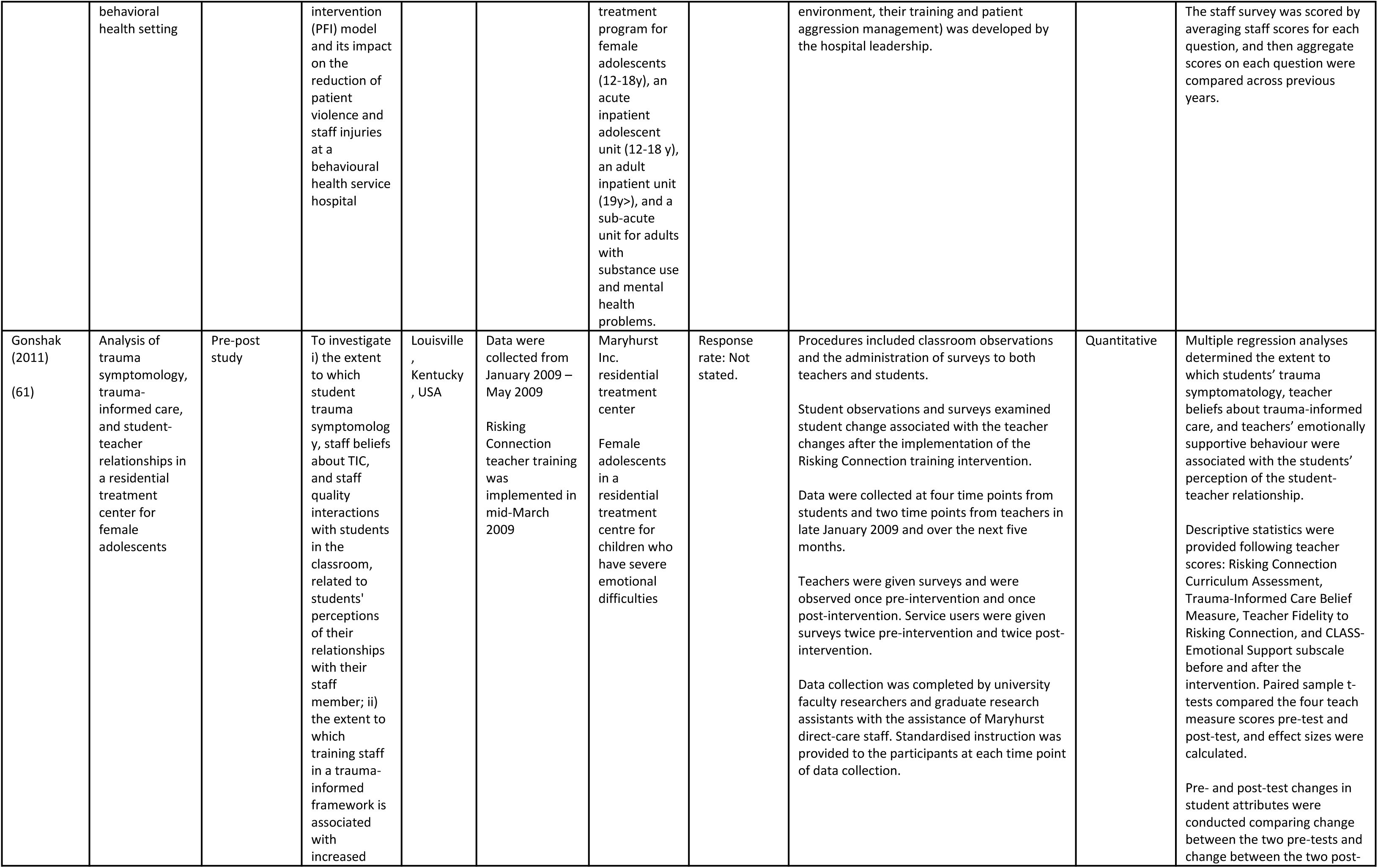
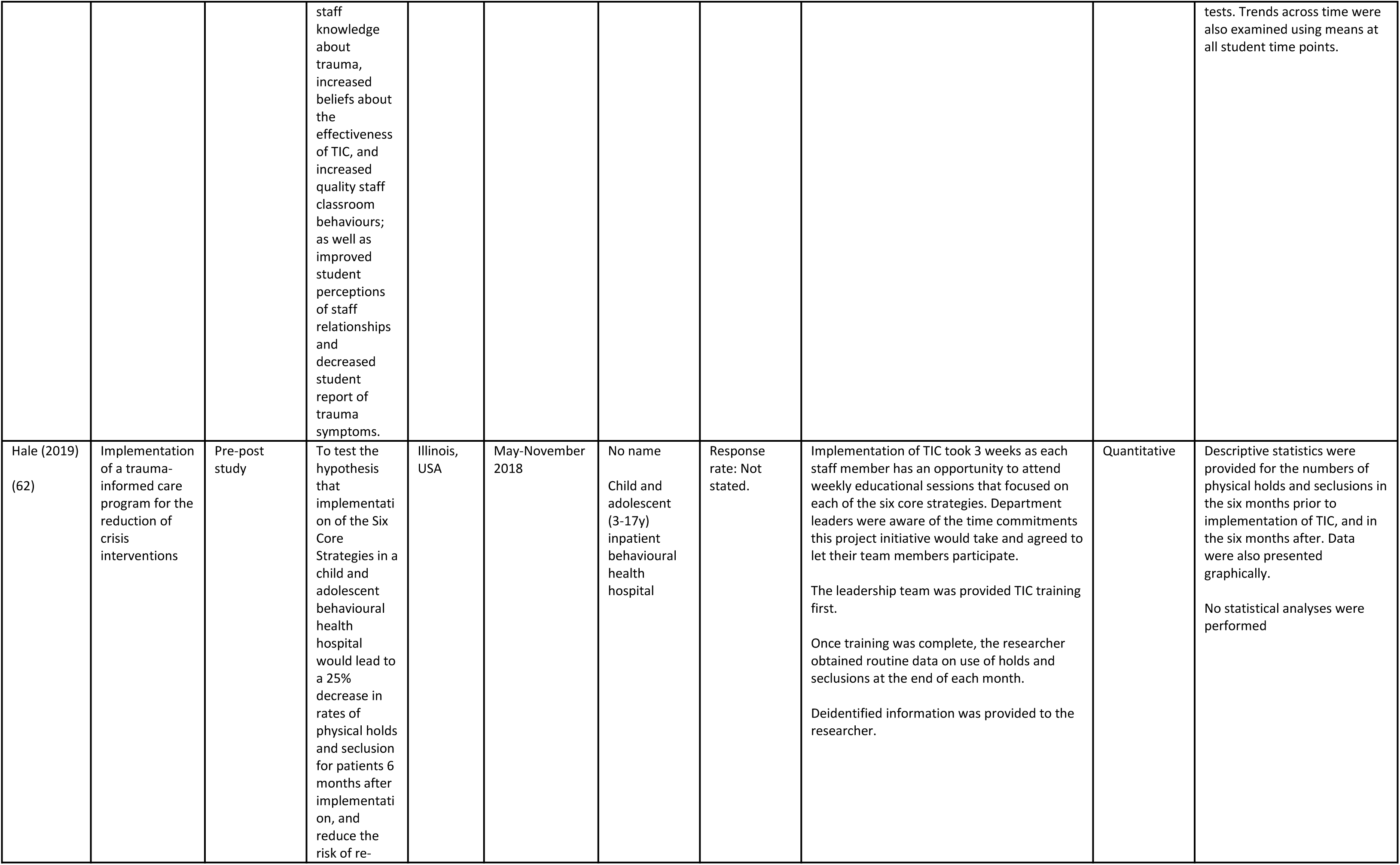
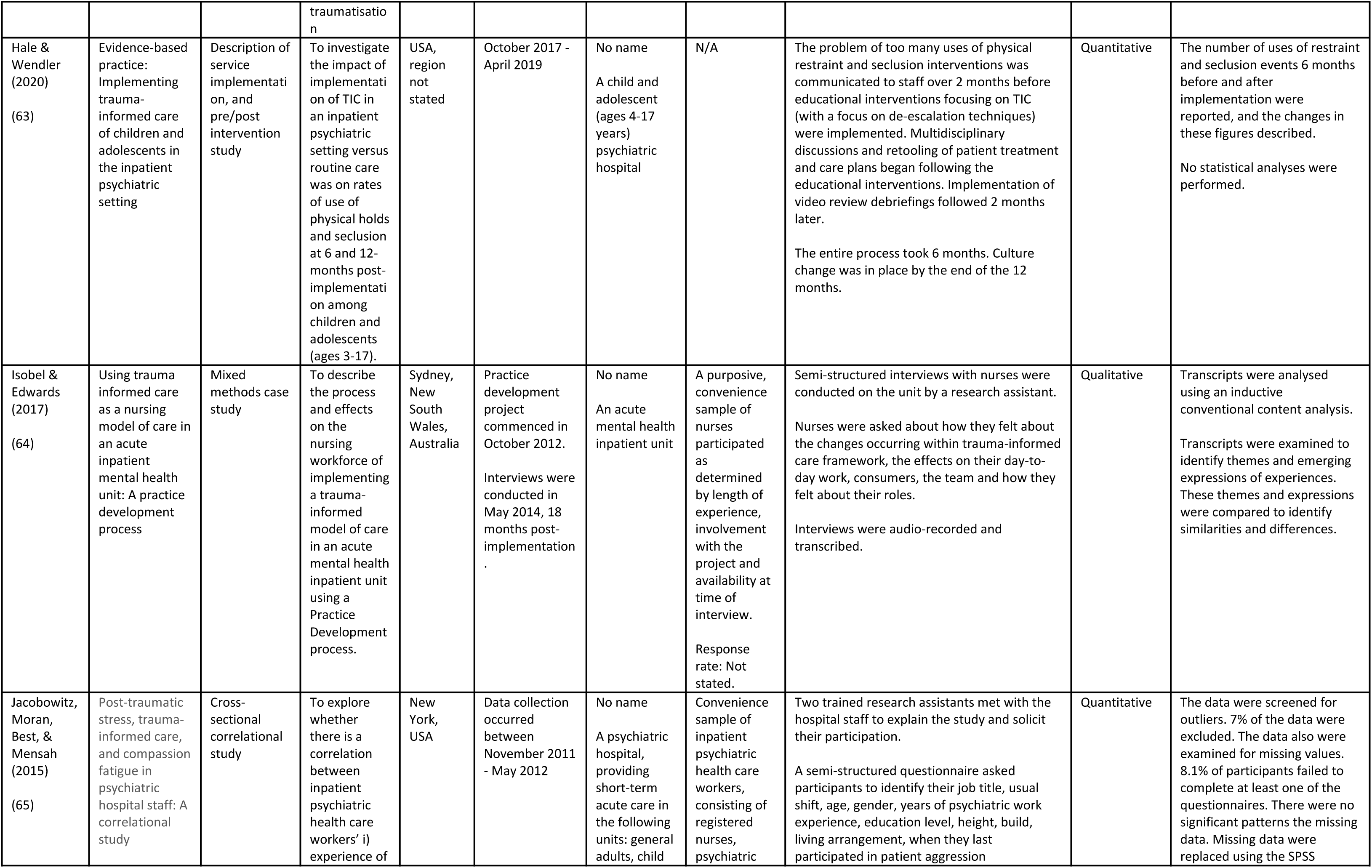
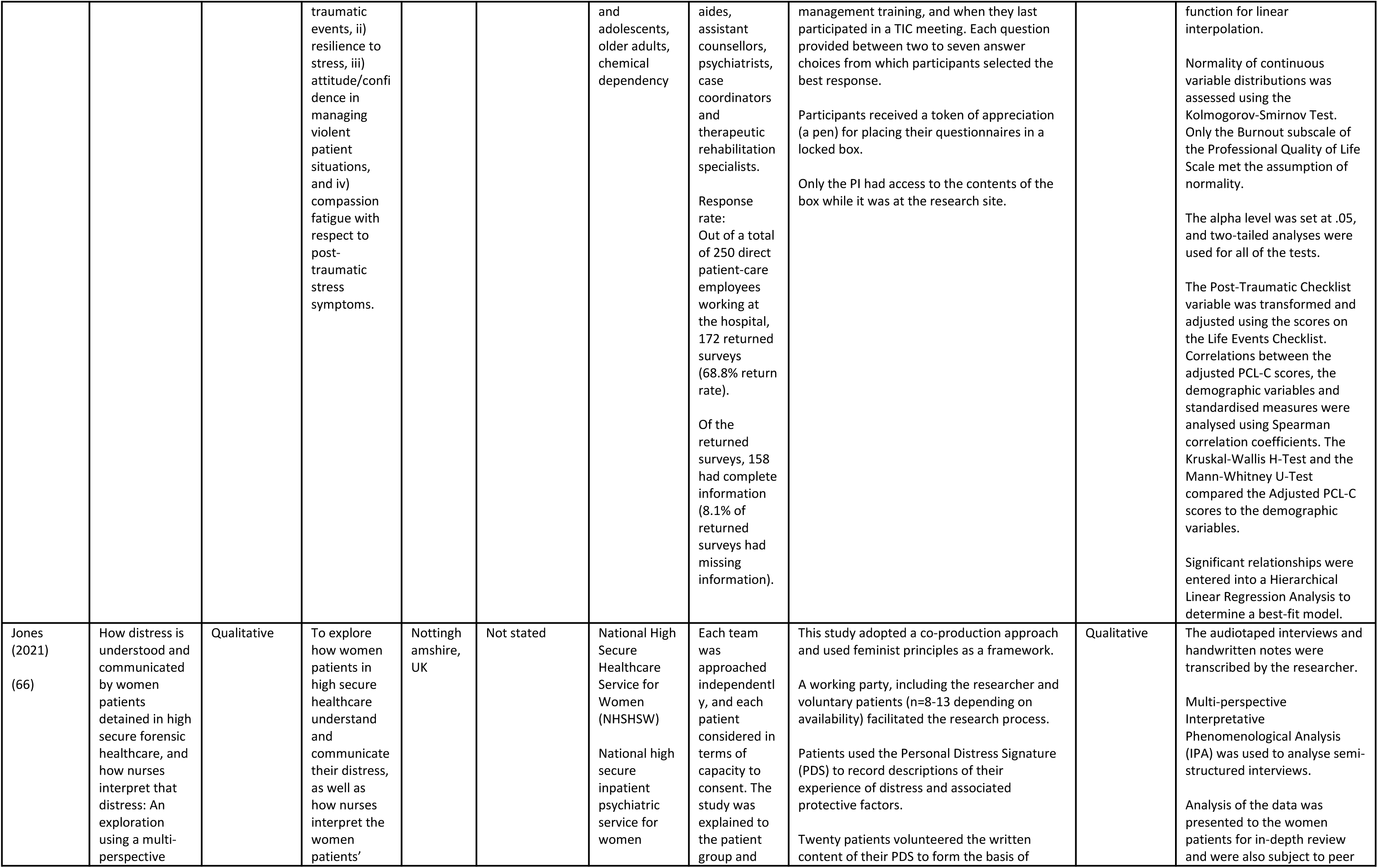
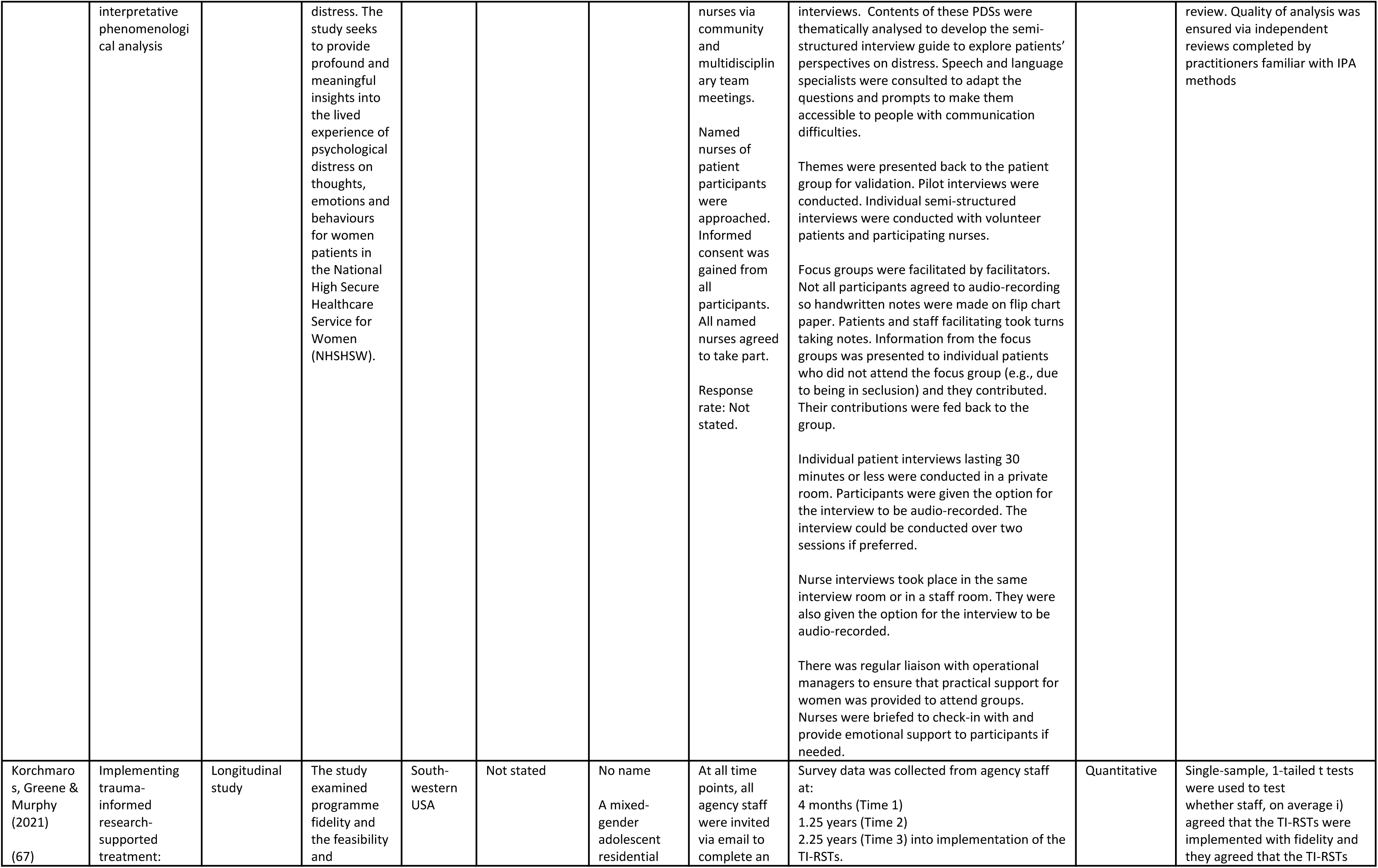
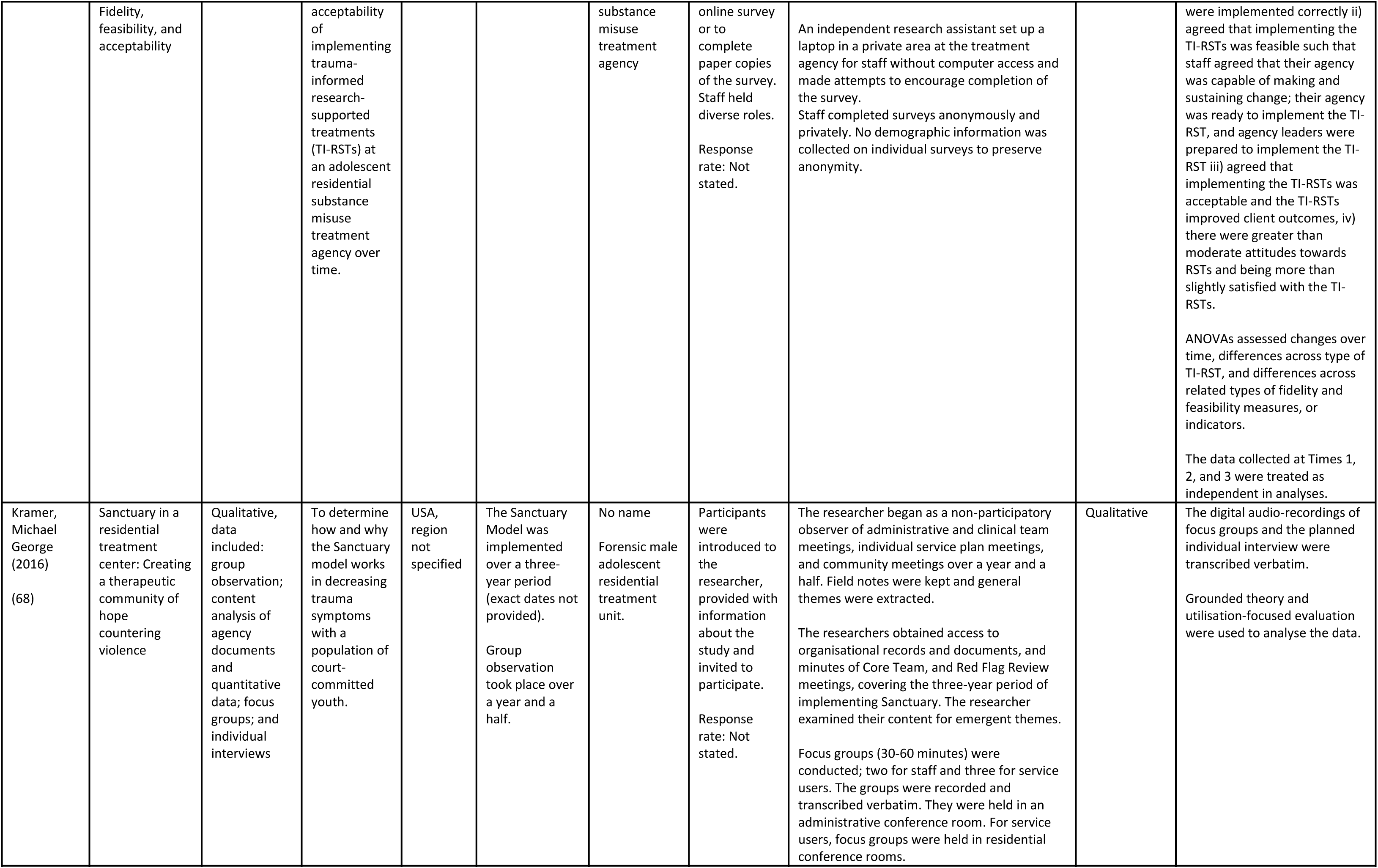
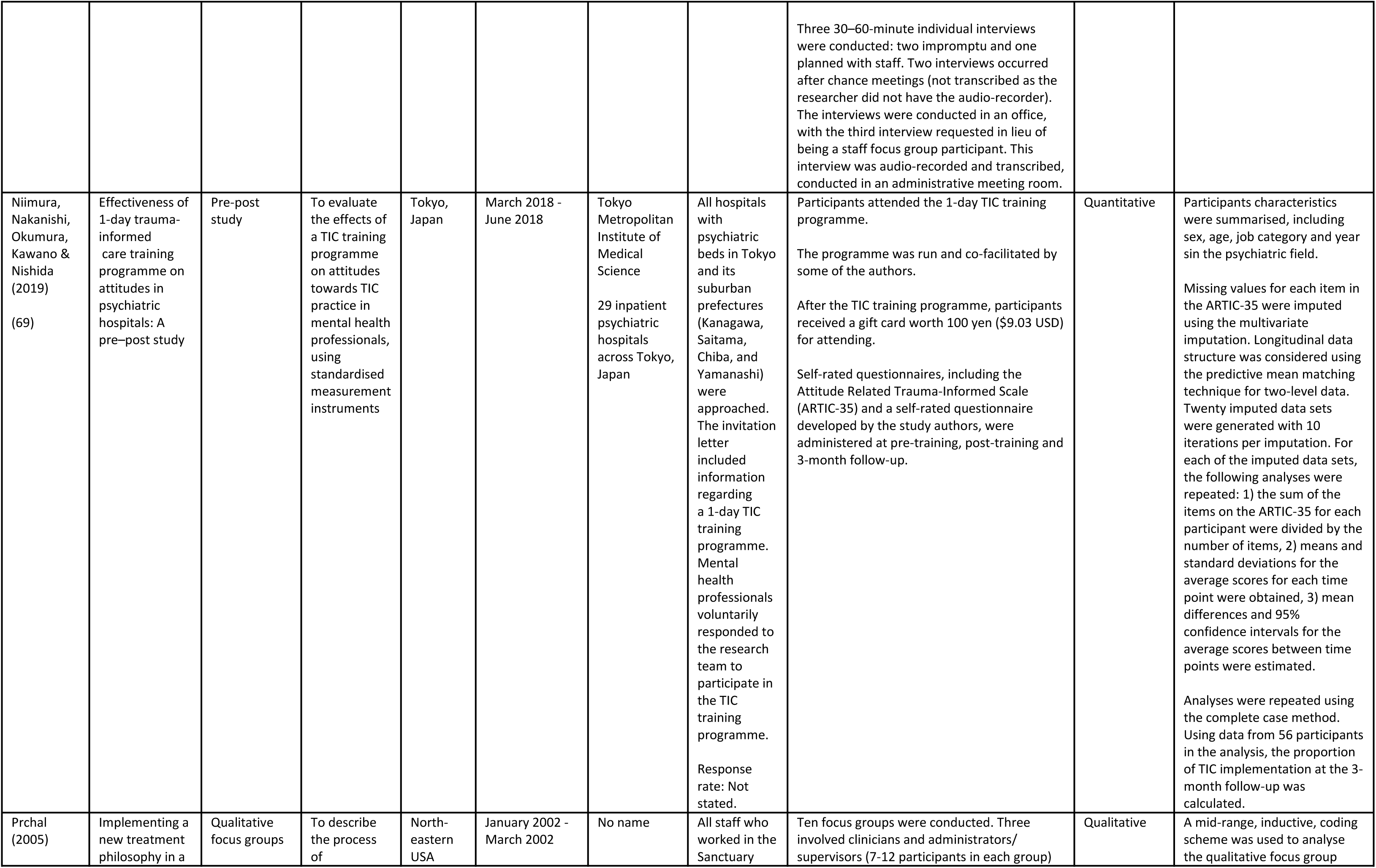
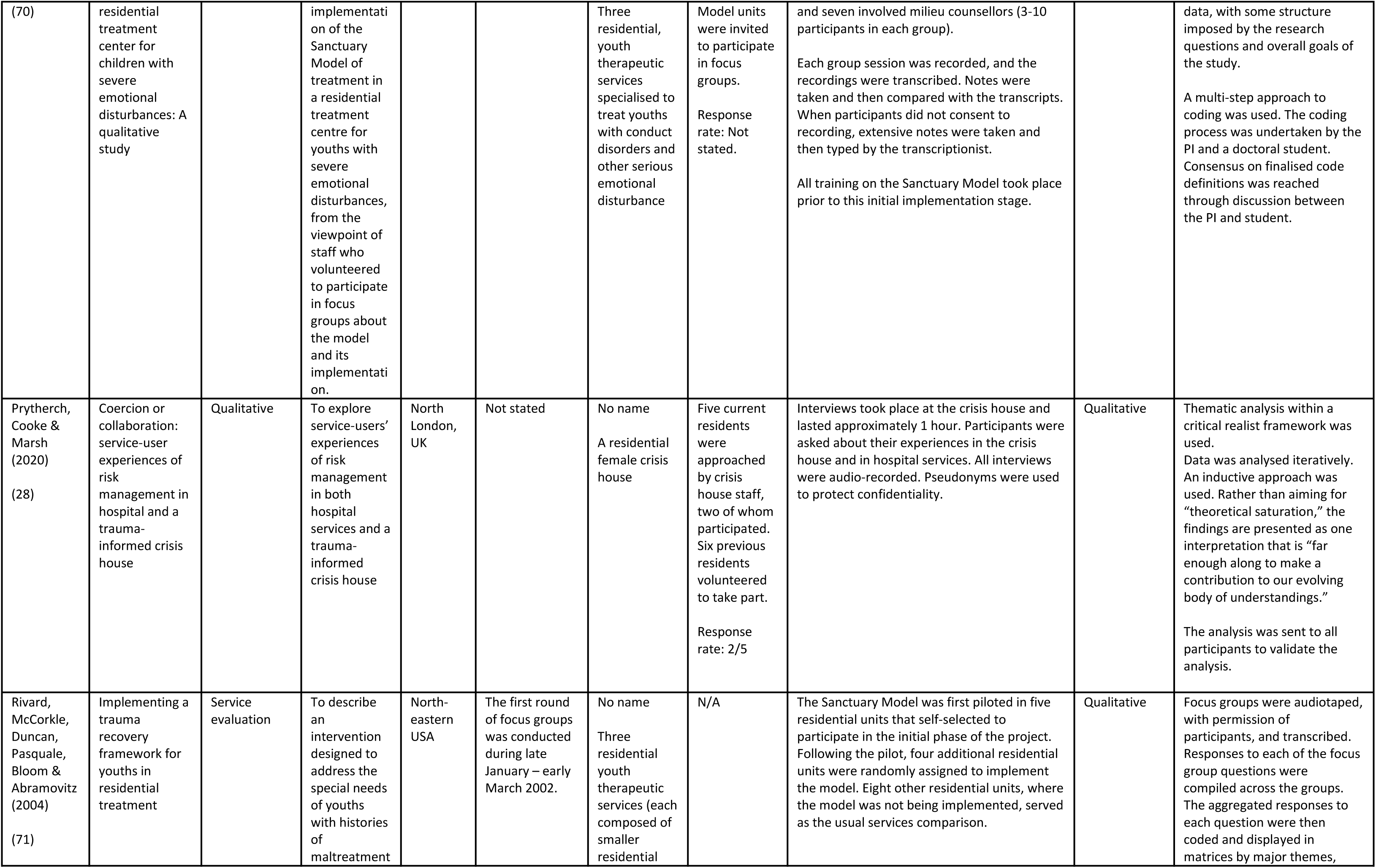
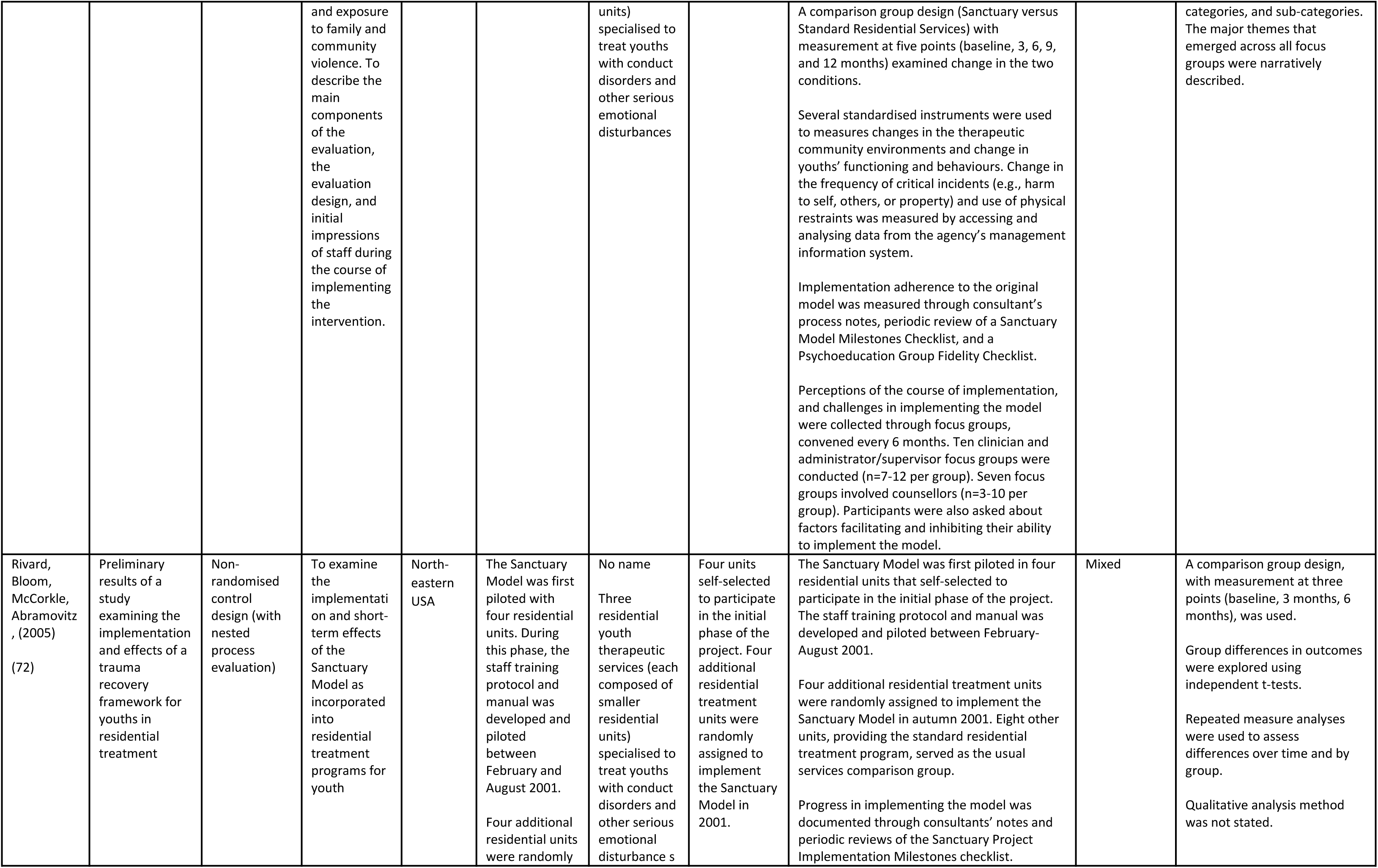
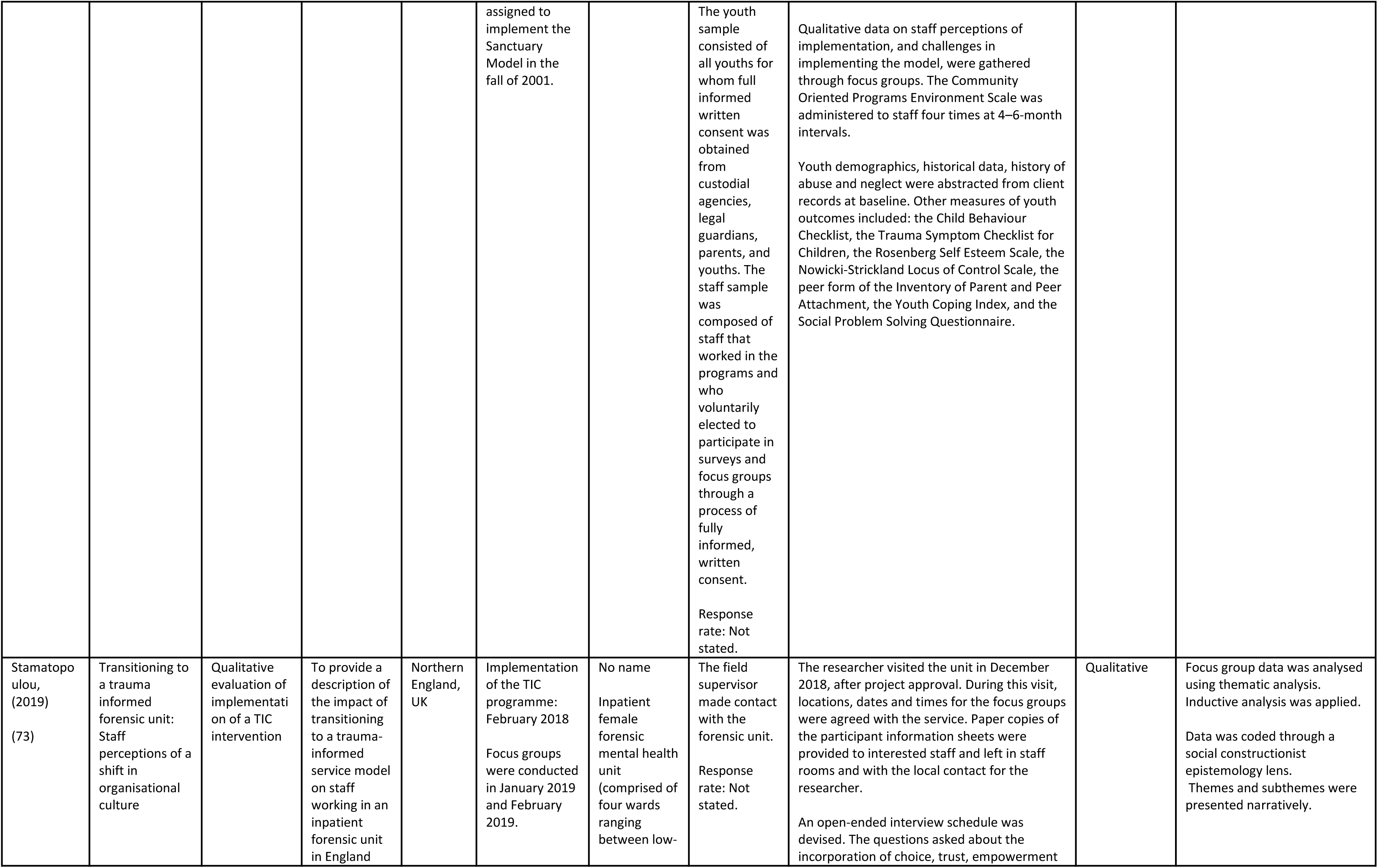
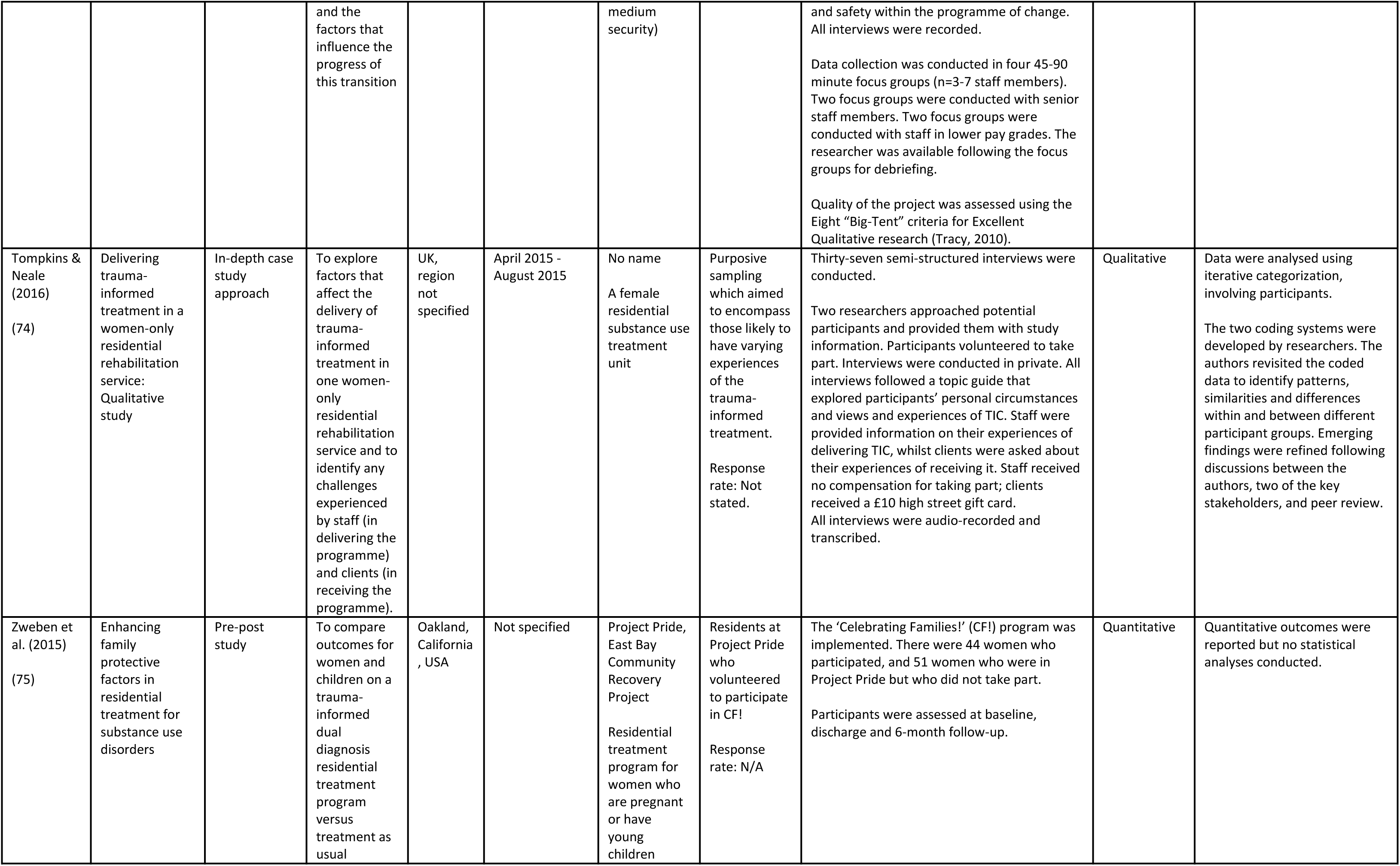
Study characteristics table

| Authors | Study Title | Type of report | Purpose of study/ study aims | Region, Country | Date of data collection / service delivery | Name of and type of service/ setting/ context | Sample recruitment<br>Response rate | Procedure | Qualitative or qualitative data? | Analysis |
| --- | --- | --- | --- | --- | --- | --- | --- | --- | --- | --- |
| Aremu, Hill, McNeal, Petersen, Swanberg, Delaney (2018)<br>(46) | Implementation of trauma-informed care and brief solution-focused therapy<br>A quality improvement project aimed at increasing engagement on an inpatient psychiatric unit | A quality improvement project | The purpose of the quality improvement project was to educate staff on methods to incorporate TIC into day-to-day practice | Illinois, USA | Initial training: January 2017<br>2nd wave training: April 2017.<br>Data were collected on the number of injectable medications dispensed by pharmacy to the unit per week between 20/10/13 – 14/12/13 and 01/07/15 – 5/10/15. | No name<br>Adult inpatient psychiatric unit | Response rate: Not stated. | A 2- hour patient engagement training was extended to all staff members.<br>Process evaluation involved reviewing data collected 1 month after the initial training and every 3 months post-training and measuring staff attendance at each mandatory training.<br>Outcome evaluation involved assessing staff comfort of patient engagement before and after training, reviewing shift documentation and nurses' reflective notes on engagement, and monitoring medications administered.<br>All participants completed the 'Management of Aggression and Violence Attitude Scale and Combined Assessment of Psychiatric Environments – brief version' before attending the training, and one month after. | Quantitative | Pre-/post-test scores on the CAPE and MAVAS were examined for differences using the Wilcoxon signed rank test for paired data.<br>Changes in rates of PRN IM medication administration, and rates of evidence of patient engagement in clinical notes, were described but not statistically analysed. |
| Azeem, Aujla, Rammerth, Binsfeld, and Jones (2011)<br>(47) | Effectiveness of six core strategies based on trauma informed care in reducing seclusions and restraints at a child and adolescent psychiatric hospital | Retrospective pre/post study | To determine the effectiveness of Six Core Strategies in reducing the use of seclusion and restraints with hospitalised youth. | USA, region not stated | All medical records were reviewed for youth hospitalised between July 2004 - March 2007 | No name<br>Mixed- and single-gender child and adolescent (ages 6-17) hospital psychiatric unit | N/A | Information on restraint and seclusion use was gathered on a standard form completed when a service user was placed in a restraint or seclusion.<br>In March 2005, staff at the hospital were trained on the Six Core Strategies in national training.<br>The data collected were shared in all staff meetings, regularly with clinical teams, and were posted on the respective units monthly. | Quantitative | Demographic and clinical variables were reported. Outcomes were described but no statistical analyses performed. |
| Azeem, Reddy, Wudarsky, Carabetta, Gregory & Sarofin (2015)<br>(48) | Restraint reduction at a pediatric psychiatric hospital: A ten-year journey | Service description | To reduce the number of restraints and to provide TIC in a 52-bed paediatric psychiatric hospital. | USA, region not stated | 2005 - 2014 | No name<br>Mixed- and single-gender child and adolescent (ages 6-17) unit at a psychiatric hospital | N/A | N/A | Quantitative | Described some changes in quantitative measures (e.g., rates of restraint) but no statistical analyses reported. |
| Beckett, Holmes, Phipps, Patton, & Molloy (2017)<br>(49) | Trauma-informed care and practice: Practice improvement strategies in an inpatient mental health ward | Service evaluation | To improve the quality of care provided to consumers on a mental health ward within an acute inpatient service by establishing TIC on the ward. | Sydney, New South Wales, Australia | The study evaluates the programme over a 3-year period. Specific study dates are not stated. | No name<br>A psychiatric inpatient ward, with a high dependency unit and an acute unit | N/A | Study procedures not stated. | Mixed | Not stated |
| Blair, Woolley, Szarek, Mucha, Dutka, Schwartz, Wisniowski & Goethe (2017)<br>(50) | Reduction of seclusion and restraint in an inpatient psychiatric setting: A pilot study | Pre/post implementation of intervention pilot study | To describe and evaluate the effectiveness of a quality and safety initiative designed to decrease seclusion and restraint on an inpatient psychiatric service | Connecticut, USA | Pre-intervention data: October 2008–September 2009<br>Post-intervention data: October 2010 – September 2012 | No name<br>A mixed gender psychiatric inpatient service of a large urban hospital | Baseline - all consecutive admissions during the year prior to the intervention: n=3884<br>Study sample - all consecutive admissions after the full intervention implementation: n=8029 | Baseline data (e.g., the number and duration of seclusion/restraint events and demographic data) were from all consecutive admissions during the year prior to introduction of the intervention (October 2008–September 2009). The study sample consisted of all consecutive admissions after the intervention was fully implemented (October 2010–September 2012). | Quantitative | Chi-square tests to compare differences in seclusion/restraint incidence in the study versus baseline periods<br>t tests to compare seclusion/restraint duration in the study versus baseline periods |
| Boel-Studt (2017)<br>(51) | A quasi-experimental study of trauma-informed psychiatric residential treatment for children and adolescents | Service evaluation | To examine the effectiveness of a trauma-informed approach implemented in the psychiatric residential treatment facilities of a Behavioural Health Agency serving trauma-affected | Midwestern USA | The programme commenced in 2012.<br>Data collection for youth in the trauma-informed psychiatric residential treatment group began 12 months following implementation to allow for training and full integration of | No name<br>Psychiatric residential facilities of a large Behavioural Health Agency | N=205<br>Data for this study were extracted from the case records of children and adolescents who received either traditional PRT (n=100) or TI-PRT (n=105) services.<br>Records | Data for the comparison group were extracted from the files of 100 youth who were discharged from one of the psychiatric residential treatment facilities within a 13-month period before TIC implementation.<br>Data collection for youth in the trauma-informed psychiatric residential treatment group began 12 months following implementation. There were three separate waves over a period of nine months until an adequately powered sample size was achieved.<br>Functional impairment was assessed by master's-level clinicians employed by the agency at the time of admission and again at | Quantitative | Repeated measures analysis of variance examined differences between groups on change in functional impairment from admission to discharge.<br>Multivariate regression examined differences in average length of time in treatment.<br>Logistic regression examined differences in the probability of discharging to a community-based placement versus an institutional placement. |
|  |  |  | children and adolescents compared to a traditional psychiatric treatment approach previously used. |  | the model. Data collection occurred over 9-months. |  | that contained missing data on key study variables were excluded.<br>Response rate: N/A | discharge using the Child and Adolescent Functional Assessment Scale. |  | Zero-inflated Poisson (ZIP) regression examined group differences in restraint and seclusion. |
| Borckardt et al. (2011) (52) | Systematic investigation of initiatives to reduce seclusion and restraint in a state psychiatric hospital | Cluster-randomised, controlled, cross-over trial | To examine the effect of systematic implementation of behavioural interventions on the rate of seclusion and restraint in an inpatient psychiatric hospital | South-eastern USA | Baseline phase: January 2005 – February 2006<br>Implementation phase: March 2006 – March 2008<br>Follow-up period: April 2008 – June 2008 | No name<br>Five inpatient psychiatric wards at one hospital: an adult unit, a geriatric unit, a general adult unit, a substance abuse unit, and a child and adolescent unit at one large state-funded psychiatric hospital | Response rate: Not stated. | A multiple baseline design, implementing an engagement model with four components.<br>Each unit was assigned to implement the four components in a different, randomly determined order. Each unit was assigned two separate implementation periods dedicated to making physical changes to the therapeutic environment. Each unit served as its own control from intervention to intervention.<br>Number of seclusions and restraints per patient day for each unit and each period of implementation were routinely collected.<br>Mean length of stay and mean illness severity rating were collected at each phase of the implementation schedule.<br>The patient form and staff form of the Quality of Care measure (Danielson et al) was taken before and after each phase of the intervention rollout.<br>There was an observation-only phase at the beginning of implementation. This was extended for most units into the second phase of the implementation schedule before full randomisation of the order of the intervention across units. | Quantitative | Descriptive statistics of key variables were presented.<br>The effects of the interventions on rates of seclusion and restraint in the units were measured over time. Rate of seclusion and restraint were permitted to cluster over time.<br>Independent t-tests examined changes in patient-reported factors pre- and post-implementation of TIC across all units.<br>Paired-sample t tests examined changes in staff members' Quality of Care ratings pre- and post- each stage of implementation.<br>Non-parametric Wilcoxon's rank-sum test examined the overall change in mean monthly rate of restraints and seclusions from the baseline to the follow-up phase. |
| Brown, McCauley, Navalta, & Saxe (2013) (53) | Trauma systems therapy in residential settings: Improving emotion regulation and the social environment of traumatised | Description of service implementation | To provide an overview of the successful adaptation and implementation of trauma Systems Therapy (TST) at three | Boston, USA | BOSTON Intensive Residential Treatment Program: Between September 2000-2007 | Names:<br>- Boston Intensive Residential Treatment Program<br>- The Children's Village | Response rate: Not stated. | TST was implemented in three residential programs and different sets of outcomes were tracked in each. Each of these programs is described in the article in turn. | Quantitative | Outcomes were described and some presented graphically. No statistical analyses were conducted. |
|  | children and youth in congregate care |  | residential facilities |  | The Children's Village: Between January-August 2008<br>KVC Health Systems: 2008 - 2009 | - KVC Health Systems, Inc<br>Three residential treatment units for young people with severe psychiatric disorders |  |  |  |  |
| Cadiz, Savage, Bonavota, Hollywood, Butters, Neaery & Quiros (2004)<br>(54) | The Portal Project: A layered approach to integrating trauma into alcohol and other drug treatment for women | Cross-site study, including process evaluation | To test the Portal model of integrated, gender-sensitive, culturally competent service that considers the multiple domains of trauma, alcohol and other drug problems, mental health and parenting. | New York, USA | Not stated | Names:<br>- Starhill Residential Drug Treatment Program<br>- Dreitzer - Women and Children's Residential Treatment Center<br>Residential treatment programme for women with alcohol/ drug issues, mental health problems and trauma histories. | People entering treatment at either of Palladia's two residential drug treatment programs.<br>Response rate: N/A | Client interviews, case record reviews and fidelity studies based on observations of treatment groups were developed for the Portal Project evaluation. | N/A - Service description | Not stated |
| Chandler (2008)<br>(55) | From traditional inpatient to trauma-informed treatment: Transferring control from staff to patient | Qualitative descriptive study design | To describe experiences of staff who successfully transitioned from traditional inpatient care to TIC.<br>They aimed to describe and compare i) the experiences of staff in | Massachusetts, USA | Not stated | No name<br>An inpatient psychiatric unit | Purposive sampling: Staff who worked on the unit for more than 12 years were invited to take part because they spanned the transition between the traditional program and TIC. | After participants gave written informed consent, individuals were interviewed.<br>Interviews ranged from 60 to 90 min. Narratives were tape-recorded or handwritten, depending on the interviewees' preference.<br>Participants were asked to describe the symptoms that brought patients into the unit. They were then asked to describe changes in patient care over the previous 12 years. The staff narratively described their experience, reflecting on the traditional approach and the transition to TIC. | Qualitative | Verbatim transcripts were analysed by a nine-step inductive content analysis.<br>This step-by-step analysis, an iterative process, occurred following each interview.<br>Confirmability, as a measure of scientific rigor, was determined by auditability, credibility, and fittingness. |
|  |  |  | reducing patient symptoms in a traditional inpatient model and in TIC and ii) how the staff created a trauma-informed culture of safety. |  |  |  | Response rate: 10/36 | Participants' tape-recorded responses were transcribed verbatim. |  |  |
| Chandler (2012)<br>(56) | Reducing use of restraints and seclusion to create a culture of safety | Qualitative single case study | To describe the structure that empowered staff of a locked community hospital unit to reduce the use of restraints and seclusion to create a culture of safety. | Massachusetts, USA | Not stated | No name<br>An inpatient psychiatric unit | Researchers attended a staff meeting to explain the study. Following the meeting, individual staff volunteered for the study directly with the researcher.<br>Patients were told that the researchers were visiting to observe a community meeting. Patients asked to recount their experience of the difference between this unit and others they have experienced.<br>Response rate: Not stated. | The PI interviewed staff and leadership; reviewed unit policies on restraint and seclusion; and used participant observation.<br>Individual interviews about restraint, seclusions and safety were arranged with staff in a private location.<br>Voluntary informed consent was obtained, anonymity was maintained through numerically coding digitally recorded and transcribed interviews, and data was stored in a locked location which was only accessed by the PI.<br>Bracketing was used prior to data collection to note the investigators' preconceptions and biases. | Qualitative | An eight-step inductive content analysis was used with the verbatim interview transcripts following each interview and for field notes.<br>This step-by-step analysis was an iterative process that occurred after every interview.<br>Confirmability of the findings was established by auditability, credibility, and fittingness. |
| Duxbury et al. (2019)<br>(57) | Minimising the use of physical restraint in acute mental health services: The outcome of a restraint reduction programme ('REsTRAIN YOURSELF'). | Non-randomised controlled trial (this paper reports only on outcomes of physical restraint reduction, however) | To test the hypothesis that restraint use would be 40% lower on intervention wards after the introduction of REsTRAIN YOURSELF | North-west England, comprising five counties; Cheshire, Greater Manchester, Merseyside, Lancashire and Cumbria, UK | January 2015 - February 2016.<br>The total study duration was 16.7 months on all wards. | No name<br>11 mixed-gender and 3 single-gender adult, acute mental health wards from seven mental health hospitals | Two acute care wards were targeted from all eligible acute wards within each participating Trust. Matched wards were allocated for each Trust, taking into account restraint use, number of beds and patient demographics. Some Trusts were limited in the wards they could use due to competing interventions being introduced; therefore, non-matched samples had to be used in some instances.<br>Response rate: Not stated. | Within all participating Trusts, a range of innovations were implemented on the implementation wards within a 'Six Core Strategy' framework. A dedicated improvement adviser worked on the wards one day a week to support the implementation of the approach.<br>There were three study phases during the course of the project. These were baseline, implementation and adoption.<br>The implementation phase referred to when the REsTRAIN YOURSELF adviser was active on the ward (duration mean per ward=5 months, range=3.5-5.5 months).<br>The baseline phase (mean duration=13.6 months, range=8.1–18.3 months) referred to the study period before implementation.<br>The adoption phase (mean duration=7.9 months, range=2.4-13.1 months) referred to the improvement stage when the adviser stopped visiting the ward. Staff were encouraged to carry on REsTRAIN YOURSELF implementation without external support from the project and the continued use of their local champions. | Mixed (this paper only reports quantitative results) | Restraint event rates per 1000 bed-days with 95% confidence intervals were calculated for the intervention and comparator wards across the study period.<br>Chi-squared tests were used to analyse associations between exposure to the intervention and restraint frequencies. |
| Farragher & Yanosy (2005)<br>(58) | Creating a trauma-sensitive culture in residential treatment | Service change description | To explain how their staff used the Sanctuary Model to bring about significant changes in the children, staff, and organisation as a whole. | New York, USA | They began the process of learning about and implementing TIC in 2001 | Andrus Children's Center<br>Residential mixed-gender youth treatment unit for children with severe emotional problems | N/A | N/A | None | N/A |
| Forrest, Gervais, Lord, Sposato, Martin, Beserra & Spinazzola (2018)<br>(59) | Building communities of care: A comprehensive model for trauma-informed youth capacity building and behaviour management in residential services | Service evaluation | To conduct a preliminary evaluation of two programs investigating whether Building Communities of Care's (BCC) unique approach to embedded behaviour management in treatment of youth results in reduced restraint, improved safety, and shortened length of stay | Massachusetts, USA | Introduction of BCC:<br>Program 1 - December 2013.<br>Program 2 - March 2016.<br>Retrospective, naturalistic, aggregate data was collected from 2012-2017.<br>Data on length of stay, number of restraints, and worker's compensation claims was included from January 2012 to September 2017.<br>Data on position of restraints, number of client restraint-related injury, and number of staff restraint-related injuries was included from January 2014 when available. | No name<br>Residential mixed-gender treatment programmes for young people (ages 12-22 years) with mental disorders | N/A | Length of stay data was recorded by program directors in months and reported to quality improvement and management personnel monthly.<br>Number of restraints was routinely collected internally by the agency as part of its critical incidents tracking and risk reduction efforts.<br>Program directors reported the data in aggregate to quality improvement and management personnel at the end of each quarter.<br>The programs' insurer provided number and total yearly monetary value of worker's compensation claims.<br>Data on position of restraints, and number of client and staff restraint-related injuries were collected from annual reports to an overseeing government agency. Position of restraints data was collected immediately after restraint occurrence in a mandatory report of the incident.<br>Similarly, data on client restraint-related injury and staff restraint-related injury was collected immediately after the incident, with injuries categorised as either minor (requiring on site medical treatment) or major (requiring further medical assistance). | Quantitative | Quantitative measures are described and presented graphically, but no statistical analyses were conducted.<br>Total yearly monetary value of worker's compensations claims was averaged per claim and per quarter for this evaluation.<br>Length of stay was averaged across all clients by year for examination. |
| Goetz & Taylor-Trujillo (2012)<br>(60) | A change in culture: violence prevention in an acute | Service evaluation | To describe the development of the patient-focused | Midwestern USA | The PFI model was initiated in 2005 | No name<br>Four psychiatric services: a residential | N/A | The data on seclusion and restraint, Code Gray episodes, and staff injuries were collected by the hospital as part of their routine monitoring. The staff safety survey (is a biannual survey with 10 Likert-style questions focusing on staff's perceptions of the | Quantitative | Descriptive statistics describing the data for each indicator were reported and presented graphically. |
|  | behavioral health setting |  | intervention (PFI) model and its impact on the reduction of patient violence and staff injuries at a behavioural health service hospital |  |  | treatment program for female adolescents (12-18y), an acute inpatient adolescent unit (12-18 y), an adult inpatient unit (19y>), and a sub-acute unit for adults with substance use and mental health problems. |  | environment, their training and patient aggression management) was developed by the hospital leadership. |  | The staff survey was scored by averaging staff scores for each question, and then aggregate scores on each question were compared across previous years. |
| Gonshak (2011)<br>(61) | Analysis of trauma symptomology, trauma-informed care, and student-teacher relationships in a residential treatment center for female adolescents | Pre-post study | To investigate i) the extent to which student trauma symptomology, staff beliefs about TIC, and staff quality interactions with students in the classroom, related to students' perceptions of their relationships with their staff member; ii) the extent to which training staff in a trauma-informed framework is associated with increased staff knowledge about trauma, increased beliefs about the effectiveness of TIC, and increased quality staff classroom behaviours; as well as improved student perceptions of staff relationships and decreased student report of trauma symptoms. | Louisville , Kentucky , USA | Data were collected from January 2009 – May 2009<br>Risking Connection teacher training was implemented in mid-March 2009 | Maryhurst Inc. residential treatment center<br>Female adolescents in a residential treatment centre for children who have severe emotional difficulties | Response rate: Not stated. | Procedures included classroom observations and the administration of surveys to both teachers and students.<br>Student observations and surveys examined student change associated with the teacher changes after the implementation of the Risking Connection training intervention.<br>Data were collected at four time points from students and two time points from teachers in late January 2009 and over the next five months.<br>Teachers were given surveys and were observed once pre-intervention and once post-intervention. Service users were given surveys twice pre-intervention and twice post-intervention.<br>Data collection was completed by university faculty researchers and graduate research assistants with the assistance of Maryhurst direct-care staff. Standardised instruction was provided to the participants at each time point of data collection. | Quantitative | Multiple regression analyses determined the extent to which students' trauma symptomatology, teacher beliefs about trauma-informed care, and teachers' emotionally supportive behaviour were associated with the students' perception of the student-teacher relationship.<br>Descriptive statistics were provided following teacher scores: Risking Connection Curriculum Assessment, Trauma-Informed Care Belief Measure, Teacher Fidelity to Risking Connection, and CLASS-Emotional Support subscale before and after the intervention. Paired sample t-tests compared the four teach measure scores pre-test and post-test, and effect sizes were calculated.<br>Pre- and post-test changes in student attributes were conducted comparing change between the two pre-tests and change between the two post- tests. Trends across time were also examined using means at all student time points. |
| Hale (2019)<br>(62) | Implementation of a trauma-informed care program for the reduction of crisis interventions | Pre-post study | To test the hypothesis that implementation of the Six Core Strategies in a child and adolescent behavioural health hospital would lead to a 25% decrease in rates of physical holds and seclusion for patients 6 months after implementation, and reduce the risk of re- traumatisation | Illinois, USA | May-November 2018 | No name<br>Child and adolescent (3-17y) inpatient behavioural health hospital | Response rate: Not stated. | Implementation of TIC took 3 weeks as each staff member has an opportunity to attend weekly educational sessions that focused on each of the six core strategies. Department leaders were aware of the time commitments this project initiative would take and agreed to let their team members participate.<br>The leadership team was provided TIC training first.<br>Once training was complete, the researcher obtained routine data on use of holds and seclusions at the end of each month.<br>Deidentified information was provided to the researcher. | Quantitative | Descriptive statistics were provided for the numbers of physical holds and seclusions in the six months prior to implementation of TIC, and in the six months after. Data were also presented graphically.<br>No statistical analyses were performed |
| Hale & Wendler (2020) (63) | Evidence-based practice: Implementing trauma-informed care of children and adolescents in the inpatient psychiatric setting | Description of service implementation, and pre/post intervention study | To investigate the impact of implementation of TIC in an inpatient psychiatric setting versus routine care was on rates of use of physical holds and seclusion at 6 and 12-months post-implementation among children and adolescents (ages 3-17). | USA, region not stated | October 2017 - April 2019 | No name<br>A child and adolescent (ages 4-17 years) psychiatric hospital | N/A | The problem of too many uses of physical restraint and seclusion interventions was communicated to staff over 2 months before educational interventions focusing on TIC (with a focus on de-escalation techniques) were implemented. Multidisciplinary discussions and retooling of patient treatment and care plans began following the educational interventions. Implementation of video review debriefings followed 2 months later.<br>The entire process took 6 months. Culture change was in place by the end of the 12 months. | Quantitative | The number of uses of restraint and seclusion events 6 months before and after implementation were reported, and the changes in these figures described.<br>No statistical analyses were performed. |
| Isobel & Edwards (2017) (64) | Using trauma informed care as a nursing model of care in an acute inpatient mental health unit: A practice development process | Mixed methods case study | To describe the process and effects on the nursing workforce of implementing a trauma-informed model of care in an acute mental health inpatient unit using a Practice Development process. | Sydney, New South Wales, Australia | Practice development project commenced in October 2012.<br>Interviews were conducted in May 2014, 18 months post-implementation . | No name<br>An acute mental health inpatient unit | A purposive, convenience sample of nurses participated as determined by length of experience, involvement with the project and availability at time of interview.<br>Response rate: Not stated. | Semi-structured interviews with nurses were conducted on the unit by a research assistant.<br>Nurses were asked about how they felt about the changes occurring within trauma-informed care framework, the effects on their day-to-day work, consumers, the team and how they felt about their roles.<br>Interviews were audio-recorded and transcribed. | Qualitative | Transcripts were analysed using an inductive conventional content analysis.<br>Transcripts were examined to identify themes and emerging expressions of experiences. These themes and expressions were compared to identify similarities and differences. |
| Jacobowitz, Moran, Best, & Mensah (2015) (65) | Post-traumatic stress, trauma-informed care, and compassion fatigue in psychiatric hospital staff: A correlational study | Cross-sectional correlational study | To explore whether there is a correlation between inpatient psychiatric health care workers' i) experience of | New York, USA | Data collection occurred between November 2011 - May 2012 | No name<br>A psychiatric hospital, providing short-term acute care in the following units: general adults, child | Convenience sample of inpatient psychiatric health care workers, consisting of registered nurses, psychiatric | Two trained research assistants met with the hospital staff to explain the study and solicit their participation.<br>A semi-structured questionnaire asked participants to identify their job title, usual shift, age, gender, years of psychiatric work experience, education level, height, build, living arrangement, when they last participated in patient aggression | Quantitative | The data were screened for outliers. 7% of the data were excluded. The data also were examined for missing values. 8.1% of participants failed to complete at least one of the questionnaires. There were no significant patterns the missing data. Missing data were replaced using the SPSS |
|  |  |  | traumatic events, ii) resilience to stress, iii) attitude/confidence in managing violent patient situations, and iv) compassion fatigue with respect to post-traumatic stress symptoms. |  |  | and adolescents, older adults, chemical dependency | aides, assistant counsellors, psychiatrists, case coordinators and therapeutic rehabilitation specialists.<br>Response rate:<br>Out of a total of 250 direct patient-care employees working at the hospital, 172 returned surveys (68.8% return rate).<br>Of the returned surveys, 158 had complete information (8.1% of returned surveys had missing information). | management training, and when they last participated in a TIC meeting. Each question provided between two to seven answer choices from which participants selected the best response.<br>Participants received a token of appreciation (a pen) for placing their questionnaires in a locked box.<br>Only the PI had access to the contents of the box while it was at the research site. |  | function for linear interpolation.<br>Normality of continuous variable distributions was assessed using the Kolmogorov-Smirnov Test. Only the Burnout subscale of the Professional Quality of Life Scale met the assumption of normality.<br>The alpha level was set at .05, and two-tailed analyses were used for all of the tests.<br>The Post-Traumatic Checklist variable was transformed and adjusted using the scores on the Life Events Checklist. Correlations between the adjusted PCL-C scores, the demographic variables and standardised measures were analysed using Spearman correlation coefficients. The Kruskal-Wallis H-Test and the Mann-Whitney U-Test compared the Adjusted PCL-C scores to the demographic variables.<br>Significant relationships were entered into a Hierarchical Linear Regression Analysis to determine a best-fit model. |
| Jones (2021) (66) | How distress is understood and communicated by women patients detained in high secure forensic healthcare, and how nurses interpret that distress: An exploration using a multi-perspective | Qualitative | To explore how women patients in high secure healthcare understand and communicate their distress, as well as how nurses interpret the women patients' | Nottinghamshire, UK | Not stated | National High Secure Healthcare Service for Women (NHSWSW)<br>National high secure inpatient psychiatric service for women | Each team was approached independently, and each patient considered in terms of capacity to consent. The study was explained to the patient group and | This study adopted a co-production approach and used feminist principles as a framework.<br>A working party, including the researcher and voluntary patients (n=8-13 depending on availability) facilitated the research process.<br>Patients used the Personal Distress Signature (PDS) to record descriptions of their experience of distress and associated protective factors.<br>Twenty patients volunteered the written content of their PDS to form the basis of | Qualitative | The audiotaped interviews and handwritten notes were transcribed by the researcher.<br>Multi-perspective Interpretative Phenomenological Analysis (IPA) was used to analyse semi-structured interviews.<br>Analysis of the data was presented to the women patients for in-depth review and were also subject to peer |
|  | interpretative phenomenological analysis |  | distress. The study seeks to provide profound and meaningful insights into the lived experience of psychological distress on thoughts, emotions and behaviours for women patients in the National High Secure Healthcare Service for Women (NHSHSW). |  |  |  | nurses via community and multidisciplinary team meetings.<br>Named nurses of patient participants were approached. Informed consent was gained from all participants. All named nurses agreed to take part.<br>Response rate: Not stated. | interviews. Contents of these PDSs were thematically analysed to develop the semi-structured interview guide to explore patients' perspectives on distress. Speech and language specialists were consulted to adapt the questions and prompts to make them accessible to people with communication difficulties.<br>Themes were presented back to the patient group for validation. Pilot interviews were conducted. Individual semi-structured interviews were conducted with volunteer patients and participating nurses.<br>Focus groups were facilitated by facilitators. Not all participants agreed to audio-recording so handwritten notes were made on flip chart paper. Patients and staff facilitating took turns taking notes. Information from the focus groups was presented to individual patients who did not attend the focus group (e.g., due to being in seclusion) and they contributed. Their contributions were fed back to the group.<br>Individual patient interviews lasting 30 minutes or less were conducted in a private room. Participants were given the option for the interview to be audio-recorded. The interview could be conducted over two sessions if preferred.<br>Nurse interviews took place in the same interview room or in a staff room. They were also given the option for the interview to be audio-recorded.<br>There was regular liaison with operational managers to ensure that practical support for women was provided to attend groups. Nurses were briefed to check-in with and provide emotional support to participants if needed. |  | review. Quality of analysis was ensured via independent reviews completed by practitioners familiar with IPA methods |
| Korchmaros, Greene & Murphy (2021) (67) | Implementing trauma-informed research-supported treatment: | Longitudinal study | The study examined programme fidelity and the feasibility and | South-western USA | Not stated | No name<br>A mixed-gender adolescent residential | At all time points, all agency staff were invited via email to complete an | Survey data was collected from agency staff at:<br>4 months (Time 1)<br>1.25 years (Time 2)<br>2.25 years (Time 3) into implementation of the TI-RSTs. | Quantitative | Single-sample, 1-tailed t tests were used to test whether staff, on average i) agreed that the TI-RSTs were implemented with fidelity and they agreed that the TI-RSTs |
|  | Fidelity, feasibility, and acceptability |  | acceptability of implementing trauma-informed research-supported treatments (TI-RSTs) at an adolescent residential substance misuse treatment agency over time. |  |  | substance misuse treatment agency | online survey or to complete paper copies of the survey. Staff held diverse roles.<br>Response rate: Not stated. | An independent research assistant set up a laptop in a private area at the treatment agency for staff without computer access and made attempts to encourage completion of the survey. Staff completed surveys anonymously and privately. No demographic information was collected on individual surveys to preserve anonymity. |  | were implemented correctly ii) agreed that implementing the TI-RSTs was feasible such that staff agreed that their agency was capable of making and sustaining change; their agency was ready to implement the TI-RST, and agency leaders were prepared to implement the TI-RST iii) agreed that implementing the TI-RSTs was acceptable and the TI-RSTs improved client outcomes, iv) there were greater than moderate attitudes towards RSTs and being more than slightly satisfied with the TI-RSTs.<br>ANOVAs assessed changes over time, differences across type of TI-RST, and differences across related types of fidelity and feasibility measures, or indicators.<br>The data collected at Times 1, 2, and 3 were treated as independent in analyses. |
| Kramer, Michael George (2016) (68) | Sanctuary in a residential treatment center: Creating a therapeutic community of hope countering violence | Qualitative, data included: group observation; content analysis of agency documents and quantitative data; focus groups; and individual interviews | To determine how and why the Sanctuary model works in decreasing trauma symptoms with a population of court-committed youth. | USA, region not specified | The Sanctuary Model was implemented over a three-year period (exact dates not provided).<br>Group observation took place over a year and a half. | No name<br>Forensic male adolescent residential treatment unit. | Participants were introduced to the researcher, provided with information about the study and invited to participate.<br>Response rate: Not stated. | The researcher began as a non-participatory observer of administrative and clinical team meetings, individual service plan meetings, and community meetings over a year and a half. Field notes were kept and general themes were extracted.<br>The researchers obtained access to organisational records and documents, and minutes of Core Team, and Red Flag Review meetings, covering the three-year period of implementing Sanctuary. The researcher examined their content for emergent themes.<br>Focus groups (30-60 minutes) were conducted; two for staff and three for service users. The groups were recorded and transcribed verbatim. They were held in an administrative conference room. For service users, focus groups were held in residential conference rooms. | Qualitative | The digital audio-recordings of focus groups and the planned individual interview were transcribed verbatim.<br>Grounded theory and utilisation-focused evaluation were used to analyse the data. |
|  |  |  |  |  |  |  |  | Three 30–60-minute individual interviews were conducted: two impromptu and one planned with staff. Two interviews occurred after chance meetings (not transcribed as the researcher did not have the audio-recorder). The interviews were conducted in an office, with the third interview requested in lieu of being a staff focus group participant. This interview was audio-recorded and transcribed, conducted in an administrative meeting room. |  |  |
| Niimura, Nakanishi, Okumura, Kawano & Nishida (2019) (69) | Effectiveness of 1-day trauma-informed care training programme on attitudes in psychiatric hospitals: A pre–post study | Pre-post study | To evaluate the effects of a TIC training programme on attitudes towards TIC practice in mental health professionals, using standardised measurement instruments | Tokyo, Japan | March 2018 - June 2018 | Tokyo Metropolitan Institute of Medical Science<br>29 inpatient psychiatric hospitals across Tokyo, Japan | All hospitals with psychiatric beds in Tokyo and its suburban prefectures (Kanagawa, Saitama, Chiba, and Yamanashi) were approached. The invitation letter included information regarding a 1-day TIC training programme. Mental health professionals voluntarily responded to the research team to participate in the TIC training programme.<br>Response rate: Not stated. | Participants attended the 1-day TIC training programme.<br>The programme was run and co-facilitated by some of the authors.<br>After the TIC training programme, participants received a gift card worth 100 yen (\$9.03 USD) for attending.<br>Self-rated questionnaires, including the Attitude Related Trauma-Informed Scale (ARTIC-35) and a self-rated questionnaire developed by the study authors, were administered at pre-training, post-training and 3-month follow-up. | Quantitative | Participants characteristics were summarised, including sex, age, job category and year sin the psychiatric field.<br>Missing values for each item in the ARTIC-35 were imputed using the multivariate imputation. Longitudinal data structure was considered using the predictive mean matching technique for two-level data. Twenty imputed data sets were generated with 10 iterations per imputation. For each of the imputed data sets, the following analyses were repeated: 1) the sum of the items on the ARTIC-35 for each participant were divided by the number of items, 2) means and standard deviations for the average scores for each time point were obtained, 3) mean differences and 95% confidence intervals for the average scores between time points were estimated.<br>Analyses were repeated using the complete case method. Using data from 56 participants in the analysis, the proportion of TIC implementation at the 3-month follow-up was calculated. |
| Prchal (2005) | Implementing a new treatment philosophy in a | Qualitative focus groups | To describe the process of | North-eastern USA | January 2002 - March 2002 | No name | All staff who worked in the Sanctuary | Ten focus groups were conducted. Three involved clinicians and administrators/ supervisors (7-12 participants in each group) | Qualitative | A mid-range, inductive, coding scheme was used to analyse the qualitative focus group |
| (70) | residential treatment center for children with severe emotional disturbances: A qualitative study |  | implementation of the Sanctuary Model of treatment in a residential treatment centre for youths with severe emotional disturbances, from the viewpoint of staff who volunteered to participate in focus groups about the model and its implementation. |  |  | Three residential, youth therapeutic services specialised to treat youths with conduct disorders and other serious emotional disturbance | Model units were invited to participate in focus groups.<br>Response rate: Not stated. | and seven involved milieu counsellors (3-10 participants in each group).<br>Each group session was recorded, and the recordings were transcribed. Notes were taken and then compared with the transcripts. When participants did not consent to recording, extensive notes were taken and then typed by the transcriptionist.<br>All training on the Sanctuary Model took place prior to this initial implementation stage. |  | data, with some structure imposed by the research questions and overall goals of the study.<br>A multi-step approach to coding was used. The coding process was undertaken by the PI and a doctoral student. Consensus on finalised code definitions was reached through discussion between the PI and student. |
| Prytherch, Cooke & Marsh (2020)<br>(28) | Coercion or collaboration: service-user experiences of risk management in hospital and a trauma-informed crisis house | Qualitative | To explore service-users' experiences of risk management in both hospital services and a trauma-informed crisis house | North London, UK | Not stated | No name<br>A residential female crisis house | Five current residents were approached by crisis house staff, two of whom participated. Six previous residents volunteered to take part.<br>Response rate: 2/5 | Interviews took place at the crisis house and lasted approximately 1 hour. Participants were asked about their experiences in the crisis house and in hospital services. All interviews were audio-recorded. Pseudonyms were used to protect confidentiality. | Qualitative | Thematic analysis within a critical realist framework was used.<br>Data was analysed iteratively. An inductive approach was used. Rather than aiming for "theoretical saturation," the findings are presented as one interpretation that is "far enough along to make a contribution to our evolving body of understandings."<br>The analysis was sent to all participants to validate the analysis. |
| Rivard, McCorkle, Duncan, Pasquale, Bloom & Abramovitz (2004)<br>(71) | Implementing a trauma recovery framework for youths in residential treatment | Service evaluation | To describe an intervention designed to address the special needs of youths with histories of maltreatment | North-eastern USA | The first round of focus groups was conducted during late January – early March 2002. | No name<br>Three residential youth therapeutic services (each composed of smaller residential | N/A | The Sanctuary Model was first piloted in five residential units that self-selected to participate in the initial phase of the project. Following the pilot, four additional residential units were randomly assigned to implement the model. Eight other residential units, where the model was not being implemented, served as the usual services comparison. | Qualitative | Focus groups were audiotaped, with permission of participants, and transcribed. Responses to each of the focus group questions were compiled across the groups. The aggregated responses to each question were then coded and displayed in matrices by major themes, |
|  |  |  | and exposure to family and community violence. To describe the main components of the evaluation, the evaluation design, and initial impressions of staff during the course of implementing the intervention. |  |  | units) specialised to treat youths with conduct disorders and other serious emotional disturbances |  | A comparison group design (Sanctuary versus Standard Residential Services) with measurement at five points (baseline, 3, 6, 9, and 12 months) examined change in the two conditions.<br>Several standardised instruments were used to measure changes in the therapeutic community environments and change in youths' functioning and behaviours. Change in the frequency of critical incidents (e.g., harm to self, others, or property) and use of physical restraints was measured by accessing and analysing data from the agency's management information system.<br>Implementation adherence to the original model was measured through consultant's process notes, periodic review of a Sanctuary Model Milestones Checklist, and a Psychoeducation Group Fidelity Checklist.<br>Perceptions of the course of implementation, and challenges in implementing the model were collected through focus groups, convened every 6 months. Ten clinician and administrator/supervisor focus groups were conducted (n=7-12 per group). Seven focus groups involved counsellors (n=3-10 per group). Participants were also asked about factors facilitating and inhibiting their ability to implement the model. |  | categories, and sub-categories. The major themes that emerged across all focus groups were narratively described. |
| Rivard, Bloom, McCorkle, Abramovitz, (2005)<br>(72) | Preliminary results of a study examining the implementation and effects of a trauma recovery framework for youths in residential treatment | Non-randomised control design (with nested process evaluation) | To examine the implementation and short-term effects of the Sanctuary Model as incorporated into residential treatment programs for youth | North-eastern USA | The Sanctuary Model was first piloted with four residential units. During this phase, the staff training protocol and manual was developed and piloted between February and August 2001.<br>Four additional residential units were randomly | No name<br>Three residential youth therapeutic services (each composed of smaller residential units) specialised to treat youths with conduct disorders and other serious emotional disturbances | Four units self-selected to participate in the initial phase of the project. Four additional residential treatment units were randomly assigned to implement the Sanctuary Model in 2001. | The Sanctuary Model was first piloted in four residential units that self-selected to participate in the initial phase of the project. The staff training protocol and manual was developed and piloted between February-August 2001.<br>Four additional residential treatment units were randomly assigned to implement the Sanctuary Model in autumn 2001. Eight other units, providing the standard residential treatment program, served as the usual services comparison group.<br>Progress in implementing the model was documented through consultants' notes and periodic reviews of the Sanctuary Project Implementation Milestones checklist. | Mixed | A comparison group design, with measurement at three points (baseline, 3 months, 6 months), was used.<br>Group differences in outcomes were explored using independent t-tests.<br>Repeated measure analyses were used to assess differences over time and by group.<br>Qualitative analysis method was not stated. |
|  |  |  |  |  | assigned to implement the Sanctuary Model in the fall of 2001. |  | The youth sample consisted of all youths for whom full informed written consent was obtained from custodial agencies, legal guardians, parents, and youths. The staff sample was composed of staff that worked in the programs and who voluntarily elected to participate in surveys and focus groups through a process of fully informed, written consent.<br>Response rate: Not stated. | Qualitative data on staff perceptions of implementation, and challenges in implementing the model, were gathered through focus groups. The Community Oriented Programs Environment Scale was administered to staff four times at 4–6-month intervals.<br>Youth demographics, historical data, history of abuse and neglect were abstracted from client records at baseline. Other measures of youth outcomes included: the Child Behaviour Checklist, the Trauma Symptom Checklist for Children, the Rosenberg Self Esteem Scale, the Nowicki-Strickland Locus of Control Scale, the peer form of the Inventory of Parent and Peer Attachment, the Youth Coping Index, and the Social Problem Solving Questionnaire. |  |  |
| Stamatopoulou, (2019) (73) | Transitioning to a trauma informed forensic unit: Staff perceptions of a shift in organisational culture | Qualitative evaluation of implementation of a TIC intervention | To provide a description of the impact of transitioning to a trauma-informed service model on staff working in an inpatient forensic unit in England | Northern England, UK | Implementation of the TIC programme: February 2018<br>Focus groups were conducted in January 2019 and February 2019. | No name<br>Inpatient female forensic mental health unit (comprised of four wards ranging between low- | The field supervisor made contact with the forensic unit.<br>Response rate: Not stated. | The researcher visited the unit in December 2018, after project approval. During this visit, locations, dates and times for the focus groups were agreed with the service. Paper copies of the participant information sheets were provided to interested staff and left in staff rooms and with the local contact for the researcher.<br>An open-ended interview schedule was devised. The questions asked about the incorporation of choice, trust, empowerment | Qualitative | Focus group data was analysed using thematic analysis. Inductive analysis was applied.<br>Data was coded through a social constructionist epistemology lens. Themes and subthemes were presented narratively. |
|  |  |  | and the factors that influence the progress of this transition |  |  | medium security) |  | and safety within the programme of change. All interviews were recorded.<br>Data collection was conducted in four 45-90 minute focus groups (n=3-7 staff members). Two focus groups were conducted with senior staff members. Two focus groups were conducted with staff in lower pay grades. The researcher was available following the focus groups for debriefing.<br>Quality of the project was assessed using the Eight "Big-Tent" criteria for Excellent Qualitative research (Tracy, 2010). |  |  |
| Tompkins & Neale (2016) (74) | Delivering trauma-informed treatment in a women-only residential rehabilitation service: Qualitative study | In-depth case study approach | To explore factors that affect the delivery of trauma-informed treatment in one women-only residential rehabilitation service and to identify any challenges experienced by staff (in delivering the programme) and clients (in receiving the programme). | UK, region not specified | April 2015 - August 2015 | No name<br>A female residential substance use treatment unit | Purposive sampling which aimed to encompass those likely to have varying experiences of the trauma-informed treatment.<br>Response rate: Not stated. | Thirty-seven semi-structured interviews were conducted.<br>Two researchers approached potential participants and provided them with study information. Participants volunteered to take part. Interviews were conducted in private. All interviews followed a topic guide that explored participants' personal circumstances and views and experiences of TIC. Staff were provided information on their experiences of delivering TIC, whilst clients were asked about their experiences of receiving it. Staff received no compensation for taking part; clients received a £10 high street gift card. All interviews were audio-recorded and transcribed. | Qualitative | Data were analysed using iterative categorization, involving participants.<br>The two coding systems were developed by researchers. The authors revisited the coded data to identify patterns, similarities and differences within and between different participant groups. Emerging findings were refined following discussions between the authors, two of the key stakeholders, and peer review. |
| Zweben et al. (2015) (75) | Enhancing family protective factors in residential treatment for substance use disorders | Pre-post study | To compare outcomes for women and children on a trauma-informed dual diagnosis residential treatment program versus treatment as usual | Oakland, California, USA | Not specified | Project Pride, East Bay Community Recovery Project<br>Residential treatment program for women who are pregnant or have young children | Residents at Project Pride who volunteered to participate in CF!<br>Response rate: N/A | The 'Celebrating Families!' (CF!) program was implemented. There were 44 women who participated, and 51 women who were in Project Pride but who did not take part.<br>Participants were assessed at baseline, discharge and 6-month follow-up. | Quantitative | Quantitative outcomes were reported but no statistical analyses conducted. |

### Study characteristics

The models reported on most commonly were the Six Core Strategies (n=7) and the Sanctuary Model (n=6). Most included studies were based in the USA (n=23), followed by the UK (n=5), Australia (n=2), and Japan (n=1). Most studies were undertaken in acute services (n=16) and residential treatment services (n=14), while one was undertaken in an NHS crisis house. Over a third of studies were based in child, adolescent, or youth mental health settings (n=12), while six were based in women’s only services. See Table 1 for full characteristics of all included studies.

Twenty-one studies gave no information about how they defined ‘trauma’ within their models. Of the ten studies that did provide a definition, four (56, 65, 73, 74) referred to definitions from a professional body (5, 76, 77); three studies (62, 68, 72) used definitions from peer-reviewed papers or academic texts (78–81); in two studies (55, 64), authors created their own definitions of trauma; and in one study (61), the trauma definition was derived from the TIC model manual. See full definitions in Table 2. Some studies reported details on participants’ experiences of trauma, these are reported in Table 1.

**Table 2.**
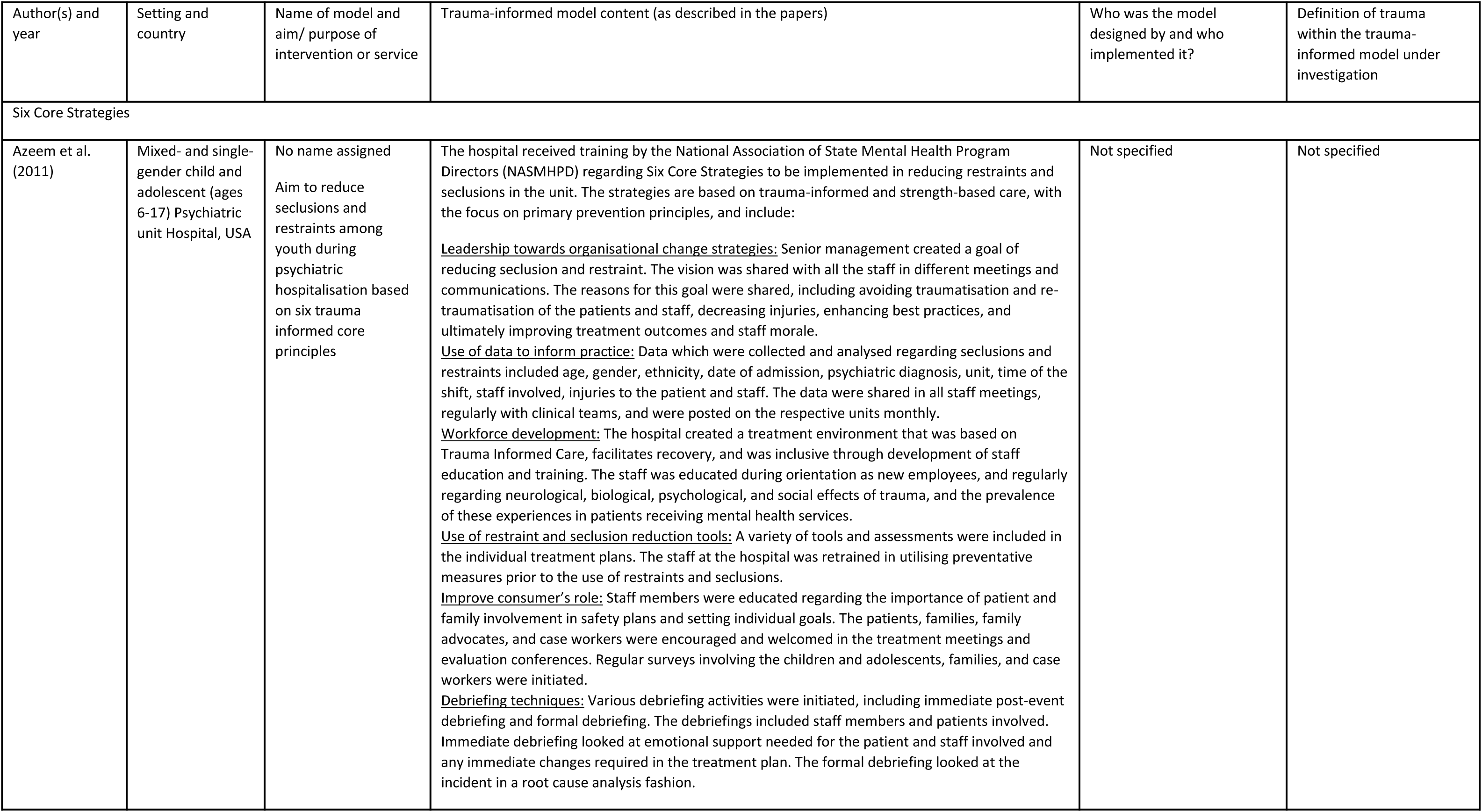
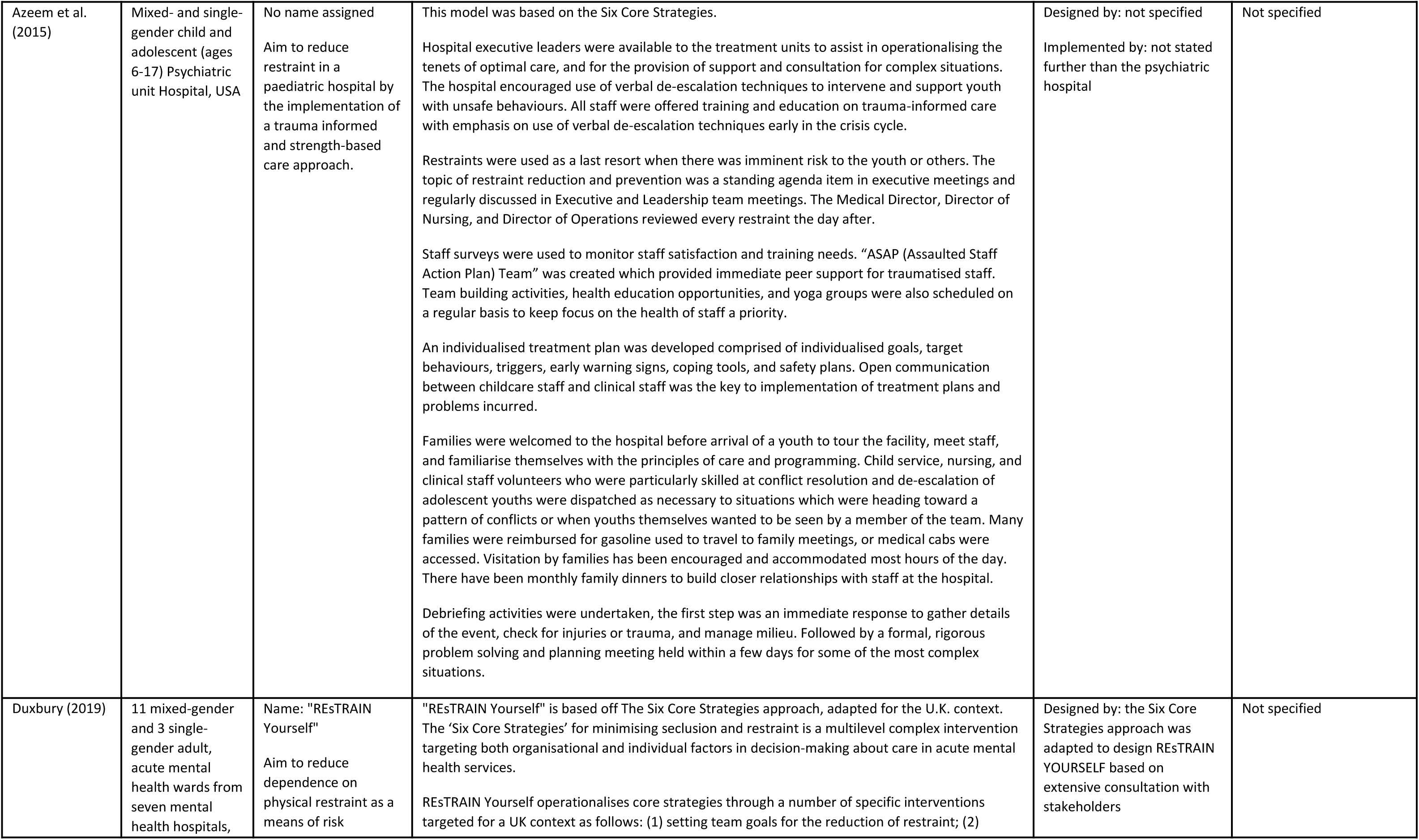
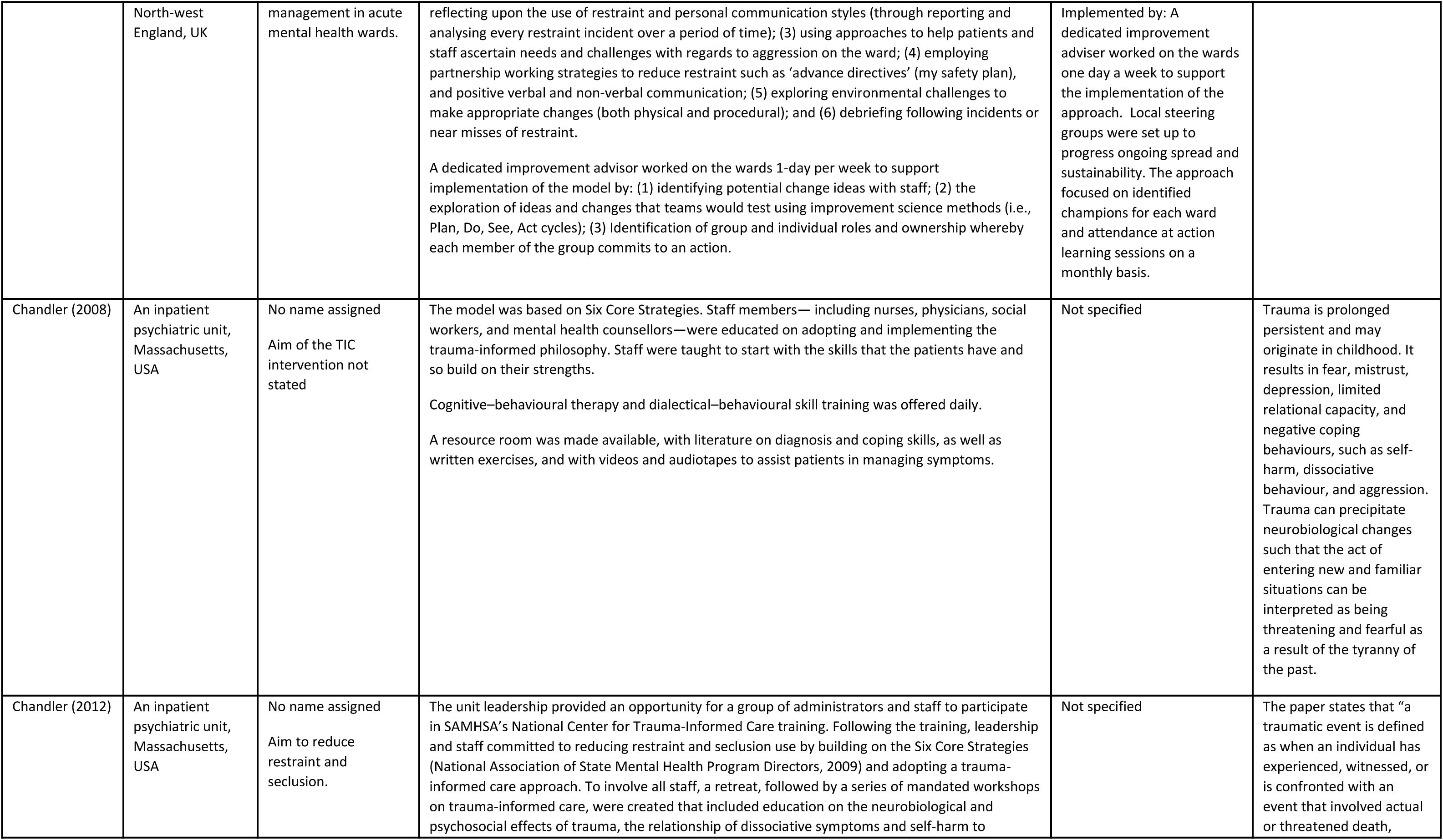
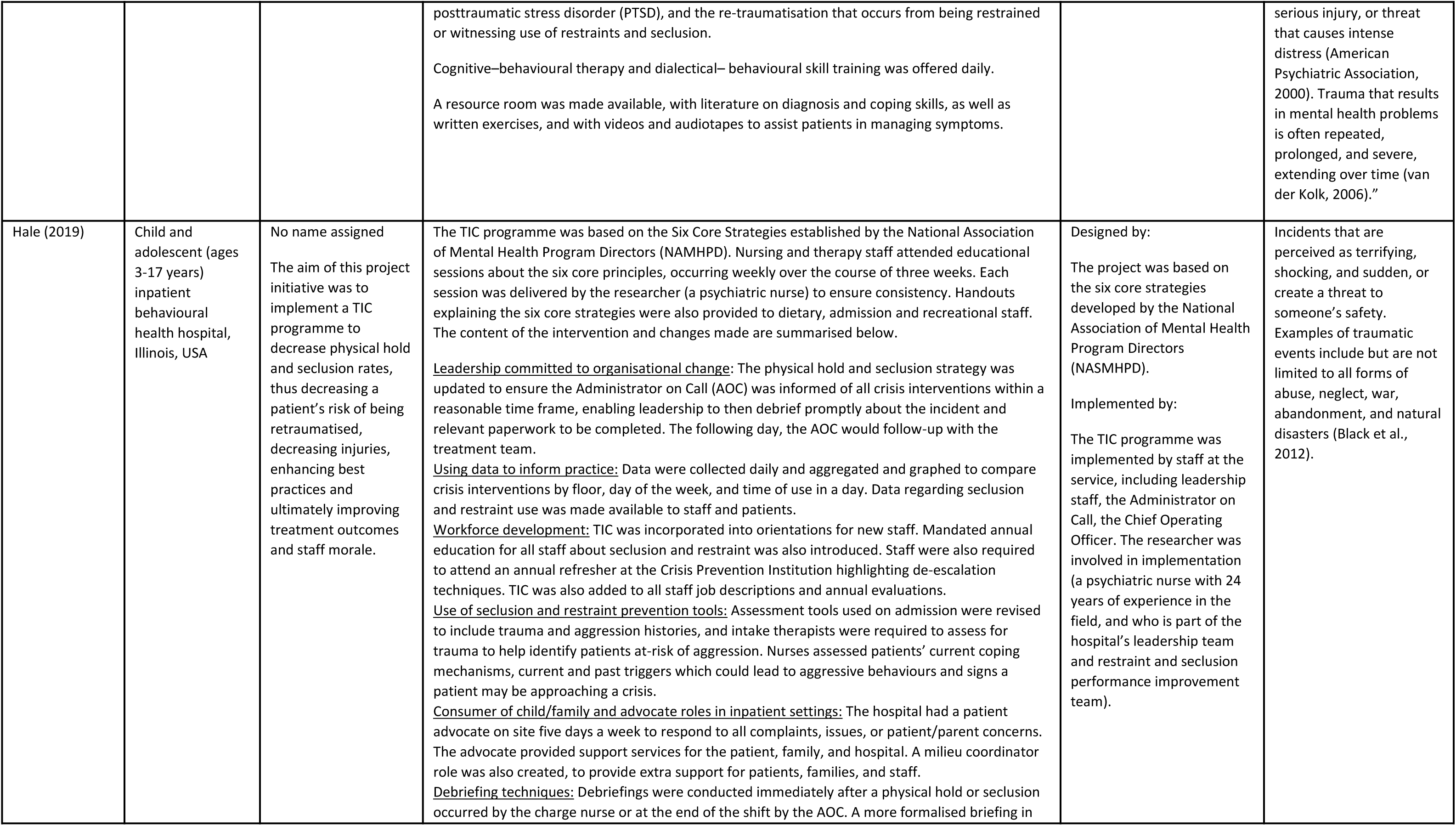
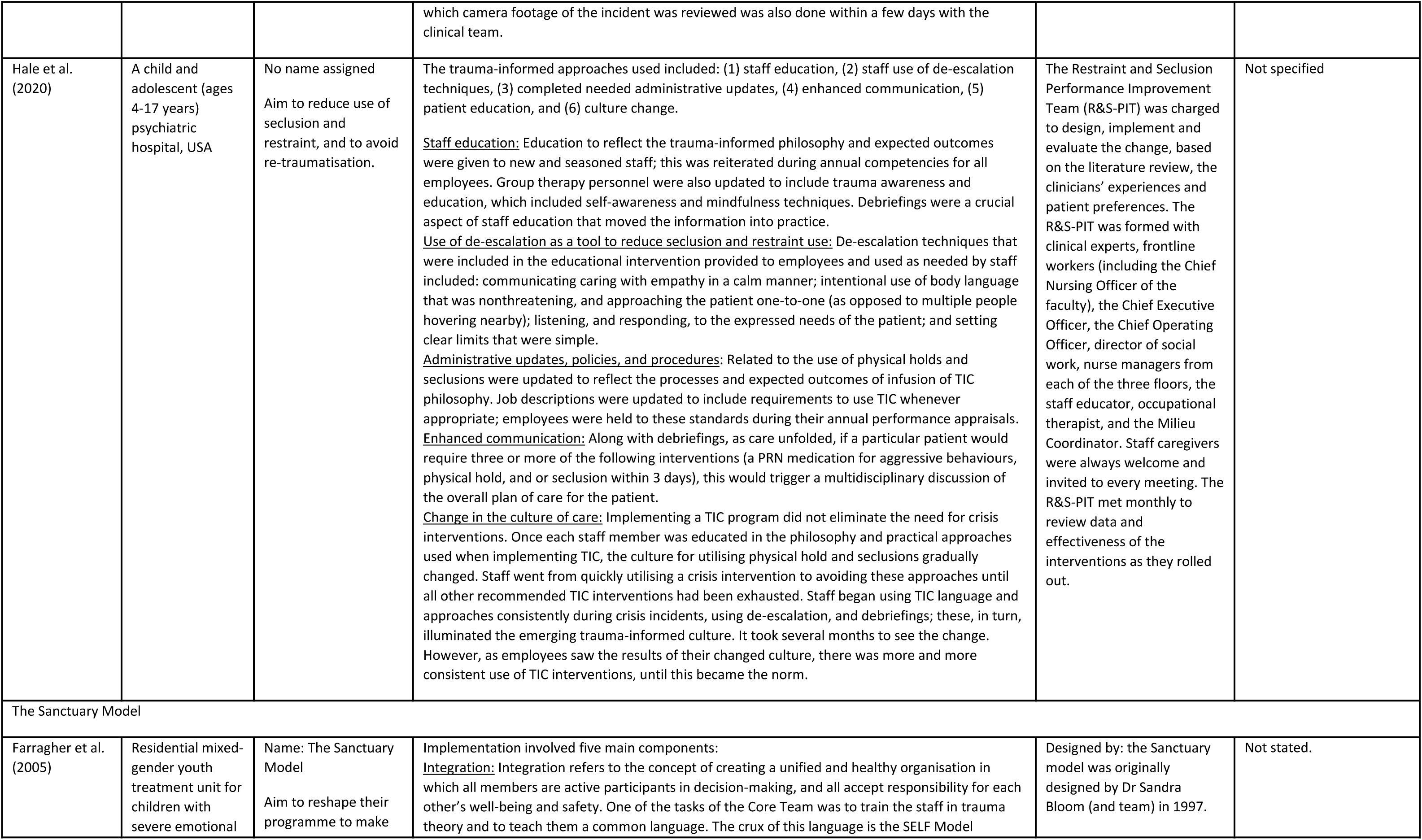
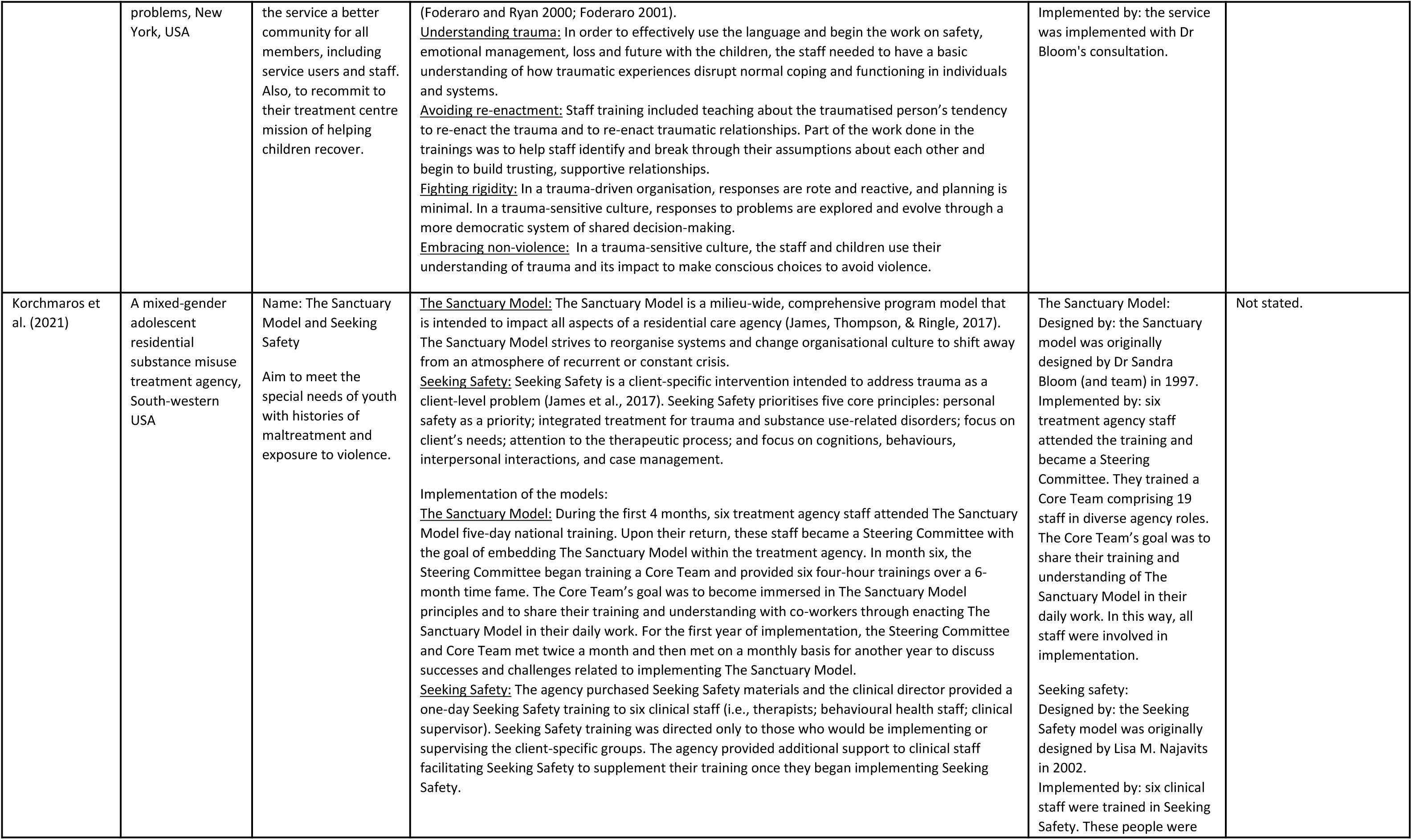
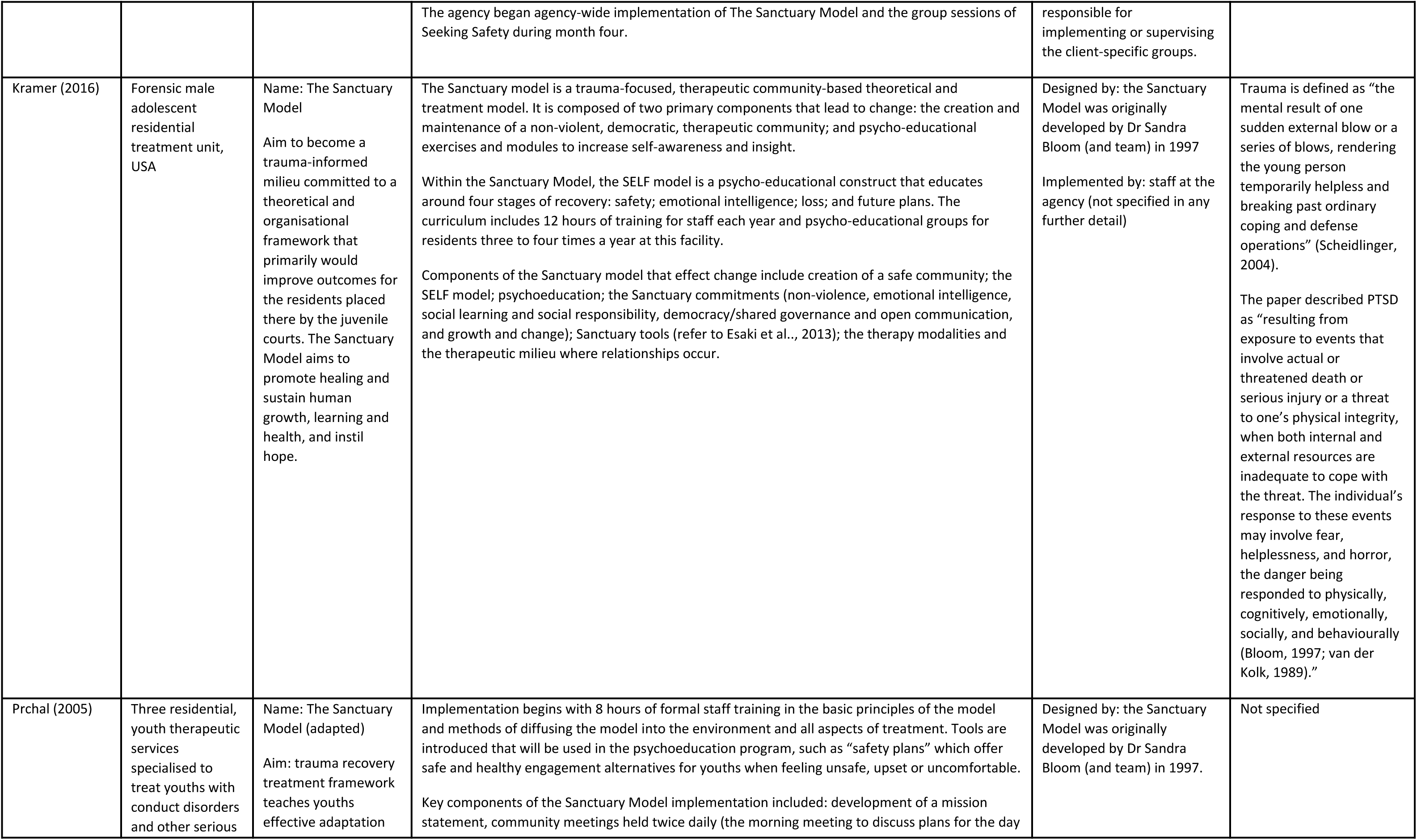
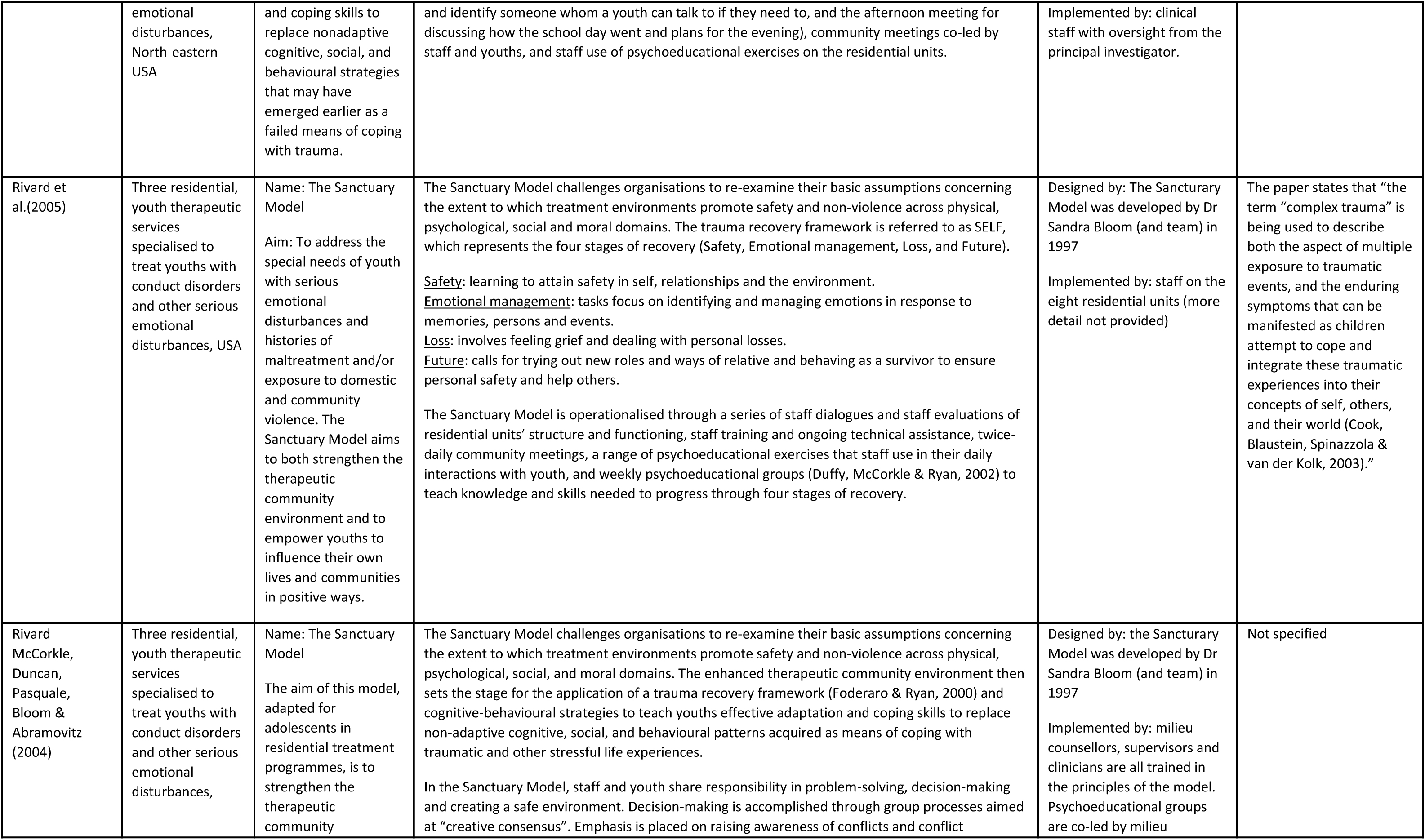
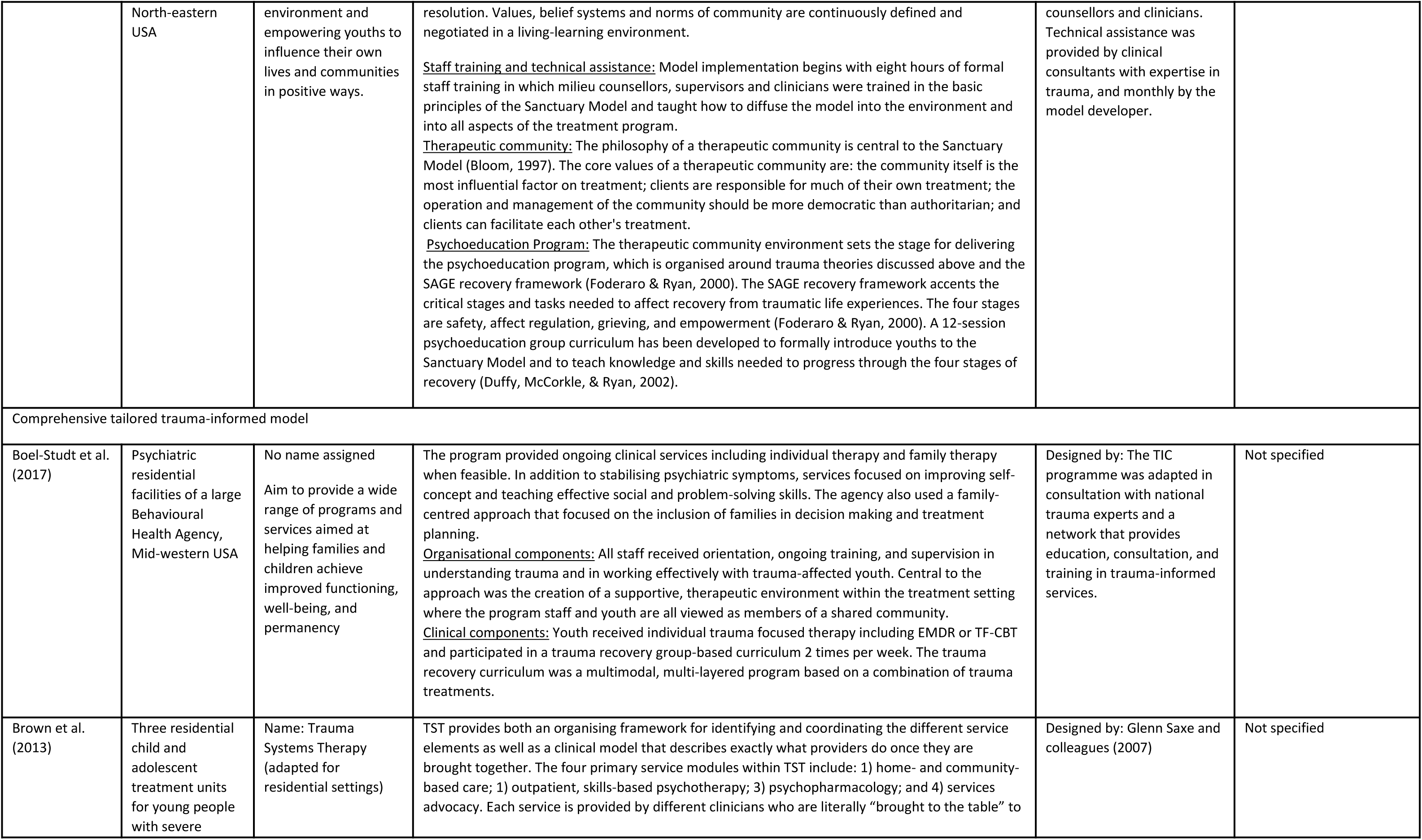
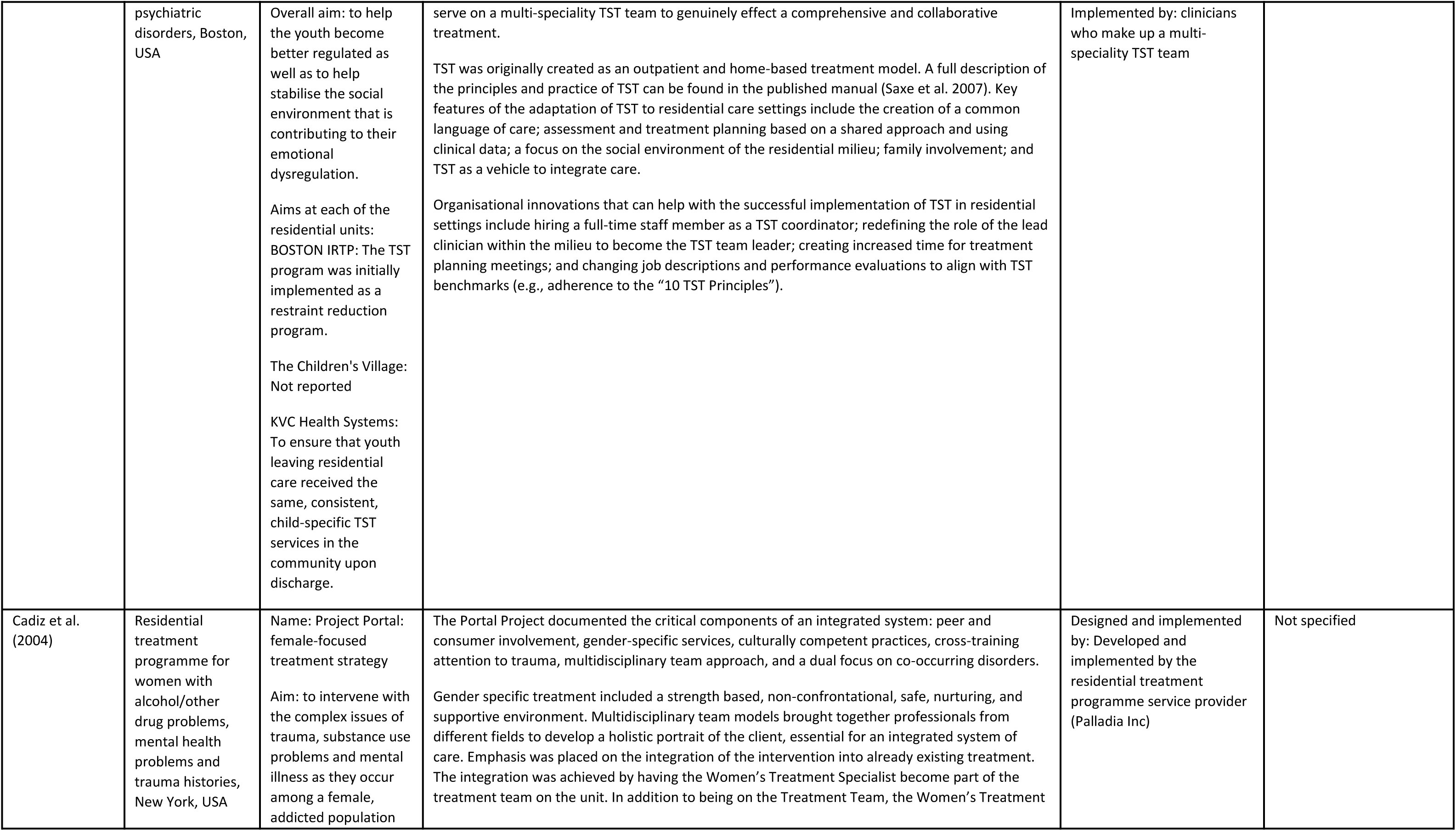
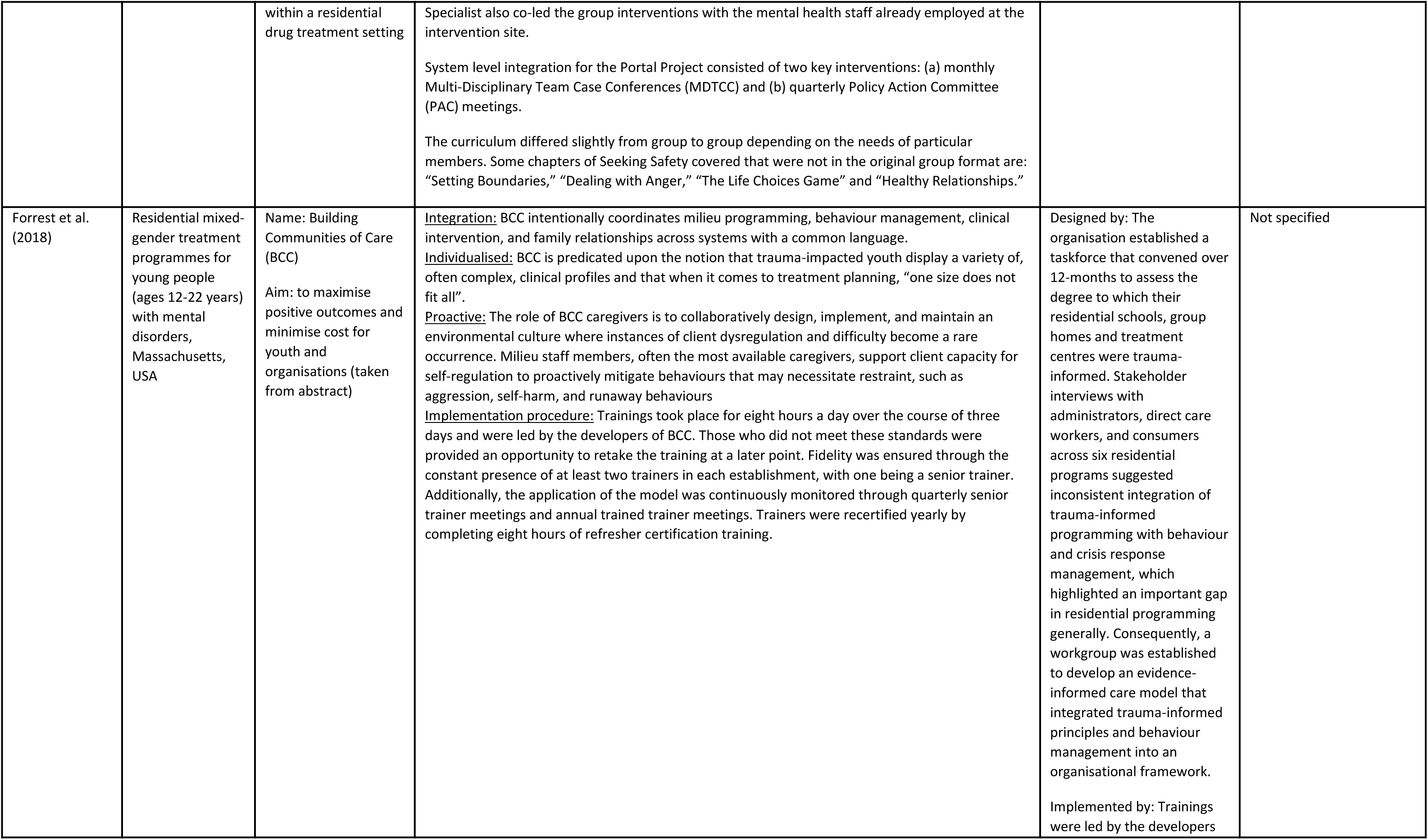
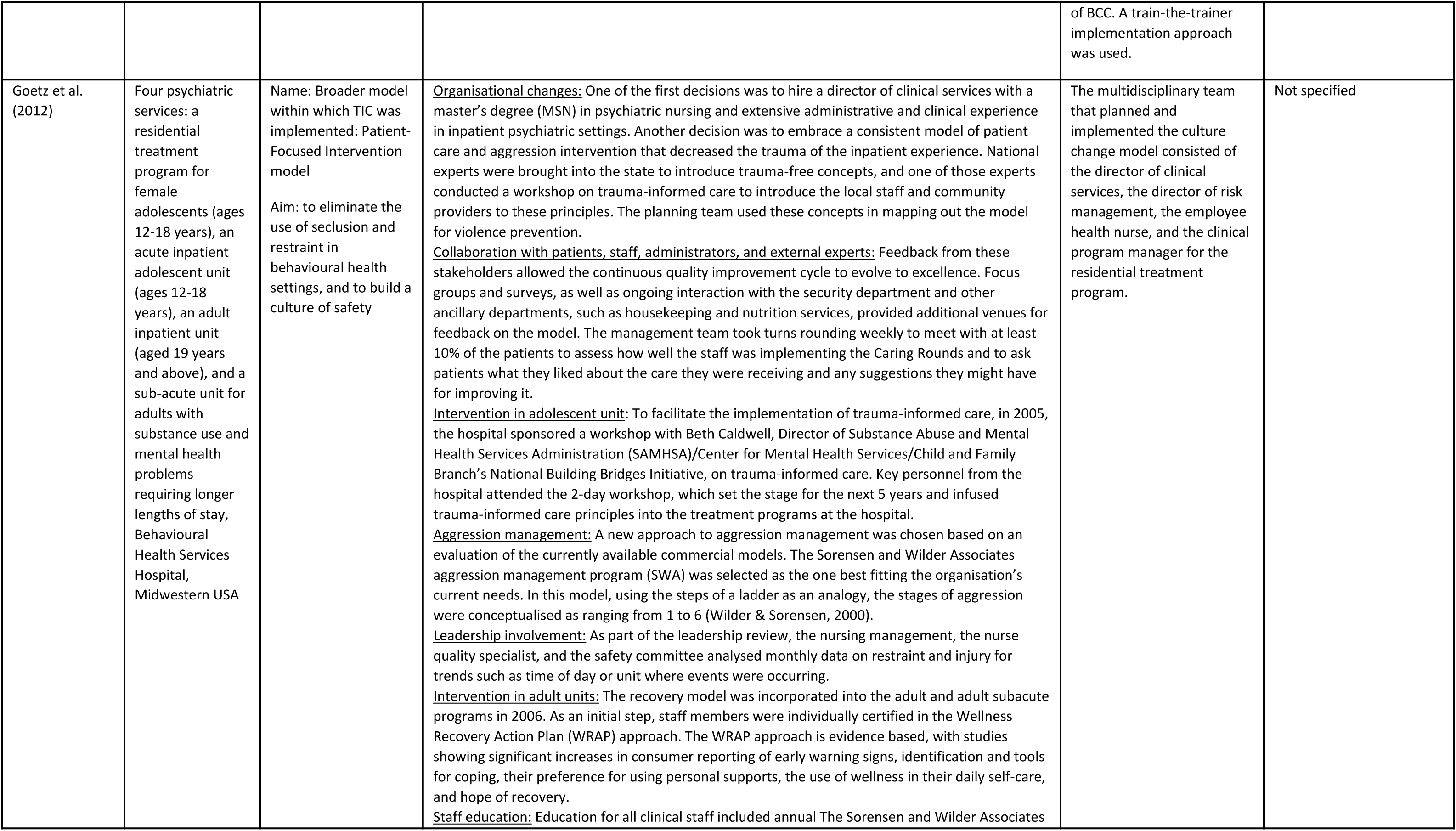
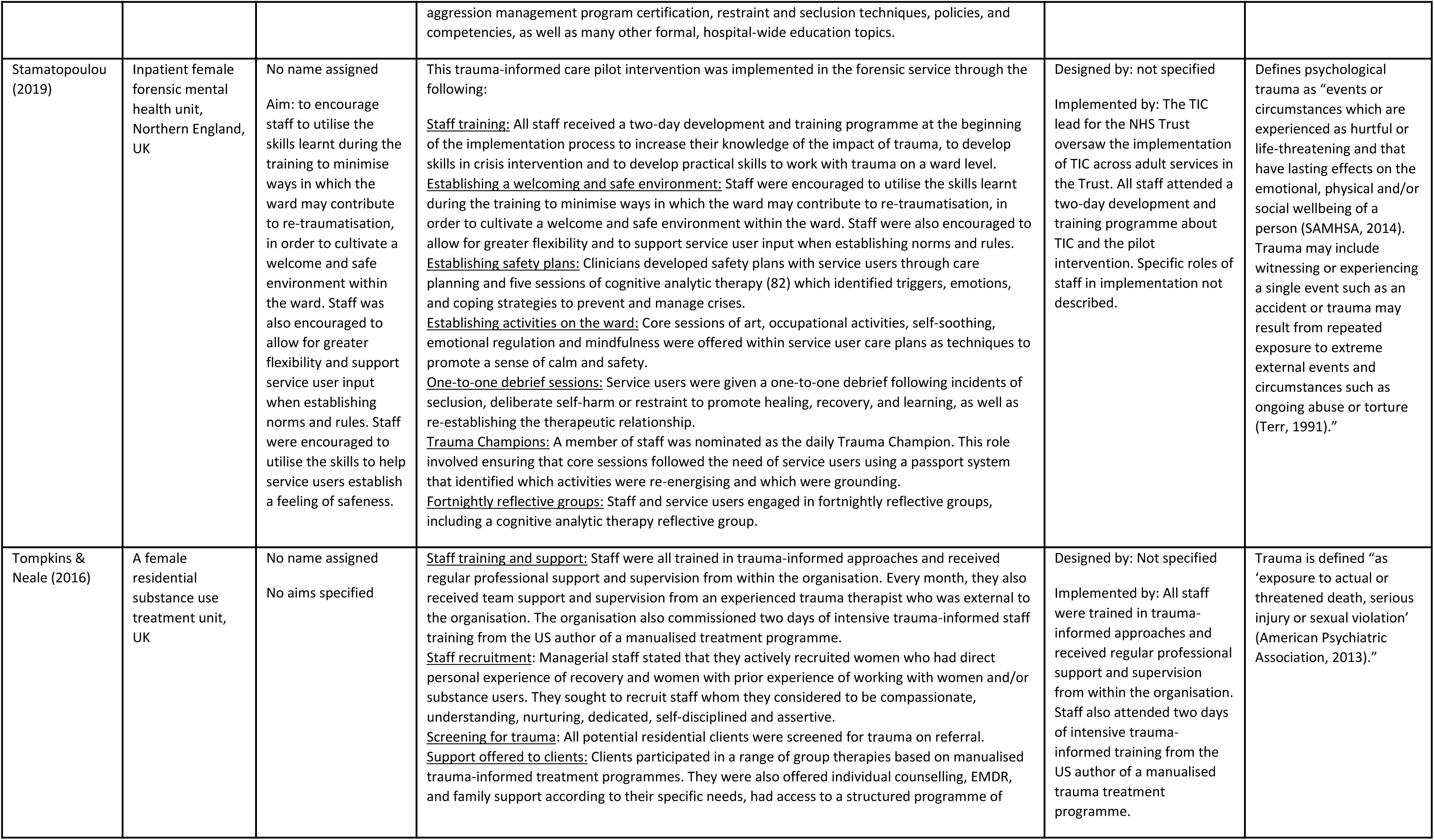
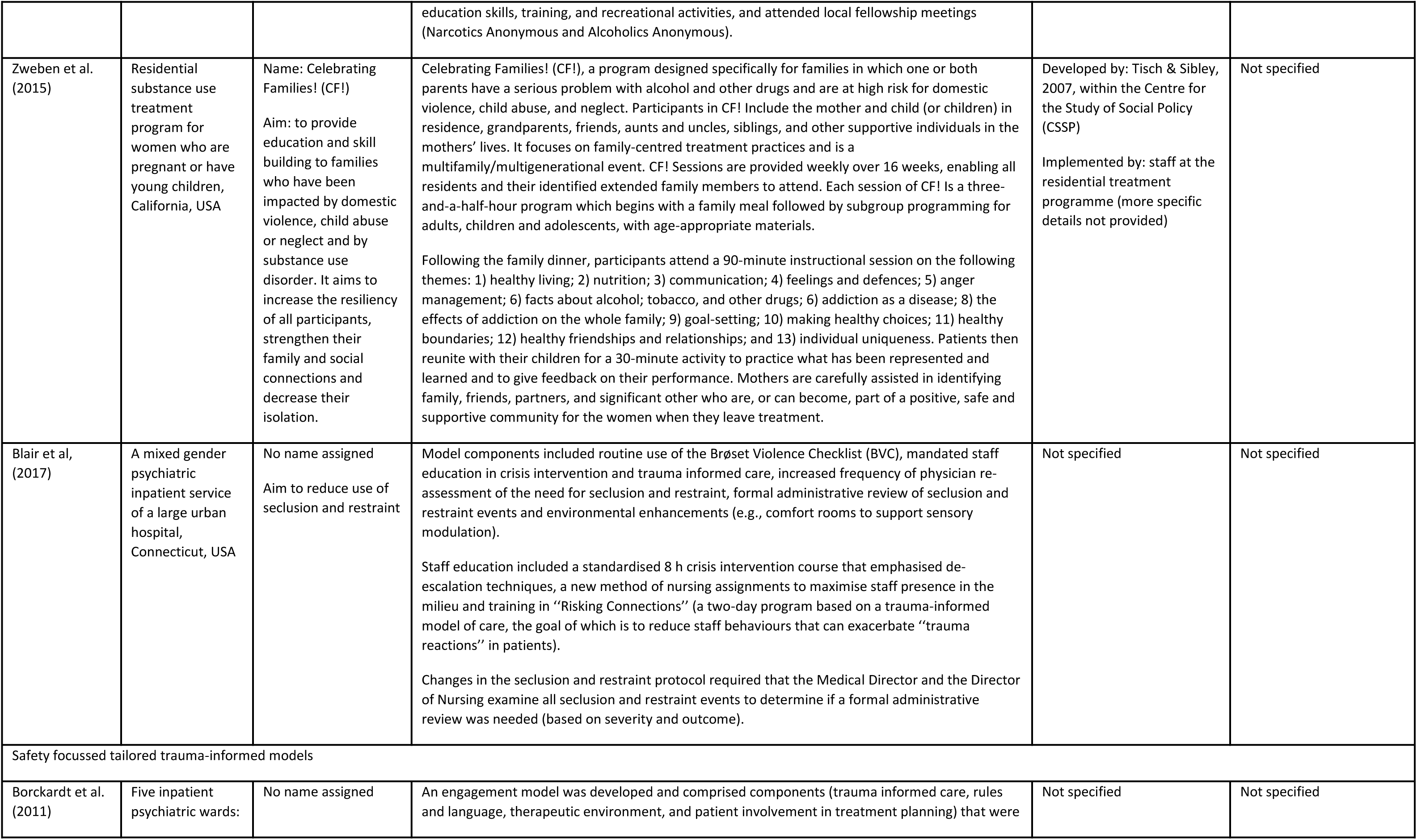
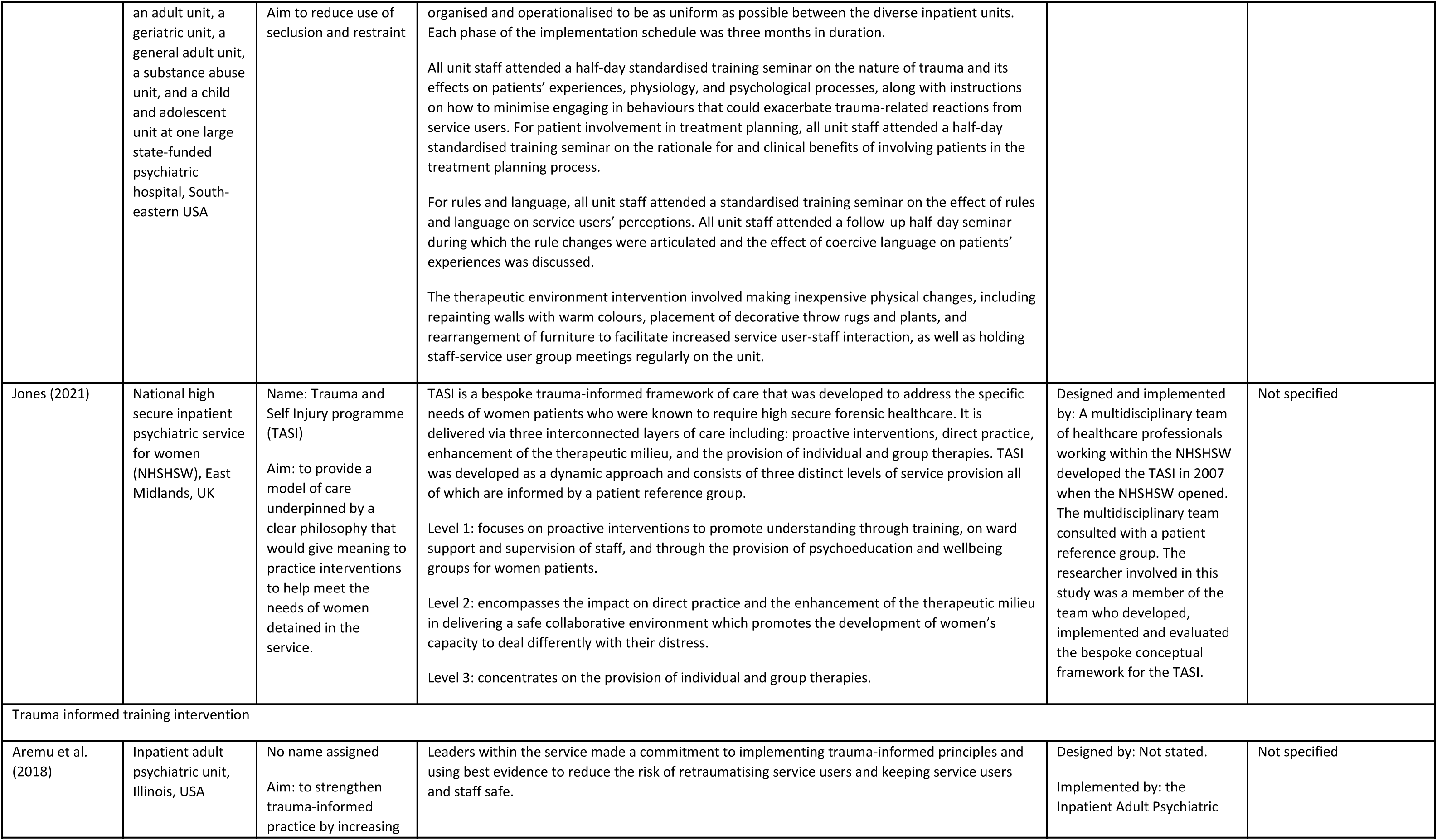
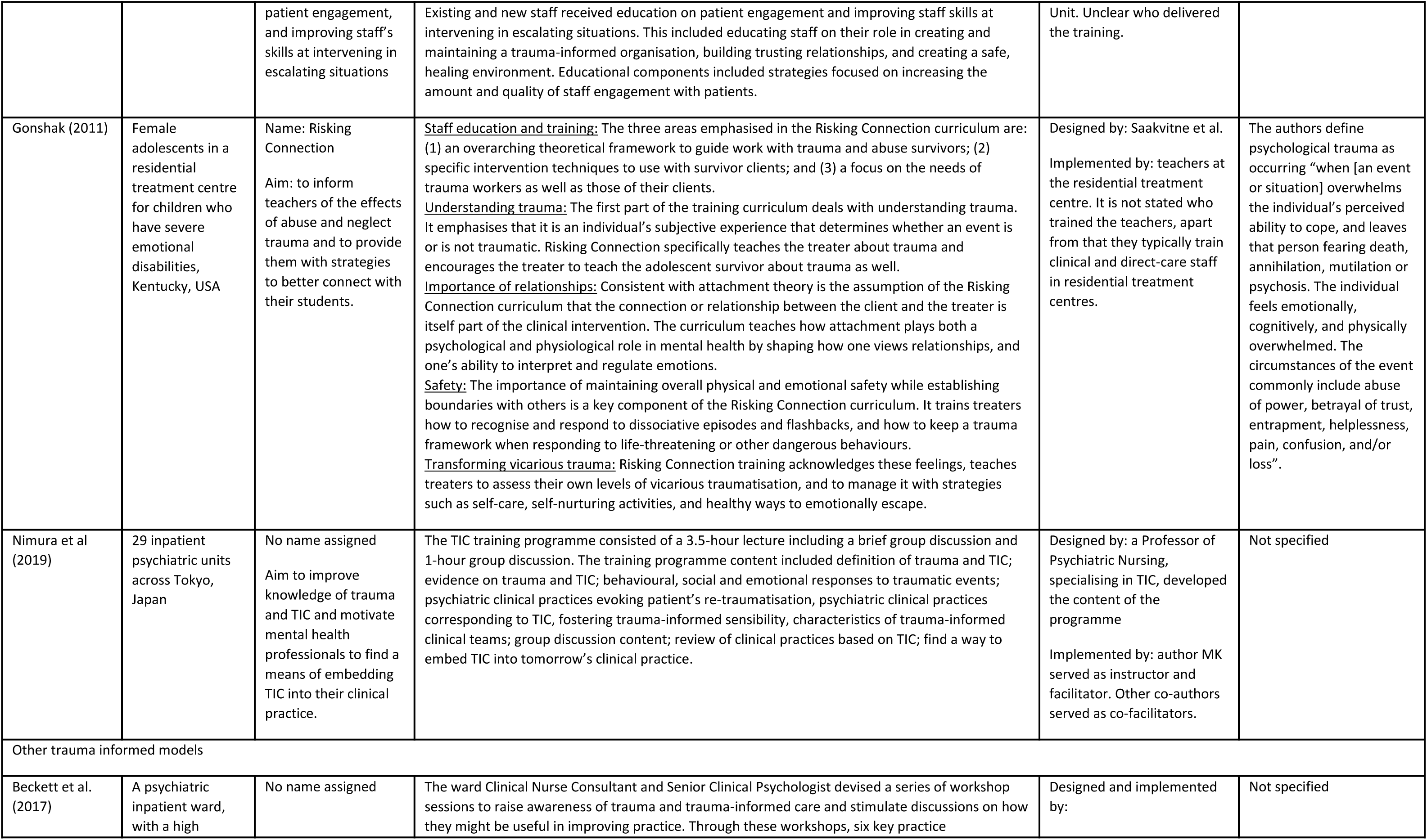
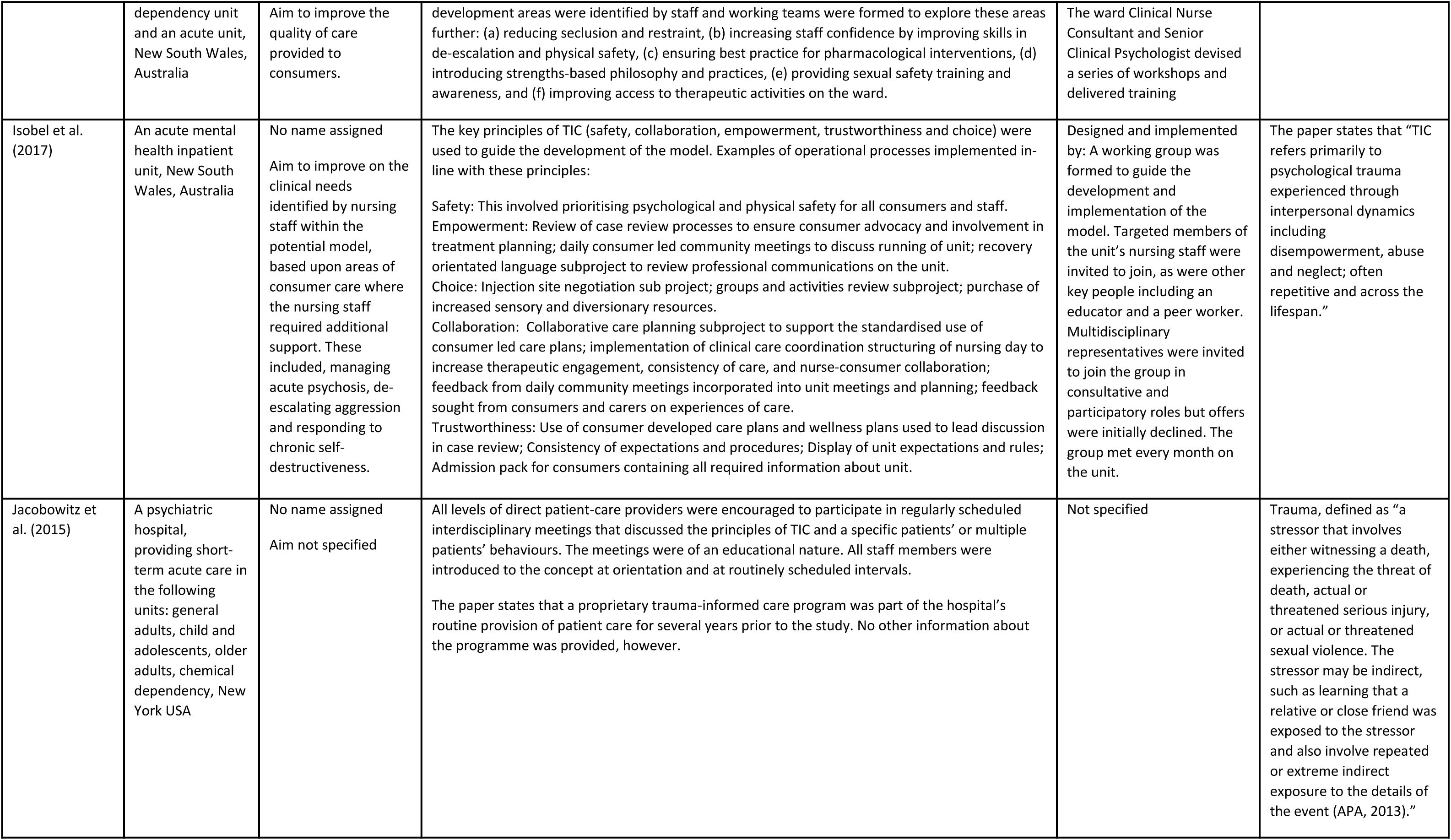
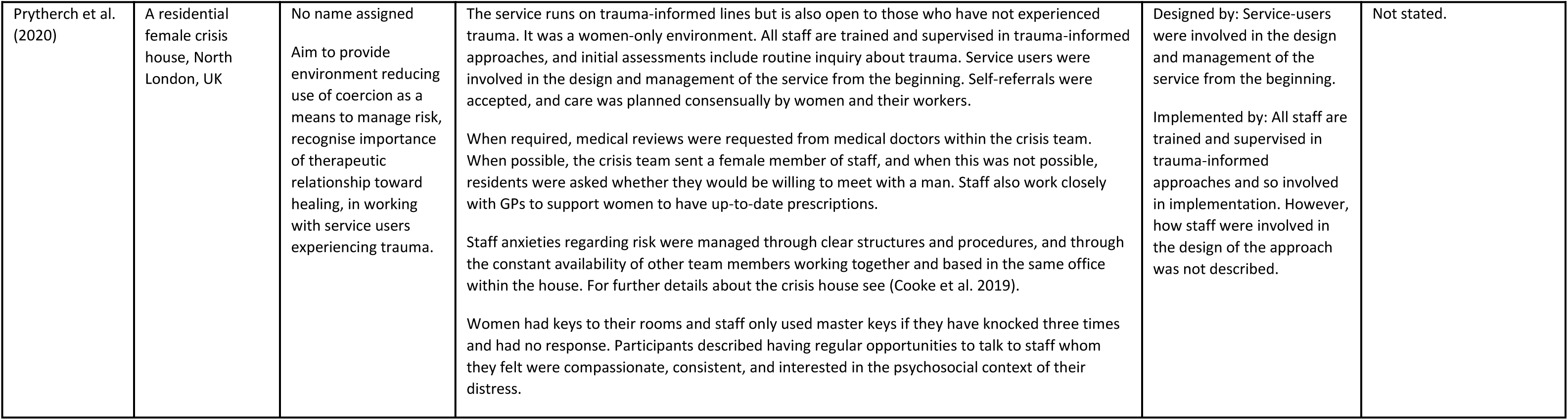
Trauma informed models

#### What trauma informed approaches are used in inpatient, crisis, emergency, and residential mental health care?

Thirty-one studies in twenty-seven different settings described the implementation of trauma-informed approaches at an organisational level in inpatient, crisis, emergency, and residential settings. The different models illustrate that the implementation of trauma informed approaches is a dynamic and evolving process which can be adapted for a variety of contexts and settings.

We have described the trauma informed approaches by model category and by setting. Child and adolescent only settings are reported separately. Summaries of the trauma informed approaches can be seen in Table 2. For full descriptions, see Appendix 3.

#### Six Core Strategies

Seven studies conducted in four different settings implemented the Six Core Strategies model of TIC practice for inpatient care (47, 48, 55–57, 62, 63). The Six Core Strategies were developed with the aim of reducing seclusion and restraint in a trauma informed way (83). The underpinning theoretical framework for the Six Core Strategies is based on trauma-informed and strengths-based care with the focus on primary prevention principles.

##### Inpatient settings: child and adolescent services

Four studies using Six Core Strategies were conducted in two child and adolescent inpatient settings using pre/post study designs (47, 48, 62, 63). Hale (2020) describes the entire process of implementing the intervention over a 6-month period and establishing a culture change by 12 months, while Azeem et al’s (2015) service evaluation documents the process of implementing the six strategies on a paediatric ward in the USA over the course of ten years.

##### Inpatient settings: adult services

Two studies focused on the use of the Six Core Strategies in adult inpatient and acute settings (55, 56). Duxbury (2019) adapted the Six Core Strategies for the UK context and developed ‘REsTRAIN YOURSELF’, described as a trauma-informed, restraint reduction programme, which was implemented in a non-randomised controlled trial design across fourteen adult acute and inpatient wards in seven hospitals in the UK (57).

##### How does the Six Core Strategies in inpatient, crisis, emergency, and residential mental health care impact on service user outcomes?

Five studies, only one of which had a control group (57), reported a reduction in restraint and seclusion practices after the implementation of the Six Core Strategies (47, 48, 57, 62, 63). Two studies (55, 56) did not report restraint and seclusion data.

##### Question 4: What is known about staff attitudes, expectations, and experiences of delivering the Six Core Strategies in inpatient, crisis, emergency, and residential mental health care?

Staff reported an increased sense of pride in ability to help people with a background of trauma (62), which came with the skills and knowledge development provided (56). Staff also showed greater empathy and respect towards service users (55, 56).

Staff recognised the need for flexibility in implementing the Six Core Strategies, and felt equipped to do this; as a result they reported feeling more fulfilled as practitioners (56). Staff shifted their perspectives on service users and improved connection with them by viewing them through a trauma lens (55). Staff also reported improved team cohesion through the process of adopting the Six Core Strategies approach (56). It was emphasised that to create a safe environment, role modelling by staff was required (55).

##### How does the Six Core Strategies impact on staff practices and staff wellbeing in inpatient, crisis, emergency, and residential mental health care?

Service users were reportedly more involved in their own care; they reviewed safety plans with staff, and were involved in their treatment planning, including decisions on medication (55, 56). Staff and service users also engaged in shared skill and knowledge building by sharing information, support, and resources on healthy coping, and trauma-informed care generally (55, 56). Staff adapted their responses to service user distress and adopted new ways of managing risk and de-escalating without using coercive practices (56, 62). Finally, service user-staff relationships were cultivated through a culture of shared learning, understanding, and trust (55).

##### How does the Six Core Strategies in inpatient, crisis, emergency and residential mental health care impact on service use and costs and what evidence exists about their cost-effectiveness?

One study reported a reduction in the duration of hospital admission (62), which was largely attributed to the reduction in the amount of documentation that staff had to complete following crisis interventions.

#### The Sanctuary Model

Six studies, referring to five different settings (58, 67, 68, 70–72), employed the ‘Sanctuary Model’ (84) as a TIC model of clinical and organisational change. One further study (67) combined the ‘Sanctuary Model’ with ‘Seeking Safety’, an integrated treatment programme for substance misuse and trauma. All studies were conducted in child and adolescent residential emotional and behavioural health settings in the USA.

The Sanctuary Model is a ‘blueprint for clinical and organisational change which, at its core, promotes safety and recovery from adversity through the active creation of a trauma-informed community’ (84). It was developed for adult trauma survivors in short term inpatient treatment settings and has formally been adapted for a variety of settings including adolescent residential treatment programmes. No studies explored the use of the Sanctuary Model as a TIC approach in adult inpatient or acute settings, and no studies specifically tested the efficacy of The Sanctuary Model in child and adolescent settings. Studies employing the Sanctuary Model used a range of designs including longitudinal, qualitative methods, service evaluations/descriptions and a non-randomised controlled design.

##### What is known about service user and carer expectations and experiences of The Sanctuary Model in acute, crisis and inpatient mental health care?

In a service description, Kramer (2016) reported that service users experienced clear interpersonal boundaries with staff, facilitated by healthy attachments and organisational culture. Service users also experienced staff responses as less punitive and judgemental.

##### How does The Sanctuary Model in acute, crisis and inpatient mental health care impact on service user outcomes?

Kramer (2016) described decreasing rates of absconscion, restraint, and removal of service users from the programme post-implementation of the Sanctuary Model, which the authors hypothesised was due to the safe environment, and the movement towards a culture of hope.

##### What is known about staff attitudes, expectations, and experiences of delivering The Sanctuary Model in acute, crisis and inpatient mental health care?

Staff received The Sanctuary Model positively (67, 70) and it gave them a sense of hopefulness (71). However, it was widely acknowledged that it was resource intensive for staff to implement TIC in practice and additional training may be needed (70).

Korchmaros et al., (2021) reported that the Sanctuary Model components most likely to be adopted were those that staff found most intuitive, though staff generally did not believe the model was acceptable in its entirety for their clinical setting. As staff communication improved, so did physical safety for staff and service users, and team meeting quality (71). Staff appreciated the improved safety – or perception of such – after implementing the Sanctuary Model (70, 71).

Staff reported that the Sanctuary Model promoted a healthy organisational culture with a commitment to a culture of social responsibility, social learning (68) and mutual respect (70).

##### How does The Sanctuary Model impact on staff practices and staff wellbeing in acute, crisis and inpatient mental health care?

Following implementation, staff focused more on service user recovery, through teaching service users adaptive ways of coping and encouraging empathy and compassion towards service users (70–72). Staff reported a holistic and compassionate understanding of the associations between trauma, adversity and service user behaviour (70). Staff also reported sharing more decisions with service users (70) and collaboratively involving service users in their treatment care, safety plans, and in the development of community rules (70, 71). Staff felt able to communicate information, ideas, and their mistakes more openly, and their ability to model healthy relationships was seen as a fundamental to treatment (71).

##### How does The Sanctuary Model in acute, crisis and inpatient mental health care impact on service use and costs and what evidence exists about their cost-effectiveness?

One study reported a reduction in staff compensation claims due to a reduction in harms from physical interventions (67).

#### Comprehensive tailored trauma informed model

We categorised eight models (51, 53, 54, 59, 60, 73–75) as ‘comprehensive tailored TIC models’. This reflects models that are holistic and multi-faceted, but do not closely follow an established TIC intervention model or blueprint, and have instead been developed locally or to specifically complement the needs of a specific setting.

##### Residential settings: child and adolescent services

Three studies describe comprehensive tailored trauma informed models in child and adolescent residential treatment settings (51, 53, 59). At an organisational level, key features of the tailored approaches include whole staff training using a trauma-informed curriculum (51, 53, 59), creating a supportive, therapeutic environment and sense of community, using a family-centred approach, structured programmes of psychoeducational, social and skills groups (51, 59) and collaborative working across different agencies (53, 59).

##### Residential settings: adult women-only substance misuse service

Three studies (54, 74, 75) describe a gender-specific trauma-informed treatment approach in a female only environment. Key components include providing training for all staff on trauma and trauma informed approaches, team support and supervision from a trained trauma therapist, social and life skills groups (74); individual therapy and family support programme (75); trauma-informed group therapies and psychoeducation (54, 74); the implementation of a ‘gender specific, strengths based, non-confrontational, safe nurturing environment’ and strategic level meetings to identify structural barriers and gaps across different agencies (54). Zweben et al. (2017) focused specifically on family-centred treatment for women with severe drug and alcohol problems (75).

##### Inpatient settings: adults and children/adolescents

The ‘Patient Focussed Intervention Model’ (60) was implemented in a variety of settings including residential child and adolescent, adult acute and adult longer term inpatient stays. This model was developed using a collaborative process with involvement from service users, staff, administrators, and external collaborators with continuous quality improvement. The model incorporated i) an individual TIC treatment model, which emphasised a patient-centred approach to building a culture and environment which is soothing and healing (e.g., by making changes to the physical environment and providing animal assisted therapy), ii) the Sorensen and Wilder Associates (SWA) aggression management program (85) and iii) staff debriefing after incidents of aggression.

##### Inpatient settings: women-only forensic service

Stamatopoulou (2019) used qualitative research methods to explore the process of transitioning to a TIC model in a female forensic mental health unit (73) using the ‘trauma informed organisational change model’ (86). Components included i) staff training on trauma-informed care, ii) co-produced safety planning through five sessions of Cognitive Analytic Therapy (82), iii) a daily ‘Trauma Champion’ role for staff, and iv) reflective practice groups for staff and service users.

##### What is known about service user and carer expectations and experiences of comprehensive tailored trauma informed models in inpatient, crisis, emergency, and residential mental health care?

In a study by Tompkins & Neale (2018), service users reported being unaware that the service they were attending was trauma informed and were therefore not anticipating being encouraged to confront and reflect on their traumas (74). However, the daily structure and routine timetable created a secure environment for their treatment experience. Service users who remained engaged with the service felt cared for by staff, and that the homely atmosphere in the service supported them to feel secure.

##### How do comprehensive tailored trauma informed models in inpatient, crisis, emergency and residential mental health care impact on service user outcomes?

Three studies reported data on seclusion and restraints; all indicated a decrease (53, 60, 75). Brown et al. (2013) quantitatively reported reductions in the use of both seclusion and restraint in the year following Trauma Systems Therapy implementation, and the reduction in use of physical restraints was significant and sustained over the following eight years. Goetz et al. (2012) also quantitatively reported that seclusion and restraint rates halved following implementation of the Patient Focused Intervention Model, as well as reductions in: i) staff injuries, ii) hours in seclusion and restraint and iii) the number of aggressive patient events. However, while the number of seclusion room placements was lower, the average number of restraints among children and young people was higher in TIC compared to usual care.

Using a pre-post design, Zweben et al. (2017) reported that service users receiving TIC reported fewer psychological and emotional problems after a month, compared to on entry to the programme, as well as reduced drug and alcohol misuse. Service user average length of stay was higher among those in the programme, compared to those not enrolled. Finally, as court and protective services became aware of the service users’ improvements, reunifications with children approached 100%.

##### What is known about staff attitudes, expectations, and experiences of delivering comprehensive tailored trauma informed models in inpatient, crisis, emergency, and residential mental health care?

Two papers reported on staff attitudes and experiences. Some staff experienced introduction of TIC as overwhelming, leaving them feeling unsure about what was expected from them throughout the training and implementation process, and that it might have been more successful had it been implemented in stages (73). Some staff also felt unsure of what TIC entailed even after training, and it took time for staff to feel competent and confident (74). Staff who had the least awareness of TIC experienced the greatest change anxiety when they were told of its implementation. When there were conflicting views within the team, the consistency of implementation was reduced (73). Conversely, however, staff reported a sense of achievement in having implemented TIC and consequently an increased sense of job satisfaction. Goetz et al. (2012) also reported reduced staff injuries in the first year after implementing TIC.

Staff reported that their own traumatic experiences can inform the way they communicate and react to situations in the clinical environment, and they needed help to set boundaries and avoid emotional over involvement (74). Implementing TIC broke down barriers between their private and professional selves, and there was an increased awareness of the personal impact of their work.

##### How do comprehensive tailored trauma informed models impact on staff practices and staff wellbeing in inpatient, crisis, emergency, and residential mental health care?

Stamatopoulou (2019) reported that with the introduction of TIC, staff moved away from a solely diagnosis-based understanding of distress and developed an ability to formulate connections between service users’ backgrounds and their clinical presentations. As a result, staff showed empathy and respect towards service users and approached sensitive situations with service users more mindfully. Staff adopted new ways of managing risk (73) and service users were collaboratively involved in the development of treatment care plans (74).

There was an increased focus on staff wellbeing as well as greater awareness of staff personal boundaries and experiences of trauma and adversity, which led to staff feeling as though they had more in common with service users than expected (73). Staff also discussed the importance of protecting their wellbeing by maintaining personal and professional boundaries, practicing mindfulness, and attending mutual aid groups (74).

Wider group relationships were reportedly redefined, and the impact of the work on staff wellbeing was acknowledged, specifically the ways in which staff’s own trauma informed their reactions to incidents on the ward (73). Staff reported a greater sense of team connectedness, and that their individual professional identities were reconstructed.

##### How do comprehensive tailored trauma informed models in inpatient, crisis, emergency and residential mental health care impact on service use and costs and what evidence exists about their cost-effectiveness?

Staff had concerns over the sustainability of the model, especially in an organisation that could not offer the time or money required for service development (73). Fidelity to a model varied depending on which staff were working; for instance, when agency staff or new joiners were working, fidelity was low and crisis incidents increased.

Adolescents in residential treatment receiving TIC spent significantly less time in treatment compared to those receiving traditional treatment, with the receipt of trauma informed psychiatric residential treatment (TI-PRT) accounting for 25% of the variance in length of stay (51). Similarly, authors observed reduced length of hospital admissions after the introduction of their TIC approach (59).

#### Safety focused tailored trauma-informed models

Two studies (50, 52) tailored their own trauma-informed approach to create a culture of safety and i) reduce restrictive practices and ii) staff injuries. A third study (66) utilised a Trauma and Self Injury (TASI) programme, which was developed in the National High Secure Service for Women.

##### Inpatient settings: adults and child/adolescent services

Blair et al. (2017) conducted a pilot trauma-informed intervention study with the aim of reducing seclusions and restraints in a psychiatric inpatient hospital facility. Intervention components included the use of Broset Violence Checklist (BVC) (87–89); 8-hour staff training in crisis intervention; two-day training in “Risking Connections” (90); formal reviewing of restraint and seclusion incidents; environmental enhancements; and individualised plans for service users.

Borckardt et al (2011) reported a ‘trauma informed care engagement model’ to reduce restraint and seclusion across five various inpatient units (acute, child and adolescent, geriatric, general and a substance misuse unit). The intervention components included TIC training, changes in rules and language to be more trauma-sensitive, patient involvement in treatment planning and physical changes to the environment.

##### Inpatient settings: women-only forensic service

Jones (2021) reported the TASI programme in a high security women’s inpatient hospital, which aimed to manage trauma and self-injury with a view to reducing life threatening risks to service users and staff. The programme: i) promotes understanding of trauma through staff training, staff support and supervision on the wards, ii) provides psychoeducation and wellbeing groups for service users, iii) focuses on the improvement of the therapeutic environment, iv) promotes service users’ ability to cope with their distress, and v) provides individual and group therapy.

##### What is known about service user and carer expectations and experiences of safety focused tailored trauma-informed models in inpatient, crisis, emergency, and residential mental health care?

In Jones (2021), some women reported feeling initially overwhelmed by and underprepared to acknowledge and work on their trauma experiences, and sometimes felt their distress was misunderstood by the nurses in their setting, which could lead to an escalation (66). Service users felt connected to themselves, to feel safe and contained, particularly in comparison to previous inpatient experiences. Overall, however, the service did not make the women feel as though their problems were fully understood.

##### How do safety-focused tailored trauma-informed models in inpatient, crisis, emergency, and residential mental health care impact on service user outcomes?

Reductions in the number of seclusions and restraint were reported (50, 52). Specifically, the trauma informed change to the physical therapeutic environment was associated with a reduction in the number of both seclusion and restraint (52). The duration of restraints reportedly increased, while duration of seclusion decreased (50).

##### What is known about staff attitudes, expectations, and experiences of delivering safety focused tailored trauma-informed models in inpatient, crisis, emergency, and residential mental health care?

In Jones (2021), nurses emphasised that shared understanding and trust were instrumental in connecting and communicating with the women in their service (66). Nurses also cultivated service user-staff therapeutic relationships, which they experienced as intensely emotional. The nurses reported becoming more critical of other staff members they perceived as lacking compassion towards the service users. Finally, staff felt that a barrier to the nurse-service user relationship was the staff’s inability to share personal information and vulnerability back to the service user.

##### How do safety focused tailored trauma-informed models impact on staff practices and staff wellbeing in inpatient, crisis, emergency, and residential mental health care?

Staff developed new tools and adapted their responses to trauma and distress, and service users were also involved in some, but not all, staff training (66). There were changes to information sharing practices (73); information on service users’ history and intervention plans were more openly shared within the team to ensure all staff had the same information and did not need to risk re-traumatising service users by asking for information again. Service users also reported being more involved in their treatment planning (52, 56) and medication decisions (55).

#### Trauma informed training intervention

Three studies focused on trauma informed training interventions for staff (46, 61, 69). All studies used a pre/post design to evaluate effectiveness.

##### Residential settings: child and adolescent services

Gonshak (2011) reported on a specific trauma-informed training programme called Risking Connections (90) implemented in a residential treatment centre for children with ‘severe emotional disabilities’. Risking Connections is a training curriculum for working with survivors of childhood abuse and includes i) an overarching theoretical framework to guide work with trauma and abuse ii) specific intervention techniques iii) a focus on the needs of trauma workers as well as those of their clients.

##### Inpatient settings: adult services

Nimura (2019) reported a 1-day TIC training intervention, covering items including the definition of trauma, evidence on trauma and behavioural, social, and emotional responses to traumatic events, on attitudes of staff in a psychiatric hospital setting. Aremu (2018) reported a training intervention to improve staff engagement which they identified as a key component of TIC. The intervention was a 2-hour training on engaging with patients, however the specific content of the training is not specified.

##### What is known about staff attitudes, expectations, and experiences of delivering trauma informed training interventions in inpatient, crisis, emergency, and residential mental health care?

Following training, half of staff reported feeling their skills or experience were too limited to implement changes and also that staff experienced difficulties when trying to share the principles of trauma informed care with untrained staff (69). However, TIC training did produce positive attitude shifts towards TIC.

##### How do trauma informed training interventions impact on staff practices and staff wellbeing in inpatient, crisis, emergency, and residential mental health care?

Staff modified their communication with service users by altering their tone and volume, and adopted new ways of managing risk without using coercive practices (69). There was also a reported a reduction in the use of as-needed medication (46).

##### How do trauma informed training interventions in inpatient, crisis, emergency, and residential mental health care impact on service use and costs and what evidence exists about their cost-effectiveness?

One study reported that educating staff about TIC in residential settings was time intensive and may require frequent or intense “booster” sessions following the initial training (61). In terms of evaluation, it took time to implement TIC in the residential setting and then to collect related outcome data.

#### Other TIC models

This category reports studies which have made attempts to shift the culture towards trauma-informed approaches but have not made full-scale clinical and organisational changes to deliver a comprehensive trauma-informed model (28, 49, 64, 65).

##### Inpatient settings: adult services

Isobel and Edwards’ (2017) case study on an Australian inpatient acute ward described TIC ‘as a nursing model of care in acute inpatient care’. This intervention was specifically targeted at nurses working on acute inpatient units but did not involve members of the multi-disciplinary team.

Beckett (2017) conducted a TIC improvement project on an acute inpatient ward (which primarily received admissions through the hospital emergency department) using workforce development and a participatory methodology. Staff devised workshops on trauma-informed approaches, through which six key practice areas were identified for improvement by the staff team.

Jacobowitz et al (2015) conducted a cross-sectional study in acute psychiatric inpatient wards to assess the association between TIC meetings and staff PTSD symptoms, resilience to stress, and compassion fatigue. Neither the content nor the structure of the ‘trauma-informed care meetings’ were described in further detail.

##### Crisis house setting

Prytherch, Cooke & March’s (2020) qualitative study of a ‘trauma informed crisis house’, based in the UK, gives a partial description of how trauma-informed approaches were embedded in the service delivery and design.

##### What is known about service user and carer expectations and experiences of other TIC models in inpatient, crisis, emergency, and residential mental health care?

Prytherch, Cooke and March (2020) reported that their TIC model created a positive experience for service users by making them feel worthwhile, respected and heard by staff (28). Service users valued being trusted by staff, for example, to have their own room keys and to maintain their social and occupational roles, which were a key source of self-worth. While service user experiences were often positive, they found being asked directly about trauma challenging. This was invalidating for those who did not initially identify as having experienced trauma. Some service users also felt the TIC programme could only support them so far, as it did not incorporate or address wider societal injustices, such as issues related to housing and benefits, that can contribute to and exacerbate distress.

##### How do other TIC models in inpatient, crisis, emergency, and residential mental health care impact on service user outcomes?

Beckett et al. (2017) reported that in the three years after the TIC workshops, rates of seclusion dropped by 80%, as did the length of time spent in seclusion, with most seclusions lasting less than an hour.

##### What is known about staff attitudes, expectations, and experiences of delivering other TIC models in inpatient, crisis, emergency, and residential mental health care?

Staff found an increased sense of confidence and motivation in managing emotional distress and behavioural disturbance (49). Staff also showed increased respect, understanding and compassion towards service users.

Similarly, staff expressed hope for improved care in future, on the basis that they were better skilled to deliver care that was consistent and cohesive, while also being individual and flexible (64). They reported a need for clarity and consistency in their role expectations and felt that changes in practice needed to be introduced slowly and framed positively. Conversely, others felt the changes introduced were too minimal to be significant and did not much vary from existing practice. Some staff expressed a fear of reduced safety that could follow changes to longstanding practice. Ambivalence to change stemmed from different degrees of understanding of TIC among staff, and others felt personally criticised by the newly introduced approach, specifically that their previous practice had been labelled traumatising.

##### How do other TIC models impact on staff practices and staff wellbeing in inpatient, crisis, emergency, and residential mental health care?

Some staff modified their communication with service users, e.g., reducing clinical jargon and focusing on the strengths of the service users (49). Regular opportunities for service users and staff to discuss and reflect on information, concerns, and experiences were established. Staff received training in physical safety and de-escalation procedures; subsequently, the need for security staff on the ward decreased. Finally, in terms of staff wellbeing, staff PTSD symptoms increased with an increase in signs of burnout and length of time between attending trauma-informed care meetings (65).

##### Implementation of TIC

###### What are the reported difficulties in implementing TIC?

Challenges to successful implementation included uncertainty, ambivalence and lack of interest in TIC from staff members (64, 69, 73), a lack of time (71), low readiness to change (66)and a lack of enthusiasm and commitment by staff (72), alongside power imbalances between staff and service users (66). Additional challenges included inadequate TIC knowledge among staff, the provision of staff training around TIC (particularly the delivery of additional training for new starters), and the implementation of staff supervision (70, 71, 74); these barriers arose as a result of insufficient time and money to support service development (73). At times there was a lack of understanding of TIC among staff, and a feeling that some models were vague (e.g., The Sanctuary model) (70). Practice changes in relation to staff working with families, and not attributing blame or increasing their feelings of guilt, was noted as challenging (72).

There were also longer-term concerns. Stamatopoulou et al. (2019) reported that TIC required significant resources, which may be draining in the long term, particularly as these services are likely to already be struggling to secure the limited resources available to them. Services may prefer instead to implement approaches which are less resource intensive to embed within their existing service structures. Similarly, maintaining the presence of a stable and well-trained team around TIC in the long term is challenging during periods of staff absence and high staff turnover (70); a key principle of TIC is to provide consistency to service users (74).

##### What is required to implement TIC effectively?

Senior staff buy-in and commitment and enthusiasm about TIC was deemed a critical first step for successful implementation (72). Slow, positively framed, and well-managed implementation is necessary to allow for a smooth transition to TIC, and staff and service users benefit from having clearly defined staff roles within TIC services (64). Positive role modelling by senior staff, such as a ward manager, was also key for successful implementation (55).

The provision of high-quality TIC training for existing staff and new starters was noted as integral to implementation (70, 71, 74). Staff must then be given sufficient time to become competent and feel comfortable delivering TIC, while ongoing support, mentoring and supervision is delivered to strengthen the development and maintenance of TIC skills (74). Specialist training to provide, for example, gender-sensitive (54) and culturally-sensitive services (49) may also result in a better adapted service. The presence of consistent, trained staff helped to support regular delivery of TIC to service users (73). TIC implementation can be further supported through peer support among staff, for example providing reflective and debrief spaces (73) and peer learning (56, 63).

Finally, it was suggested that during staff recruitment, it may be pertinent to consider each candidate’s background and experiences, and how these may complement the needs of the service users within each individual service, e.g., female staff with personal experiences which mirror those of the women in the service, or female staff who had worked with a similar service user group previously (74).

**Table 3.**
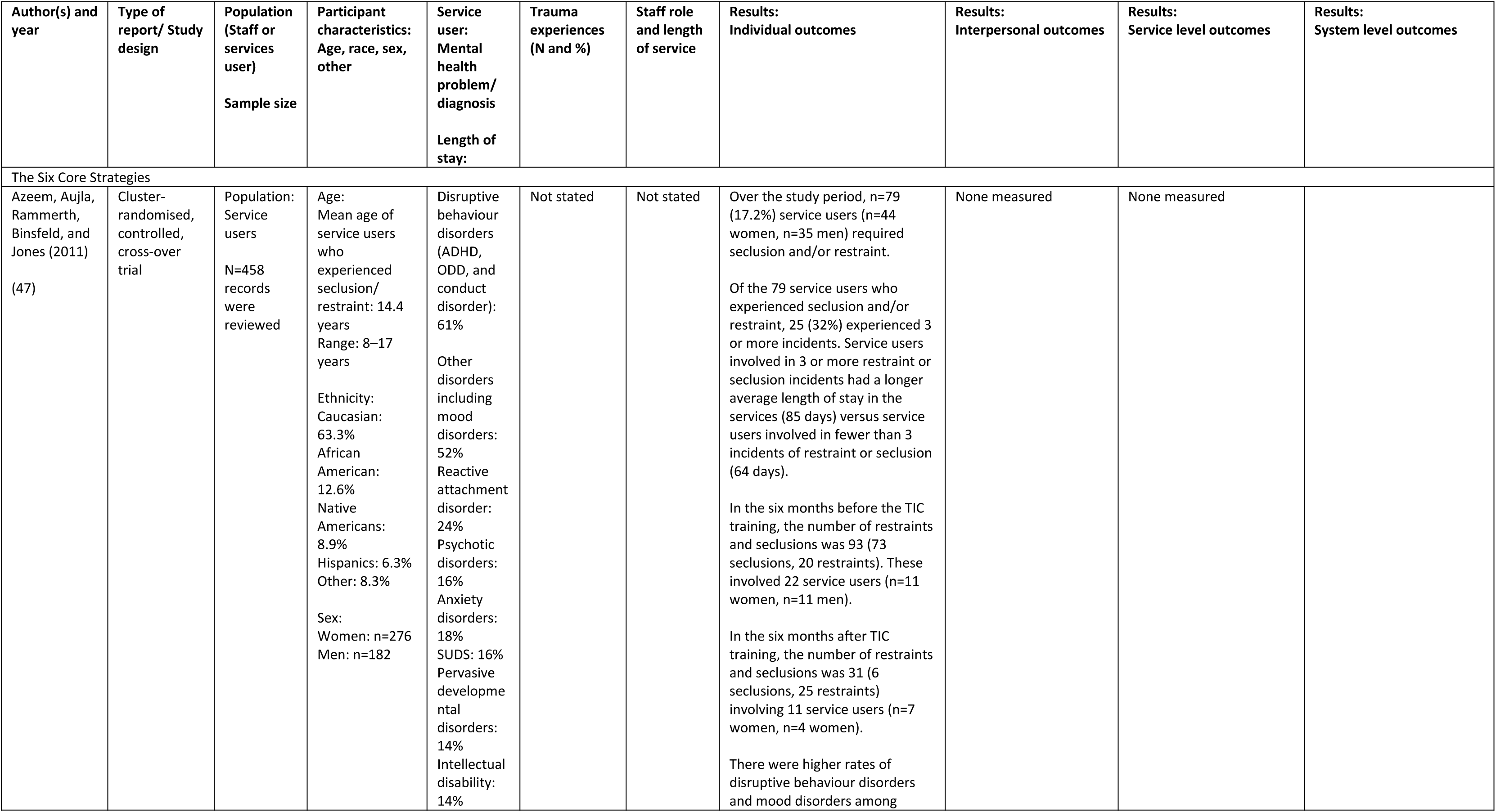
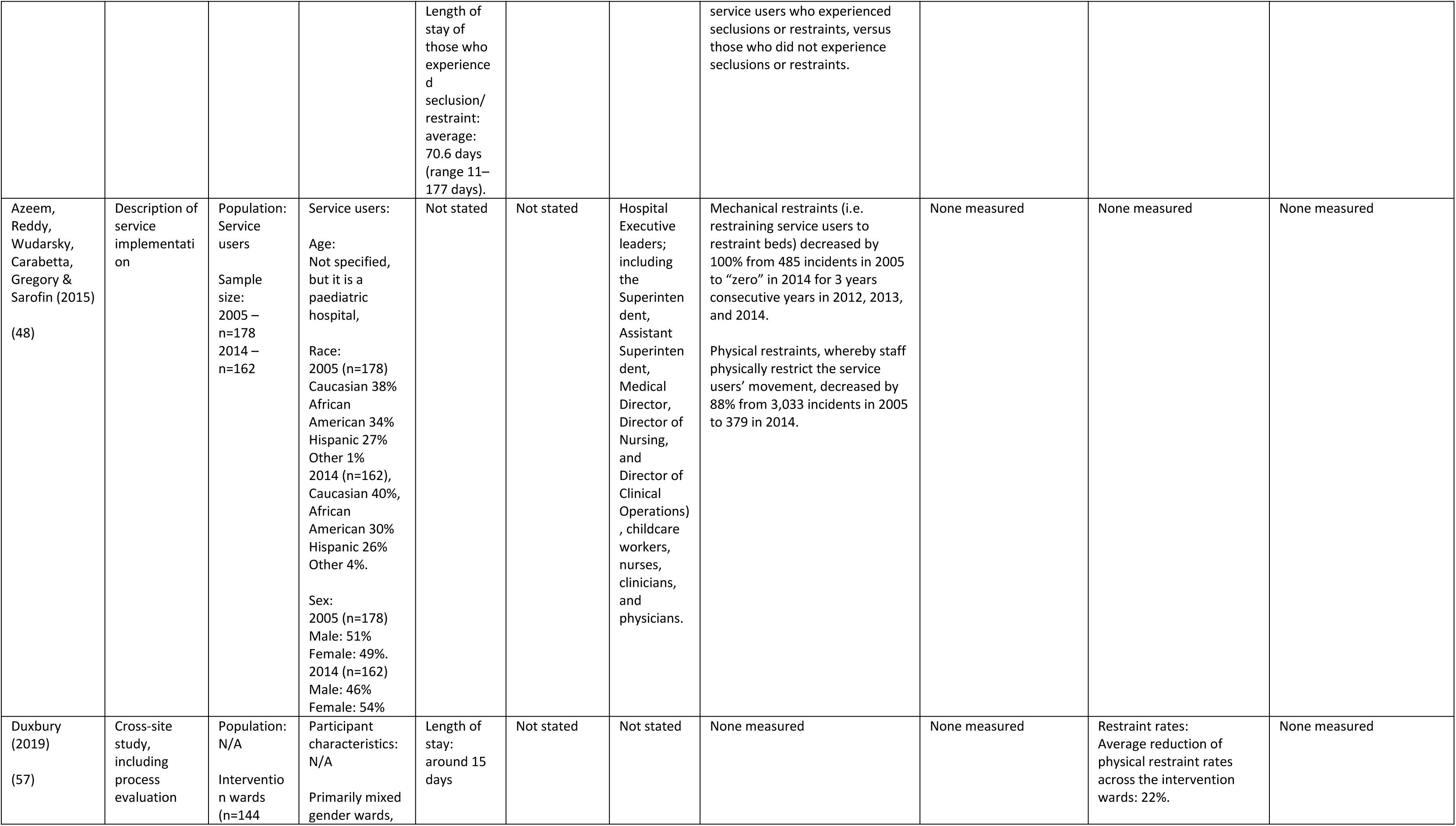
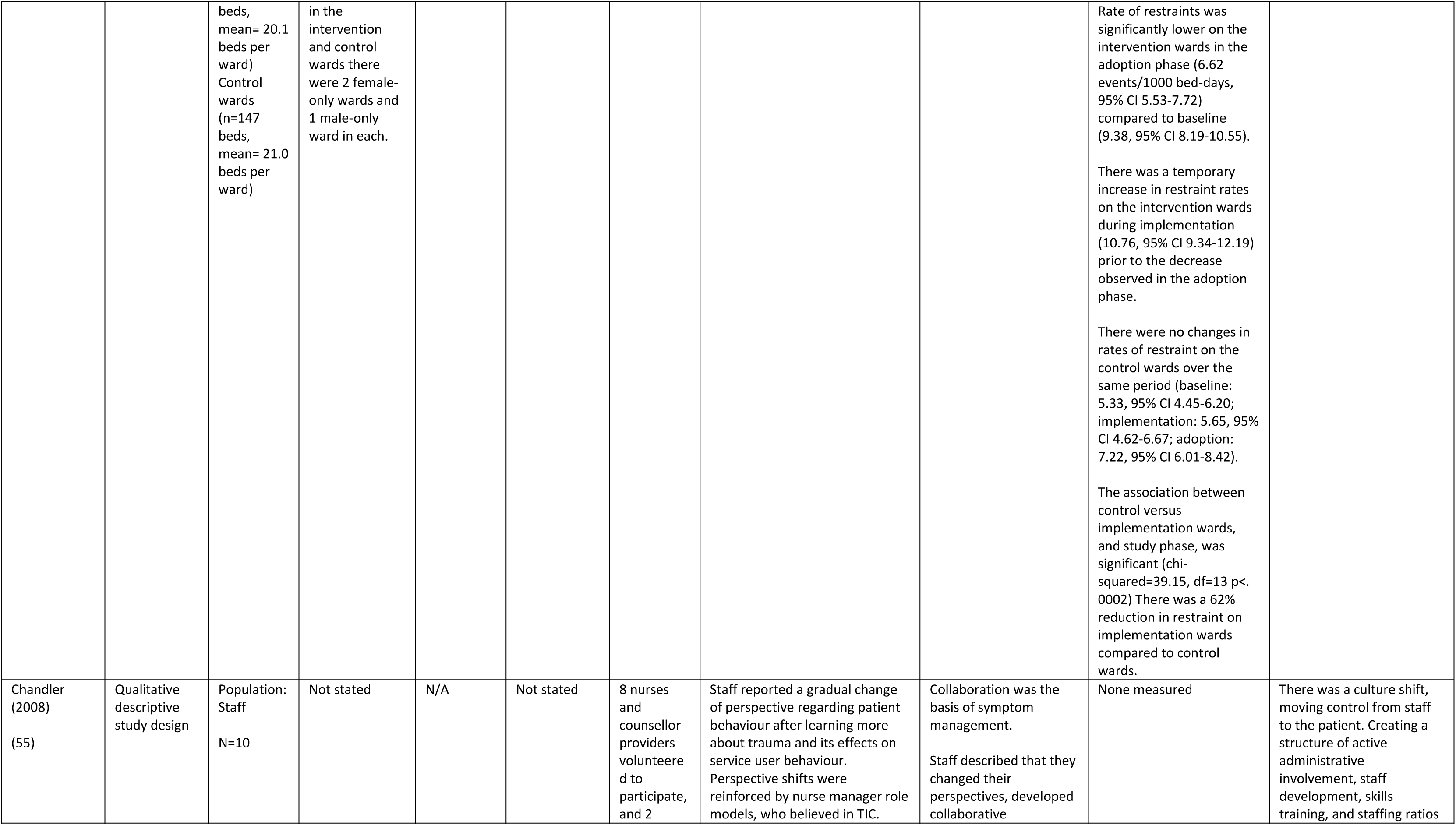
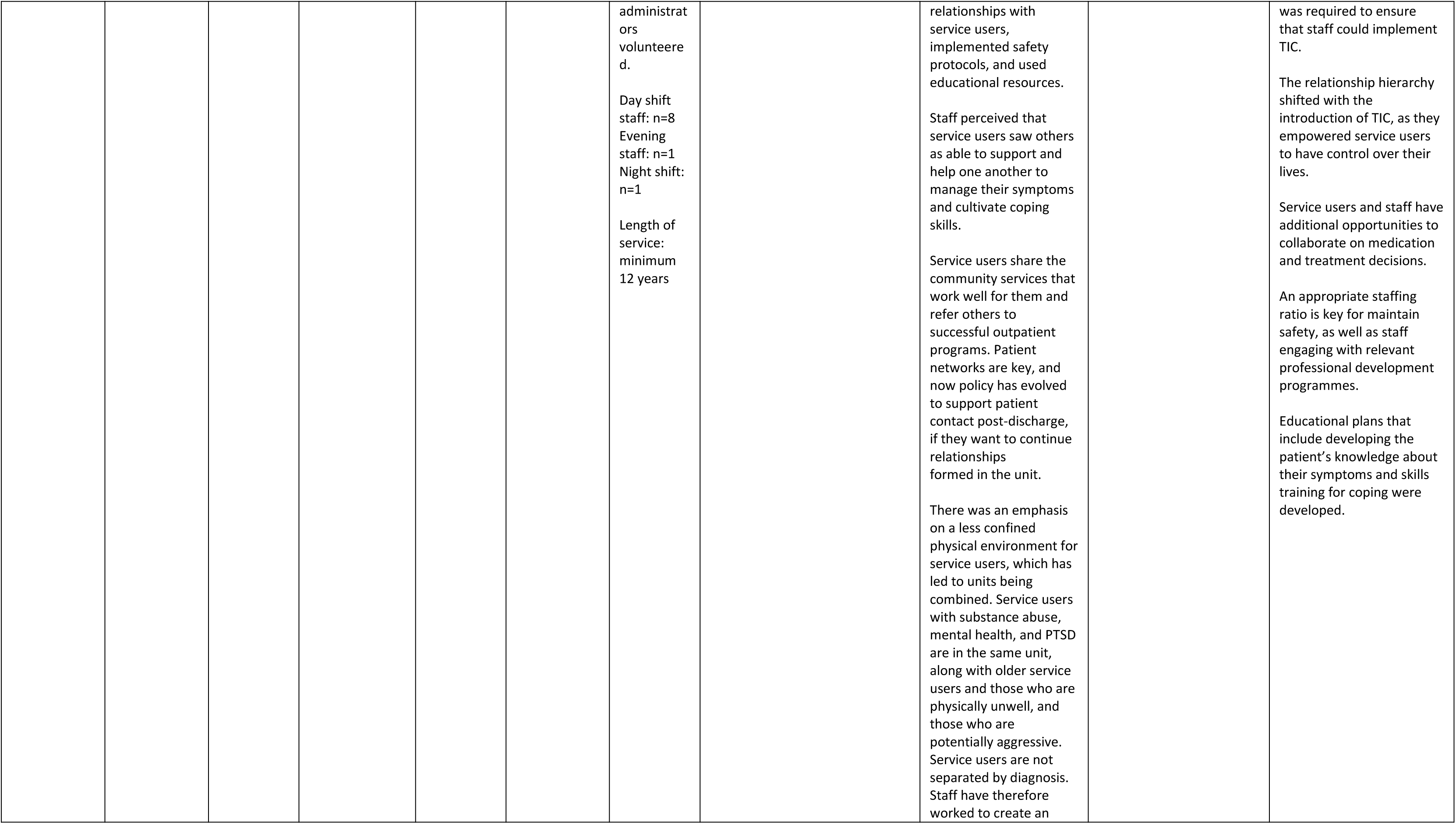
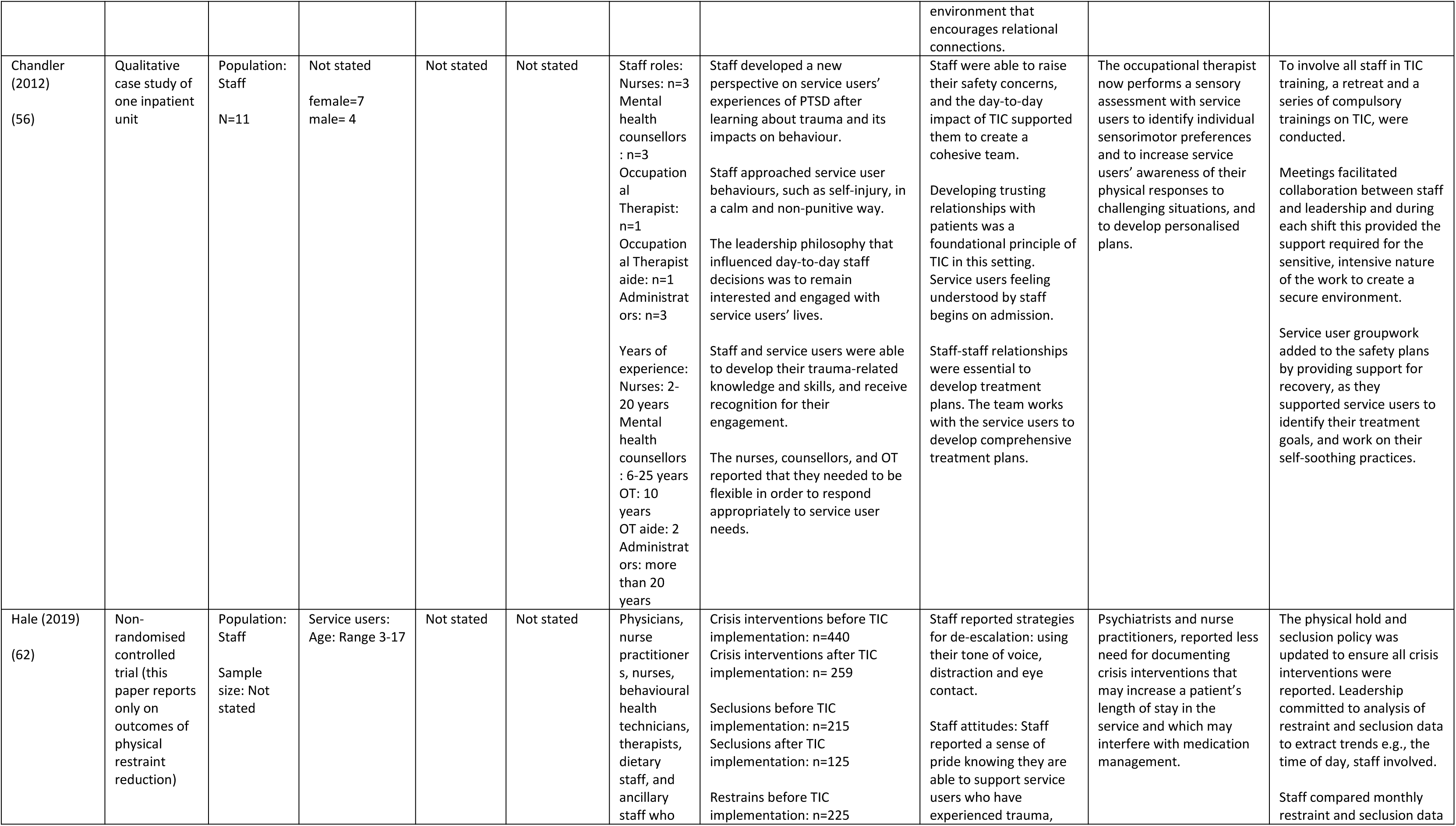
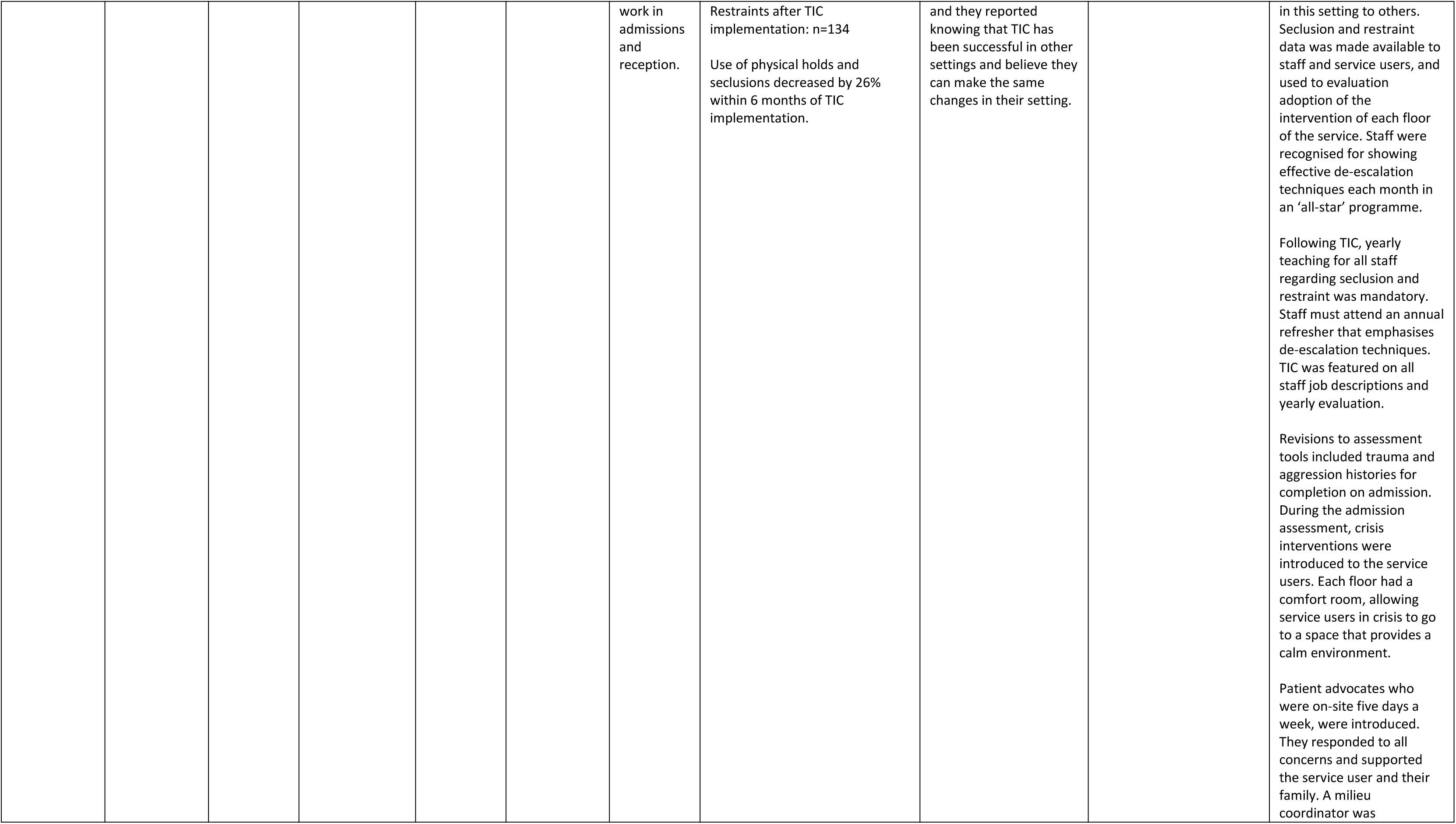
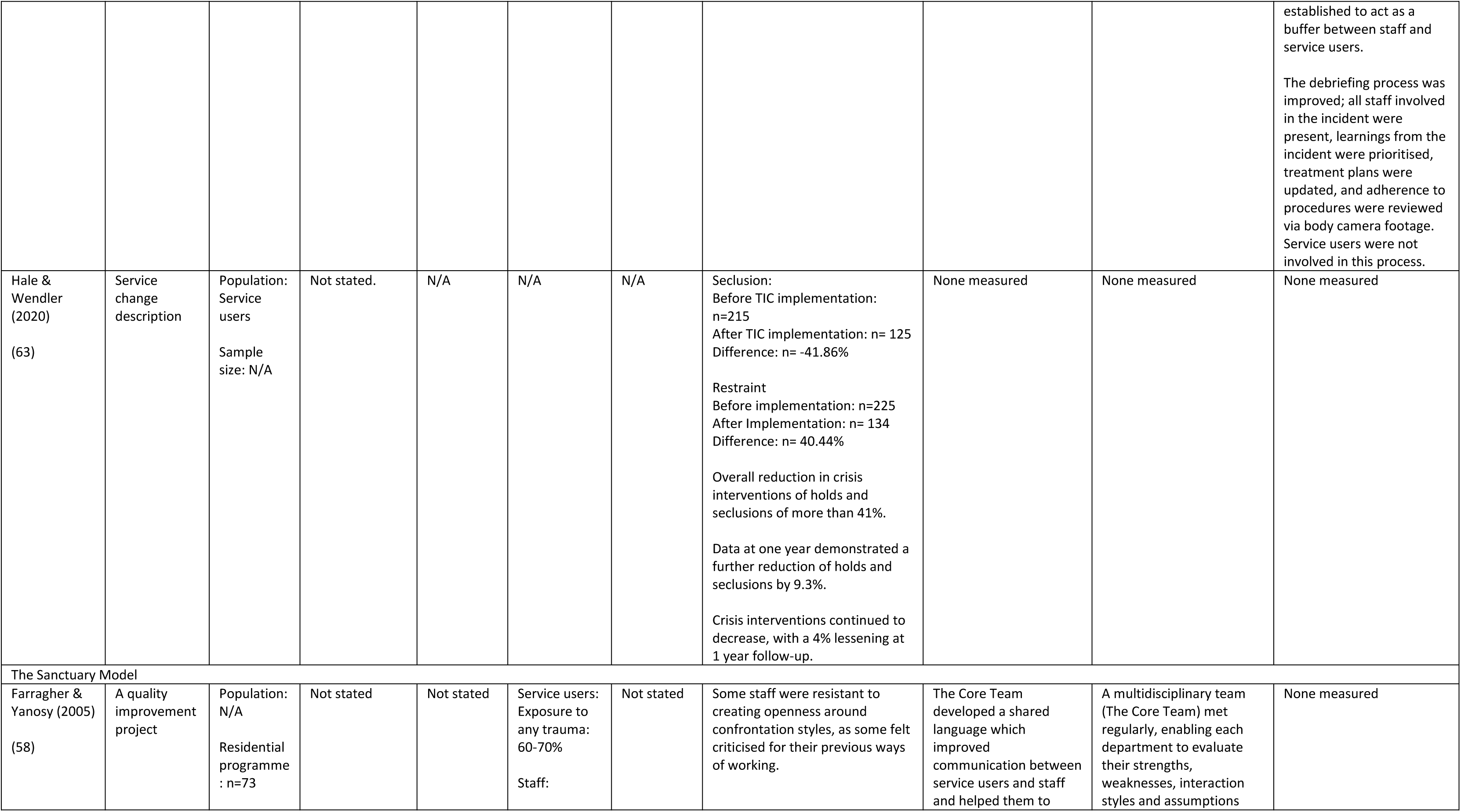
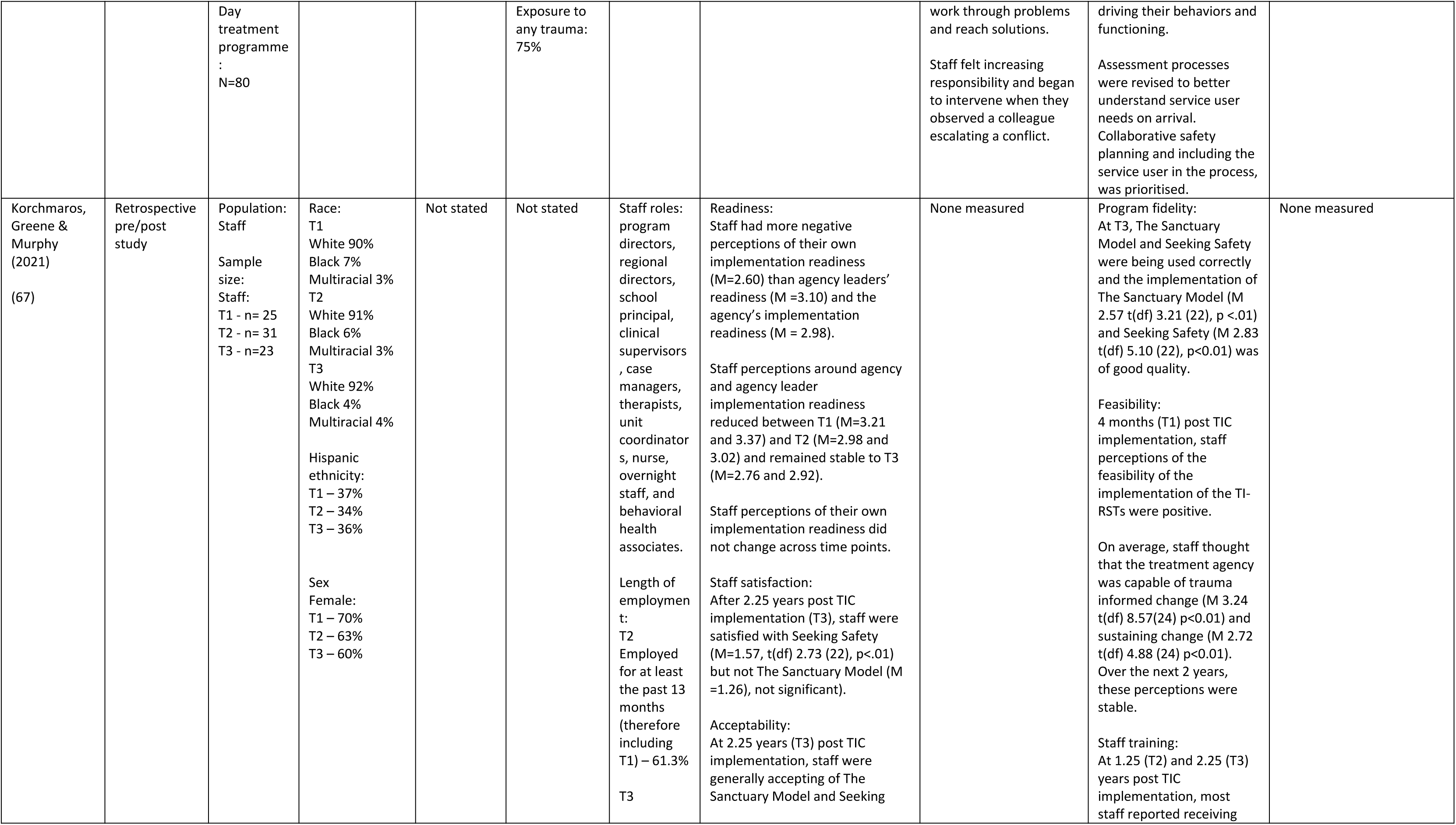
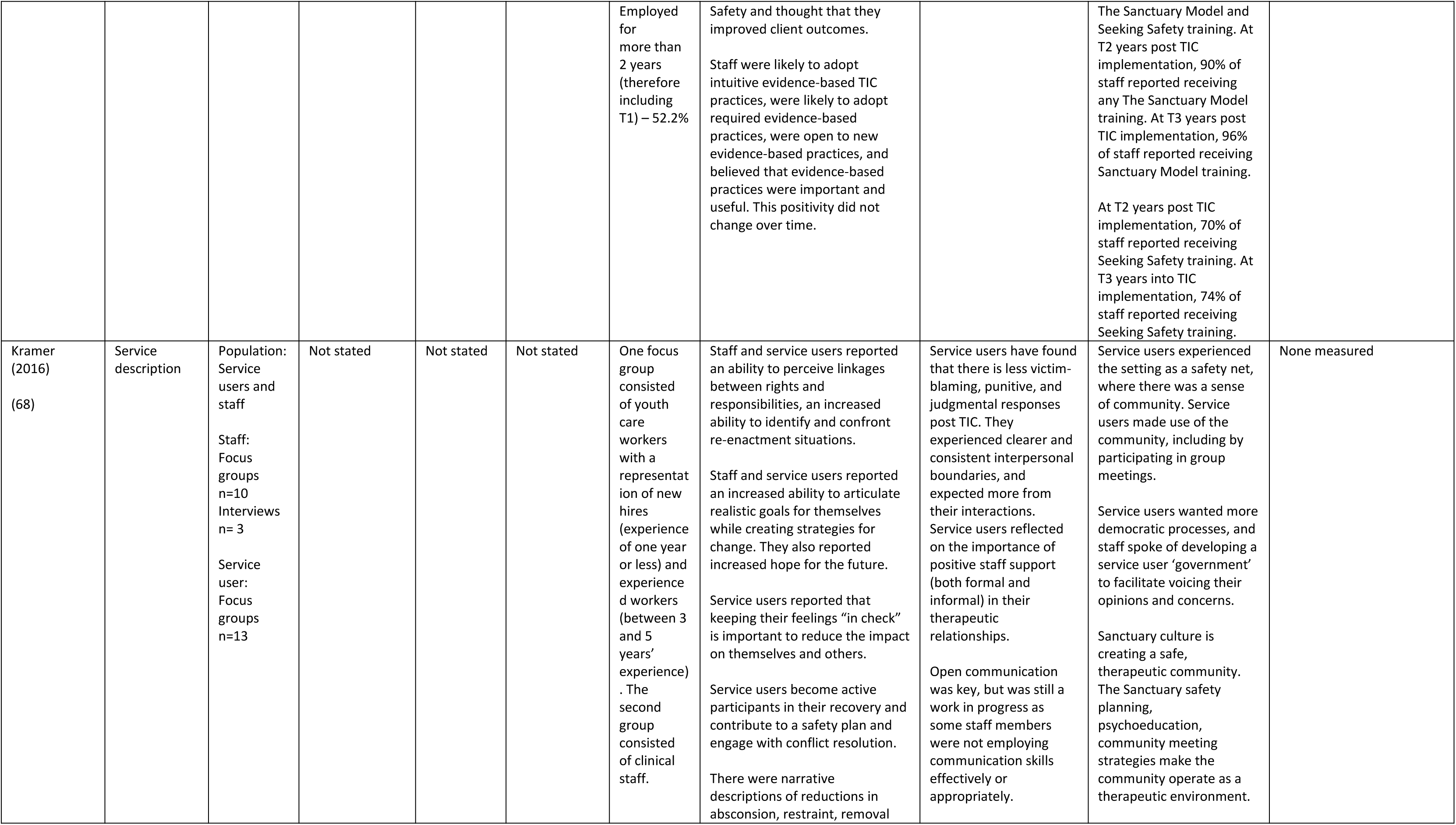
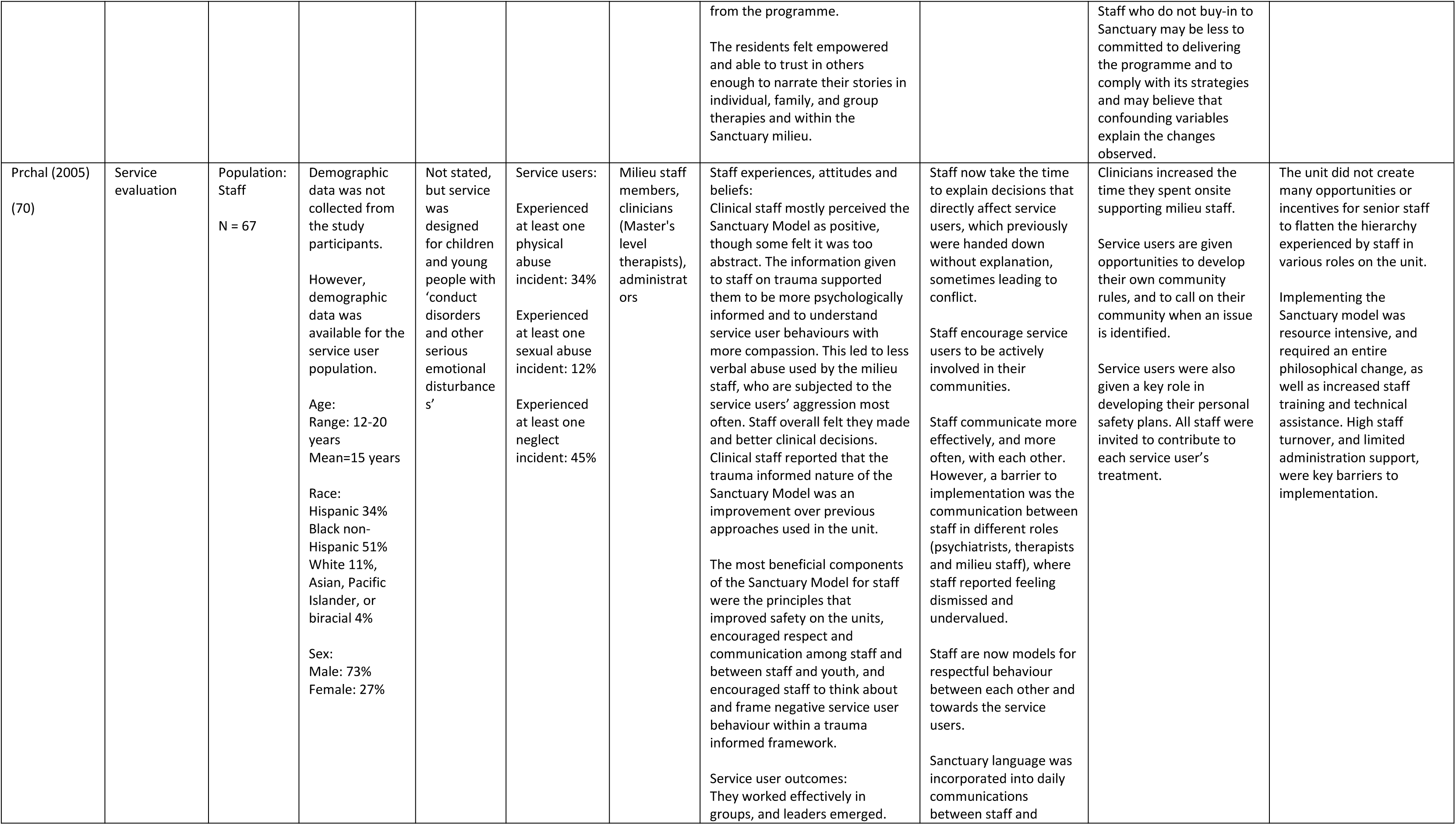
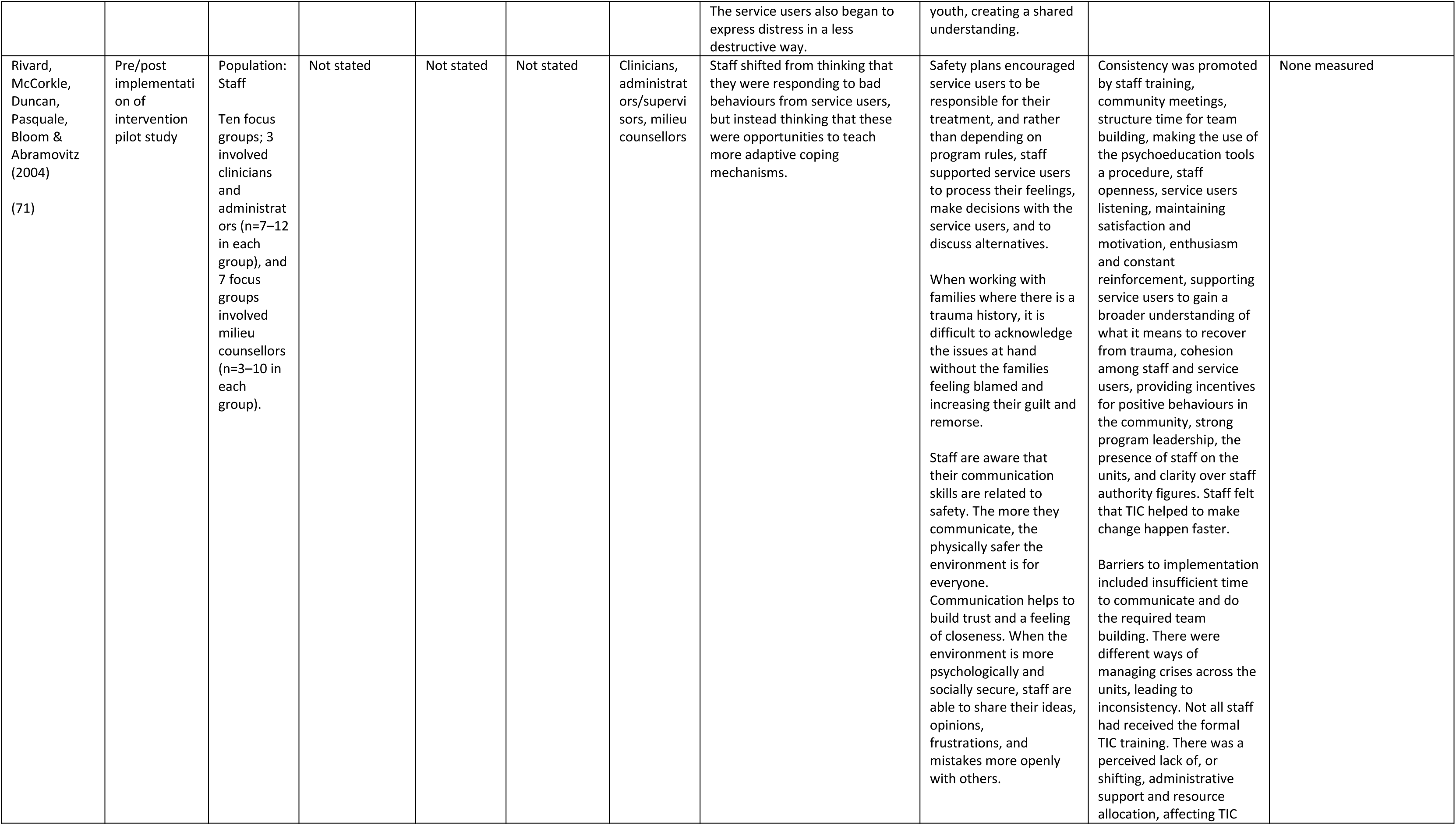
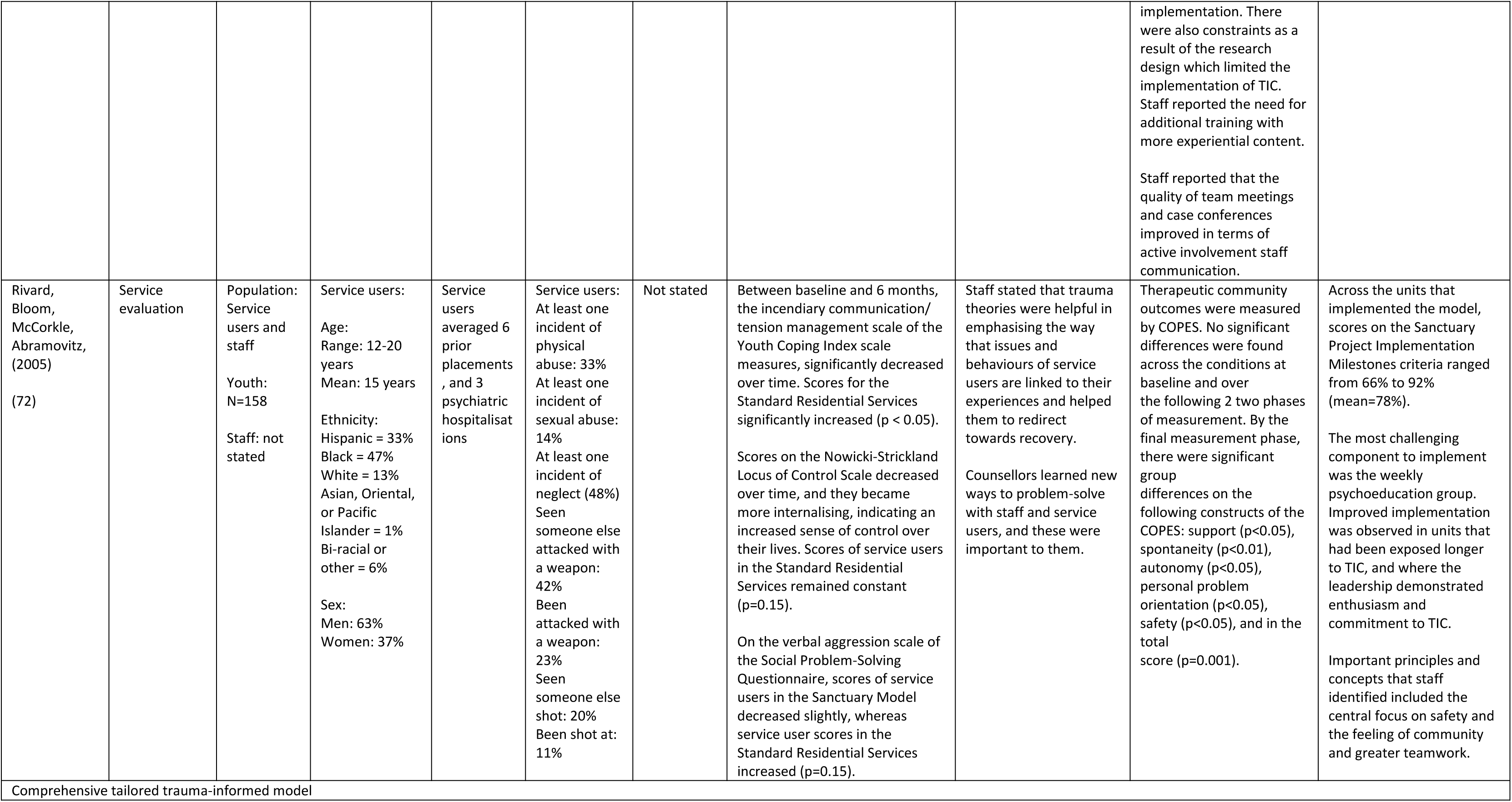
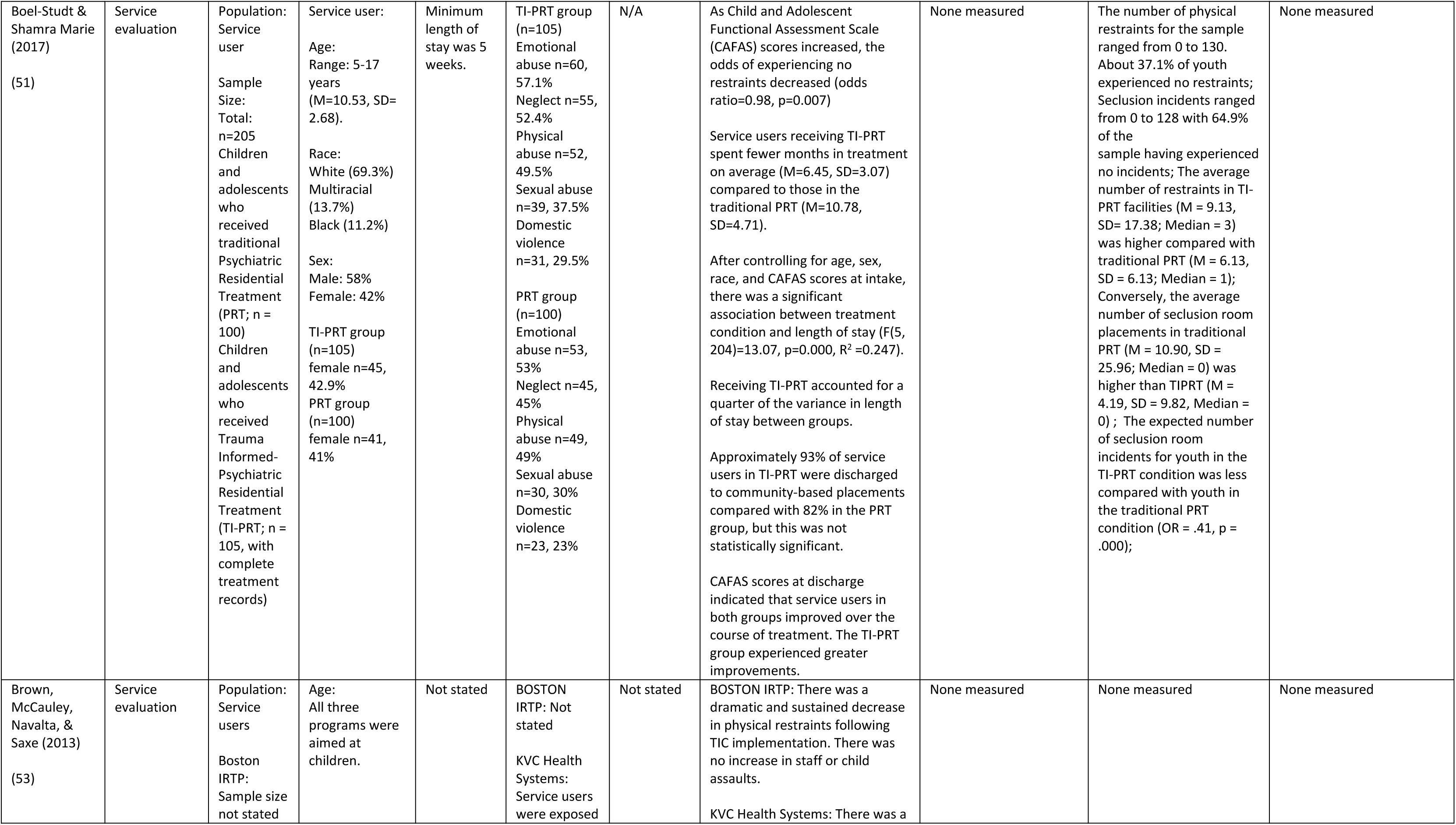
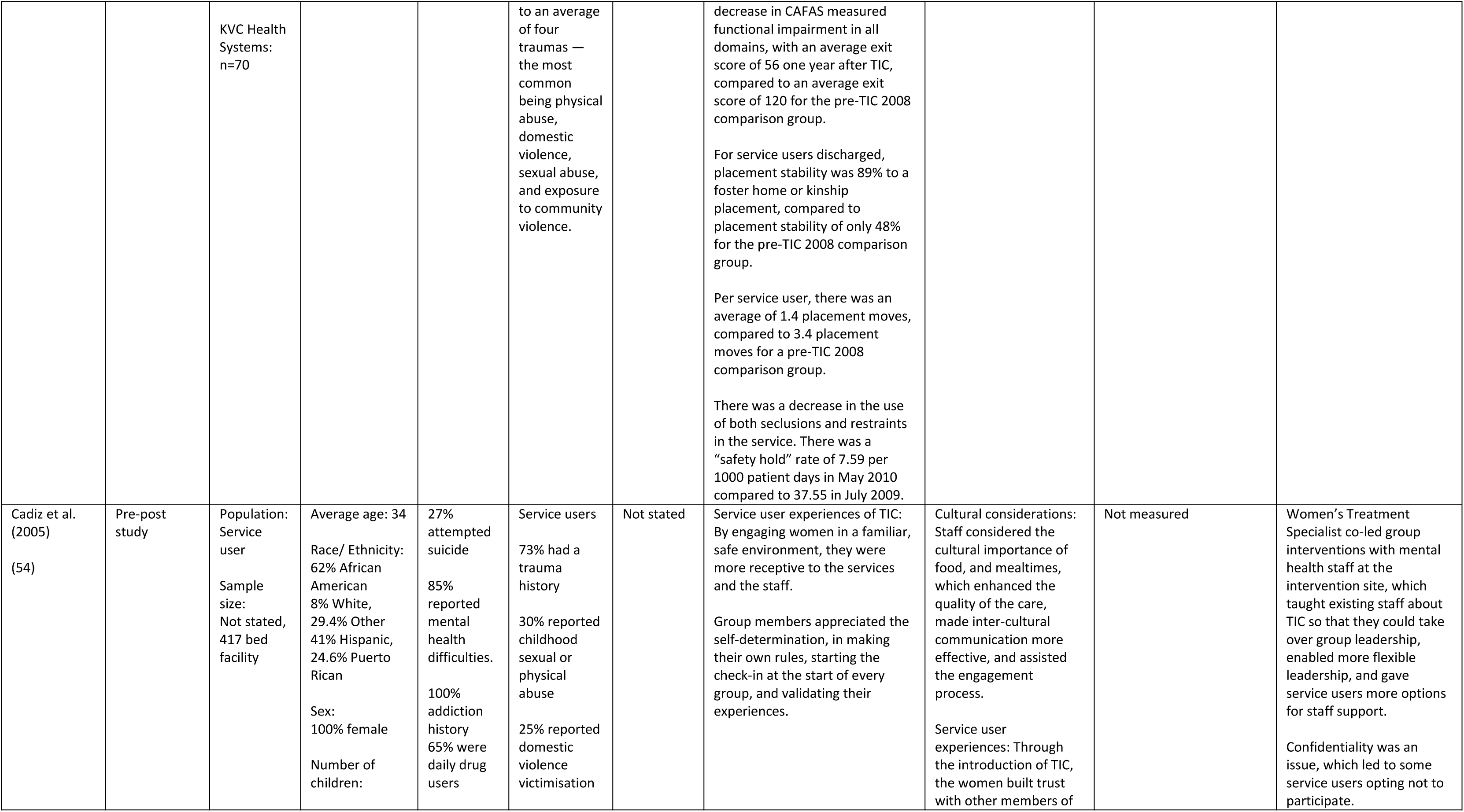
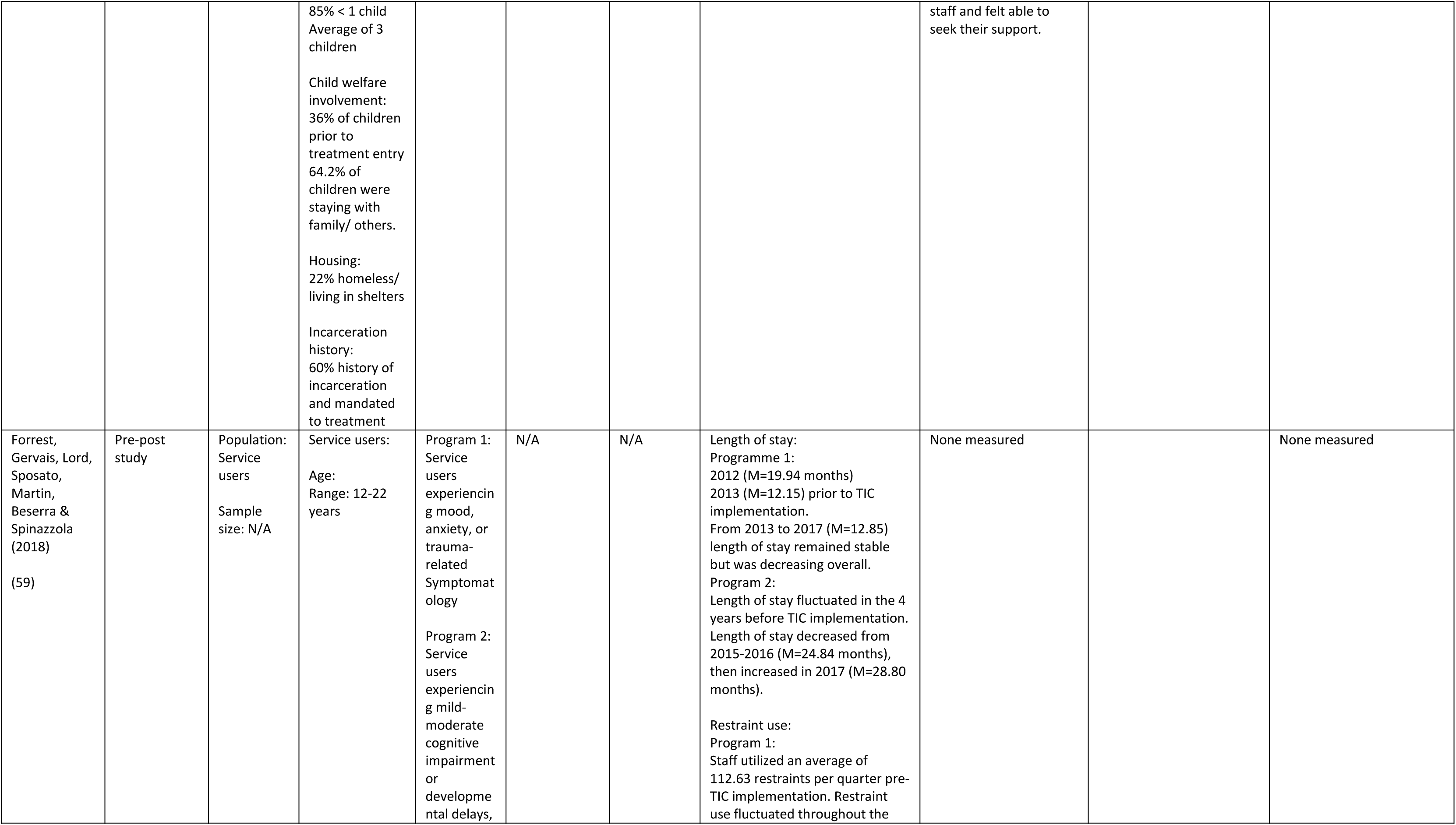
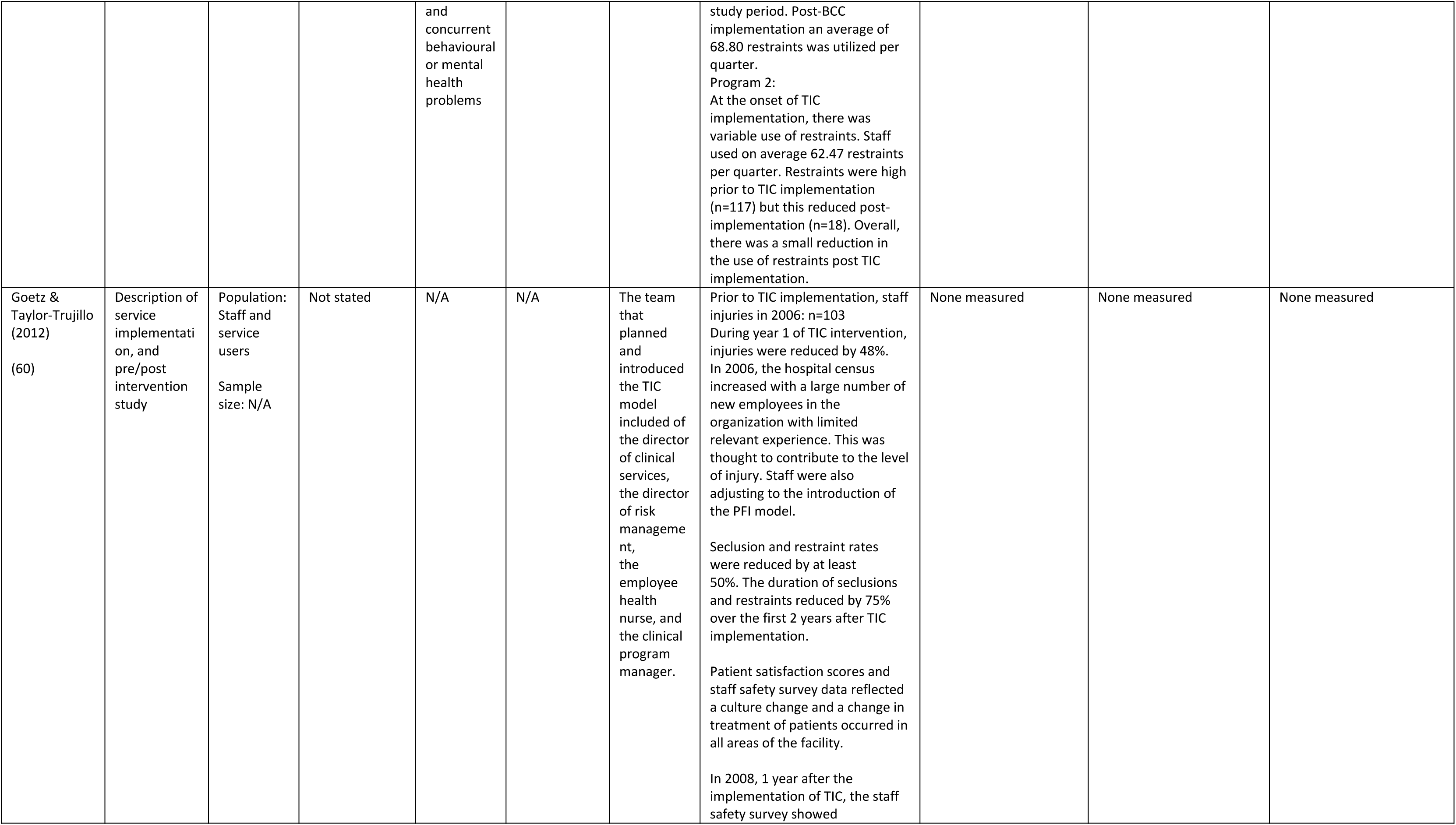
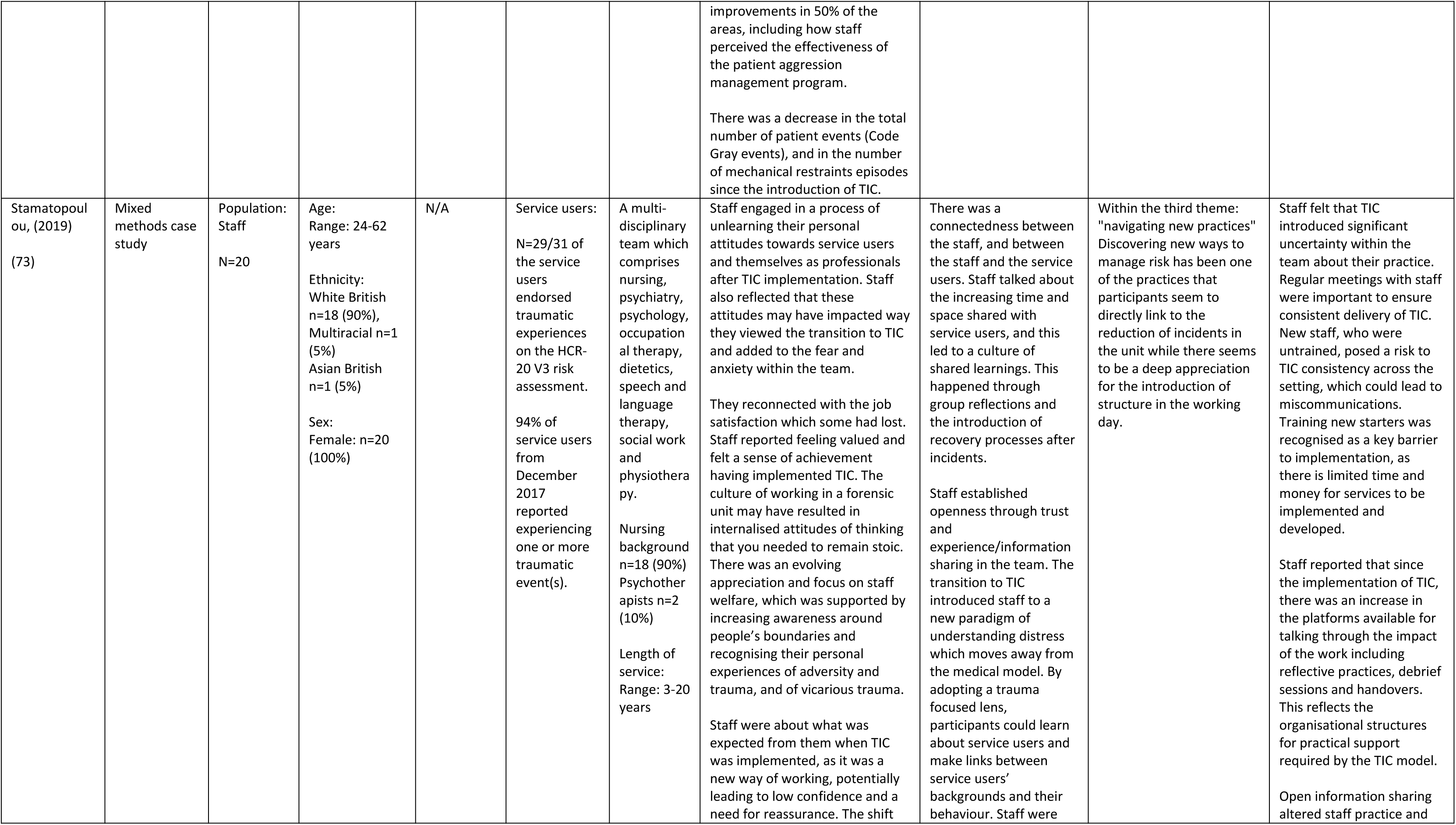
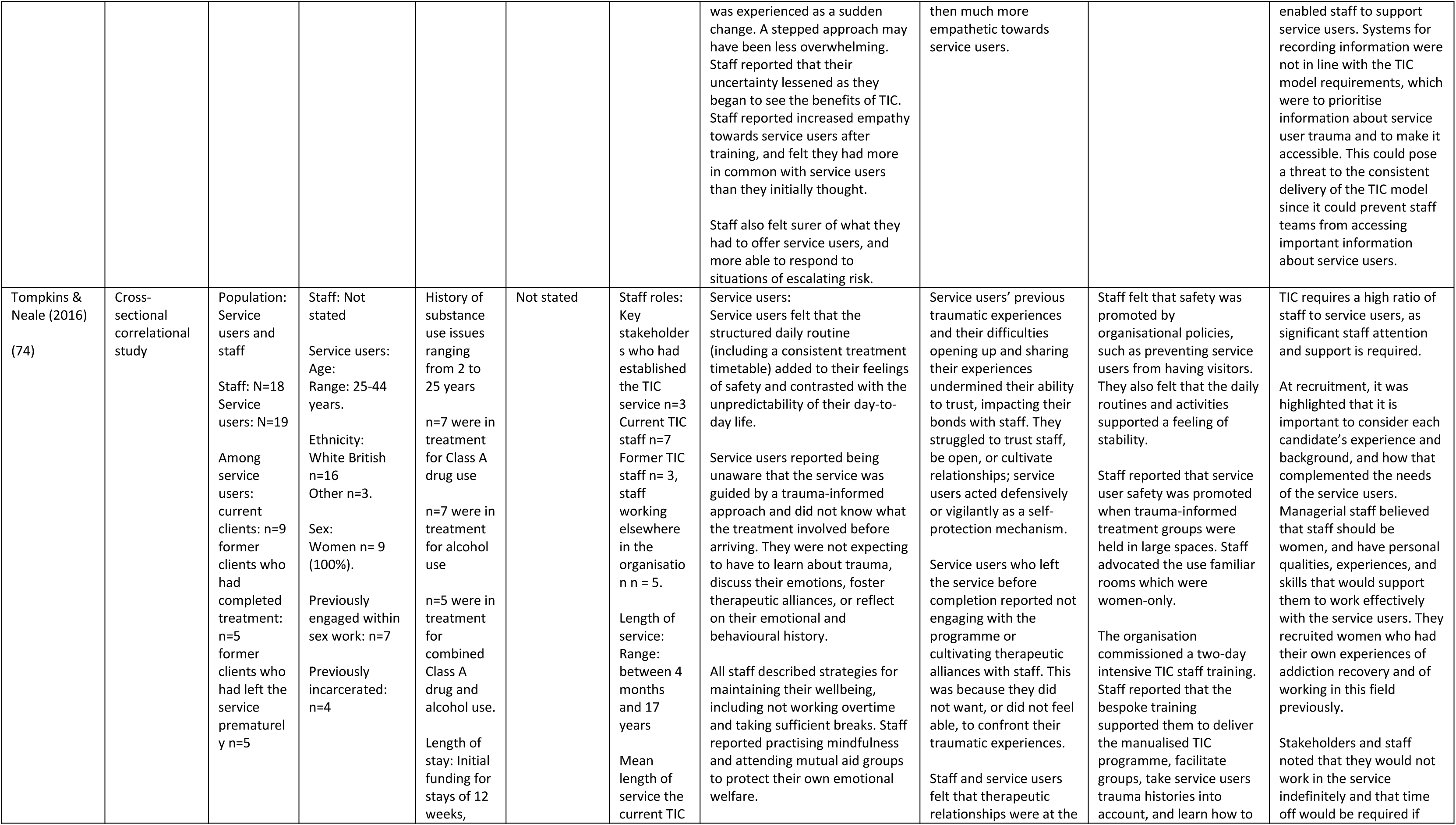
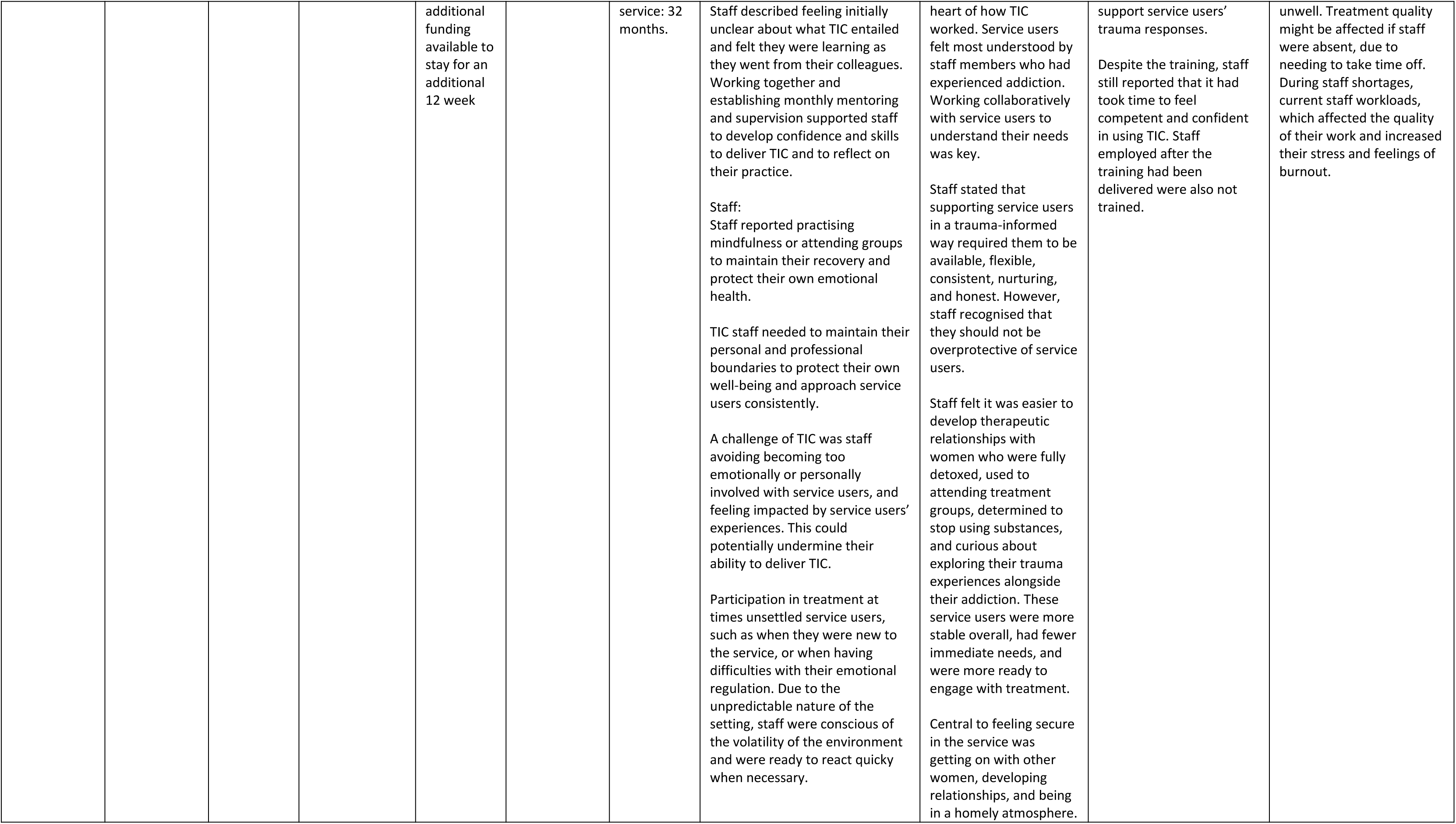
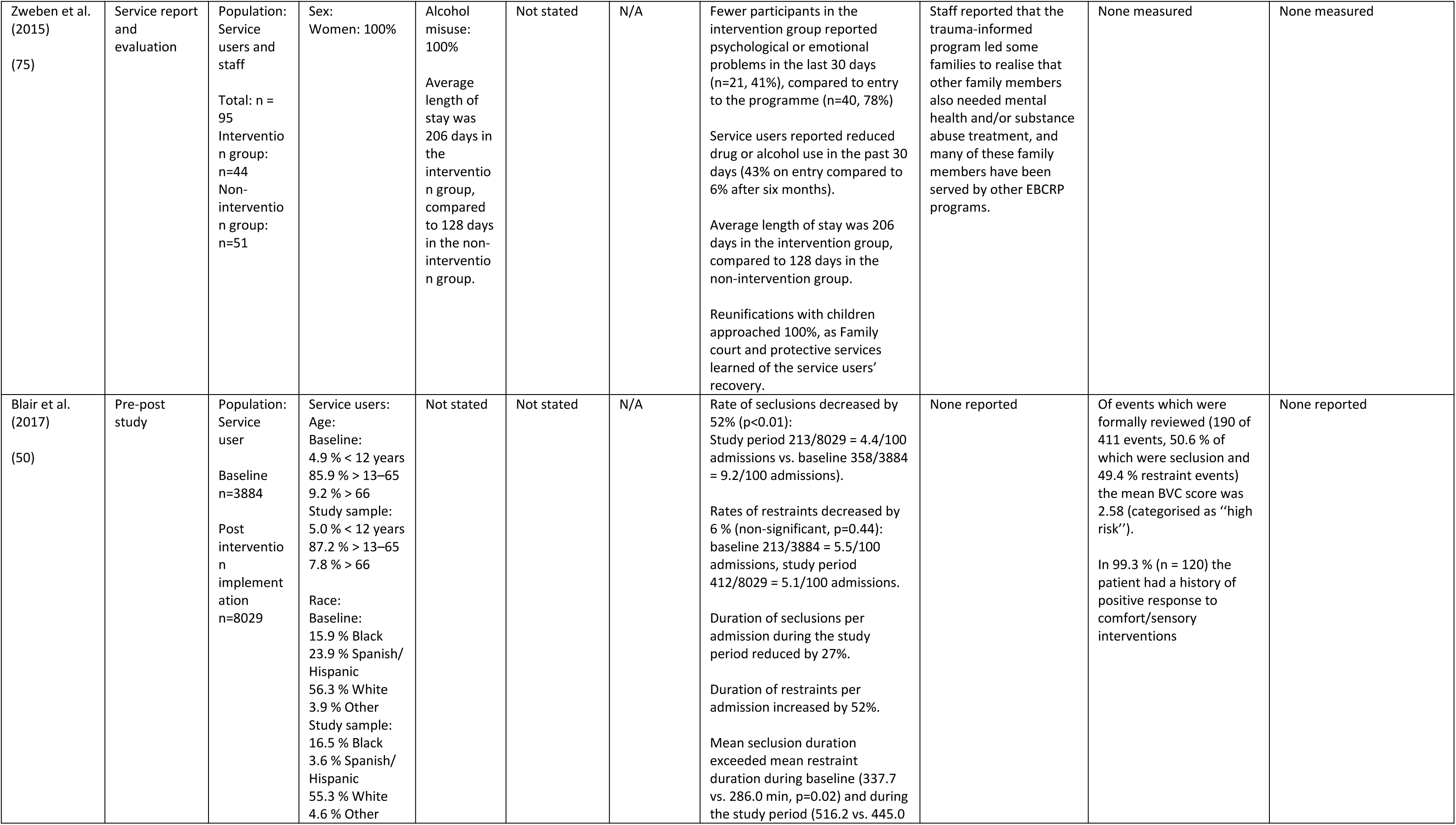
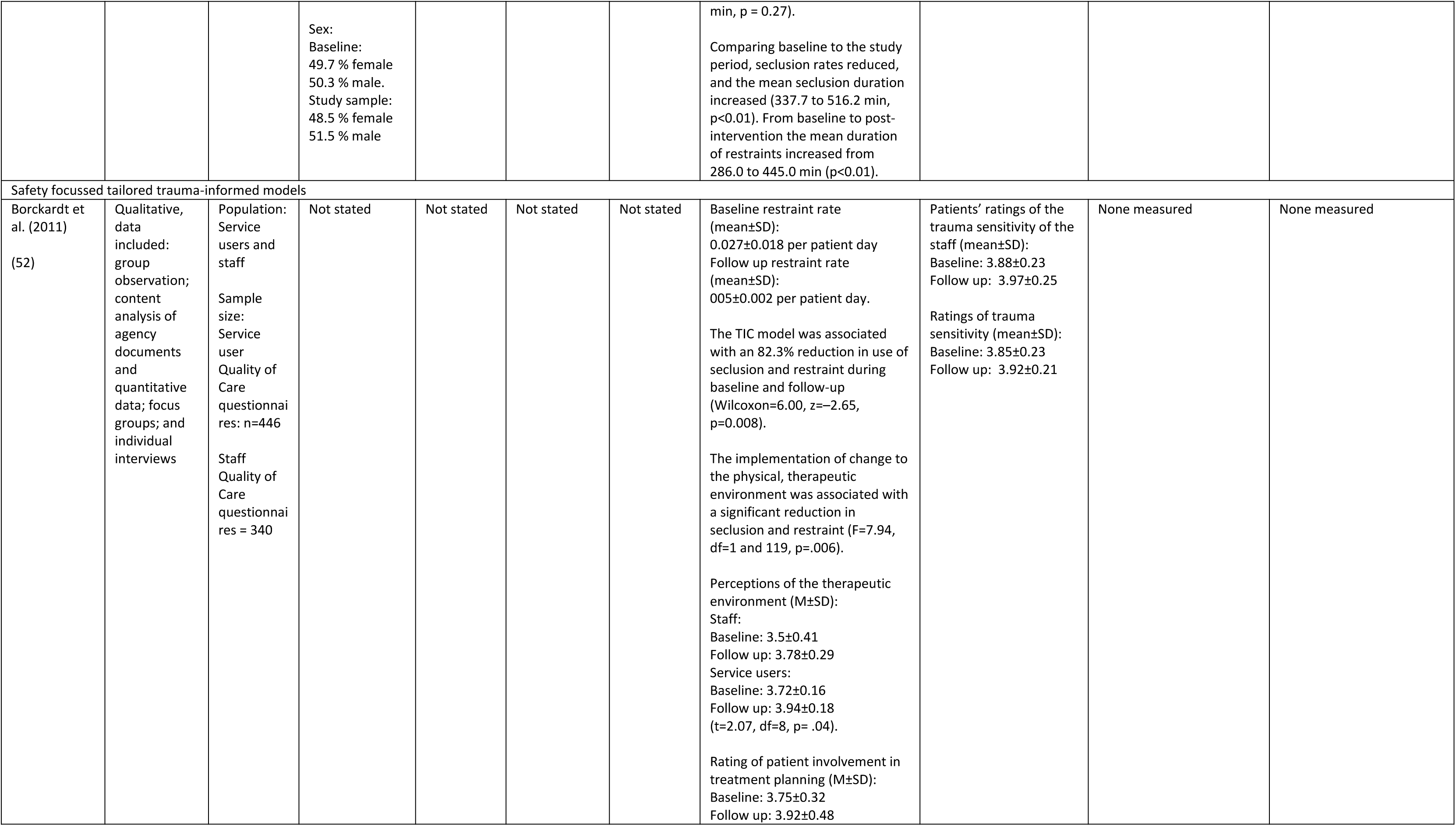
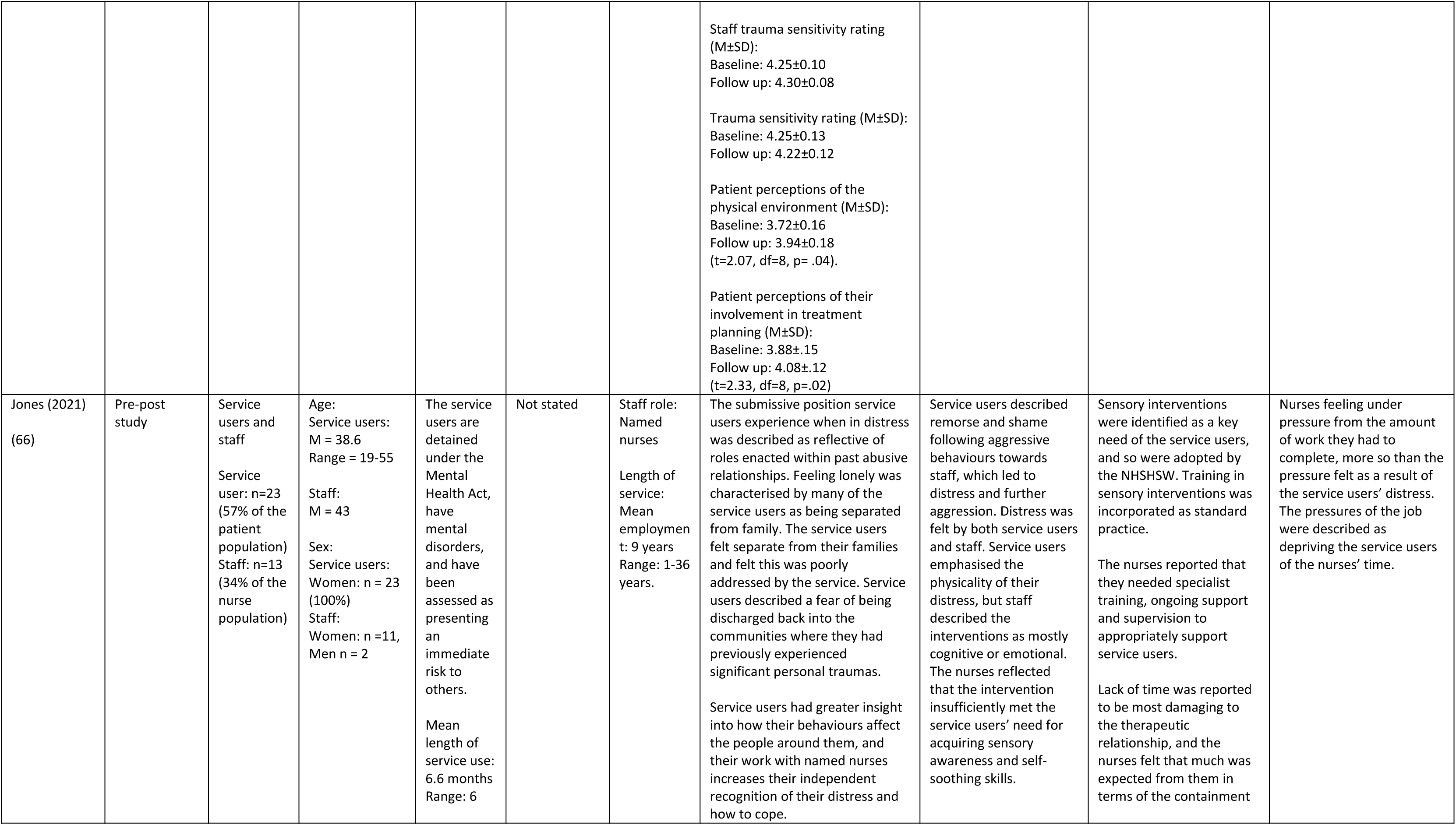
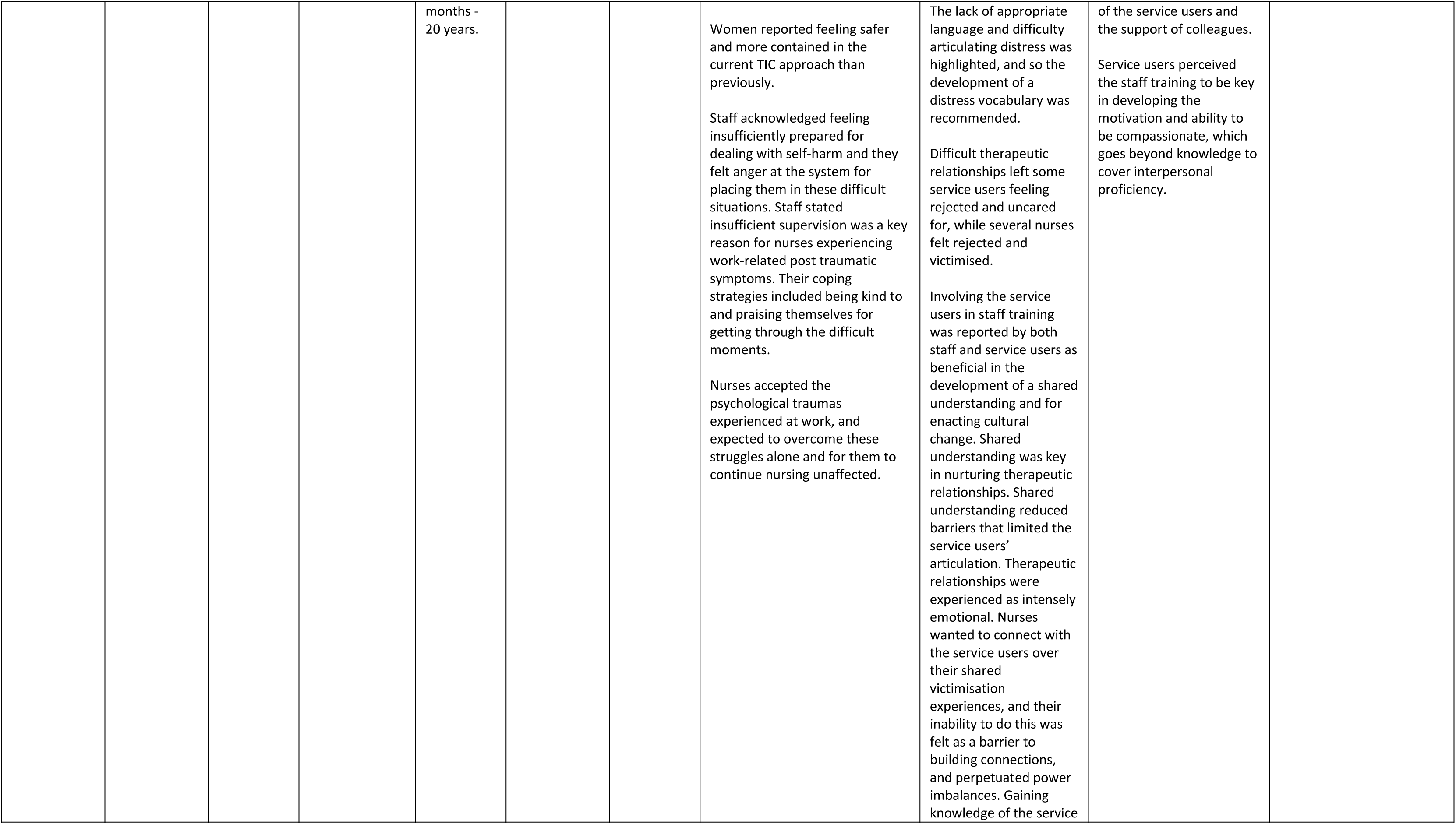
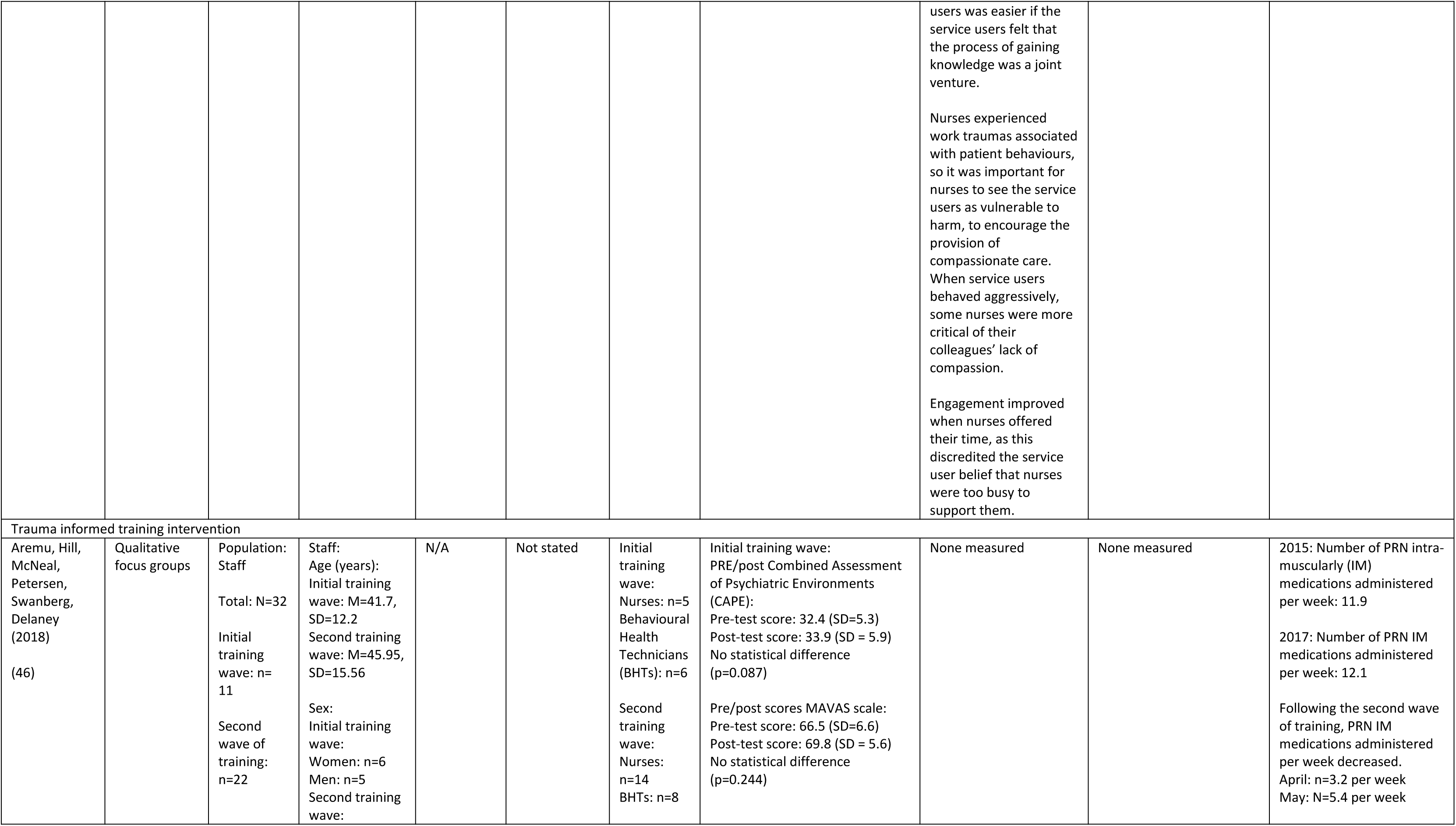
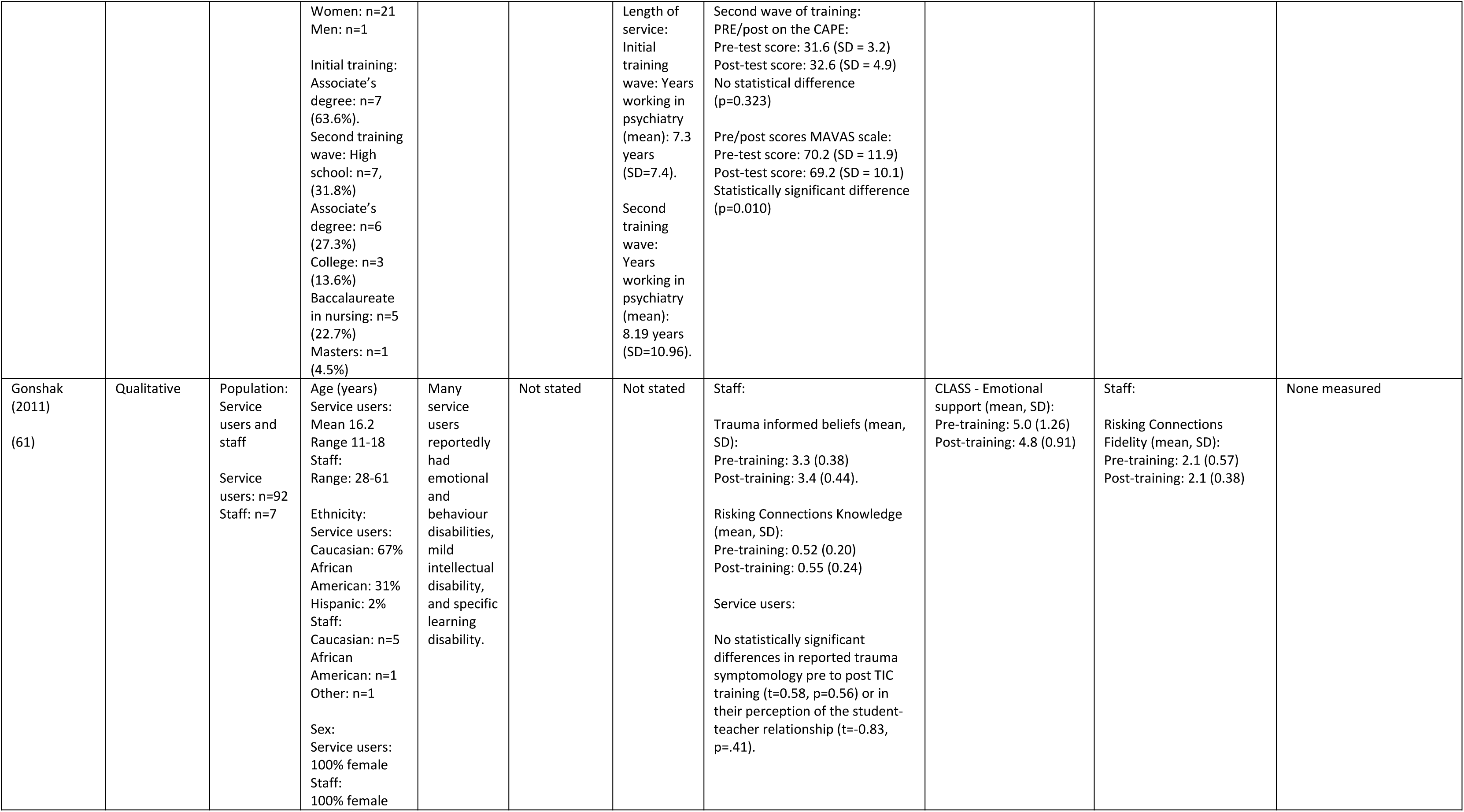
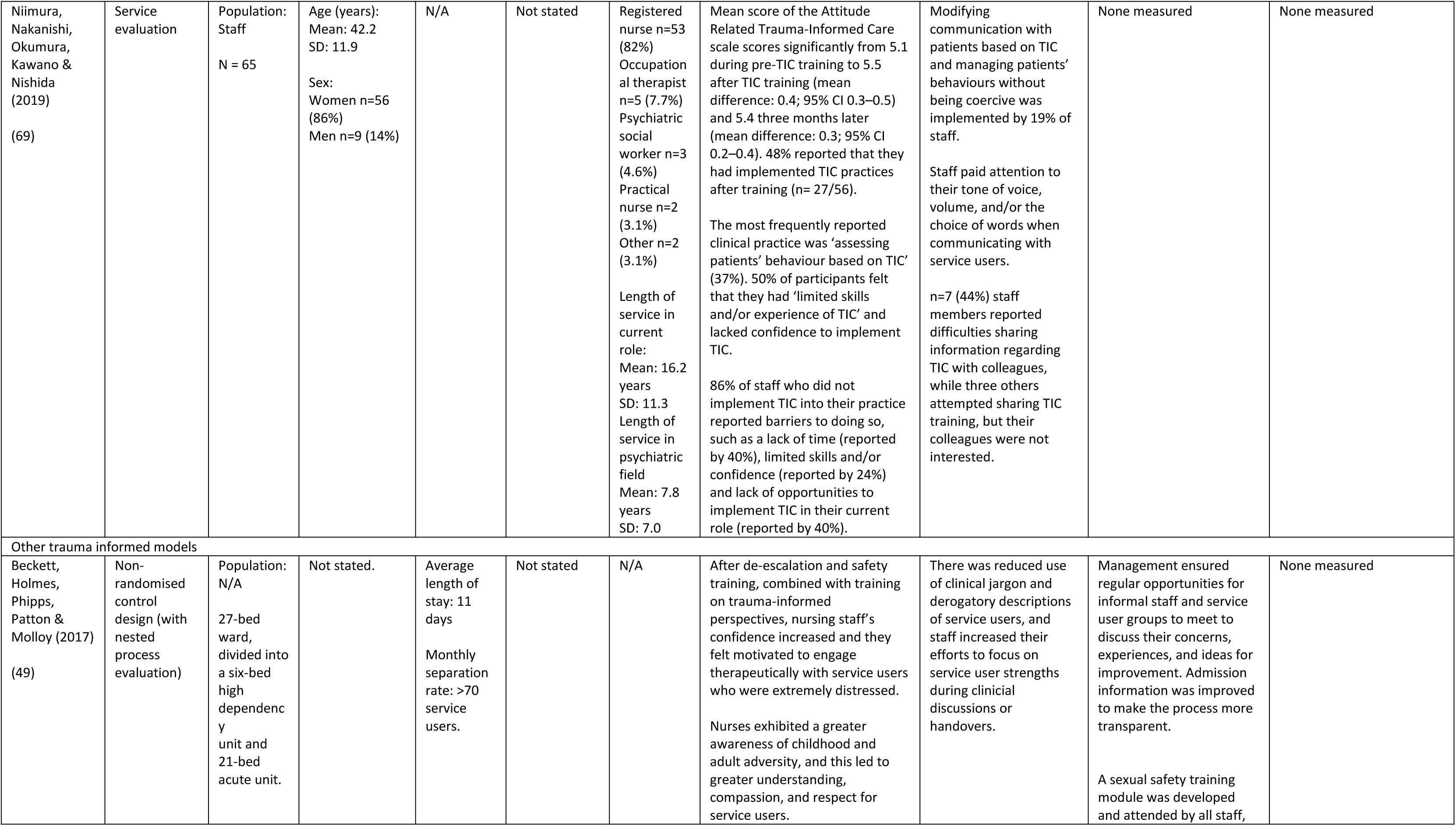
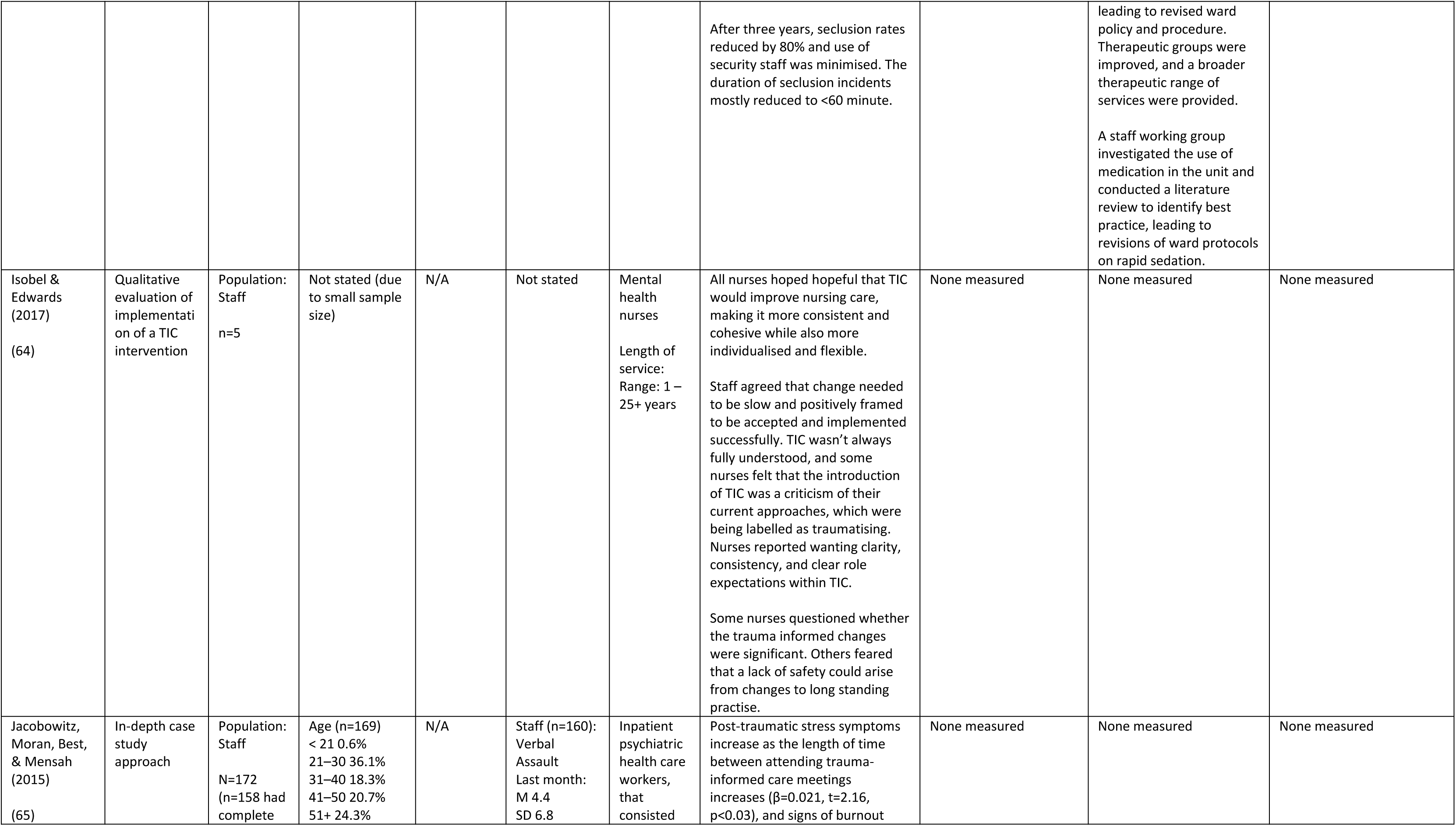
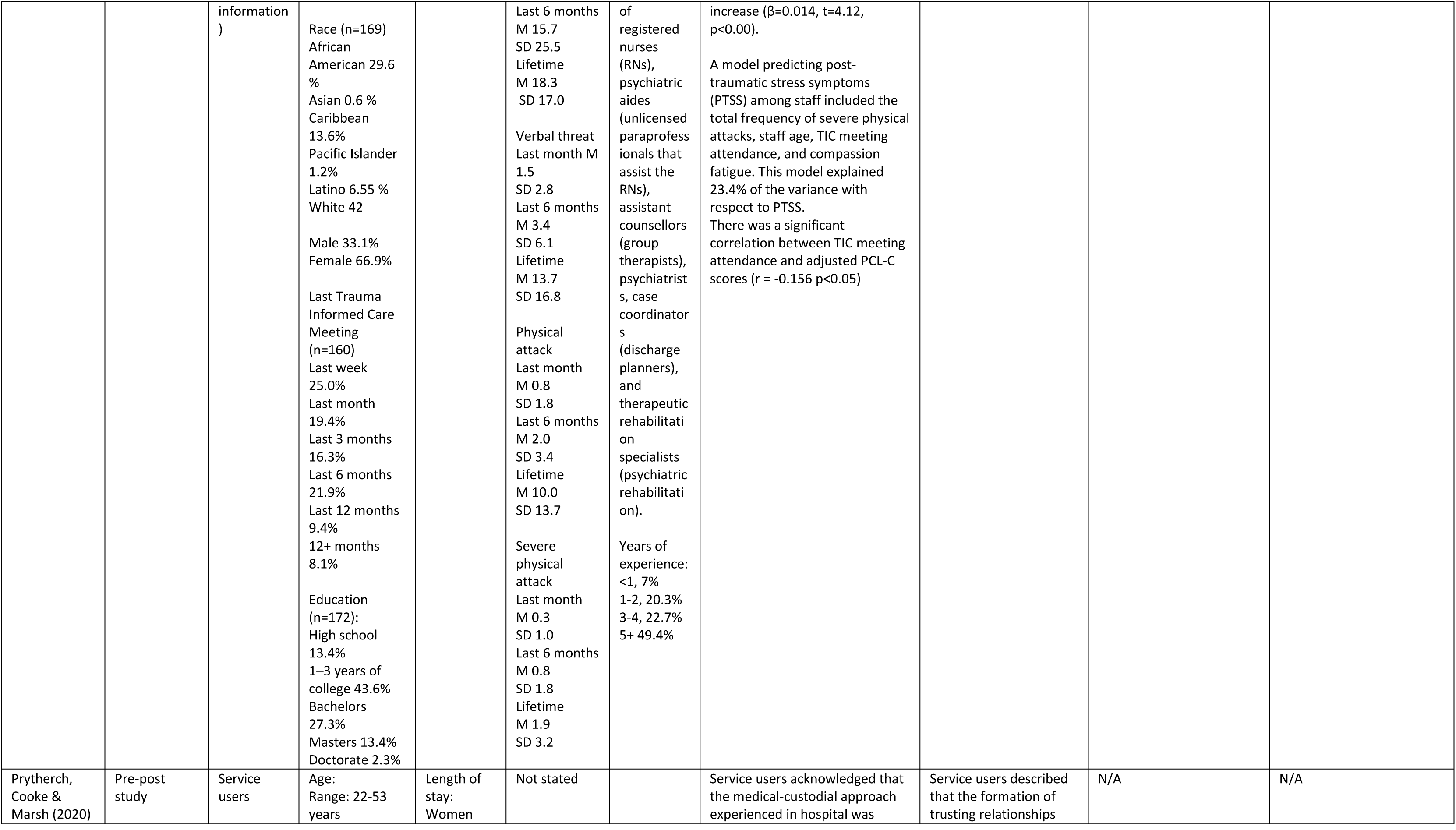
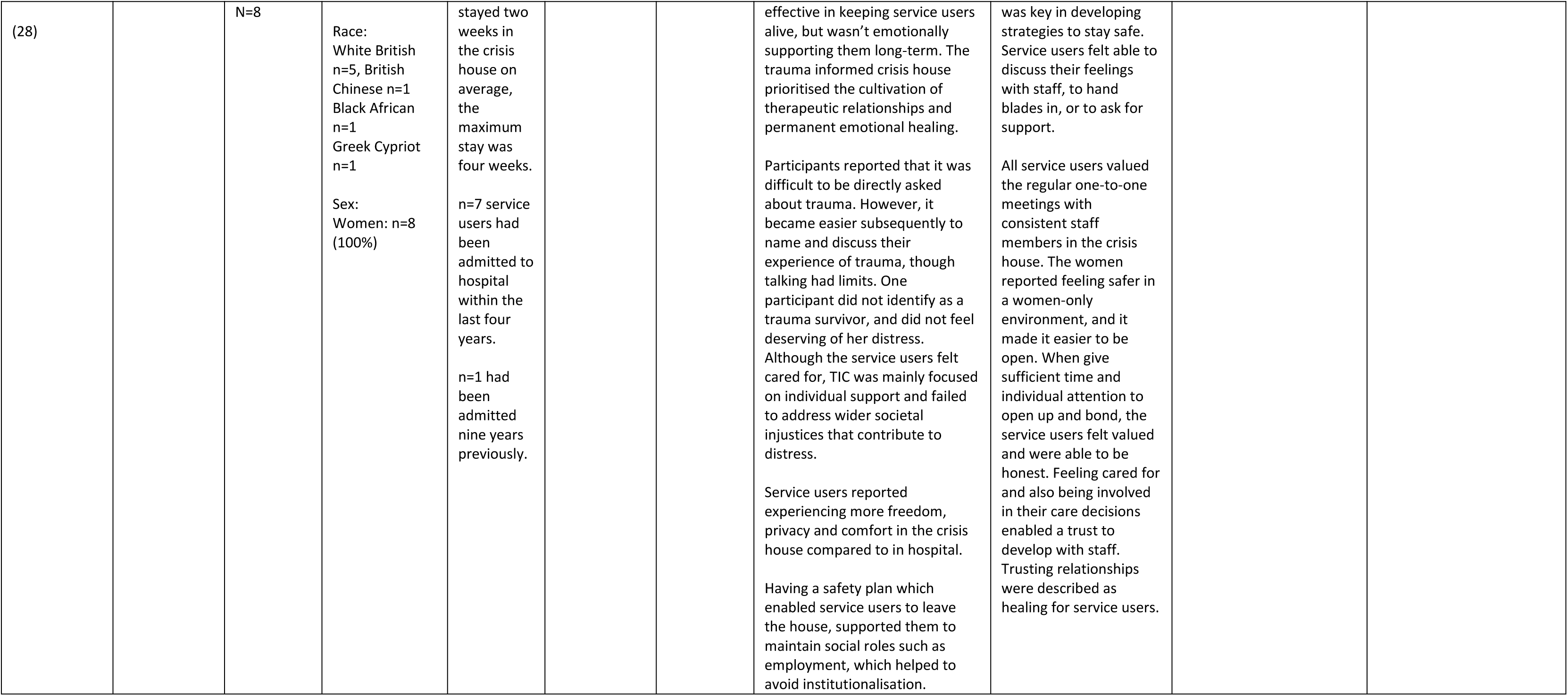
Results table

## Discussion

### Key findings

TIC approaches implemented in acute, crisis, emergency and residential mental health settings were broad and varied. Studies that utilised either the Six Core Strategies model (83) or the Sanctuary Model (84) followed a clear structure to enact organisation and clinical change. Other studies implemented TIC models tailored to their specific settings, some with a particular emphasis on improving safety. The value of staff training was highlighted across studies, but the content of training was often described in limited detail, making it difficult to draw inferences around its comprehensiveness. Two principles from Sweeney and Taggart (2018) that were often underrepresented in the TIC models were i) recognising social traumas and the intersectionality of multiple traumas and ii) working in partnership with trauma survivors to design, deliver and evaluate services. This indicates missed opportunities to use lived experience input to develop services, particularly for service user groups who have experienced multiple and social traumas. Trauma itself was poorly defined, raising the question of whether TIC implementation can be truly meaningful without a clear framework of what trauma itself entails.

Service users felt connected to their traumatic experiences and the implementation of TIC practice improved their ability to communicate their traumas. However, this could be a difficult process, and it is important to highlight that service users should not be required to share their experiences of trauma. The focus of TIC approaches in relation to empowering service users should also extend to giving them autonomy over when and how they disclose their traumatic experiences to staff (91). While service users felt cared for, trusting, and trusted by staff working in TIC services, some elements of distressing service user experiences were not reflected in the TIC materials e.g., lack of access to housing or benefits, indicating a need for closer adaptation of TIC to the needs of service user populations.

Staff initially felt overwhelmed, anxious, criticised, and even reluctant at the introduction of TIC, though TIC positively influenced staff empathy, compassion, and wellbeing. Time was a key factor for implementation, particularly time to allow for staff to build their skills and confidence. Once skills were developed, staff felt proud and satisfied with their roles. There were potential implications of delivering TIC for staff with histories of trauma also, e.g., providing staff with support and supervision to process those experiences. Future research could explore how these changes impact on staff turnover, a key concern in providing consistent care in inpatient and residential care settings.

Reductions in restraint and seclusion were observed across settings, although the quality of evidence is limited as most studies are pre/post designs and lacked a comparison group. However, a broader question remains around whether services that continue to use restraint and seclusion (even in a reduced capacity) can be considered trauma informed.

There was a lack of economic evidence available. However, if TIC is reducing rates of seclusion and restraint, reducing length of stays, as well as creating a more therapeutic environment (as reported in this review), this may have a positive impact, as conflict is costly (92), while increased patient satisfaction is associated with reduced costs (93). However, there was also a lack of data on carers, perhaps due to our focus on specific settings, and very little evidence on hospital emergency departments (where care may be experienced as traumatising (94)), and on community-based crisis assessment services, home treatment, or acute day units, which future research could investigate.

### Strengths and limitations

Our broad literature search retrieved evidence on TIC approaches in a variety of mental health settings, providing insight into implementation and outcomes by setting. By nature of reviewing existing academic and grey literature, we are behind the curve of survivor thought and experience of TIC implementation. For example, information on the potential harms of poorly implemented TIC has been documented by people with lived experience (95), such as feeling required or coerced to confront their traumatic experiences. There was heterogeneity in the reporting of outcome measures across studies, limiting the generalisability of our conclusions. There were significant methodological weaknesses across the included studies; they were primarily cross sectional, with few employing a control group or a randomised design, limiting our ability to draw causal conclusions about the impacts of TIC. There was more data on residential mental health care settings compared to other settings, mirroring other review findings (96); this may be due to their longer-term nature, which may be more amenable to the implementation and evaluation of TIC. This limits our understanding of TIC implementation and outcomes in other settings.

### Implications for policy

A central tenet of TIC is empathy and understanding of the role of trauma in shaping mental health outcomes, which are positioned as being at the core of how mental health services can best support service users. Arguably, these principles should be a basic requirement of all mental health services, whether labelled as trauma-informed or not. Introducing TIC into services may be a method by which basic practices can be integrated and maintained in a structured way (6). This requires system-level change, which may be both time and financially resource intensive, although we identified little economic evidence.

### Implications for practice

Delivering TIC in inpatient and residential settings requires clear and decisive leadership as well as clear staff roles(97). In addition, TIC needs to be implemented, reviewed and evaluated collaboratively with service users to ensure it is delivering safe and appropriate care, otherwise services may continue to cause significant harm (95). TIC could be included in pre-registration education across healthcare professions (98, 99) and used to inform and shape existing clinical practices. New starters may benefit from training as part of their inductions, and consistent staff supervision also supports the effective delivery of TIC (100). These requirements result in a sustainability challenge as the creation and delivery of training is resource intensive, and staff turnover rates are high (101). Finally, clinicians should be conscious not to cause further harm through the delivery of TIC (95), and that confronting and dealing with trauma should always be the choice of the service user, and receipt of TIC should not be dependent on service user willingness to engage with their traumatic experiences.

### Implications for future research

We have identified significant evidence gaps around TIC implementation in non-inpatient acute care, including A&E, crisis teams, crisis houses, and acute day hospitals. Most of the evidence is concentrated within the USA, and there is a paucity of data on TIC elsewhere. Very few studies included control or comparison groups, crucially limiting our ability to determine the nature and strength of change due to TIC. With the development of other approaches to improving inpatient care, for example, Safewards (102), robust research methodologies must clarify the specific advantages of TIC. Future primary research could explore TIC in acute and crisis mental health care settings, where it may be more difficult to implement a consistent and sustainable TIC approach as service users are engaged for shorter time periods. Future research could also consider the potential negative and harmful impacts of TIC. The present study could be extended to map the use of TIC in forensic settings, where poor mental health and experiences of trauma are also highly prevalent.

## Conclusion

This scoping review has demonstrated the range of TIC approaches used in acute, crisis, and residential settings as well as a range of outcomes relating to service users and staff experiences, attitudes, and practises. TIC implementation requires commitment and strong leadership to enact organisational change, as well as appropriate training and supervision for staff and the involvement of service users in the design and delivery of approaches. Future research requires robust methods to accurately measure the impacts of TIC approaches and their potential benefits over existing care practices, through the utilisation of comparator conditions to TIC models. Research would also benefit from exploring how TIC impacts on carers; how trauma is understood in emergency services (settings that are frequently used by trauma survivors); and prioritising the expertise and views of those with lived experience with respect to how best to deliver TIC across mental health services.

## Data Availability

The data used in this study is published and available from each of the included studies

## Lived experience commentary

### Written by LM and RRO

As survivors of trauma from both within and outside mental health services, we welcome approaches which recognise its importance, and are pleased to see the range of perspectives which have been studied. We hope future research can engage with more work from outside the global north, which dominates in this paper.

However, from reading this literature, we do not know whether as patients we could always distinguish a trauma-informed service from any other - particularly given implementation challenges described in services with high levels of inconsistency, agency staff, new starters, and/or staff scepticism. At times trauma-informed care seems to be a means to an end which should already be universal, i.e., treating service users with respect and supporting our autonomy.

It does not particularly matter to us whether a service is trauma-informed on paper if the support staff change day by day and cannot or will not put the theory into practice. Nor does it matter that staff believe themselves to be validating our trauma, if they also retain power: if we can be physically held down; if the versions of ourselves written in their notes overwrite our autobiographies; if we have to surrender control of our most painful experiences in exchange for care. We have experienced some of these; we have heard about others happening to our peers, sometimes in the name of TIC (e.g. https://www.psychiatryisdrivingmemad.co.uk/post/trauma-informed-care-left-me-more-traumatised-than-ever).

We welcome several studies showing reductions in restraint in trauma-informed environments (although note that this was not a universal finding). But we question whether a system founded on such violence can be anything other than traumatising in its own right. A version of TIC implemented in an under-resourced, fundamentally carceral system carries all the risks of harm associated with that system - with the added gaslighting of framing it as trauma-informed harm. As such, everyone involved in providing care they view as trauma-informed should consider what power over our stories and our bodies they are willing and able to give back.

## Acknowledgements

We would like to thank the NIHR LEWG for their contributions to this project, including researcher T.K. We would like to acknowledge the academic and lived experience experts who shared resources with us, and recommended papers for inclusion.

## Author Contribution

Formulating the research question(s): SJ, KT, AS, KS, PB, PM, NVSJ, RS, RRO, TJ, PN, TK, LM

Designing the study: SJ, KT, AS, KS, PB, PM, NVSJ, RS, LM, RRO, TJ, PN, TK

Conducting the study (including analysis): KS, KT, EM, JS, UF, FA, PB, SC, VT, MS, NVSJ, JG

Writing and revising the paper: KS, KT, SJ, AS, EM, JS, RS, UF, PB, SC, VT, MS, NVSJ, RA, JG, RRO

## Data Availability

All data used is publicly available in the published papers included in this study.

