## Appendix 2 for "A scoping review of trauma informed approaches in acute, crisis, emergency and residential mental health care"

**EMBASE SEARCH TERMS**

**Search One  - Date searched 24.02.22**

trauma-sensitive OR (trauma adj1 sensitive) OR trauma-informed OR (trauma adj1 informed) OR

trauma-integrated OR (trauma adj1 integrated) OR (trauma adj2 aware) OR trauma focus?ed OR violence informed

AND

psychiatric inpatient* OR acute psychiatric ward* OR psychiatric ward* OR psychiatr* serv* OR psych* consult* OR (liaison adj2 psych*) OR consultation liaison* OR mental health liaison OR (hospital adj1 psych*) OR crisis resolution OR crisis service* OR crisis assessment OR crisis house* OR crisis bed* OR exp psychiatric department/ OR exp psychiatric intensive care unit/ OR mental hospital/ OR psychiatric emergency service/ OR crisis intervention/

**Search two - Date searched 24.02.22**

trauma-sensitive OR trauma adj1 sensitive OR trauma-informed OR trauma adj1 informed OR

trauma-integrated OR trauma adj1 integrated OR trauma adj2 aware OR trauma focus?ed OR violence informed

AND

residential OR therapeutic communit*.tw. OR rehabilitat* AND substance OR drug OR alcohol* OR (illicit*adj3 addict*) OR abuse OR depend* OR disorder OR use* OR using OR drink* OR misuse OR treat* OR over* OR withdraw* OR exp substance abuse/ exp alcoholism/ OR exp opiate addiction/ OR withdrawal syndrome/ OR drug induced psychosis/

**Search three - Date searched 24.02.22**

trauma-sensitive OR trauma adj1 sensitive OR trauma-informed OR trauma adj1 informed OR

trauma-integrated OR trauma adj1 integrated OR trauma adj2 aware OR trauma focus?ed  OR violence informed

AND

inpatient OR hospitali?ed OR forensic.tw.

AND

prison* OR incarcerat* OR jail* OR inmate OR correctional facility OR correctional institution OR correctional offen* OR detain* OR detention OR penitentiar* OR penal institution OR juvenile detention OR offend* OR medium secur* OR high secur* OR low secur* OR forensic psychiatr* OR forensic service* OR forensic hospital OR forensic mental health OR forensic psycholog* OR therapeutic communit*
