## Appendix 3 for "A scoping review of trauma informed approaches in acute, crisis, emergency and residential mental health care"

| Author(s) and year | Setting and country | Name of model and aim/ purpose of intervention or service | Trauma-informed model content | Who was the model designed by and who implemented it? | Theoretical basis of intervention and service | Definition of trauma within the trauma-informed model under investigation |
| --- | --- | --- | --- | --- | --- | --- |
| The Sanctuary Model | | | | | | |
| Farragher et al. (2005) | Residential mixed-gender youth treatment unit for children with severe emotional problems, New York, USA | Name: The Sanctuary Model  Aim to reshape their programme to make the service a better community for all members, including service users and staff. Also, to recommit to their treatment centre mission of helping children recover. | ***Implementation:*** Creating a trauma-sensitive culture involved a deep examination and transformation of agency dynamics and included all community members, upper management, families, middle management, children, line staff as well as every department and function. The change had five components: integration, understanding trauma, avoiding re-enactment, fighting rigidity and embracing non-violence. Most importantly, creating this kind of culture was an on-going process, one that is never complete, never linear, but always evolving with lots of movement forward and a fair share of movement backward.  *I****ntegration:*** Integration refers to the concept of creating a unified and healthy organisation in which all members are active participants in decision-making, and all accept responsibility for each other’s well-being and safety. At Andrus, the Sanctuary Model provided the forum for moving toward integration. With Dr. Bloom’s help, the unit created a multi-level, multi-disciplinary Core Team that met twice monthly with her for a year to discuss agency issues and to learn about how trauma affects not only individuals, but entire organisations. Through the Core Team, representatives from each department were able to take an honest look at their own departments’ strengths, shortcomings, styles of interactions and most importantly the assumptions which had been driving their current behaviours and functioning. Beginning with these individual or small group assumptions, the Core Team was able to build shared assumptions, beliefs and values that were common to the entire agency. These shared assumptions, beliefs and values would serve to foster greater integration within the agency. The new shared beliefs were:   1. We will ask clients not “what is wrong with you”, but “what has happened to you?”   2. We must work as a non-hierarchical group.   3. All people who work or live or come to school here must be safe   4. Our job is to help people put their feelings into words rather than act them out.   5. We must help people think as a group.   6. We must approach conflict resolution as a team.   7. We must create a living learning environment.   8. At Andrus, the milieu IS the treatment.  This eighth tenet of the Core Team, to acknowledge that the milieu is the treatment, was most helpful in cementing the other seven. Agency leadership took the stance that the milieu is the most powerful force in creating change in clients, and every person, regardless of title or position, is part of that milieu. Working within this framework, all staff should feel empowered and responsible to make decisions effecting a child’s treatment. This clarification of the treatment philosophy not only allowed staff at every level of the hierarchy to participate in shaping the culture of the milieu, but it also demanded that they do so. But to effectively participate, people had to communicate with each other. To communicate with each other, people had to have a shared language. One of the tasks of the Core Team was to train the staff in trauma theory and to teach them a common language – the language of trauma. The crux of this language is the SELF Model (Foderaro and Ryan 2000; Foderaro 2001). This simple way of understanding the needs of people who have been traumatised also organises treatment goals. Part of employing this common language in the milieu is to teach the children to use it as well. Treatment plans, planning conferences, interviews and therapy sessions all construct dialogue using the language of SELF. Staff and children become explicit about which behaviours promote or hinder safety as well as when and how certain coping skills may allow children to manage their emotions in ways that will promote their own safety and well-being. The language also gives children and staff an opportunity to address loss and grief, a treatment issue that is often neglected in residential settings in favour of a focus on behaviour. Finally, the SELF model encourages a focus on the future, and that in turn encourages the children to work toward change. The use of the SELF model applies not only to the children and families, but to the staff and the agency itself as a living, dynamic entity.  ***Understanding trauma:*** In order to effectively use the language and begin the work on safety, emotional management, loss and future with the children, the staff needed to have a basic understanding of how traumatic experiences disrupt normal coping and functioning in individuals and systems. As part of their training effort, they designed a six-session training for direct care staff, including milieu therapists, social workers, teachers, as well as administrators, which covered basic psychobiology of trauma, re-enactment, the language of SELF, and vicarious trauma. They also used the trainings as an opportunity for staff to assess the agency’s functioning in various areas. They talked about values and beliefs and how our own experiences can impact their work. They also focused on understanding that working with traumatised children and families can be traumatising to the staff. Talking to the staff about vicarious trauma and ways to use each other for support was a large part of the training.  In each classroom, clinical staff and teachers began psycho-education groups to teach the children about trauma. Acknowledging the important role of parents, and their need to use the tools of SELF too, children were encouraged to teach their families what they had learned. The forums of open-school night, family day, and graduation were used to allow the children opportunities to present what they had learned. Social workers incorporated the language into family therapy sessions, and taught families about trauma in that setting as well. The assessment process was also updated and were designed to include several standardised trauma assessments, the Traumatic Events Screening Inventory and the Trauma Symptom Checklist for Children to measure exposure to trauma and trauma symptoms.  ***Avoiding re-enactment:*** Staff training included teaching about the traumatised person’s tendency to re-enact the trauma and to re-enact traumatic relationships. Part of the work done in the trainings was to help staff identify and break through their assumptions about each other and begin to build trusting, supportive relationships. An expectation of solid teamwork was created, and it was mandated that cottage teams create times to meet with each other to plan and to address emerging problems. Part of creating a trauma-sensitive culture is creating safe space for confrontation. During trainings, discussions were had about race and culture, and how some personal styles of interacting with children might be seen as inappropriate or re-traumatising. They talked about creating a trauma-sensitive culture in residential treatment, about how certain staff were seen as heavy handed, and how co-workers feared confronting these methods out of fear of retribution. Staff were encouraged to use the “tapping out” technique to let a colleague know that he or she needs some time away from the child. Supervision was used to reinforce the use of tapping out and constructive crisis resolution as being more valued than saving face with a child.  ***Fighting rigidity:*** In a trauma-driven organisation, responses are rote and reactive, and planning is minimal. In a trauma-sensitive culture, responses to problems are explored and evolve through a more democratic system of shared decision-making. A democratic environment creates space for creativity. Staff need to know that it is safe to try new things, if they stay within the shared values and beliefs of the organisation. In a trauma-sensitive residential culture, all members of the team are providing treatment by influencing and creating the therapeutic milieu. Clinicians at Andrus were seen as the treatment coordinators for each of their cases. Some of the other ways that they fought rigidity in how they conceptualise therapy was through outside training and a focus on trauma-focused interventions, including new group and individual therapy models.  ***Embracing non-violence:*** In a trauma-sensitive culture, the staff and children use their understanding of trauma and its impact to make conscious choices to avoid violence. One way to accomplish this is through safety planning. Having a formal meeting in which staff along with a child can document triggers to aggression, staff responses that might exacerbate aggression and staff responses that the child finds most helpful can lead to more appropriate interventions and a decrease in violent and aggressive behaviour. The team, including the child, not only discusses what the staff should do to help a child avoid aggression, but the child also must have a plan of action, outlining steps he or she will take to avoid violence when triggered. Embracing non-violence also means embracing shared power and decision making and reducing the abuse of power. One way to avoid abusive power is through open dialogue with members of the community at every level. Rather than becoming punitive with children or staff when they make poor choices, staff began to use these incidents as learning and teaching tools. This was facilitated by creating a Red Flag Review, a team meeting to understand and reformulate a plan, any time there was an incident of violence on campus. They also created mandatory reviews every time a restraint was performed. The staff involved met with their supervisor to discuss alternative or proactive responses for the future, and the team and the child met to debrief the incident and revise the safety plan. | Designed by: the Sanctuary model was originally designed by Dr Sandra Bloom (and team) in 1997.  Implemented by: the service was implemented with Dr Bloom's consultation. | Not stated in this paper. | Not stated. |
| Korchmaros et al. (2021) | A mixed-gender adolescent residential substance misuse treatment agency, South-western USA | Name: The Sanctuary Model and Seeking Safety  Aim to meet the special needs of youth with histories of maltreatment and exposure to violence. | The Sanctuary Model: The Sanctuary Model is a milieu-wide, comprehensive program model that is intended to impact all aspects of a residential care agency (James, Thompson, & Ringle, 2017). The Sanctuary Model strives to reorganise systems and change organisational culture to shift away from an atmosphere of recurrent or constant crisis.  Seeking Safety: Seeking Safety is a client-specific intervention intended to address trauma as a client-level problem (James et al., 2017). Seeking Safety prioritises five core principles: personal safety as a priority; integrated treatment for trauma and substance use-related disorders; focus on client’s needs; attention to the therapeutic process; and focus on cognitions, behaviours, interpersonal interactions, and case management.  ***Implementation of the models:***  The Sanctuary Model: During the first 4 months, six treatment agency staff attended The Sanctuary Model five-day national training. Upon their return, these staff became a Steering Committee with the goal of embedding The Sanctuary Model within the treatment agency. In month six, the Steering Committee began training a Core Team and provided six four-hour trainings over a 6-month time fame. The Core Team was comprised of 19 staff in diverse agency roles. The Core Team’s goal was to become immersed in The Sanctuary Model principles and to share their training and understanding with co-workers through enacting The Sanctuary Model in their daily work. For the first year of implementation, the Steering Committee and Core Team met twice a month and then met on a monthly basis for another year to discuss successes and challenges related to implementing The Sanctuary Model tools (e.g., red flag reviews; safety plans), embedding overall The Sanctuary Model tenets, and ongoing training needs.  Seeking Safety: the agency purchased Seeking Safety materials and the clinical director provided a one-day Seeking Safety training to six clinical staff (i.e., therapists; behavioural health staff; clinical supervisor). Other staff’s knowledge of the training and subsequent implementation of Seeking Safety was limited as Seeking Safety was considered a client-specific service. Therefore, Seeking Safety training was directed only to those who would be implementing or supervising the client-specific groups. The agency provided additional support to clinical staff facilitating Seeking Safety to supplement their training once they began implementing Seeking Safety. Core Team members observed group Seeking Safety sessions guided by the Seeking Safety Adherence Scale-Brief Version (Najavits, Liese, & Health, 2007) and provided verbal and written feedback to each facilitator multiple times.  The agency began agency-wide implementation of The Sanctuary Model and the group sessions of Seeking Safety during month four. | **The Sanctuary Model:**  Designed by: the Sanctuary model was originally designed by Dr Sandra Bloom (and team) in 1997.  Implemented by: six treatment agency staff attended the training and became a Steering Committee. They trained a Core Team comprising 19 staff in diverse agency roles. The Core Team’s goal was to share their training and understanding of The Sancturary Model in their daily work. In this way, all staff were involved in implementation.  **Seeking safety:**  Designed by: the Seeking Safety model was originally designed by Lisa M. Najavits in 2002.  Implemented by: six clinical staff (i.e., therapists, behavioural health staff, clinical supervisor) were trained in Seeking Safety. These people were responsible for implementing or supervising the client-specific groups. Core Team members observed group Seeking Safety sessions multiple times. | The Sanctuary Model: System changes are based on the active creation and maintenance of a nonviolent community of shared governance; in turn, this approach builds a socially responsive, emotionally intelligent community and safe environment that facilitates and encourages growth and change (Esaki et al., 2013; Leigh-Smith, Toth, Lehmann, & Sanders, 2014). The Sanctuary Model purports system changes as an essential shift for agencies that serve youth with trauma histories because over time the organisational structure and functions instigate traumatic re-enactment. This impacts staff ability to problem solve, effectively communicate, and provide meaningful services.  Seeking Safety:  Seeking Safety is a client-specific intervention which prioritises five core principles: personal safety as a priority; integrated treatment for trauma and substance use-related disorders; focus on client’s needs; attention to the therapeutic process; and focus on cognitions, behaviours, interpersonal interactions, and case management. | Not stated. |
| Kramer (2016) | Forensic male adolescent residential treatment unit, USA | Name: The Sanctuary Model  Aim to become a trauma-informed milieu committed to a theoretical and organisational framework that primarily would improve outcomes for the residents placed there by the juvenile courts. The Sanctuary Model aims to promote healing and sustain human growth, learning and health, and instil hope. | The Sanctuary model is a trauma-focused, therapeutic community-based theoretical and treatment model. It is composed of two primary components that lead to change: the creation and maintenance of a non-violent, democratic, therapeutic community; and psycho-educational exercises and modules to increase self-awareness and insight.  Within the Sanctuary Model, the SELF model is a psycho-educational construct that educates around four stages of recovery: safety; emotional intelligence; loss; and future plans. The curriculum includes 12 hours of training for staff each year and psycho-educational groups for residents three to four times a year at this facility. Residents receive a handbook explaining all of The Sanctuary Model’s commitments, the SELF model, tools/strategies, I.e., the “common language” of the Sanctuary Model community. Education in safety includes physical, social, emotional and moral safety. Emotional intelligence teaches how to manage emotions, connecting them to cognitions and behaviours. Moving through loss teaches how to grieve and deal with personal losses. Future planning calls for aspiring to new roles and a hopeful future.  Components of the Sanctuary model that effect change include creation of a safe community; the SELF model; psychoeducation; the Sanctuary commitments (non-violence, emotional intelligence, social learning and social responsibility, democracy/shared governance and open communication, and growth and change); Sanctuary tools (refer to Esaki et al.., 2013); the therapy modalities and the therapeutic milieu where relationships occur. Therapeutic relationships involve caring, supportive and healthy interactions, and staff taking an active interest in the lives of residents. The resultant relationships are shaped within the therapeutic community that is safe and enriching, encouraging and coaching residents to express feelings. | Devised by: the Sanctuary Model was originally developed by Dr Sandra Bloom (and team) in 1997  Implemented by: staff at the agency (not specified in any further detail) | The premise of the Sanctuary Model is that trauma deconstructs the social and personal world of the individual, attachment constructs this world, and the creation of Sanctuary reconstructs and restores the social and personal world of the individual. As an organisational change model, organisations and communities can be transformed through integration, understanding trauma, avoiding re-enactments, fighting rigidity, and embracing non-violence (Farragher and Yanosy, 2005). When an organisation can respond effectively to its own trauma issues, it can be more effective in treating traumatised youth entrusted to its care. This is done by enhancing the therapeutic community to strengthen mutual support, autonomy, and cognitive, social and behavioural strategies (Rviard et al., 2005). The community can be enhanced by faithfulness to the Sanctuary commitments of non-violence, emotional intelligence, democracy, open communication, social responsibility social learning and growth and change (Esaki et al. 2013). | The concept of trauma as applied to children is defined as “the mental result of one sudden external blow or a series of blows, rendering the young person temporarily helpless and breaking past ordinary coping and defense operations” (Scheidlinger, 2004).  The paper described PTSD as “resulting from exposure to events that involve actual or threatened death or serious injury or a threat to one’s physical integrity, when both internal and external resources are inadequate to cope with the threat. The individual’s response to these events may involve fear, helplessness, and horror, the danger being responded to physically, cognitively, emotionally, socially and behaviourally (Bloom, 1997l van der Kolk, 1989).” |
| Prchal (2005) | Three residential, youth therapeutic services specialised to treat youths with conduct disorders and other serious emotional disturbances, North-eastern USA | Name: The Sanctuary Model (adapted)  Aim: within the context of safe, supportive, stable, and socially responsible therapeutic communities, a trauma recovery treatment framework is used to teach youths effective adaptation and coping skills to replace nonadaptive cognitive, social, and behavioural strategies that may have emerged earlier as a failed means of coping with traumatic life experiences. | The Sanctuary Model was originally developed for adults and older adolescents in short-term inpatient treatment. It was adapted for purpose in this study for application with youths aged 12-20 years in residential treatment programs. While adapted for a younger target population, it was implemented in accord with the basic tenets of the original model  The implementation of the Sanctuary Model and subsequent changes in behaviour and organisational climate require an iterative learning process. Implementation begins with 8 hours of formal staff training (for milieu counsellors, supervisors and clinicians) in the basic principles of the model and methods of diffusing the model into the environment and all aspects of treatment. For example, in training sessions staff members begin to draft a Sanctuary Model mission statement for their units. Tools are introduced that will be used in the psychoeducation program, such as “safety plans” which offer safe and healthy engagement alternatives for youths when feeling unsafe, upset or uncomfortable.  Training sessions covered the following topics: (a) opening communication channels and team building, (b) using trauma theories to understand how traumatic life experiences can influence youths’ coping, adaptation, and social skills, © how to apply therapeutic community philosophy and a trauma recovery framework in working with youths, (d) how to diffuse the Sanctuary Model into team meetings and individual treatment planning, (e) how to facilitate therapeutic community meetings and involve youth in leadership roles, and (f) how to incorporate the Sanctuary Model in the larger environments of school, community, home and family.  Key components of the Sanctuary Model implementation included: development of a mission statement, community meetings held twice daily (the morning meeting to discuss plans for the day and identify someone whom a youth can talk to if they need to, and the afternoon meeting for discussing how the school day went and plans for the evening), community meetings co-led by staff and youths, and staff use of psychoeducational exercises on the residential units. | Devised by: the Sanctuary Model was originally developed by Dr Sandra Bloom (and team) in 1997.  Implemented by: clinical staff with oversight from the principal investigator | The Sanctuary Model is based in social psychiatry, trauma theories, therapeutic community philosophy, and cognitive-behavioural approaches. It rests on the premise that the therapeutic environment is a critical determinant in facilitating the recovery process. Successful implementation of the model not only requires the implementation of new treatment protocols but also requires change in the program philosophy and milieu toward a nonviolent and community-oriented paradigm, change in the organisation culture, and change in attitudes and behaviour of youth and staff. The treatment approach is organised around theoretical assumptions about the effects of trauma and a therapeutic framework for facilitating client movement through four stages of recovery (SAGE): (a) safety (attaining safety in self, relationships, and environment); (b) affect modulation (identifying levels of affect and modulating affect in response to memories, persons, events); (c) grieving (feeling grief and dealing with personal losses); and (d) empowerment (trying out new roles and ways of relating and behaving as a “survivor” to ensure personal safety and to help others). This framework is integrated into the primary therapeutic modalities of the Sanctuary Model, which include the therapeutic community itself, community meetings, and psychoeducation exercises and groups. | Not specified |
| Rivard et al.(2005) | Three residential, youth therapeutic services specialised to treat youths with conduct disorders and other serious emotional disturbances, USA | Name: The Sanctuary Model  To address the special needs of youth with serious emotional disturbances and histories of maltreatment and/or exposure to domestic and community violence. The Sanctuary Model aims to both strengthen the therapeutic community environment and to empower youths to influence their own lives and communities in positive ways. | The fundamental premise of the Sanctuary Model is that the treatment environment is a core modality for modelling healthy relationships among interdependent community members. The Sanctuary Model challenges organisations to re-examine their basic assumptions concerning the extent to which treatment environments promote safety and non-violence across physical, psychological, social and moral domains. The enhanced therapeutic community environment then sets the stage for the application of a trauma recovery framework (Foderaro & Ryan, 2000) and cognitive-behavioural strategies to teach youths effective adaptation and coping skills.  The trauma recovery framework is referred to as SELF, which represents the four stages of recovery (Safety, Emotional management, Loss, and Future).  ***Safety:*** is translated as learning to attain safety in self, relationships and the environment.  ***Emotional management:*** tasks focus on identifying and managing emotions in response to memories, persons and events.  ***Loss:*** involves feeling grief and dealing with personal losses.  ***Future:*** calls for trying out new roles and ways of relative and behaving as a survivor to ensure personal safety and help others.  The Sanctuary Model is operationalised through a series of staff dialogues and staff evaluations of residential units’ structure and functioning, staff training and ongoing technical assistance, twice-daily community meetings, a range of psychoeducational exercises that staff use in their daily interactions with youth, and weekly psychoeducational groups (Duffy, McCorkle & Ryan, 2002) to teach knowledge and skills needed to progress through four stages of recovery.  The psychoeducational groups are organised around the SELF recovery framework with elements of the therapeutic community philosophy. The contend of the curriculum used to conduct the groups is summarised below:  Sessions 1 and 2: Trauma Theory - The first two sessions are designed to help youths understand what is known and understood about the effects of trauma and violence; how people react to overwhelming stress with fight, flight, or freeze responses; how these stressful experiences affect peoples’ thinking, feelings, and behaviours; and how these experiences can overwhelm people’s ability to cope.  Session 3: Tools that Help People Build a Better Future - This session presents an overview of the SELF recovery framework and introduces concepts and tools that will be covered in following sessions. The primary theme of this session is that youths will learn healthy ways of coping and problem solving that will help them build better futures for themselves.  Session 4 and 5: Safety - The next two sessions are designed to help youths increase their understanding of safety and to teach tools needed to establish and maintain safety on physical, psychological, social, and moral levels. The terms are explained to youths in the following ways. Physical safety includes safety within the environment where basic needs for nutrition, shelter, and sleep are met; where individuals are free from harm; and where comfort is provided. Psychological safety includes feeling safe in one’s own mind. It means not hurting your own or others’ feelings. It includes the way you talk to your self and the way you talk to your peers, staff, and your family. Social safety means feeling safe and trusting other people. It is about choosing friends that you can trust. It also means being able to manage rough times and situations safely by talking rather than fighting. Moral safety means feeling safe enough to do the right thing, making good choices and doing your best to hold to them, respecting others, and having values that you and the people you are with live up to. If a peer is planning to hurt someone else, moral safety is telling the truth to protect your peers and staff.  Session 6: Safety and Boundaries – This session helps youths learn and practice safety principles with regard to boundaries. Youths are taught how to differentiate between physical and emotional boundaries, how to set their own boundaries, how to say yes or no when others want to come into their boundaries, and how to recognise when they violate the boundaries of others.  Sessions 7 and 8: Emotions – These two sessions focus on what feelings are and how feelings are an essential tool to have on the journey toward growing up healthy. Exercises show youths how to give names to feelings, how to think about what causes feelings, how to recognise various feeling signals in mind and body, how to understand ourselves better through our feelings, and how to manage the intensity of feelings without numbing or losing control.  Sessions 9 and 10: Loss – In these sessions youths learn how healing from loss is connected to safety and emotions and how this is connected to a better future, how difficult it is to grieve, how people can get stuck when they are not able to grieve their losses, and how people need support when they are grieving so their safety can be maintained.  Sessions 11 and 12: Future – These sessions provide an opportunity for youths to begin to think about their futures. The emphasis is on teaching youth to navigate through each day with a better understanding of how their futures are determined by their abilities to keep themselves safe, to manage their emotions, to overcomes losses they have experienced, and to make choices that will help them reach their desired futures. Exercises show youths that they have a choice in creating their futures and plant the seeds of hope for something different than what many youths have experienced in the past. | Created by: The Sancturary Model was developed by Dr Sandra Bloom (and team) in 1997  Implemented by: staff on the eight residential units (more detail not provided) | The Sanctuary Model integrates an enhanced therapeutic community philosophy (Bloom, 1997), trauma theories (Bloom, 1997), and cognitive-behavioural approaches. There is the idea that maltreatment and trauma can disrupt children’s abilities to form attachments with others, to process social information, to problem-solve, to discriminate between positive and negative behaviour, to regulate affect, and to develop accurate perceptions of self (Friedrich, 1996). Repeated exposure to maltreatment or witnessing family or community violence can result in accommodating to chronic stress in maladaptive ways (Connor, 2002; Fletcher, 1996; Terr, 1991). The Sanctuary Model proposes that within the context of safe, supportive, stable and socially responsible therapeutic communities, a trauma recovery treatment framework could be used to teach youths effective adaptation and coping skills to replace non-adaptive cognitive, social and behavioural strategies acquired as a means of coping with traumatic life experiences. | The paper states that “the term “complex trauma” is being used to describe both the aspect of multiple exposure to traumatic events, and the enduring symptoms that can be manifested as children attempt to cope and integrate these traumatic experiences into their concepts of self, others, and their world (Cook, Blaustein, Spinazzola & van der Kolk, 2003).” |
| Rivard et al. (2004) | Three residential, youth therapeutic services specialised to treat youths with conduct disorders and other serious emotional disturbances, North-eastern USA | Name: The Sanctuary Model  The aim of this model, adapted for adolescents in residential treatment programmes, is to strengthen the therapeutic community environment and empowering youths to influence their own lives and communities in positive ways. | The fundamental premise of the Sanctuary Model is that the treatment environment is a core modality for modelling healthy relationships among interdependent community members. The Sanctuary Model challenges organisations to re-examine their basic assumptions concerning the extent to which treatment environments promote safety and non-violence across physical, psychological, social and moral domains. The enhanced therapeutic community environment then sets the stage for the application of a trauma recovery framework (Foderaro & Ryan, 2000) and cognitive-behavioural strategies to teach youths effective adaptation and coping skills to replace non-adaptive cognitive, social and behavioural patterns acquired as means of coping with traumatic and other stressful life experiences.  In the Sanctuary Model, staff and youth share responsibility in problem-solving, decision-making and creating a safe environment. Youth are responsible for much of their own treatment and help each other in goal attainment. Decision-making is accomplished through group processes aimed at “creative consensus”. Emphasis is placed on raising awareness of conflicts and conflict resolution. Values, belief systems and norms of community are continuously defined and negotiated in a living-learning environment.  ***Staff training and technical assistance:*** Model implementation begins with eight hours of formal staff training in which milieu counsellors, supervisors and clinicians were trained in the basic principles of the Sanctuary Model and taught how to diffuse the model into the environment and into all aspects of the treatment program. Sessions cover topics including: (a) opening communication channels and team building; (b) using trauma theories to understand how traumatic life experiences can influence youth’s coping, adaptation, and social skills; (c) how to apply therapeutic community philosophy and a trauma recovery network in working with youth; (d) how to diffuse the Sanctuary Model into team meetings and individual treatment planning; (e) how to facilitate therapeutic community meetings and involve youth in leadership roles; and (f) how to incorporate the Sanctuary Model in the large environment – school, community, home and family. A major emphasis is placed on providing on-going technical assistance and consultation to staff in each residential unit to translate the Sanctuary Model philosophy, principles and language into daily programming, team meetings, treatment planning, community meetings, and work with families.  ***Therapeutic community:*** the philosophy of a therapeutic community is central to the Sanctuary Model (Bloom, 1997). The core values of a therapeutic community are: the community itself is the most influential factor on treatment; clients are responsible for much of their own treatment; the operation and management of the community should be more democratic than authoritarian; and clients can facilitate each other's treatment.  The Sanctuary Model adds to these values an emphasis on creating a “living-learning environment” (Bloom, 1997, p. 127) which is physically, psychologically, socially, and morally safe for both clients and staff. Establishing and maintaining a therapeutic community in the Sanctuary Model requires an active process of breaking down institutional, societal, professional, and communication barriers that isolate both staff and youths. Simultaneously, the re-building process involves consciously learning new ways to relate as interdependent community members, creating and modelling healthy and supportive relationships between individuals, and developing an atmosphere of hope and non-violence.  Diffusion of this enhanced therapeutic community philosophy begins with the development of mission statements, which are unique to each residential unit, but built upon the core Sanctuary Model principals of non-violence and shared community action. The notion of community is reinforced twice daily in community that are aimed at teaching youths how to rely on their community and how to become caring and responsible community members. A protocol is followed in which all community members share feelings, state their goals for the day, ask for specific help from other members in achieving their goals, share successes at the end of the day, and discuss ways to solve community problems. In addition, incentives for prosocial community behaviours are provided through community reward programs that are tailored to the needs and goals of individual residential units.    ***Psychoeducation Program:*** the therapeutic community environment sets the stage for delivering the psychoeducation program, which is organised around trauma theories discussed above and the SAGE recovery framework (Foderaro & Ryan, 2000). The SAGE recovery framework accents the critical stages and tasks needed to affect recovery from traumatic life experiences. The four stages are safety, affect regulation, grieving, and empowerment (Foderaro & Ryan, 2000).  The terminology used to describe the framework was adapted slightly to facilitate youths’ understanding of the meaning of these stages. For example, there are four types of safety. Physical safety includes safety within the environment where basic needs for nutrition, shelter, and sleep are met; where individuals are free from harm; and where comfort is provided. Psychological safety encompasses feeling safe in one’s own mind, not hurting one’s own or others’ feelings, and the ways you talk to yourself, peers, staff, and family. Social safety refers to feeling safe with and trusting other people and learning how to choose friends that you can trust. It also means being able to manage rough times and situations safely by talking rather than fighting. Moral safety means feeling safe enough to do the right thing, making good choices and doing your best to hold to them, respecting others, and having values that you live up to. Affect modulation is translated as emotional management; grieving is rephrased as loss; and empowerment is interpreted as focusing on the future.  A 12-session psychoeducation group curriculum has been developed to formally introduce youths to the Sanctuary Model and to teach knowledge and skills needed to progress through the four stages of recovery (Duffy, McCorkle, & Ryan, 2002). Groups are convened weekly and are co-led by clinicians and milieu counsellors. Each session has learning objectives, exercises, and ways of transporting information and practicing skills between sessions.  **Session 1-2:** Trauma and stress Concepts covered:   - Definition of stress and trauma - How people respond to stressful life experience in various ways (flight/fight/freeze) - How stress interferes with feeling, thinking and acting - How our relationships and experiences teach us to cope - How some coping patterns are not healthy and how people can learn new ways to cope that may be more helpful   Skills rehearsed:   - Recognising events that are stressful, threatening or dangerous - Identifying and understanding how stressful events trigger physical responses (adrenaline) and behavioural responses (fight, flight, freeze) - Recognising unhealthy ways that people learn to cope with unhealthy stress (withdrawing, excessive activity, addiction, fighting) - How to be aware when you are reacting to past stressful events in the present - Monitoring your stress levels and responses   **Session 3:** Four Stage recovery framework  Concepts covered:   - Introduction to the four stages: being safe, managing emotions, dealing with loss and change, moving on and making choices for the future - Linking stages with behaviours - How moving through the stages can help young people learn healthy ways of coping and problem-solving   Skills rehearsed:   - Differentiating the four stages and using the language - Mapping one’s life choices - Using the stages as guideposts in mapping the future - Practising self-help skills   **Sessions 4-5:** Safety  Concepts covered:   - Definition of the four types of safety: physical, psychological, social and moral - Applying the safety principles and learning new skills to establish and maintain safety   Skills rehearsed:   - Recognising and avoiding unsafe situations (i.e., threats of self-harm or threats of harm from or toward others) - Identifying ways individuals create safety in communities - Developing and practising how to use individual safety plans   **Session 6:** Boundaries  Concepts covered:   - Definition of personal boundaries and why people need them - How to create boundaries by asserting oneself - How to recognise when you might be entering another person’s boundaries   Skills rehearsed:   - Differentiating between physical and emotional boundaries - Setting personal boundaries - Saying yes or no when others want to enter your boundaries - Recognising when you violate the boundaries of others   **Sessions 7-8:** Affect regulation  Concepts covered:   - Feelings are natural and help us understand ourselves – what we like, what we don’t like - Links between emotions, behaviours, and decision-making - Recognising and managing emotions gives people more control over the present and helps us move towards our goals - The role of feelings in conflict resolution   Skills rehearsed:   - Naming feelings - Becoming aware of the cause of feelings - Recognising various feeling signals in mind and body - Managing the intensity of feelings without numbing or losing control - Using self-soothing skills - Identifying and practising healthy ways to express feelings   **Sessions 9-10:** Loss and grief  Concepts covered:   - How dealing with change and loss affects our safety, feelings, and ability to move on to the future - How people can get stuck when they are not able to grieve their losses - How people need support when they are grieving so their safety can be maintained   Skills rehearsed:   - Recognising personal losses - Reaching out to others when feeling sad - Supporting others who are feeling sad because of their losses   **Sessions 11-12:** Future  Concepts covered:   - How one’s future is connected to the ways we keep ourselves safe, manage our feelings, and move beyond our losses - How to make choices that will get you to the future you want - Planning for the future - Review of coping skills   Skills rehearsed:   - Identifying life skills that keep us safe - Describing oneself in the future - Listing steps to take to get toward the desired future - Practising the steps   Activities in the sessions include viewing and discussing videos, interactive activities, role plays, pencil and paper exercises, and writing exercises. At the last session participants are rewarded with a certificate of completion and encouraged to become role models as they begin to live their lives more aware of how connected they are to others and how their emotions, safety, and successes affect others.  Acknowledging that recovery is a non-linear and cyclical process, the 12 sessions are designed to be ongoing. Youths newly admitted to the program can enter the group at whatever stage is currently being covered. Youths who have completed a full round of the psychoeducation groups are expected to continue participating in the group with a new awareness of their own stage of recovery, to acquire new learning, and to bring their new skills and experience to help other members of the community.  Various exercises, used in the formal psychoeducation groups, were compiled into a supplemental manual that could be used during everyday program operations on the residential units.  Individual safety plans, jointly developed by youths and their counsellors to identify alternative ways to feel safe in unpleasant or stressful situations, could be put into action when youths showed signs of distress. At times when the whole community felt unsafe due to conflict or acting out behaviour, group exercises could be used to assist in identifying the cause of the conflict and alternative ways of problem-solving and working through conflict. Various paper and pencil exercises used in the psychoeducation groups for visualising the future, such as mazes and drawing oneself in the future, could be pulled out as needed to help youths consider possible outcomes of different choices. | Created by: the Sancturary Model was developed by Dr Sandra Bloom (and team) in 1997  Implemented by: milieu counsellors, supervisors and clinicians are all trained in the principles of the model. Psychoeducational groups are co-led by milieu counsellors and clinicians. Technical assistance was provided by clinical consultants with expertise in trauma, and monthly by the model developer. | The Sanctuary Model integrates an enhanced therapeutic community philosophy (Bloom, 1997), trauma theories (Bloom, 1997), and cognitive-behavioural approaches. There is the idea that maltreatment and trauma can disrupt children’s abilities to form attachments with others, to process social information, to problem-solve, to discriminate between positive and negative behaviour, to regulate affect, and to develop accurate perceptions of self (Friedrich, 1996). Repeated exposure to maltreatment or witnessing family or community violence can result in accommodating to chronic stress in maladaptive ways (Connor, 2002; Fletcher, 1996; Terr, 1991). The Sanctuary Model proposes that within the context of safe, supportive, stable and socially responsible therapeutic communities, a trauma recovery treatment framework could be used to teach youths effective adaptation and coping skills to replace non-adaptive cognitive, social and behavioural strategies acquired as a means of coping with traumatic life experiences. | Not specified |
| Six Core Strategies | | | | | | |
| Azeem et al. (2011) | Mixed- and single-gender child and adolescent (ages 6-17) Psychiatric unit Hospital, USA | No name assigned  Aim to reduce seclusions and  restraints among youth during psychiatric hospitalisation based on six trauma informed core principles | The hospital received training by the National Association of State Mental Health Program Directors (NASMHPD) regarding Six Core Strategies to be implemented in reducing restraints and seclusions in the unit. The strategies are based on trauma-informed and strength-based care, with the focus on primary prevention principles, and include: (a) leadership towards organisational change, (b) use of data to inform practice, (c) workforce development, (d) use of restraint and seclusion reduction tools, (e) improve consumer’s role in inpatient setting, and (f) vigorous debriefing techniques.  Senior administrative, medical, nursing, and senior school staff at the hospital were trained on the six core NASMHPD strategies.  ***Leadership towards organisational change strategies***: senior management created a goal of reducing seclusion and restraint. The vision was shared with all the staff in different meetings and communications. The reasons for this goal were shared, including avoiding traumatisation and re-traumatisation of the patients and staff, decreasing injuries, enhancing best practices, and ultimately improving treatment outcomes and staff morale. Hospital leadership developed a plan for reducing seclusion and restraints and allocating resources and looking at removing barriers to the plan. Looking and analysing seclusion and restraint data became a standing agenda item for Hospital Leadership Meetings and Medical Executive Meetings. The team that received the initial training met regularly to look at the progress and to implement various strategies and was called the Trauma Reduction Team. Goals and targets were established for the inpatient units with the primary teams including child psychiatrists, child psychiatric nurse practitioners, nurses, psychologists, social workers, and other direct care staff, and the information received at the national training was disseminated to the team members. Further measures to explain and inform staff included regular grand rounds, all staff meetings, and staff training besides monthly meetings to evaluate the progress of various steps.  ***Use of data to inform practice:*** data which were collected and analysed regarding seclusions and restraints included age, gender, ethnicity, date of admission, psychiatric diagnosis, unit, time of the shift, staff involved, injuries to the patient and staff. The data were shared in all staff meetings, regularly with clinical teams, and were posted on the respective units monthly. The data were helpful in creating healthy competition between the units and monitoring progress. Various supportive resources were infused for units that might be having struggles at different times. Clinical reviews were conducted in a non- judgmental and supportive way for patients requiring a higher number of seclusions and restraints, to look at various reasons for seclusions and restraints and what can be done in the future to prevent them.  ***Workforce development:*** the hospital created a treatment environment that was based on Trauma Informed Care, facilitates recovery, and was inclusive through development of staff education and training. The staff was educated during orientation as new employees, and regularly regarding neurological, biological, psychological, and social effects of trauma, and the prevalence of these experiences in patients receiving mental health services. The principles of recovery-oriented care, including person-centred care, respect, dignity, partnerships, and self-management, became an integral part of the staff trainings. These principles were also included in job descriptions, competencies, and performance evaluations. The staff was trained to avoid judgmental terms like “manipulative,” “borderline,” “attention seeking,” and “noncompliant.” This was regularly emphasised in morning reports and various clinical meetings.  ***Use of restraint and seclusion reduction tools:*** a variety of tools and assessments were included in the individual treatment plans. The staff at the hospital was retrained in utilising preventative measures including awareness about the patient’s trauma history, formulating and utilising safety plans, use of comfort rooms, occupational therapy techniques, and de-escalation approaches prior to the use of restraints and seclusions. Therapeutic communication between the patient, family, and staff was encouraged as the team set out to identify triggers and warning signs about each individual patient. Efforts were made to share reports of near misses and what worked in certain situations.  ***Improve consumer’s role:*** staff members were educated regarding the importance of patient and family involvement in safety plans and setting individual goals. The patients, families, family advocates, and case workers were encouraged and welcomed in the treatment meetings and evaluation conferences. All the units held regular community meetings involving the patients who gave suggestions for improvements on the units. Regular surveys involving the children and adolescents, families, and case workers were initiated. Complaints were addressed by management in a timely fashion.  ***Debriefing techniques***: Various debriefing activities were initiated, including immediate post-event debriefing and formal debriefing done within 48 to 72 hr. The debriefings were done in a nonpunitive and supportive way. The debriefings included the staff members and the patient involved. The immediate debriefing look at the emotional support needed for the patient and staff involved and any immediate changes required in the treatment plan. The formal debriefing looked at the incident in a root cause analysis fashion. This rigorous problem-solving procedure identifies what went wrong, what could have been done differently, and how to avoid similar incidents in the future. Interventions to mitigate the impact of traumatisation and re-traumatisation to the patient and staff were implemented. | Not specified | Specific guidelines to reduce the use of restraints and seclusion from the National Association of State Mental Health Program Directors. | Not specified |
| Azeem et al. (2015) | Mixed- and single-gender child and adolescent (ages 6-17) Psychiatric unit Hospital, USA | No name assigned  Aim to reduce restraint in a paediatric hospital by the implementation of a trauma informed and strength-based care approach. | This model was based on the Six Core Strategies.  Mechanical restraint beds once used at this facility were deconstructed and a symbolic “Healing Bench” was fabricated from the wooden frames of these beds.  The hospital encouraged use of verbal de-escalation techniques to intervene and support youth with unsafe behaviours.  Hospital executive leaders were available to the treatment units to assist in operationalising the tenets of optimal care, and for the provision of support and consultation for complex situations.  Restraints were used as a last resort when there was imminent risk to the youth or others. The topic of restraint reduction and prevention was a standing agenda item in executive meetings and regularly discussed in Executive and Leadership team meetings. The Medical Director, Director of Nursing, and Director of Operations reviewed every restraint the day after. The time interval was shortened for nursing and physician re-evaluation when a youth was restrained. Any youth who had two or more interventions in 12 hr or endured an episode lasting longer than a certain time period required notification to the Medical Director or designee. Use of the mechanical restraint bed was prohibited unless both the physician and the supervising nurse were present at the unit.  Data was collected on restraint use in each unit and shared across the facilities and clinical reviews with youth involved in multiple incidents of restraint were conducted non judgementally and supportively with staff, with the aim of building strategies to pre-empt future occurrences.  All staff were offered training and education on trauma-informed care with emphasis on use of verbal de-escalation techniques early in the crisis cycle.  New employee orientation was revised to include proactive approaches to engage youth through various coaching activities.  Many training initiatives were undertaken for hospital staff for learning and implementing specific principles of Dialectical Behaviour Therapy and Attachment, Regulation and Competency (ARC) framework. Trauma-Focused Cognitive Behavioural Therapy training was offered to clinical staff. Trainers were available for consultation during implementation.  Staff were given opportunities to visit other facilities with experience in restraint reduction.  Staff surveys were used to monitor staff satisfaction and training needs. “ASAP (Assaulted Staff Action Plan) Team” was created which provided immediate peer support for traumatised staff. The “ASAP Team” sponsored events to educate and reinforce the importance of staff’s well-being through self-care and to address vicarious trauma.  Team building activities, health education opportunities, and yoga groups were also scheduled on a regular basis to keep focus on the health of staff a priority.  Families were welcomed to the hospital before arrival of a youth to tour the facility, meet staff, and familiarise themselves with the principles of care and programming. The hospital clinical teams visited complex youth transferring from other facilities to begin the process of engagement and ease their transition into the hospital program.  An individualised treatment plan was developed comprised of individualised goals, target behaviours, triggers, early warning signs, coping tools, and safety plans. Youth’s specific interests, hobbies, coping skills, and motivators are incorporated into their plans.  Open communication between childcare staff and clinical staff was the key to implementation of treatment plans and problems incurred.  The occupational therapy consultant was utilised liberally to assess the sensory needs of youth and has appropriate tools available like sensory brushes, weighted blankets and vests, balls, and play dough that were used widely.  Youths had access to comfort rooms. Music, video games, a pool, gymnasium, and walks with staff have been frequently used as calming activities. Various sports were offered.  Staff worked with family members to identify strategies that work for de-escalation of youth. a STAR Team (group of staff members selected to be peer consultants and support specialists) was created. Child service, nursing, and clinical staff volunteers who were particularly skilled at conflict resolution and de-escalation of adolescent youths were dispatched as necessary to situations which were heading toward a pattern of conflicts or when youths themselves wanted to be seen by a member of the team.  Debriefing activities were undertaken, the first step was an immediate response to gather details of the event, check for injuries or trauma, and manage milieu. Followed by a formal, rigorous problem solving and planning meeting held within a few days for some of the most complex situations. Staff focus on any new information learned, and changes needed to safety plan and any alternative options for effective intervention were highlighted. Staff brainstormed around any interpersonal or environmental precipitant which could have been prevented. Youth debriefing focuses on chain analysis of the incident: understand what was upsetting, what was helpful, and what can be done differently next time to prevent the incident.  Many families were reimbursed for gasoline used to travel to family meetings, or medical cabs were accessed. Visitation by families has been encouraged and accommodated most hours of the day. There have been monthly family dinners to build closer relationships with staff at the hospital. The trust built between families and staff has given a deeper understanding of the family, youth, and their specific strengths and needs. Hospital staff visited the homes of adolescents to help with parent–youth interactional coaching for successful transition of youth to the communities.  For adolescents and families who were monolingual in their native language, interpreters were provided for the communication needs of those treating and those being treated when communication to staff was necessary.  Youths were given clothing vouchers, so outfits of their own liking were purchased.  Youth surveys implemented to identify areas of service and care in need of improvement.  Protection and advocacy staff have been present in the facility, and have educated, in a group setting, youths of their civil rights while hospitalised. | Designed by: not specified  Implemented by: not stated further than the psychiatric hospital | Not specified | Not specified |
| Duxbury (2019) | 11 mixed-gender and 3 single-gender adult, acute mental health wards from seven mental health hospitals, North-west England, UK | Name: "REsTRAIN Yourself"  Aim to reduce dependence on physical restraint as a means of risk management in acute mental health wards. | "REsTRAIN Yourself" is based off The Six Core Strategies approach, adapted for the U.K. context. The ‘Six Core Strategies’ for minimising seclusion and restraint is a multilevel complex intervention targeting both organisational and individual factors in decision-making about care in acute mental health services. The core strategies are leadership toward organisational change; the use of data to inform practice; workforce development; person-centred tools; service user roles within inpatient settings; and debriefing techniques.  REsTRAIN Yourself operationalises core strategies through a number of specific interventions targeted for a UK context as follows: (1) setting team goals for the reduction of restraint; (2) reflecting upon the use of restraint and personal communication styles (through reporting and analysing every restraint incident over a period of time); (3) using approaches to help patients and staff ascertain needs and challenges with regards to aggression on the ward; (4) employing partnership working strategies to reduce restraint such as ‘advance directives’ (my safety plan), and positive verbal and non-verbal communication; (5) exploring environmental challenges to make appropriate changes (both physical and procedural); and (6) debriefing following incidents or near misses of restraint.  A dedicated improvement advisor worked on the wards 1-day per week to support implementation of the model by: (1) identifying potential change ideas with staff; (2) the exploration of ideas and changes that teams would test using improvement science methods (i.e., Plan, Do, See, Act cycles); (3) Identification of group and individual roles and ownership whereby each member of the group commits to an action  A “train the trainer” model was rolled out. An online toolkit was developed alongside a 2-day face-to-face training event.  Local steering groups were established to track progress and sustainability of intervention. | Designed by: the Six Core Strategies approach was adapted to design REsTRAIN YOURSELF based on extensive consultation with stakeholders  Implemented by: A dedicated improvement adviser worked on the wards one day a week to support the implementation of the approach. Local steering groups were set up to progress ongoing spread and sustainability. The approach focused on identified champions for each ward and attendance at action learning sessions on a monthly basis. It is not mentioned whether these comprised of persons with lived experience. | The ‘Six Core Strategies’, which ReSTRAIN Yourself is based on, aim to minimise seclusion and restraint in a multilevel complex intervention targeting both organisational and individual factors in decision-making about care in acute mental health services. It is informed by trauma theory, and evidence that variations in restraint and seclusion rates are largely influenced by environmental or contextual factors. REsTRAIN Yourself adapted the six core strategies approach by revising language and generally modifying to suit the UK context. | Not specified |
| Chandler (2008) | An inpatient psychiatric unit, Massachusetts, USA | No name assigned  Aim of the TIC intervention not stated | The model was based on Six Core Strategies. Staff members— including nurses, physicians, social workers, and mental health counsellors—were educated on adopting and implementing the trauma-informed philosophy. Staff were taught to start with the skills that the patients have and so build on their strengths.  Cognitive–behavioural therapy and dialectical–behavioural skill training was offered daily.  A resource room was made available, with literature on diagnosis and coping skills, as well as written exercises, and with videos and audiotapes to assist patients in managing symptoms. | Not specified | Not mentioned specifically, but authors state: The foundation of trauma-informed care principles lies in the recognition that violence and victimisation play a central role in the lives of hospitalised consumers (Huckshorn, 2004). | Trauma is prolonged persistent and may originate in childhood. It results in fear, mistrust, depression, limited relational capacity, and negative coping behaviours, such as self-harm, dissociative behaviour, and aggression. Trauma can precipitate neurobiological changes such that the act of entering new and familiar situations can be interpreted as being threatening and fearful as a result of the tyranny of the past. |
| Chandler (2012) | An inpatient psychiatric unit, Massachusetts, USA | No name assigned  Aim to reduce restraint and seclusion. | The unit leadership provided an opportunity for a group of administrators and staff to participate in SAMHSA’s National Center for Trauma-Informed Care training. Following the training, leadership and staff committed to reducing restraint and seclusion use by building on the six core strategies (National Association of State Mental Health Program Directors, 2009) and adopting a trauma-informed care approach. To involve all staff, a retreat, followed by a series of mandated workshops on trauma-informed care, were created that included education on the neurobiological and psychosocial effects of trauma, the relationship of dissociative symptoms and self-harm to posttraumatic stress disorder (PTSD), and the re-traumatisation that occurs from being restrained or witnessing use of restraints and seclusion.  Staff members - including nurses, physicians, social workers, and mental health counsellors - were educated on adopting and implementing the trauma-informed philosophy. Staff were taught to start with the skills that the patients have and so build on their strengths  Cognitive–behavioural therapy and dialectical– behavioural skill training was offered daily.  A resource room was made available, with literature on diagnosis and coping skills, as well as written exercises, and with videos and audiotapes to assist patients in managing symptoms. | Not specified | Not stated | The paper states that “a traumatic event is defined as when an individual has experienced, witnessed, or is confronted with an event that involved actual or threatened death, serious injury, or threat that causes intense distress (American Psychiatric Association, 2000). Trauma that results in mental health problems is often repeated, prolonged, and severe, extending over time (van der Kolk, 2006).” |
| Hale (2019) | Child and adolescent (ages 3-17 years) inpatient behavioural health hospital, Illinois, USA | No name assigned  The aim of this project initiative was to implement a TIC programme to decrease physical hold and seclusion rates, thus decreasing a patient’s risk of being retraumatised, decreasing injuries, enhancing best practices and ultimately improving treatment outcomes and staff morale. | The TIC programme was based on the six core strategies established by the National Association of Mental Health Program Directors (NAMHPD).  Nursing and therapy staff attended educational sessions about the six core principles (see below), occurring weekly over the course of three weeks. Each session focused on two core strategies. The strategies were explained and examples of how they could be applied discussed. Each session was delivered by the researcher (a psychiatric nurse) to ensure consistency. Handouts explaining the six core strategies were also provided to dietary, admission and recreational staff. The content of the intervention and changes made are summarised below.  ***Leadership committed to organisational change:*** the physical hold and seclusion strategy was updated to ensure the Administrator on Call (AOC) was informed of all crisis interventions within a reasonable time frame, enabling leadership to then debrief promptly about the incident and relevant paperwork to be completed. The following day, the AOC would follow-up with the treatment team. Leadership also analysed trends in physical hold and seclusion data (including comparing data to other psychiatric hospitals within the corporate division). The Chief Operating Officer started town hall meetings to discuss staff’s concerns about the TIC programme. An all-star program was developed to recognise one staff member a month who had used de-escalation techniques effectively, and a ceremony held on their unit to celebrate this with both patients and staff (with refreshments provided).  ***Using data to inform practice:*** data were collected daily and aggregated and graphed to compare crisis interventions by floor, day of the week, and time of use in a day. Data regarding seclusion and restraint use was made available to staff and patients. Each month, the floor that used the fewest physical holds and seclusions was given a reception for staff and patients to celebrate it, and a banner was hung outside of the unit to mark this achievement.  ***Workforce development:*** TIC was incorporated into orientations for new staff. Mandated annual education for all staff about seclusion and restraint was also introduced. Presentation slides were distributed to staff annually about updates to policy changes and new interventions to reduce physical holds and seclusions – staff were also completed a test on this annually. Staff were also required to attend an annual refresher at the Crisis Prevention Institution highlighting de-escalation techniques. TIC was also added to all staff job descriptions and annual evaluations.  ***Use of seclusion and restraint prevention tools:*** assessment tools used on admission were revised to include trauma and aggression histories, and intake therapists were required to assess for trauma to help identify patients at-risk of aggression. Crisis interventions were explained to patients on admission. Nurses assessed patients’ current coping mechanisms, current and past triggers which could lead to aggressive behaviours and signs a patient may be approaching a crisis. If crisis interventions occurred more than three times during a patient’s stay, an additional meeting with the treatment team was held to discuss what could be done further to avoid restrictive crisis interventions. Break boxes (containing stress balls, playing cards, hand radios and other hand trinkets) were created to help patients de-escalate. Each floor had a comfort room providing a calm environment with music, a rocking chair, bean bag and sensory tools to also aid de-escalation.  ***Consumer of child/family and advocate roles in inpatient settings:*** the hospital had a patient advocate on site five days a week to respond to all complaints, issues or patient/parent concerns. The advocate provided support services for the patient, family and hospital. A milieu coordinator role was also created, to provide extra support for patients, families and staff and help to de-escalate situations to reduce the need for restrictive crisis interventions.  ***Debriefing techniques:*** debriefings were conducted immediately after a physical hold or seclusion occurred by the charge nurse or at the end of the shift by the AOC. A more formalised briefing in which camera footage of the incident was reviewed was also done within a few days with the clinical team. The focus of debriefings was to learn from the incident, and to update patients’ treatment plans and interventions to reduce the use of restrictive crisis interventions. | Designed by: The project was based on the six core strategies developed by the National Association of Mental Health Program Directors (NASMHPD).  Implemented by:  The TIC programme was implemented by staff at the service, including leadership staff, the Administrator on Call, the Chief Operating Officer. The researcher was involved in implementation (a psychiatric nurse with 24 years of experience in the field, and who is part of the hospital’s leadership team and restraint and seclusion performance improvement team). | The TIC programme was based on one developed by NASMHPD (Azeem et al., 2011): a comprehensive and holistic approach to management of behavioural health issues that includes six core strategies for prevention of traumatisation within the behavioural health setting. These core strategies include the following: a leadership team committed to organisational change, the use of data (internal and external) to inform the practice change, development of the workforce/staff, the use of specific tools (such as de-escalation) to reduce the use of physical holds and seclusion, ensuring that patients/family members have input, and the use of debriefing techniques as a learning tool when crisis interventions are used. | Incidents that are perceived as terrifying, shocking, and sudden, or create a threat to someone’s safety. Examples of traumatic events include but are not limited to all forms of abuse, neglect, war, abandonment, and natural disasters (Black et al., 2012). |
| Hale et al. (2020) | A child and adolescent (ages 4-17 years) psychiatric hospital**,** USA | No name assigned  Aim to reduce use of seclusion and restraint, and to avoid re-traumatisation. | The trauma-informed approaches used included: (1) staff education, (2) staff use of de-escalation techniques, (3) completed needed administrative updates, (4) enhanced communication, (5) patient education, and (6) culture change.  ***Staff education:*** education to reflect the trauma-informed philosophy and expected outcomes were given to new and seasoned staff; this was reiterated during annual competencies for all employees. Group therapy personnel were also updated to include trauma awareness and education, which included self-awareness and mindfulness techniques. Debriefings were a crucial aspect of staff education that moved the information into practice; much learning took place within this structure. If an employee used a crisis intervention, three debriefings were held. The first debriefing included employees involved and was an effort to get data before the end of the shift, while the occurrence was fresh on everyone’s mind. The second debriefing took place between the patient and a staff member who was identified as having the best rapport with the patient within 24 hours; patient input from this debriefing was included in the development of subsequent care plans. The goal was to record the patient’s perspectives on the experience, after they had had an opportunity to process and reflect on what had happened. These debriefings were routine prior to implementation of trauma-informed care at the facility. What was new was the final debriefing, which occurred within 48 hours of the incident, and included a reviewal of video documentation of the incident, before, during and after it occurred. This debriefing was led by the Restraint and Seclusion Performance Improvement Team (R&S-PIT) and included as well, the employees involved in the crisis intervention and the social worker assigned to the patient. Psychological safety of staff members was addressed by the R&S PIT team as each individual could themselves experience a traumatising reaction when reviewing the video. Psychological safety was assured as each debriefing began with a supportive statement identifying possible reactions on reviewing the incident and during debriefing. One member from the R&S PIT team was designated to meet with individual staff members as needed based on staff’s responses during the final debriefing. Private support meetings would occur as needed. Staff who may have been triggered by the debriefing would also be encouraged to contact the Employee Assistance Program as needed. This final debriefing became a powerful educational tool for all; the open discussion of the situation and results facilitated learning about how to infuse TIC into practice consistently; this opportunity to use the language and philosophy of TIC facilitated culture change as well.  ***Use of de-escalation as a tool to reduce seclusion and restraint use:*** de-escalation techniques that were included in the educational intervention provided to employees and used as needed by staff included: communicating caring with empathy in a calm manner; repeating simple messages as needed until they were heard; intentional use of body language that was nonthreatening, and approaching the patient one-to-one (as opposed to multiple people hovering nearby); listening, and responding, to the expressed needs of the patient; and setting clear limits that were simple. A major component of de-escalation in this population is the therapeutic use of self. This includes monitoring one’s own body language, speaking in a low and calm voice, using eye contact, and most important, expressing a comportment of empathy. These techniques were evaluated when the debriefings occurred, and if opportunities to substitute de-escalation for a physical hold or seclusion intervention was seen, it was used as a teaching point during the final crisis intervention debriefing.  ***Administrative updates, policies and procedures:*** related to the use of physical holds and seclusions were updated to reflect the processes and expected outcomes of infusion of TIC philosophy. Job descriptions were updated to include requirements to use TIC whenever appropriate; employees were held to these standards during their annual performance appraisals. Recognition of excellent employee use of TIC actions were recognised with an award, selected by R&S PIT committee members. All patient documentation from admission to discharge was updated and/or created to uphold the principles and practices of TIC.  ***Enhanced communication:*** along with debriefings, as care unfolded, if a particular patient would require three or more of the following interventions (a PRN medication for aggressive behaviours, physical hold, and or seclusion within 3 days), this would trigger a multidisciplinary discussion of the overall plan of care for the patient. Included in this discussion was the physician, the social worker/therapist, the nurse, and the Chief Nursing Officer. A deep discussion about the patient’s treatment plan; medication management; milieu management and family dynamics. At the end of this meeting, the care plan was updated to reflect the new interventions or new/current medications changes. In this way, individual patient data were used to inform practice and next steps.  ***Change in the culture of care:*** implementing a TIC program did not eliminate the need for crisis interventions as each crisis situation is unique. Once each staff member was educated in the philosophy and practical approaches used when implementing TIC, the culture for utilising physical hold and seclusions gradually changed. Staff went from quickly utilising a crisis intervention to avoiding these approaches until all other recommended TIC interventions had been exhausted. As well, staff learned that a crisis intervention involving a patient restraint episode did not necessarily require the patient to be placed, automatically, in seclusion. Staff began to see that other alternatives, such as having a patient go in their own room where they remained unrestricted could also be effective and was congruent with TIC. The TIC program provided staff with a heightened level of self-awareness. They were able to see an aggressive child in a different way that allowed them not to perceive their behaviour as personal or a reflection of poor care. Staff began to use de-escalation techniques learned during education and following debriefings and were able to see that this approach was successful in maintaining the dignity and care of the child, while keeping staff safe. Staff began using TIC language and approaches consistently during crisis incidents, using de-escalation, and debriefings; these, in turn, illuminated the emerging trauma-informed culture. A consensus of understanding developed with the staff that using crisis interventions increased a patient’s risk for re-traumatisation; and began approaching each patient situation as if the patient, indeed, had a history of trauma, whether or not they did. This became an underlying assumption of the TIC culture. This culture change process did not occur overnight; it took several months to see the change. However, as employees saw the results of their changed culture, there was more and more consistent use of TIC interventions, until this became the norm.  ***Summary of program implementation timeline:*** Awareness of the problem of too many uses of physical restraint and seclusion interventions led to a sense of urgency and buy-in from leadership which escalated the need for change. This was communicated to staff over a period of about 2 months before the educational interventions focusing on TIC with an emphasis on de-escalation techniques were rolled out. During this time, all needed administrative updates were completed. Multidisciplinary discussions and retooling of patient treatment and care plans began following the educational interventions as well. Implementation of the debriefing with video review followed approximately 2 months later. The entire process took about 6 months; culture change was in place by the end of 12 months. | The Restraint and Seclusion Performance Improvement Team (R&S-PIT) was charged to design, implement and evaluate the change, based on the literature review, the clinicians’ experiences and patient preferences. The R&S-PIT was formed with clinical experts, frontline workers (including the Chief Nursing Officer of the faculty), the Chief Executive Officer, the Chief Operating Officer, director of social work, nurse managers from each of the three floors, the staff educator, occupational therapist, and the Milieu Coordinator. Staff caregivers were always welcome and invited to every meeting. The R&S-PIT met monthly to review data and effectiveness of the interventions as they rolled out. | The TIC programme was based on the Six Core Strategies approach developed by NASMHPD (Azeem et al. 2011). This is a comprehensive and holistic approach to management of behavioural health issues that includes six core strategies for prevention of re-traumatisation within the behavioural health setting. These core strategies include the following: a leadership team committed to organisational change, the use of data (internal and external) to inform the practice change, development of the workforce/staff, the use of specific tools (such as de-escalation) to reduce the use of physical holds and seclusion, ensuring that patients/family members have input, and the use of debriefing techniques as a learning tool when crisis interventions are used. | Not specified |
| Comprehensive tailored trauma-informed model | | | | | | |
| Boel-Studt et al. (2017) | Psychiatric residential  facilities of a large Behavioural Health Agency, Mid-western USA | No name assigned  Aim to provide a wide range of programs and services aimed at helping families and children achieve improved functioning, well-being, and permanency | Prior to the agency integrating trauma-informed programming, the facilities provided 24 hr supervision and services to meet youths’ daily needs. The program provided ongoing clinical services including individual therapy and family therapy when feasible. In addition to stabilising psychiatric symptoms, services focused on improving self-concept and teaching effective social and problem-solving skills. The agency also used a family-centred approach that focused on the inclusion of families in decision making and treatment planning. The trauma-informed model included all the same service components as before in the traditional facilities along with several trauma-focused enhancements.  ***Organisational components:*** All staff received orientation, ongoing training, and supervision in understanding trauma and in working effectively with trauma-affected youth. Central to the approach was the creation of a supportive, therapeutic environment within the treatment setting where the program staff and youth are all viewed as members of a shared community. A key element of creating a trauma-informed culture and setting the stage for recovery involves working to achieve a sense of safety and self-awareness. All members, including the staff and youth, engage in safety planning in which they identify stressors, physiological cues triggered by stress, and strategies to proactively respond in stressful situations. Each member documented his or her safety plan and always kept it with them. Additionally, members identified a mission (i.e., goals and objectives that they hope to accomplish) and a shadow mission (underlying thoughts and behaviours that undermine their achieving the mission). Member check-ins occurred daily among youth and staff to discuss any issues or red flags that may be triggering psychological or physiological stress.  ***Clinical components***: Youth received individual trauma focused therapy including EMDR or TF-CBT and participated in a trauma recovery group-based curriculum 2 times per week. The trauma recovery curriculum was a multimodal, multi-layered program based on a combination of trauma treatments. The curriculum initially focused on helping youth achieve a sense of psychological and physical safety and trust through psychoeducation and supplementary exercises. The next phases focused on helping youth recognise the impacts of trauma, teaching effective coping strategies, and establishing a sense of hopefulness and goals for the future. The groups were led by staff who are trained in the curriculum and are comprised of approximately 8–10 youth matched by age. Program staff and therapists also worked with caregivers to provide trauma education and teach skills to help them support their child’s treatment. Within the treatment environment and as part of the individual treatment and trauma recovery groups, the youth were consistently exposed to prosocial and adaptive processes versus power and control. Although all youth were encouraged to participate in member activities, as a critical aspect of creating a culture of safety and support, participation was optional, and members could opt out without threat of consequences. | Designed by: The TIC programme was adapted in consultation with national trauma experts and a network that provides education, consultation, and training in trauma-informed services. | Similar to the previously described models (Attachment regulation and competency, the fairy tale model, trauma systems therapy), the approach is grounded in trauma knowledge and trauma-informed principles of care and includes both clinical and organisational components. | Not specified |
| Brown et al. (2013) | Three residential child and adolescent treatment units for young people with severe psychiatric disorders, Boston, USA | Name: Trauma Systems Therapy (adapted for residential settings)  Aim to help the youth become better regulated as well as to help stabilise the social environment that is contributing to their emotional dysregulation.  The specific aims at each of the three residential units were:  BOSTON IRTP: The TST program was initially implemented as a restraint reduction program.  The Children's Village: Not reported  KVC Health Systems: The overarching goal of the project was to ensure that youth leaving residential care received the same, consistent, child-specific TST services in the community upon discharge. | TST provides both an organising framework for identifying and coordinating the different service elements as well as a clinical model that describes exactly what providers do once they are brought together. The four primary service modules within TST include: 1) home- and community-based care; 1) outpatient, skills-based psychotherapy; 3) psychopharmacology; and 4) services advocacy. Each service is provided by different clinicians who are literally “brought to the table” to serve on a multi-speciality TST team to genuinely effect a comprehensive and collaborative treatment.  TST was originally created as an outpatient and home-based treatment model. A full description of the principles and practice of TST can be found in the published manual (Saxe et al. 2007). Key features of the adaptation of TST to residential care settings include:  ***The creation of a common language of care:*** TST helps to create a common language as well as shared goals and values to which all staff members, youth and families are exposed. For instance, staff members and families are taught to understand behaviour typically labelled as “bad”, “difficult” or “conduct disordered” instead as a youth in a dysregulated state who is reacting either to reminders of traumatic events or significant environmental stressors. Creating a context in which this kind of shared mutual understanding exists helps to change the nature of the therapeutic milieu to bring about lasting and meaningful change both in the individual and larger system.  ***Assessment and treatment planning based on a shared approach and using clinical data:*** A shared approach to assessment and treatment planning is important, as is using clinical data to focus on a small number of “priority problems”. Once these priority problems are defined, strategies are developed for how each service of the residential care setting (e.g., clinical, milieu, education) addresses the child’s priority problems. A primary assessment tool for identifying priority problems is the “moment by moment assessment”. This explores episodes in which the child has become dysregulated to look for patterns of contextual stimulation and emotional and behavioural responses.  ***A focus on the social environment of the residential milieu:*** an important adaptation is assessing for distress or threat within the social milieu. This acknowledges the fact that youth in residential treatment live within a unique social environment that may itself play a significant role in contributing to traumatic triggers/reminders or significant stressors and so dysregulation of the youth. Two types of potential distress or threat that were identified for assessment in the milieu included: violation of boundaries and violations of “team integrity”. Residential-based teams have a system and format for regularly discussing the functioning of the team and ascertaining whether individual team members (or the team as a whole) are contributing to a distressed or threatening environment. This process allows the team to develop solutions and proactively identify and address problems in a more productive manner.  ***Family involvement:*** family involvement in treatment is an essential component of assessing and addressing the child’s social environment. This can be achieved by rating the social environment. Residential staff using TST have been trained to engage caregivers by utilising the TST “Ready-Set-Go" module. This process involves aligning with caregivers by helping them identify and communicate what is most important in their lives – their life goals and “major source(s) of pain”. This allows the clinical to form a treatment alliance with the caregiver based on their own priorities rather than priorities imposed by the clinician. The clinician is trained to elicit the priorities of both the caregiver and youth and to consequently develop a treatment plan that is equally based on their input as well as the assessment conducted by the residential TST team.  ***TST as a vehicle to integrate care:*** TST is a systems-based treatment that can provide a framework agency-wide and across all staff. It is not a “one size fits all” intervention. It is a flexible, systematic approach to treatment planning based on frequent reviews of relevant data. TST is also an organisational model for systems change. TST requires commitment from agency leadership to make changes throughout every level of the organisation. TST emphasises the importance of training reaching all staff (including direct care staff), and all team members contributing.  Organisational innovations that can help with the successful implementation of TST in residential settings include hiring a full-time staff member as a TST coordinator; redefining the role of the lead clinician within the milieu to become the TST team leader; creating increased time for treatment planning meetings; and changing job descriptions and performance evaluations to align with TST benchmarks (e.g., adherence to the “10 TST Principles”).  The discussion in the paper states that successful implementation of TST in residential settings requires training staff at all levels (i.e., direct care and clinical staff); reconfiguring staff roles and responsibilities as well as creating new positions; increasing treatment planning time; and sharing TST concepts and language with youth and families. | Designed by: Glenn Saxe and colleagues (2007)  Implemented by: clinicians who make up a multi-speciality TST team | TST is a trauma-informed, flexible, systems oriented, empirically supported treatment model. It has a dual focus on a youth’s emotions and behaviours, and the role a distressed or threatening social environment may play in keeping a traumatised youth in a dysregulated state (both coming together to form a trauma system). TST proposes that it is important to assess the trauma system (both a youth’s emotional regulation capacity and the functioning of the social environment in which they live) as this will help to guide treatment. Treatment modalities are designed to help the youth become better regulated as well as to help stabilise the social environment that is contributing to the dysregulation. In TST, the clinical model is embedded in an organisational model. TST therefore describes what to do clinically, and how to integrate different clinical interventions so that children receive the right level of care, at the right moment in time, and in a tightly integrated manner. | Not specified |
| Cadiz et al. (2004) | Residential treatment programme for women with alcohol/other drug problems, mental health problems and trauma histories, New York, USA | Name: Project Portal: female-focused treatment strategy  Aim to intervene with the complex issues of trauma, substance use problems and mental illness as they occur among a female, addicted population within a residential drug treatment setting | The Portal Project documented the critical components of an integrated system: peer and consumer involvement, gender-specific services, culturally competent practices, cross-training attention to trauma, multidisciplinary team approach, and a dual focus on co-occurring disorders.  Gender specific treatment included a strength based, non-confrontational, safe, nurturing, and supportive environment. Multidisciplinary team models brought together professionals from different fields to develop a holistic portrait of the client, essential for an integrated system of care.  Emphasis was placed on the integration of the intervention into already existing treatment. The treatment setting had a strong behaviourally based milieu approach that has a long-standing history of difficulty adapting to new perspectives of human development. The integration was achieved by having the Women’s Treatment Specialist become part of the treatment team on the unit. In addition to being on the Treatment Team, the Women’s Treatment Specialist also co-led the group interventions with the mental health staff already employed at the intervention site.  System level integration for the Portal Project consisted of two key interventions: (a) monthly Multi-Disciplinary Team Case Conferences (MDTCC) and (b) quarterly Policy Action Committee (PAC) meetings. Each served to address the system level needs of the target population. The purpose of the MDTCC was to assemble expertise and representation from multiple service arenas; facilitate access to services from a variety of agencies and systems; foster positive change in the service delivery and practice areas; maximise information sharing and identify gaps and barriers to service. The PAC was designed to identify structural barriers and gaps in services among multiple systems. A core group of collaborators attended quarterly meetings with the aim of enacting policy changes that were supportive of the integrated treatment approach.  The curriculum differed slightly from group to group depending on the needs of particular members. Some chapters of Seeking Safety covered that were not in the original group format are: “Setting Boundaries,” “Dealing with Anger,” “The Life Choices Game” and “Healthy Relationships.” Another chapter that they more or less incorporated was the chapter on “Grounding”. | Designed and implemented by: Developed and implemented by the residential treatment programme service provider (Palladia Inc) | A prominent philosophy of this project and intervention is to treat a person from a holistic perspective; she is not a diagnosis, a trauma victim, or an alcoholic or addict, but a person, a mother, a friend, a daughter. The Portal model recognises that the issue of gender exists within many contexts: race, culture, age, sexual orientation, socioeconomic, class, ability, living situation, health status, and motherhood. In working with people with alcohol and drug misuse disorders and related syndromes, integrating treatment approaches to improve the chances for a successful outcome is critical (Kofed, Friedman & Peck, 1993; Sullivan & Evans, 1994). An integrated treatment approach means conceptualising the two syndromes of alcohol and other drug problems and mental illness as independent co-existing disorders, assessing clients for the presence of the disorders simultaneously (Sullivan & Evans, 1994), and understanding the diversity of substance-using and abusing women. | Not specified |
| Forrest et al. (2018) | Residential mixed-gender treatment programmes for young people (ages 12-22 years) with mental disorders, Massachusetts, USA | Name: Building Communities of Care (BCC)  Aim to maximise positive outcomes and minimise cost for youth and organisations (taken from abstract) | ***Integration***: BCC intentionally coordinates milieu programming, behaviour management, clinical intervention, and family relationships across systems with a common language. BCC was intentionally designed as a singular package on the premise that trauma-focused interventions are more effective if they are applied in a trauma-informed culture that minimises uncertainty and unpredictability.  ***Individualised:*** BCC is predicated upon the notion that trauma-impacted youth display a variety of, often complex, clinical profiles and that when it comes to treatment planning, “one size does not fit all”. Caregivers maintain routines, policies, and systems upon which they individualise care with a menu-based approach, including collaboratively developing treatment plans, individualising the environment for maximum predictability, comfort, and safety, and planning youth-selected activities. For example, bedrooms are decorated prior to clients’ arrival with the interests of each client in mind, while simultaneously eliminating possible triggers of dysregulation. Additionally, during intake parents and their children approve the types of non-verbal, verbal, and physical interventions appropriate if necessary. By individualising behaviour management interventions, the focus remains on the individual child’s needs; the goal becomes de-escalation through minimisation of threatening physical positions and maximisation of validation as opposed to applying standardised interventions to stop unsafe behaviour. Most importantly, caregivers are trained to attune to the unique needs of each youth through a continual practice of reflective listening and supporting client self-regulation.  ***Proactive:*** The role of BCC caregivers is to collaboratively design, implement, and maintain an environmental culture where instances of client dysregulation and difficulty become a rare occurrence. Milieu staff members, often the most available caregivers, support client capacity for self-regulation to proactively mitigate behaviours that may necessitate restraint, such as aggression, self-harm, and runaway behaviours. Caregivers are trained to attune to initial signs of dysregulation and respond with validating strategies to de-escalate before a negative strategy is needed. Attuning includes remaining aware of how caregiver body language and facial expression affects the situation and maintaining a nonthreatening approach. In the event a restraint occurs, caregivers utilise the least intrusive, preapproved restraint while remaining attuned to the child’s needs and level of dysregulation, so release occurs promptly. Post-restraint a debriefing period involves finding the dysregulation and escalation triggers in order to remove them as well as addressing potential re-traumatisation.  ***Implementation procedure:*** Trainings took place for eight hours a day over the course of three days and were led by the developers of BCC. Those who did not meet these standards were provided an opportunity to retake the training at a later point. Fidelity was ensured through the constant presence of at least two trainers in each establishment, with one being a senior trainer. Additionally, the application of the model was continuously monitored through quarterly senior trainer meetings and annual trained trainer meetings. Trainers were recertified yearly by completing eight hours of refresher certification training. | Designed by: The organisation established a taskforce that convened over 12-months to assess the degree to which their residential schools, group homes and treatment centres were trauma-informed. Stakeholder interviews with administrators, direct care workers, and consumers across six residential programs suggested inconsistent integration of trauma-informed programming with behaviour and crisis response management, which highlighted an important gap in residential programming generally. Consequently, a workgroup was established to develop an evidence-informed care model that integrated trauma-informed principles and behaviour management into an organisational framework. Development was guided by suggestions of leading organisations in residential care implementation (Whittaker et al., 2016) and trauma-informed out-of-home care.  Implemented by: Trainings were led by the developers of BCC. A train-the-trainer implementation approach was used. The application of the model was continuously monitored through quarterly senior trainer meetings and annual trained trainer meetings. | BCC is grounded in the empirically supported Attachment, Regulation, and Competency (ARC) Model (Blaustein & Kinniburgh, 2010; Hodgdon et al., 2015), which has been cited as a promising strategy for implementing trauma-informed care for children, youth and families impacted by chronic and severe exposure to maltreatment, violence or neglect (Bryson et al., 2017; Menschner & Maul, 2016). The ARC foundation constitutes enhancing children’s caregiver-child relationships (attachment), skills to manage internal, and interpersonal experiences (regulation), and key capacities associated with resilience (competency). ARC primarily supports applying skills associated with attachment, regulation, and competency to the processing of traumatic experiences (Hodgdon et al., 2015). BCC integrates ARC’s goals across systems through strength-based, individualised milieu strategies, and proactive and relationally driven behaviour management. | Not specified |
| Goetz et al. (2012) | Four psychiatric services: a residential treatment program for female adolescents (ages 12-18 years), an acute inpatient adolescent unit (ages 12-18 years), an adult inpatient unit (aged 19 years and above), and a sub-acute unit for adults with substance use and mental health problems requiring longer lengths of stay, Behavioural Health Services Hospital, Midwestern USA | Name: patient-focused intervention model  Aims to eliminate the use of seclusion and restraint in behavioural health settings, and to build a culture of safety | ***Organisational changes:*** one of the first decisions was to hire a director of clinical services with a master’s degree (MSN) in psychiatric nursing and extensive administrative and clinical experience in inpatient psychiatric settings. Another decision was to embrace a consistent model of patient care and aggression intervention that decreased the trauma of the inpatient experience. The state of Nebraska’s Behavioral Health Division created an organisation called Coercion Free Nebraska, which was committed to helping behavioural health providers who helped children move toward trauma-free care environments. As part of this initiative, national experts were brought into the state to introduce trauma-free concepts, and one of those experts conducted a workshop on trauma-informed care to introduce the local staff and community providers to these principles. The planning team used these concepts in mapping out the model for violence prevention.  ***Collaboration with patients, staff, administrators, and external experts:*** Feedback from these stakeholders allowed the continuous quality improvement cycle to evolve to excellence. Focus groups and surveys, as well as ongoing interaction with the security department and other ancillary departments, such as housekeeping and nutrition services, provided additional venues for feedback on the model. Collaboration with patients continued by conducting management rounds weekly to assess the impact of Caring Rounds and trauma-informed care. The management team took turns rounding weekly to meet with at least 10% of the patients to assess how well the staff was implementing the Caring Rounds and to ask patients what they liked about the care they were receiving and any suggestions they might have for improving it. Collaboration with all levels of staff in the facility was promoted through a shared governance model where representatives from all disciplines and shifts met on a monthly basis with a member of the management to make recommendations for changes in current practices, policies, and procedures and the current model of care. During monthly meetings with nursing and ancillary support staff, staff involvement continued to be reinforced, particularly through open discussion on issues related to safety and the quality improvement data.  ***Intervention in adolescent unit:*** To facilitate the implementation of trauma-informed care, in 2005, the hospital sponsored a workshop with Beth Caldwell, Director of Substance Abuse and Mental Health Services Administration (SAMHSA)/Center for Mental Health Services/Child and Family Branch’s National Building Bridges Initiative, on trauma-informed care. Key personnel from the hospital attended the 2-day workshop, which set the stage for the next 5 years and infused trauma-informed care principles into the treatment programs at the hospital. These components were incorporated into the initial psychosocial assessment through questions about past trauma; triggers that evoke anxiety, fear, and anger; and queries about which coping skills have been successful in the past. From this psychological assessment, for each patient, a safety plan that identified triggers and coping skills was collaboratively developed. Clear expectations for patient behaviours and involvement in daily activities were incorporated. An emphasis on patient responsibility was integrated into the staff’s approach to patients in the milieu. Power struggles were reduced as the staff focused on providing a healing environment. All these efforts were seen to help reduce the violence potential of the patients.  ***Aggression management:*** A new approach to aggression management was chosen based on an evaluation of the currently available commercial models. The Sorensen and Wilder Associates aggression management program (SWA) was selected as the one best fitting the organisation’s current needs. In this model, using the steps of a ladder as an analogy, the stages of aggression were conceptualised as ranging from 1 to 6 (Wilder & Sorensen, 2000). De-escalation techniques and hands-on skills were taught to every staff member. To assist with the adoption of the SWA process, 14 clinical staff were educated as trainers. The goal was that these trainers would increase trust in the SWA process and act as role models for other staff. Since instituting the program, staff has been recertified yearly in SWA techniques. Every 3 years, all staff complete a full certification. During their orientation phase, all new staff coming into the facility became certified prior to becoming involved in managing a violent patient event. The staff members were taught to use a “show of support” instead of a “show of force” with patients. This terminology was incorporated into the verbal interaction with the patient to reframe the situation into a less confrontational episode. All staff at this facility now consider touching or going “hands on” as a failure in the de-escalation process. Code event review: As the trauma-informed care and SWA techniques for managing aggressive patients were implemented, each of the aggressive patient events, known as a Code Gray, were subjected to a very specific review. Every incident was reviewed during a brief staff huddle (called a debriefing) immediately following an incident. In these reviews, the staff involved discussed not only the specifics of the event but also the techniques used or not used and the success of those used. To assist the staff in debriefing the event, an assigned scribe completed the “review of event” form during the discussion. This form collected data such as the patient’s aggression level, effectiveness of the response, safety concerns, and future recommendations. The debriefings during the initial six months included the clinical manager, the director of clinical services, and/ or a clinical educator, as well as unit staff. After the initial 6 months, the clinical program manager, the director of clinical services, or the clinical educator periodically attended an event debriefing to monitor the process and offer support to the staff. The main purpose of their participation in the debriefing was to reinforce education of the model and identify areas for improvement. Over the years, information gleaned from this process has resulted in environmental, policy, program, and physician practice changes that have also contributed to the continuous improvement process and the building of a safe environment.  ***Leadership involvement:*** As part of the leadership review, the nursing management, the nurse quality specialist, and the safety committee analysed monthly data on restraint and injury for trends such as time of day or unit where events were occurring. The team also conducted monthly in-depth reviews of the information on the “review of event” forms to determine areas of success and opportunities for improvement in the staff interventions being implemented. In 2009, daily leadership reviews of episodes of seclusion and restraint were initiated to increase the involvement of the medical staff and administration in the process. During the weekdays, there was a review of any patient aggression events with the leadership team of the behavioural health division. This review included the medical director, the director of behavioural health services, the nursing services manager, and the manager of clinical programs. A hand-off communication given by the house supervisors from the previous evening and night shifts was discussed with the oncoming nursing management staff. This frequent review by the leadership team allowed for discussion of difficult cases and formulation of recommendations for alterations in treatment plans for particular patients. It also promoted observation of trends in interventions with patients particularly the use of physical restraint and seclusion.  ***Intervention in adult units:*** The recovery model was incorporated into the adult and adult subacute programs in 2006. As an initial step, staff members were individually certified in the Wellness Recovery Action Plan (WRAP) approach. The WRAP approach is evidence based, with studies showing significant increases in consumer reporting of early warning signs, identification and tools for coping, their preference for using personal supports, the use of wellness in their daily self-care, and hope of recovery. The WRAP approach model fits well with trauma-informed care principles and provided a more specific focus on assisting patients to develop their own notebook of recovery. The unit staff assisted patients in developing daily maintenance plans, identifying triggers and early warning signs of aggression, and developing a crisis plan, advanced directives, and a postcrisis plan. WRAP groups were implemented on the adult unit and became part of the daily language used on the unit and within the broader violence prevention model. Patients often bring their WRAP notebooks back with them if they return for an inpatient stay. As part of this program, a peer specialist was employed as part of the adult unit staff and functioned as a patient advocate and liaison to the treatment team and administration. One of the activities of the peer specialist was conducting WRAP groups. The Caring Rounds are independent of routine 15-minute safety checks. Caring Rounds is a multidisciplinary set of rounds with the specific intent of assessing each patient’s feeling of safety, pain control, and medication response. At this time, patients can also raise questions regarding their treatment plan. The rounds, conducted by nurses and therapists, occurred at least three times per day to give each patient the opportunity to voice concerns and ask specific questions about his or her care.  ***Staff education:*** Education for all clinical staff included annual The Sorensen and Wilder Associates aggression management program certification, restraint and seclusion techniques, policies, and competencies, as well as many other formal, hospital-wide education topics. The main focus for education related to The Sorensen and Wilder Associates aggression management program and restraint/seclusion topics was de-escalation techniques. In addition, all new employees were oriented to the principles of trauma-informed care, safe boundaries, and patient centred care interventions. All nurses were expected to complete a series of four modules related to the role of the psychiatric nurse, basic psychiatric diagnoses and nursing interventions, and fundamental psychopharmacology. | The multidisciplinary team that planned and implemented the culture change model consisted of the director of clinical services, the director of risk management, the employee health nurse, and the clinical program manager for the residential treatment program. | The theory used to manage the change was based on the continuous quality improvement cycle of Plan, Do, Check, and Act (PDCA), also known as a Shewhart cycle or Deming circle (American Society for Quality, n.d.). | Not specified |
| Stamatopoulou (2019) | Inpatient female forensic mental health unit, Northern England, UK | No name assigned  Aim to encourage staff to utilise the skills learnt during the training to minimise ways in which the ward may contribute to re-traumatisation, in order to cultivate a welcome and safe environment within the ward. Staff was also encouraged to allow for greater flexibility and support service user input when establishing norms and rules. Staff were encouraged to utilise the skills to help service users establish a feeling of safeness. | This trauma-informed care pilot intervention was implemented in the forensic service through the following:  ***Staff training:*** all staff received a two-day development and training programme at the beginning of the implementation process to increase their knowledge of the impact of trauma, to develop skills in crisis intervention and to develop practical skills to work with trauma on a ward level. Content of the training included:   - Why do we need trauma-informed care, what do we mean by it and what does it look like? - Vicarious trauma and staff wellbeing-why are we all burnt out? - Understanding why the ward can trigger someone’s trauma - Attachment theory and links with abuse and violence - Insecure Attachment styles - Relationship difficulties - Patient-Nurse interpersonal complexities - The importance of reflecting on these relationships - Implications for clinical work - What is trauma and acute stress reactions - how do people become traumatised? - PTSD and Complex PSTD - Diagnosis vs formulation - Developing compassion - The effect on our emotions - Trauma informed interventions - Overview of processing therapies: EMDR, CAT, narrative therapy, schema therapy, CBT, DBT - 7 domains of skill development: mindfulness, multi-sensory grounding, emotional regulation, distress tolerance, interpersonal effectiveness, meaningful activity, positive action/connection/recovery - Opportunity for everyone to discuss the roll out of TIC in the unit, fears and expectations - Next steps, what now, how do we get this off the ground, ideas, what support do you need? - Review of pilot   ***Establishing a welcoming and safe environment:*** staff were encouraged to utilise the skills learnt during the training to minimise ways in which the ward may contribute to re-traumatisation, in order to cultivate a welcome and safe environment within the ward. Staff were also encouraged to allow for greater flexibility and to support service user input when establishing norms and rules. Staff were encouraged to utilise the skills to help service users establish a feeling of safeness.  ***Establishing safety plans:*** clinicians developed safety plans with service users through care planning and five sessions of cognitive analytic therapy which identified triggers, emotions and coping strategies to prevent and manage crises.  ***Establishing activities on the ward:*** core sessions of art, occupational activities, self-soothing, emotional regulation and mindfulness were offered within service user care plans as techniques to promote a sense of calm and safety.  ***One-to-one debrief sessions:*** service users were given a one-to-one debrief following incidents of seclusion, deliberate self-harm or restraint to promote healing, recovery and learning, as well as re-establishing the therapeutic relationship.  ***Trauma Champions:*** a member of staff was nominated as the daily Trauma Champion. This role involved ensuring that core sessions followed the need of service users using a passport system that identified which activities were re-energising and which were grounding. The Trauma Champion also noted down issues which needed to be raised within reflective spaces.  ***Fortnightly reflective groups:*** staff and service users engaged in fortnightly reflective groups, including a cognitive analytic therapy reflective group. Staff were also offered weekly supervision to explore any issues relating to the TIC pilot. | Designed by: not specified  Implemented by: The TIC lead for the NHS Trust oversaw the implementation of TIC across adult services in the Trust. All staff attended a two-day development and training programme about TIC and the pilot intervention. Specific roles of staff in implementation not described. | This TIC pilot intervention was based on principles of the trauma-informed organisational change model proposed by Harris and Fallot (2001). The authors state that TIC involves having “knowledge of service users’ history of trauma and its impact and on the other hand to utilise this knowledge to design service systems which accommodate service users’ vulnerabilities while allowing them to actively participate in their treatment (Harris and Fallot, 2001)”. | Defines psychological trauma as “events or circumstances which are experienced as hurtful or life-threatening and that have lasting effects on the emotional, physical and/or social wellbeing of a person (SAMHSA, 2014). Trauma may include witnessing or experiencing a single event such as an accident or trauma may result from repeated exposure to extreme external events and circumstances such as ongoing abuse or torture (Terr, 1991).” |
| Tompkins & Neale (2016) | A female residential substance use treatment unit, UK | No name assigned  No aims specified | ***Staff training and support:*** staff were all trained in trauma-informed approaches and received regular professional support and supervision from within the organisation. Every month, they also received team support and supervision from an experienced trauma therapist who was external to the organisation. The organisation also commissioned two days of intensive trauma-informed staff training from the US author of a manualised treatment programme.  ***Staff recruitment:*** managerial staff stated that they actively recruited women who had direct personal experience of recovery and women with prior experience of working with women and/or substance users. They sought to recruit staff whom they considered to be compassionate, understanding, nurturing, dedicated, self-disciplined and assertive.  ***Screening for trauma:*** all potential residential clients were screened for trauma on referral.  ***Support offered to clients:*** clients participated in a range of group therapies based on manualised trauma-informed treatment programmes. They were also offered individual counselling, EMDR, and family support according to their specific needs, had access to a structured programme of education skills, training, and recreational activities, and attended local fellowship meetings (Narcotics Anonymous and Alcoholics Anonymous). | Designed by: Not specified  Implemented by: All staff were trained in trauma-informed approaches and received regular professional support and supervision from within the organisation. Every month, they also received team support and supervision from an external experienced trauma therapist. Staff also attended two days of intensive trauma-informed training from the US author of a manualised trauma treatment programme. | Trauma-informed working is based on the assumption that all addiction treatment clients have experienced some form of trauma, so the approach focuses on their personal strengths to avoid disempowering them further or re-traumatising them (Covington, 2008; Elliott et al. 2005; Killeen, Back & Brady, 2015). Trauma-informed working can draw upon a range of integrated treatment programmes such as the Trauma, Recovery and Empowerment Model (Harris, 1998), Seeking Safety (Najavits, 2002) and Helping Women Recovery (Covington, 1999). Trauma-specific psychotherapy treatments may also be used, such as EMDR, which aims to reduce the long-lasting effects for distressing memories by developing more adaptive coping mechanisms. The paper also states in the introduction that “safety, trustworthiness, collaboration, choice and empowerment have been identified as the five core values of trauma-informed programmes (Fallot & Harris, 2009; Harris & Fallot, 2001; Harris & Fallot, 2004)”. | Trauma is defined “as ‘exposure to actual or threatened death, serious injury or sexual violation’ (American Psychiatric Association, 2013).” |
| Zweben et al. (2015) | Residential substance use treatment program for women who are pregnant or have young children, California, USA | Name: Celebrating Families! (CF!)  Aims to provide education and skill building to families who have been impacted by domestic violence, child abuse or neglect and by substance use disorder. It aims to increase the resiliency of all participants, strengthen their family and social connections and decrease their isolation. | Celebrating Families! (CF!), a program designed specifically for families in which one or both parents have a serious problem with alcohol and other drugs and are at high risk for domestic violence, child abuse, and neglect.  Participants in CF! Include the mother and child (or children) in residence, grandparents, friends, aunts and uncles, siblings, and other supportive individuals in the mothers’ lives. It focuses on family-centred treatment practices and is a multifamily/multigenerational event.  CF! Sessions are provided weekly over 16 weeks, enabling all residents and their identified extended family members to attend. Each session of CF! Is a three-and-a-half-hour program which begins with a family meal followed by subgroup programming for adults, children and adolescents, with age-appropriate materials.  Following the family dinner, participants attend a 90-minute instructional session on the following themes: 1) healthy living; 2) nutrition; 3) communication; 4) feelings and defences; 5) anger management; 6) facts about alcohol; tobacco, and other drugs; 6) addiction as a disease; 8) the effects of addiction on the whole family; 9) goal-setting; 10) making healthy choices; 11) healthy boundaries; 12) healthy friendships and relationships; and 13) individual uniqueness.  Patients then reunite with their children for a 30-minute activity to practice what has been represented and learned and to give feedback on their performance.  Mothers are carefully assisted in identifying family, friends, partners, and significant other who are, or can become, part of a positive, safe and supportive community for the women when they leave treatment.  Implementing CF! In Project Pride contains elements of a prevention strategy since many family members are at high risk for child abuse, as well as an intervention strategy since the programme provides direct treatment to children who are trauma survivors. | Developed by: Tisch & Sibley, 2007, within the Centre for the Study of Social Policy (CSSP)  Implemented by: staff at the residential treatment programme (more specific details not provided) | CF! is an evidence-based family-focused cognitive behavioural support group model. CF! aims to increase resiliency and decrease risk factors, integrating addiction recovery concepts with family living skills. It emphasises building strengths rather than identifying weaknesses. A broad definition of “family” is encouraged. | Not specified |
| Safety focussed tailored trauma-informed models | | | | | | |
| Blair et al, (2017) | A mixed gender psychiatric inpatient service of a large urban hospital, Connecticut, USA | No name assigned  Aim to reduce use of seclusion and restraint | Model components included routine use of the Brøset Violence Checklist (BVC), mandated staff education in crisis intervention and trauma informed care, increased frequency of physician re-assessment of the need for seclusion and restraint, formal administrative review of seclusion and restraint events and environmental enhancements (e.g., comfort rooms to support sensory modulation).  The BVC was incorporated into the required daily documentation and was completed by a physician on admission and by nursing staff during each of the three nursing shifts throughout the hospitalisation. This instrument assesses six behaviours associated with increased risk of violence: confusion, irritability, boisterousness, verbal threats, physical threats, and attacks on inanimate objects. The investigators added, with written permission from the BVC authors, a checklist of staff intervention options: verbal de-escalation, diverting activity, reduced stimulation, sensory modulation/comfort measures, medication, continuous supervision, seclusion, and restraint. This checklist was part of the required documentation for seclusion and restraint events.  Staff education included a standardised 8 h crisis intervention course that emphasised de-escalation techniques, a new method of nursing assignments to maximise staff presence in the milieu and training in ‘‘Risking Connections’’ (a two-day program based on a trauma-informed model of care, the goal of which is to reduce staff behaviours that can exacerbate ‘‘trauma reactions’’ in patients). There were also changes in hospital policy and procedures about the physician order for renewal of seclusion and restraint: the frequency of physician review was increased to every 2 h from every 4 h for patients over age 18 (frequency for patients\18 remained every 2 h).  Changes in the seclusion and restraint protocol required that the Medical Director and the Director of Nursing examine all seclusion and restraint events to determine if a formal administrative review was needed (based on severity and outcome) and that they personally conduct all such reviews, the format for which included questions about staff knowledge of the patient (e.g., BVC scores, history of violence, medications prescribed), the specific de-escalation interventions used and the communication about the patient’s status prior to the event.  Environmental enhancements included assessing the patient’s “sensory diet” on admission (e.g., identifying personalised coping strategies for decreasing anxiety/agitation) and creating comfort rooms (e.g., areas with calming lights, sensory items, music). | Not specified | The intervention was based on published evidence-based therapeutic practices for reducing violence/aggression. | Not specified |
| Borckardt et al. (2011) | Five inpatient psychiatric wards: an adult unit, a geriatric unit, a general adult unit, a substance abuse unit, and a child and adolescent unit at one large state-funded psychiatric hospital, South-eastern USA | No name assigned  Aim to reduce use of seclusion and restraint | An engagement model was developed and comprised components (trauma- informed care, rules and language, therapeutic environment, and patient involvement in treatment planning) that were organised and operationalised to be as uniform as possible between the diverse inpatient units. Each phase of the implementation schedule was three months in duration.  For trauma-informed care, all unit staff attended a half-day standardised training seminar on the nature of trauma and its effects on patients’ experiences, physiology, and psychological processes, along with instructions on how to minimise engaging in behaviours that could exacerbate trauma-related reactions from service users.  For rules and language, all unit staff attended a standardised training seminar on the effect of rules and language on service users’ perceptions. The rules and language intervention included the establishment of a team for each unit that was tasked with reviewing and modifying unit rules and policies to be less restrictive to patients or eliminating unit rules that were too restrictive. All unit staff attended a follow-up half-day seminar during which the rule changes were articulated and the effect of coercive language on patients’ experiences was discussed.  All signs on the units were reviewed and revised as necessary to ensure that they reflected the new, less restrictive rules and used non-coercive language.  The therapeutic environment intervention involved making inexpensive physical changes, including repainting walls with warm colours, placement of decorative throw rugs and plants, and rearrangement of furniture to facilitate increased service user-staff interaction, as well as holding staff-service user group meetings regularly on the unit. Two separate therapeutic environment interventions were allotted to each unit. The second intervention included replacing worn-out furniture and continuing with environmental changes initiated during the first intervention.  For patient involvement in treatment planning, all unit staff attended a half-day standardised training seminar on the rationale for and clinical benefits of involving patients in the treatment planning process. | Not specified | Not specified | Not specified |
| Jones (2017) | National high secure inpatient psychiatric service for women, East Midlands, UK | Name: Trauma and Self Injury programme (TASI)  Aimed to provide a model of care underpinned by a clear philosophy that would give meaning to practice interventions to help meet the needs of women detained in the service. | TASI is a bespoke trauma-informed framework of care that was developed to address the specific needs of women patients who were known to require high secure forensic healthcare. It is delivered via three interconnected layers of care including: proactive interventions, direct practice, enhancement of the therapeutic milieu, and the provision of individual and group therapies.  TASI was developed as a dynamic approach and consists of three distinct levels of service provision all of which are informed by a patient reference group.  ***Level 1:*** focuses on proactive interventions to promote understanding through training, on ward support and supervision of staff, and through the provision of psychoeducation and wellbeing groups for women patients.  ***Level 2:*** encompasses the impact on direct practice and the enhancement of the therapeutic milieu in delivering a safe collaborative environment which promotes the development of women’s capacity to deal differently with their distress. The use of Individual Distress Signatures, multidisciplinary team formulation and a framework for responses along with increased expertise in dealing with physical need are considered central.  ***Level 3:*** concentrates on the provision of individual and group therapies.  On-going evaluation of the TASI programme is integral to the development and review of overall impact and effectiveness. | Designed and implemented by: A multidisciplinary team of healthcare professionals working within the National High Secure Health Care Service for Women (NHSHSW) developed the TASI in 2007 when the NHSHSW opened. The multidisciplinary team consulted with a patient reference group. The researcher involved in this study was a member of the team who developed, implemented and evaluated the bespoke conceptual framework for the TASI. The researcher was a member of the clinical team working within the service, and so was known to all the participants in the study. | Care delivered within the NHSHSW is grounded within the guiding principles of trauma-informed environments. This philosophy of care understands that women patients in high secure forensic healthcare experience heightened and usually chronic levels of distress which can be communicated through violent and dangerous behaviour (McMillan & Aiyegbusi, 2009). | Not specified |
| Trauma informed training intervention | | | | | | |
| Aremu et al. (2018) | Inpatient adult psychiatric unit, Illinois, USA | No name assigned  Aim to strengthen trauma-informed practice by increasing patient engagement, and improving staff’s skills at intervening in escalating situations | Leaders within the service made a commitment to implementing trauma-informed principles and using best evidence to reduce the risk of retraumatising service users and keeping service users and staff safe.  Existing and new staff received education on patient engagement and improving staff skills at intervening in escalating situations. This included educating staff on their role in creating and maintaining a trauma-informed organisation, building trusting relationships, and creating a safe, healing environment.  Educational components included strategies focused on increasing the amount and quality of staff engagement with patients. | Designed by: Unclear  Implemented by: the Inpatient Adult Psychiatric Unit. Unclear who delivered the training. | Specific theoretical model not stated. However, the intervention aligned with trauma-informed practice principles that argue how engagement and improving skills are key components of trauma-informed practice. | Not specified |
| Gonshak (2011) | Female adolescents in a residential treatment centre for children who have severe emotional disabilities, Kentucky, USA | Name: Risking Connection  Aim to inform teachers of the effects of abuse and neglect trauma and to provide them with strategies to better connect with their students. | ***Staff education and training***: the three areas emphasised in the Risking Connection curriculum are: (1) an overarching theoretical framework to guide work with trauma and abuse survivors; (2) specific intervention techniques to use with survivor clients; and (3) a focus on the needs of trauma workers as well as those of their clients.  The Risking Connection curriculum emphasises how relationships can be transformative, whether they are brief or long-term, whether in a one-to-one or in a group context. The training curriculum deals with understanding trauma, the importance of relationship between the client and the treater, safety and transforming vicarious trauma experienced by the treater.  ***Understanding trauma:*** the first part of the training curriculum deals with understanding trauma. It emphasises that it is an individual’s subjective experience that determines whether an event is or is not traumatic. Risking Connection specifically teaches the treater about trauma and encourages the treater to teach the adolescent survivor about trauma as well. The curriculum suggests the treater be alert to “teachable moments” – times when the client can use specific information, dispute a belief, or when the treater can directly model a desired skill or offer a new perspective.  ***Importance of relationships:*** consistent with attachment theory is the assumption of the Risking Connection curriculum that the connection or relationship between the client and the treater is itself part of the clinical intervention. The curriculum teaches how attachment plays both a psychological and physiological role in mental health by shaping how one views relationships, and one’s ability to interpret and regulate emotions. It states that a therapeutic alliance with a survivor of childhood abuse works in the following ways: (1) it contradicts the client’s assumptions that all relationships will be abusive or exploitative; (2) when alliances last over time, the client can use them as a basis for forming a secure attachment, this can be to one person, a team, or to an agency or system; and (3) it diminishes the isolation experienced by many survivor clients. Risking Connection identifies the four components of a growth-producing therapeutic relationship: Respect (validation), Information, Connection and Hope. It also considers the biological function of attachment. It emphasises that when treaters make it a priority to help the adolescent calm down physiologically and later help them understand and interpret their emotions, they are increasingly able to self-regulate and make connections between their past traumatic experiences and current functioning. Specific strategies include: prioritising relationship-building; directly teaching about the effects of trauma; providing assistance with and role modelling effective calming techniques; collaboration of treatment goals; and using “restorative tasks” as a consequence when an adolescent has done something to harm a relationship.  ***Safety:*** the importance of maintaining overall physical and emotional safety while establishing boundaries with others is a key component of the Risking Connection curriculum. It trains treaters how to recognise and respond to dissociative episodes and flashbacks, and how to keep a trauma framework when responding to life-threatening or other dangerous behaviours. The trauma framework helps treaters manage their own anxiety, keep events in context, and points them toward helpful responses to the client’s symptoms. Treaters focus on self-capacity development such as (1) managing feelings; (2) building an inner connection to others; and (3) increasing self-worth.  ***Transforming vicarious trauma:*** the Risking Connections curriculum includes acknowledging and understanding vicarious trauma experienced by the treater. There is the idea that if treaters ignore their own needs, levels of stress and emotional experiences then they will be more likely to respond to clients in ways that create distance and disconnection or discontinue working with them altogether. This can reinforce many of the negative cognitive schemas the adolescent is working to change. Risking Connection training acknowledges these feelings, teaches treaters to assess their own levels of vicarious traumatisation, and to manage it with strategies such as self-care, self-nurturing activities, and healthy ways to emotionally escape. Ultimately, treaters learn to transform their vicarious trauma by creating meaning, challenging negative beliefs, and participating in community building, all of which is modelled to the adolescent client. | Designed by: Saakvitne et al.  Implemented by: teachers at the residential treatment centre. It is not stated who trained the teachers, apart from that they typically train clinical and direct-care staff in residential treatment centres. | Regarding the theoretical foundation of Risking Connection, two of the four authors of the curriculum, Laurie Anne Pearlman and Karen W. Saakvitne, also developed Constructivist Self-Development Theory (CSDT). As such, CSDT is the framework for the curriculum. It emphasises (1) the healing power of the relationship between the treater and the survivor; (2) views symptoms as adaptations (i.e., seeks to understand the meaning of behaviours rather than solely focusing on controlling them); (3) posits that crises can best be managed and eventually reduced through the development of 'feeling skills;' and (4) expects the work to have an impact on the treater that parallels the impact of trauma on the survivor (Saakvitne et aI., 2003). | The authors define psychological trauma as occurring “when [an event or situation] overwhelms the individual’s *perceived* ability to cope, and leaves that person fearing death, annihilation, mutilation or psychosis. The individual feels emotionally, cognitively, and physically overwhelmed. The circumstances of the event commonly include abuse of power, betrayal of trust, entrapment, helplessness, pain, confusion, and/or loss”. |
| Nimura et al (2019) | 29 inpatient psychiatric units across Tokyo, Japan | No name assigned  Aim to improve knowledge of trauma and TIC and motivate mental health professionals to find a means of embedding TIC into their clinical practice. | The TIC training programme consisted of a 3.5-hour lecture including a brief group discussion and 1-hour group discussion. The training programme content was described as follows:  Lecture including short group discussions (3.5 hours):   1. Definition of trauma and TIC 2. Evidence on trauma and TIC 3. Behavioural, social and emotional responses to traumatic events 4. Psychiatric clinical practices evoking patient’s re-traumatisation, psychiatric clinical practices corresponding to TIC, fostering trauma-informed sensibility, characteristics of trauma-informed clinical teams   Group discussion content (1 hour)   - Review of clinical practices based on TIC - Find a way to embed TIC into tomorrow’s clinical practice | Designed by: one author (MK), Professor of Psychiatric Nursing, specialising in TIC, developed the content of the programme  Implemented by: author MK served as instructor and facilitator. Other co-authors (JN, MN, and AN) served as co-facilitators. | “The content of the programme was based on the literature regarding TIC (Hopper *et al.* 2010; Substance Abuse and Mental Health Services Administration, 2014a) and the Gellinudd Recovery Centre, managed by a third-sector organisation named Hafal in Wales, UK (Hafal, 2018)”. | Not specified |
| Other trauma informed models | | | | | | |
| Beckett et al. (2017) | A psychiatric inpatient ward, with a high dependency unit and an acute unit, New South Wales, Australia | No name assigned  Aim to improve the quality of care provided to consumers. The approach taken to achieve this goal was based on the belief that embedding the principles of TICP in ward practices, which promoted values such as choice, collaboration, trustworthiness, safety, and empowerment, would lead to practice improvement and cultural change. | The ward Clinical Nurse Consultant and Senior Clinical Psychologist devised a series of workshop sessions to raise awareness of trauma and trauma-informed care and stimulate discussions on how they might be useful in improving practice. Through these workshops, six key practice development areas were identified by staff and working teams were formed to explore these areas further: (a) reducing seclusion and restraint, (b) increasing staff confidence by improving skills in de-escalation and physical safety, (c) ensuring best practice for pharmacological interventions, (d) introducing strengths-based philosophy and practices, (e) providing sexual safety training and awareness, and (f) improving access to therapeutic activities on the ward.  Staff received training in de-escalation and physical safety combined with trauma-informed perspectives, including the integration of strengths-based philosophies and practice. Staff were encouraged to reduce their use of clinical jargon and pejorative descriptions of consumers (e.g., chronic schizophrenic) and to focus on consumer strengths and resources during clinical discussions and handover. The training also focused on increasing staff awareness of childhood and adult adversity to help foster greater understanding, compassion, and respect for service users.  A working party comprising staff from nursing, medicine, pharmacy, and consumer backgrounds was established to investigate the use of medication in the unit. A literature review to identify best practice in the use of pharmacological interventions resulted in the revision of rapid sedation protocols on the ward, emphasising lower doses of sedating medication and more specific protocols for different groups of consumers, such as those who were neuroleptic naive, frail, or elderly.  One of the nursing staff developed a sexual safety training module, which all staff attended, and revised the ward policy and procedures to ensure best practice. A section of bedrooms, in clear view of the ward office, were designated as female-only, providing further security to female service users. Avoiding the use of male staff in the restraint of female service users was also a key focus of care.  Nursing and allied health staff worked together to improve the range and number of therapeutic activities on the ward. Many groups focused on relaxation, self-care, and positive relationships, including art, yoga, meditation, and strengths- and recovery-focused conversations.  The Consumer Participation Officer (D.H.) redeveloped the ward information booklet to improve the quantity and quality of information provided to service users about the admission process. The Officer also ensured that there were regular opportunities on the ward for informal groups in which staff and service users could meet to talk about concerns, experiences, and ideas for improvement. | Designed and implemented by:  The ward Clinical Nurse Consultant and Senior Clinical Psychologist devised a series of workshops and delivered training | The approach taken to achieve this goal was based on the belief that embedding the principles of trauma-informed care in ward practices, which promoted values such as choice, collaboration, trustworthiness, safety, and empowerment, would lead to practice improvement and cultural change. | Not specified |
| Isobel et al. (2017) | An acute mental health inpatient unit, New South Wales, Australia | No name assigned  Aim to improve on the clinical needs identified by nursing staff within the potential model, based upon areas of consumer care where the nursing staff required additional support. These included, managing acute psychosis, de-escalating aggression and responding to chronic self-destructiveness. | The key principles of TIC (safety, collaboration, empowerment, trustworthiness and choice) were used to guide the development of the model. Below are examples of operational processes implemented in-line with these principles.  ***Safety****:* this involved prioritising psychological and physical safety for all consumers and staff. Examples of operational processes included:   - Review all incidents of seclusion and restraint at weekly structured meeting through a trauma informed care lens - Structured nursing debriefs following incidents - Structured consumer debriefs following incidents - Sexual safety subproject to assess and improve sexual safety on the unit - Closed clinical supervision groups for all nurses - Reflective communications workshops developed and held for all nurses - De-escalation workshops developed and held for all nurses - Trial implementation of consumer safety planning tool to identify individualised triggers, responses and care needs - Awareness raising of vicarious trauma effects, prevalence and supports - Mechanisms for constructive professional peer feedback built into unit processes - Weekly clinical review sessions for nurses to discuss approaches to care of consumers, particularly those with complex trauma histories   ***Empowerment:***   - Review of case review processes to ensure consumer advocacy and involvement in treatment planning - Daily consumer led community meetings to discuss running of unit, issues and day plans - Recovery orientated language subproject to review professional communications on the unit - Review of processes of tribunal hearings to better accommodate needs of carers and consumers - Ongoing education and resources provided to all nurses on the prevalence and effects of trauma - Resources for consumers on effects of trauma and access to support services - Access for nurses to information and support around the effects of vicarious trauma   ***Choice:***   - Injection site negotiation sub project - Groups and activities review subproject - Purchase of increased sensory and diversionary resources   ***Collaboration:***   - Collaborative care planning subproject to support the standardised use of consumer led care plans - Implementation of clinical care coordination structuring of nursing day to increase therapeutic engagement, consistency of care, and nurse-consumer collaboration - Feedback from daily community meetings incorporated into unit meetings and planning - Feedback sought from consumers and carers on experiences of care   ***Trustworthiness:***   - Use of consumer developed care plans and wellness plans used to lead discussion in case review - Rules review sub project (see Isobel, 2015) resulting in regular and transparent processes of reviewing and communicating unit rules - Consistency of expectations and procedures - Display of unit expectations and rules - Admission pack for consumers containing all required information about unit - Daily display of timetable and events on whiteboard - Documentation of treating team members and pathways of communication for all consumers - Review of all unit routines - Ongoing in-service program developed to improve nursing therapeutic skills. | Designed and implemented by: A working group was formed to guide the development and implementation of the model. The working group began with key senior members of nursing staff including the Director of Nursing. Targeted members of the unit’s nursing staff were invited to join, as were other key people including an educator and a peer worker. Increasingly, nurses from the unit joined the group as they took on roles in the project and eventually the meetings of the group were made open to any interested and available nurses from the unit. Multidisciplinary representatives were invited to join the group in consultative and participatory roles but offers were initially declined. The group met every month on the unit and prioritised goals, identified actions and discussed barriers, obstacles and progress. | The use of TIC as the philosophical model of care stemmed from literature that acknowledges the high prevalence of traumatic experiences in persons receiving mental health care (Mueser et al. 2004; Muskett 2014; National Executive Training Institute 2005), the profound impact of trauma upon individuals (Read et al. 2007) and the responsibility of health services to provide physically and emotionally safe environments (Barton et al. 2009; Cusack et al. 2003). A model of care that is derived from a trauma-informed base aims to value the consumer in all aspects of care, enable the development of individual and flexible care plans and recognise the aspects of mental health inpatient units that can be overtly and covertly traumatising. | The paper states that “TIC refers primarily to psychological trauma experienced through interpersonal dynamics including disempowerment, abuse and neglect; often repetitive and across the lifespan.” |
| Jacobowitz et al. (2015) | A psychiatric hospital, providing short-term acute care in the following units: general adults, child and adolescents, older adults, chemical dependency, New York USA | No name assigned  Aim not specified | All levels of direct patient-care providers were encouraged to participate in regularly scheduled interdisciplinary meetings that discussed the principles of TIC and a specific patients’ or multiple patients’ behaviours. The meetings were of an educational nature. All staff members were introduced to the concept at orientation and at routinely scheduled intervals.  The paper states that a proprietary trauma-informed care program was part of the hospital’s routine provision of patient care for several years prior to the study. No other information about the programme was provided, however. | Not specified | Not specified | Participants were only included if they had experienced trauma, defined as “a stressor that involves either witnessing a death, experiencing the threat of death, actual or threatened serious injury, or actual or threatened sexual violence. The stressor may be indirect, such as learning that a relative or close friend was exposed to the stressor and also involve repeated or extreme indirect exposure to the details of the event (APA, 2013).” |
| Prytherch et al. (2020) | A residential female crisis house, North London, UK | No name assigned  Aim to provide environment reducing use of coercion as a means to manage risk, recognise importance of therapeutic relationship toward healing, in working with service users experiencing trauma. | The service runs on trauma-informed lines but is also open to those who have not experienced trauma. It was a women-only environment. The staff team were appointed based on their skills, attitudes, knowledge and experience rather than professional qualifications. All staff are trained and supervised in trauma-informed approaches, and initial assessments include routine inquiry about trauma.  When required, medical reviews were requested from medical doctors within the crisis team. When possible, the crisis team sent a female member of staff, and when this was not possible, residents were asked whether they would be willing to meet with a man. Staff also work closely with GPs to support women to have up-to-date prescriptions.  Risk was managed through psychological (safety planning, regular one-to-one sessions, agreed phone calls and check-ins) rather than physical containment, recognising the key role played by relationships and the potential for coercive measures to be re-traumatising.  Staff anxieties regarding risk were managed through clear structures and procedures, and through the constant availability of other team members working together and based in the same office within the house. For further details about the crisis house see (Cooke et al. 2019).  All care was consensual and rather than “observations”, there were regular “contacts” where staff checked-in verbally with women. Women had keys to their rooms and staff only used master keys if they have knocked three times and had no response. Participants described having regular opportunities to talk to staff whom they felt were compassionate, consistent and interested in the psychosocial context of their distress.  Service users were involved in the design and management of the service from the beginning. Self-referrals were accepted, and care was planned consensually by women and their workers.  Participants pointed out that only women who are considered able to keep themselves safe within the approach offered by the crisis house are offered a place. Those who are not may be admitted to hospital, sometimes under compulsion. Some might argue then that even within the trauma-informed crisis house, coercion still exists, albeit it covertly through the possibility of being referred to hospital. | Designed by: Service-users were involved in the design and management of the service from the beginning.  Implemented by: All staff are trained and supervised in trauma-informed approaches and so involved in implementation. However, how staff were involved in the design of the approach was not described. | The introduction of the paper states that trauma-informed approaches “developed out of psychological models of trauma (e.g., Herman, 2015; Van Der Kolk, 1987). TIAs recognise that many people who come into contact with mental health services will have experienced trauma and adversity, and that their distress is in many cases an understandable response to such experiences. TIAs also attend to the social, political and cultural context within which adversity is experienced. Since trauma often occurs within the context of relationships, TIAs view collaboration and trusting relationships as central to healing (Sweeney et al., 2016). Coercive measures are seen as potentially re-traumatising and as inimical to the development of trusting relationships and therefore to good care (Sweeney et al., 2018)”. | Not stated. |
